# The effect of long-term adherence to physical activity recommendations in midlife on plasma proteins associated with frailty in the Atherosclerosis Risk in Communities (ARIC) study

**DOI:** 10.1101/2024.11.04.24316702

**Authors:** Fangyu Liu, Jennifer A. Schrack, Keenan A. Walker, Jeremy Walston, Rasika A. Mathias, Michael E. Griswold, Priya Palta, B. Gwen Windham, John W. Jackson

## Abstract

Clinical trials have shown favorable effects of exercise on frailty, supporting physical activity (PA) as a treatment and prevention strategy. Proteomics studies suggest that PA alters levels of many proteins, some of which may function as molecules in the biological processes underlying frailty. However, these studies have focused on structured exercise programs or cross-sectional PA-protein associations. Therefore, the effects of long-term PA on frailty-associated proteins remain unknown. Among 14,898 middle-aged adults, we emulated a target trial that assigned individuals to either (i) achieve and maintain the recommended PA level (≥150 minutes/week of moderate-to-vigorous physical activity [MVPA]) through 6 (±0.3) years of follow-up or (ii) follow a “natural course” strategy, where all individuals engage in various amounts of habitual MVPA. We estimated the effects of long-term adherence to recommended MVPA versus the natural course strategy on 45 previously identified frailty-associated proteins (log 2 transformed and standardized) at the end of the follow-up using inverse probability of weighting (IPW) and iterative conditional expectations (ICE). We found that long-term adherence to recommended MVPA improved the population levels of many frailty-associated proteins (ranged from 0.04 to 0.11 standard deviation); the greatest benefits were seen in proteins involved in the nervous system (e.g., voltage-dependent calcium channel subunit alpha-2/delta-3 [CACNA2D3], contactin-1 [CNTN1], neural cell adhesion molecule 1 [NCAM1], and transmembrane protein 132D [TMEM132D]) and inflammation (e.g., high-temperature requirement serine protease A1 [HTRA1] and C-reactive protein [CRP]). Our findings suggest long-term engagement in adequate habitual PA may reduce frailty risk through specific nervous systems and inflammatory proteins. (250 words)

## Introduction

Treating and preventing frailty, a state of reduced reserve and increased vulnerability, among older adults is an important public health priority, as frailty predicts adverse health outcomes, including higher healthcare utilization and cost, disability, death, and lower quality of life.^1,2^ Physical activity (PA) has been shown to potentially regulate systems involved in the onset and progression of frailty, including metabolism, inflammation, and mitochondrial function.^3,4^ This multisystem benefit, congruent with the multifactorial etiology of frailty, supports the use of PA as a promising strategy to treat and prevent frailty. Many trials have shown that PA, accumulated mostly through structured exercise programs, can reduce frailty scores^5–12^ and incident phenotypic frailty^13–15^. However, these trials collected no or a limited number of biomarkers that would otherwise provide insight into the etiological pathways linking PA to a reduction in frailty. Therefore, an essential unanswered question centers on which frailty-associated biological mechanisms are affected by PA. Using frailty-associated proteins as the outcome, we can better understand the mechanistic pathways that link PA to reduced frailty risk, informing the biological basis and effectiveness of PA as an intervention strategy for frailty.

Proteomics studies of PA and exercise in middle-aged and older adults are emerging (Table 1).^16–24^ Some studies examined how proteins were differentially expressed across various daily levels of PA/exercise,^21–24^ while others examined how proteins changed before and after exercise training.^16–20^ Though not directly targeting proteins or pathways implicated in frailty, many of these studies reported pathways modulated by PA that overlapped with pathways related to frailty risk, e.g., angiogenesis, extracellular matrix, metabolism, and inflammation.^25^ However, two gaps remain. First, studies on protein changes pre-and post-exercise have focused on structured exercise programs with one fixed activity type (e.g., aerobic, resistance, or high-intensity interval training), frequency, and duration.^16–20^ Yet, free-living PA consists of a mixture of activity types and is performed in a less structured manner. To this end, it is unknown whether adequate free-living PA can alter the etiologic pathways of frailty. Second, studies on PA/exercise in daily living have been exclusively cross-sectional.^21–24^ Therefore, little is known about the long-term effects of engaging in adequate free-living PA on frailty-associated proteins.

**Table 1.**
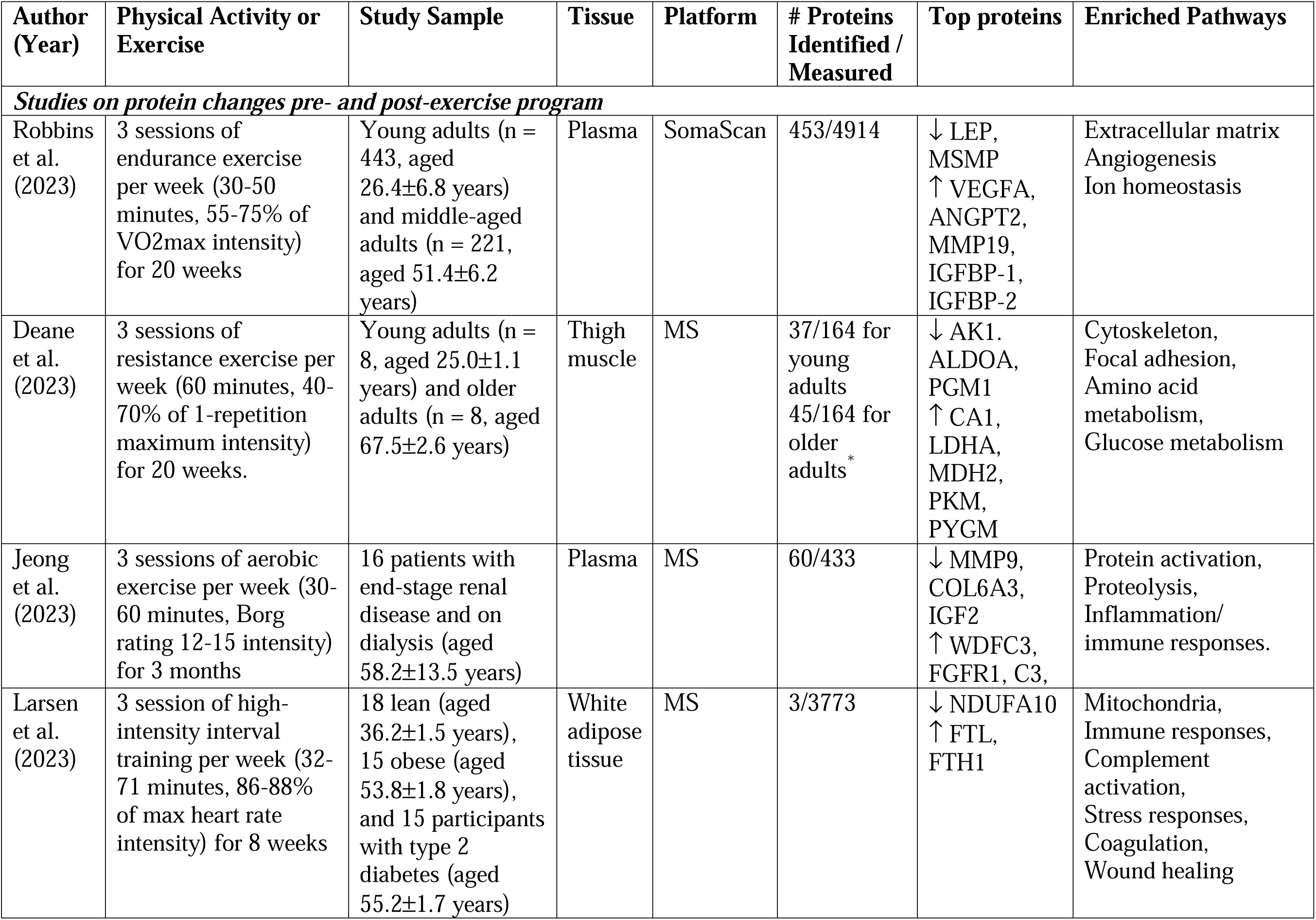
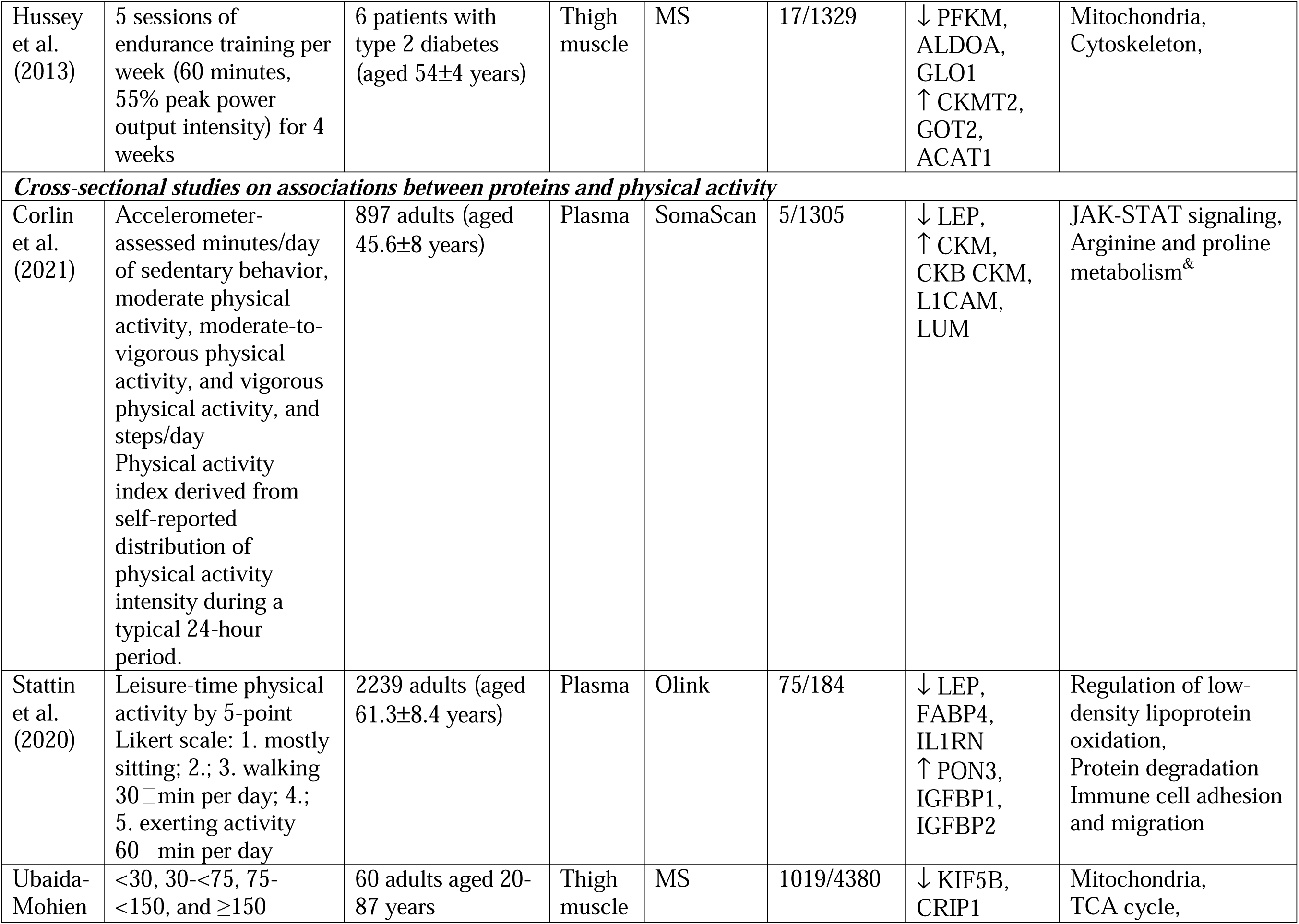
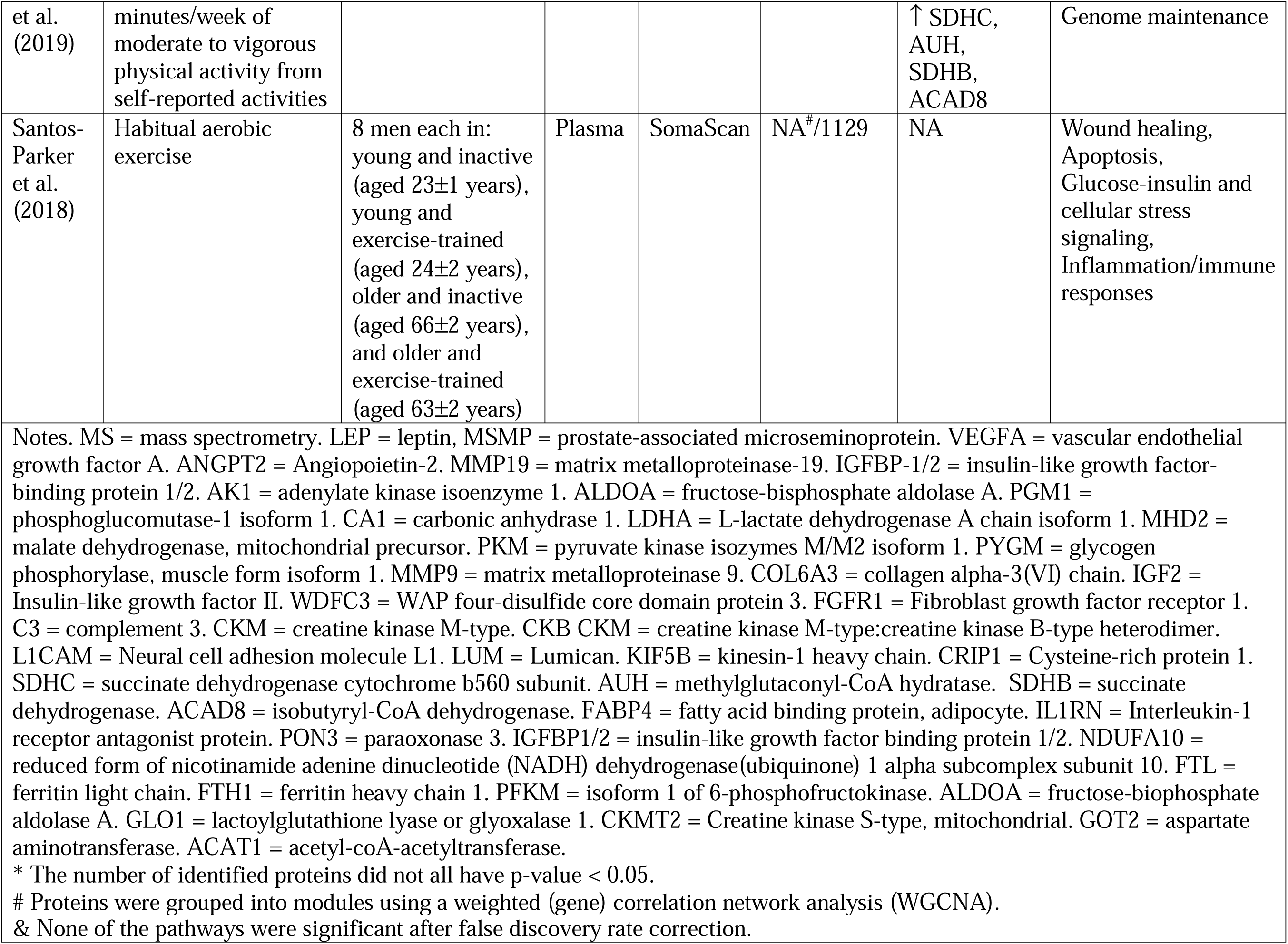
Current literature of proteomics of physical activity or exercise.

To address these research gaps, this study examined the effects of long-term adherence to the United States Centers for Disease Control and Prevention (US CDC) recommended ≥150 minutes/week of moderate to vigorous PA (MVPA)^26^ in midlife on 45 proteins previously linked to frailty in later life.^27^ Among 14,898 middle-aged participants from the Atherosclerosis Risk in Communities (ARIC) study, we emulated a trial where participants were randomized at enrollment to either: (i) receive an intervention strategy that ensures participants achieved and maintained ≥150 minutes/week of MVPA during 6-year follow-up or (ii) a “natural course” strategy where participants engaged in various amounts of habitual MVPA. We estimated the per-protocol effects of strategy (i) versus (ii) on frailty-associated proteins in this emulated trial using parametric g-formula based on iterative conditional expectations (ICE) and inverse probability of weighting (IPW) to account for non-adherence to the assigned strategy. These per-protocol effects (which contrast the population-level differences in the protein outcomes at the end of follow-up between the two strategies) are measures of generalized impact, a generalization of population attributable risk^28^ that informs the population-level impact of everyone meeting or exceeding the guidelines for MVPA.^29^ We hypothesized that achieving and maintaining ≥150 minutes/week of MVPA would increase the levels of proteins associated with lower risks of frailty and decrease the levels of proteins associated with higher risks of frailty.

## Methods

### Target trial specification

The target trial emulation framework entails specifying a high-level protocol (e.g., eligibility, intervention strategies, follow-up, outcome assessment, and statistical analysis) of a hypothetical randomized trial, i.e., a target trial, that answers the research question of interest before describing how the observed data will be used to mimic the target trial.^30^ This framework guides the estimation of the causal effect using the gold standard for estimating the effect of an intervention strategy and allows for transparent evaluation of causal estimates.

The target trial protocol for this paper is summarized in Table 2. Briefly, to assess the effect of long-term adherence to the recommended ≥150 minutes/week of MVPA on frailty-associated plasma proteins, the target trial randomizes participants who are capable of engaging in MVPA to either: (i) receive the intervention strategy that ensures the achievement and maintenance of the recommended MVPA level throughout the follow-up, or (ii) a “natural course” strategy that allows participants to engage in a habitual MVPA level, which may or may not be below the recommended level. Participants assigned to either strategy are free to engage in different levels of MVPA as long as they meet the strategy’s MVPA threshold (≥150 minutes/week for the intervention strategy and ≥0 minute/week for “the natural course” strategy). Such strategies are an example of a representative intervention.^31,32^

**Table 2.** Target trial specification and emulation.

| **Component** | **Target Trial** | **Emulation using observational data** |
| --- | --- | --- |
| Eligibility criteria | Adults aged 45-66 years who are free of functional limitations that prevent them from engaging in physical activity. | Same, except that whether a participant can engage in physical activity was not directly measured in ARIC. However, it can be argued that the number of participants with functional limitations severe enough to prevent physical activity is rare given the relatively young age.<br>We excluded participants who reported neither White nor Black for self-reported race due to extremely small number (n = 49). |
| Intervention strategy | Intervention: achieving the CDC recommended 150 minutes/week of MVPA.<br>Control: performing MVPA as the participants choose. | Same. |
| Treatment assignment | Treatment is randomized at enrollment into the study. | The randomization is emulated by adjusting for confounding factors required to meet the exchangeability assumption including age, sex, race, education, smoking status, number of chronic conditions, BMI, eGFR, and total cholesterol (see Methods and Supplemental Methods for details). |
| Baseline | At enrollment (at randomization) | Same, except enrollment is taken as the date of Visit 1 (1987-1989) |
| Follow-up | Starts at enrollment and ends at 6 years after enrollment, death, or loss to follow-up, whichever occurs first. | Same, except enrollment is taken as the date of Visit 1 (1987-1989) and 6-year follow-up is taken as Visit 3 (1993-1995). The actual follow-up time is approximately 6 years but may vary among participants. We assumed that participants who met the guideline at both visits also met the guideline during the intervening period between the two visits. |
| Outcome | Frailty-associated plasma protein levels at the end of 6-year follow-up. | Same, except that the actual follow-up time may vary around 6 years. |
| Causal contrast of interest | Per-protocol effect, i.e., the effect of adhering to the assigned intervention/control strategy during the 6-years of follow-up. | Same, except that the long-term adherence is approximated by two measurements of time spent in MVPA per week at Visits 1 and 3. |
| Statistical analysis | Per-protocol effect estimated via comparison between the intervention/control groups using g-formula or MSM by IPW methods to adjust for pre- and post-baseline prognostic factors associated with adherence to the intervention and loss to follow-up including death. | Same, specifically g-formula via ICE and MSM by IPW methods were used. |

Note. MVPA = moderate-to-vigorous physical activity. BMI = body mass index. eGFR = estimated glomerular filtration rate. ICE = iterative conditional expectation. MSM = marginal structural model. IPW = inverse probability weighting.

The effect of long-term adherence to the recommendation is a per-protocol effect^33^ as it compares outcomes between individuals adhering to two strategies, intervention versus “natural course”, throughout follow-up. Under the “natural course” strategy, regardless of what level of MVPA these participants engage in during follow-up, they are always adherent to this strategy because no constraints are placed on their MVPA level. Under the intervention strategy, participants are no longer adherent to the assigned strategy when they engage in <150 minutes/week of MVPA at any time during follow-up. Because post-randomization factors can influence whether participants adhere to the assigned strategy or remain in the trial, appropriate statistical methods such as the parametric g-formula and IPW are needed to account for these factors even in a trial with non-adherence.^33,34^

### Emulation of the target trial using the ARIC study

We emulated the target trial described above using 14,898 participants at Visit 1 (1987-1989) of the ARIC study who were followed until Visit 3 (1993-1995, Figure 1A). The ARIC study is an ongoing community-based cohort study that enrolled middle-aged participants (aged 45-64 years) from 4 communities across the United States: Washington County, MD; Forsyth County, NC; northwestern suburbs of Minneapolis, MN; and Jackson, MS.^35^

**Figure 1.**
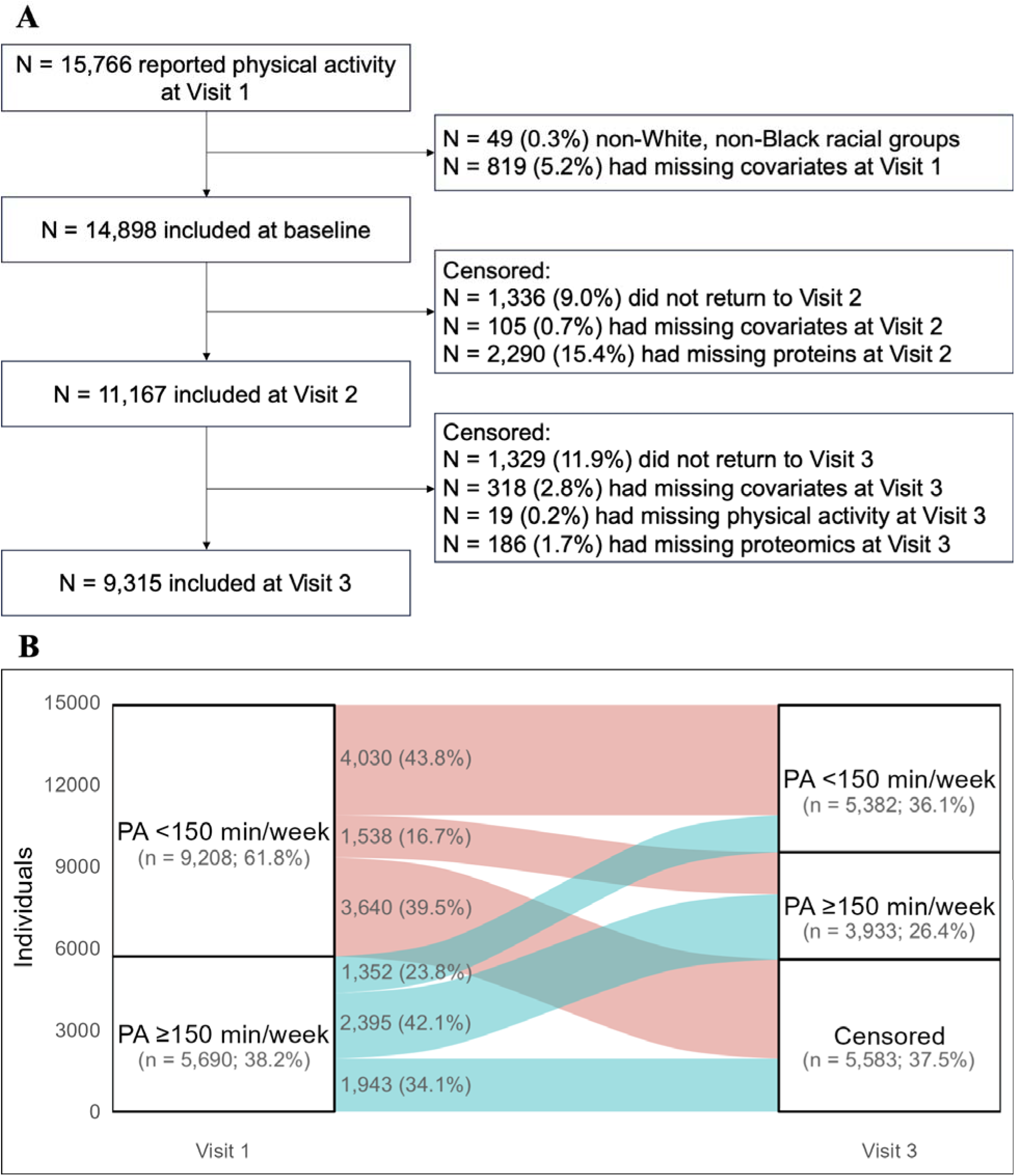
Sample selection of the emulated trial (A) and transitions between achieving and not achieving ≥150 minutes/week of moderate-to-vigorous physical activity (MVPA) during the follow-up (B).

The emulated trial closely follows the protocol of the target trial with three exceptions (Table 2). The deviations (assumption that no participants had limitations in engaging in PA, exclusion of races with a small number of participants in the sample, and approximation of long-term adherence to PA by measurements at two study visits only) and the rationales are described in Supplemental Methods. To emulate the randomization at baseline, based on the assumed causal structure of the observed data depicted in Figure 2, we assumed that whether a participant met the PA guidelines at baseline was as if randomized after conditioning on time-fixed covariates (i.e., age, sex, race, and education) and the baseline values of time-varying covariates (i.e., smoking status, number of chronic conditions, body mass index [BMI], glomerular filtration rate [eGFR], and total cholesterol). At follow-up, as we would in the target trial, we censored participants who did not return to study visits, had missing covariates and protein measurements, or had missing physical activity measurements at Visit 3. We assumed that adherence to the assigned strategy and censoring were both as if randomized conditional on the baseline time-fixed and time-varying covariates, the post-baseline values of the time-varying covariates (at the current and additionally, for continuous covariates, the previous two visits), baseline MVPA, and protein levels at the last visit (if available; proteins were not measured at Visit 1). We used the ICE approach^36^ of the parametric g-formula method and the IPW method^32^ (described below) to appropriately emulate the randomization at baseline and properly handle the covariates associated with non-adherence to their assigned strategy or censoring. These two methods also emulate the representative intervention. Under the specified intervention strategy, both methods implicitly assign participants an MVPA level as a random draw from the observed MVPA distribution among all participants who have observed MVPA ≥150 minutes/week and the same covariate history.^31,32,36^ This implicit assignment was performed at each visit.

**Figure 2.**
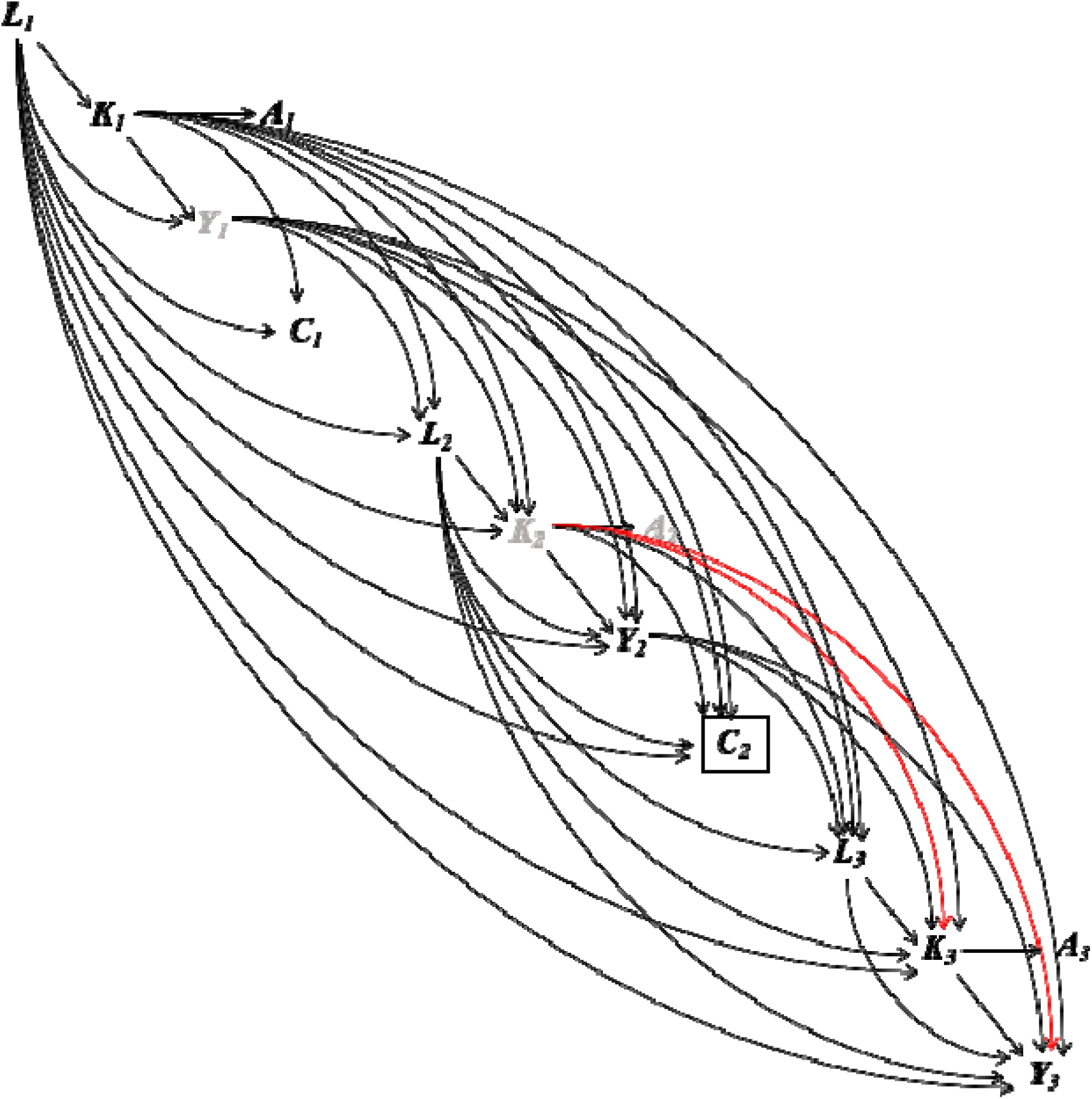
Causal structure of the emulated trial in ARIC Notes. The subscript denotes the three visits of the ARIC study used in the study. At each visit, *K_t_* denotes the minutes of MVPA. *A_t_* indicates whether the recommended ≥150 minutes/week of MVPA is achieved. *A_t_* is a coarsening variable of *K_t_*. Therefore, there is one arrow from *K_t_* to *A_t_*, and all other arrows in the diagram are to and from *K_t_*. *Y_t_* denotes the plasma protein level. *L_t_* denotes the confounders including smoking status, number of chronic conditions (0, 1, or ≥2), BMI, eGFR, and total cholesterol. *L_1_*also includes time-fixed confounding including age, sex, race, and education. *C_t_* denotes loss to follow-up after visit *t*. The box around *C_t_* denotes that only participants remained in the study were included in the study. *Y_1_* (protein level at Visit 1) and *K_2_* (MVPA minutes at Visit 2) are denoted by gray color because they were unmeasured in ARIC. The red path is the open backdoor path between *K_3_* and *Y_3_* (the outcome of interest) even after conditioning on all confounders and *Y_2_*.

### PA measurement in ARIC

ARIC participants were asked to report up to 4 sports or exercises in which they most frequently engaged at Visit 1 and Visit 3. For each reported sport or exercise, the participants reported the duration (hours/week) and the frequency (number of months/year).^37^ To assess whether a participant met the PA guidelines, the intensity of each reported sport or exercise was assigned a metabolic equivalent of task (MET) based on the 2011 Compendium of Physical Activities.^38^ Sports or exercises with MET >3 but ≤6 were considered moderate, and those with MET > 6 were considered vigorous. For each sport or exercise that met moderate or vigorous intensity levels, the weekly level (minutes/week) was estimated by multiplying the frequency and duration by 4.35 weeks/month and dividing by 52.18 weeks/year. A participant’s MVPA in minutes/week was then calculated as the sum of the weekly levels of all moderate-intensity sports or exercises and two times the weekly levels of all vigorous-intensity sports or exercises.^37^ Participants not reporting any sport or exercise meeting the MVPA intensity were assigned zero for MVPA minutes/week. Participants with MVPA minutes/week ≥150 were considered to meet the PA guidelines.

### Midlife frailty-associated plasma proteins

The 45 proteins used in this analysis were selected based on evidence from our previous study on the midlife proteomics of frailty in later life.^27^ These proteins were measured using the SomaScan platform (Version 4.0, SomaLogic, Inc., Boulder, Colorado). The SomaScan platform quantifies the relative abundances of the plasma proteins and protein complexes using single strands of DNA with chemically modified nucleotides, called modified aptamers or “SOMAmers”, which act as protein-binding reagents with defined three-dimensional structures and unique nucleotide sequences. The abundances of the SOMAmers are quantified using dynamic DNA detection technology and represent the levels of the proteins in plasma.^39^ The 45 proteins were significantly differentially expressed in midlife (Visit 3) among participants who were frail (meeting at least 3 of the 5 criteria of the physical frailty phenotype: weight loss, weakness, slowness, exhaustion, and low PA^40^) in late-life (Visits 5-7) compared to participants who were robust (meeting none of the 5 criteria) in late-life after Bonferroni correction and adjustment for age, sex, race-center, education, family income, drinking and smoking status, dietary protein intake, total cholesterol, eGFR, history of hypertension, diabetes, coronary heart disease, heart failure, cancer, and chronic lung disease, and functional limitation (Figure 3).^27^

**Figure 3.**
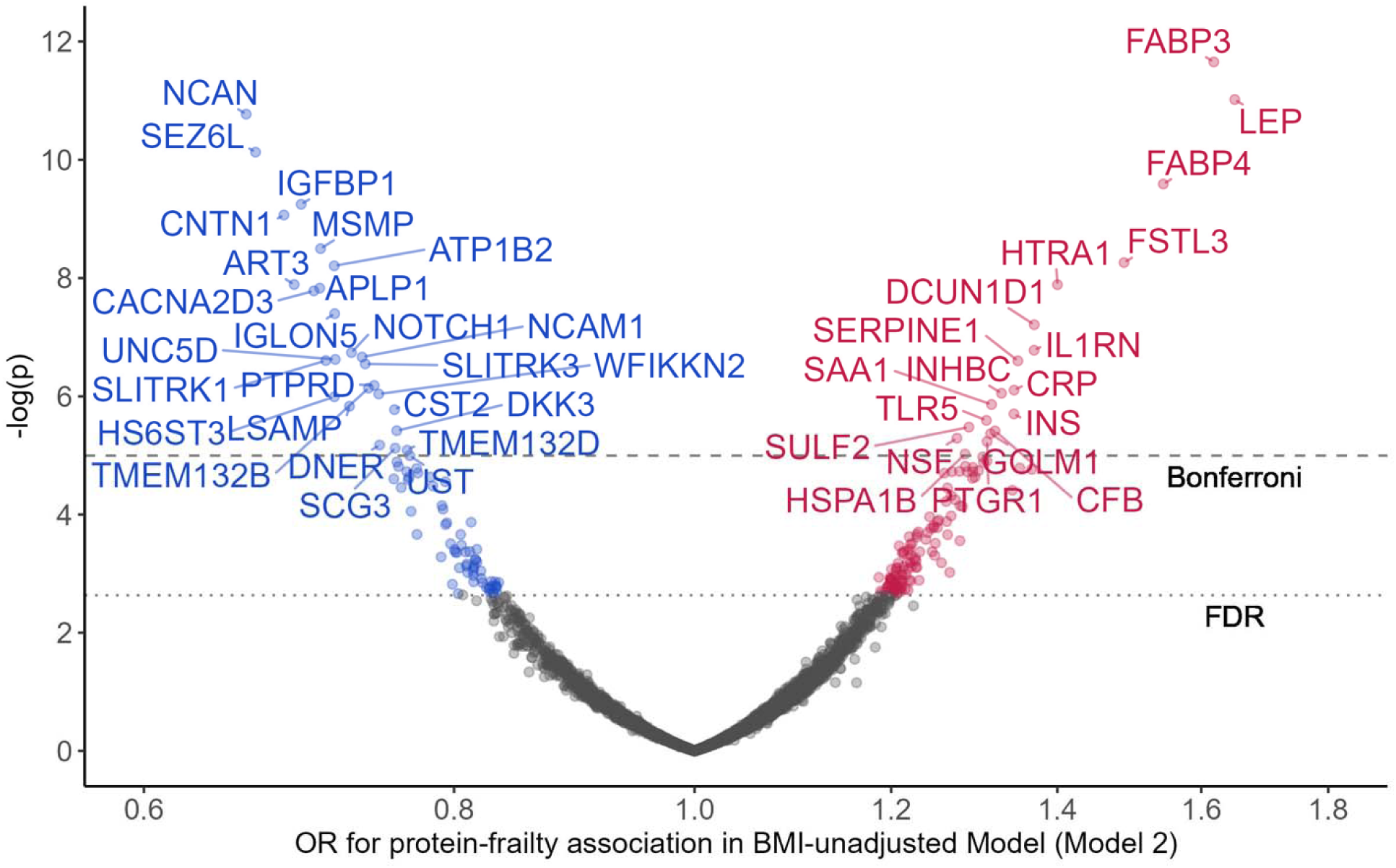
The 45 midlife proteins associated with late-life frailty from previous proteomics study of frailty (reproduced from ref ^27^).

### Main statistical analysis

The ICE and IPW methods can estimate the per-protocol effect of a representative intervention under the assumptions of exchangeability (or no unmeasured confounding), positivity, consistency, and no model misspecification. (see Supplemental Methods for discussions on these assumptions).^31,32,36^ These two methods condition, at each time point, on the time-varying confounders up to that time point, properly handling the “exposure-confounder feedback”, i.e., a feedback loop of MVPA → confounder → MVPA. An overview of these methods has been described.^41^

We used the following functional forms for the covariates (see Supplemental Methods for the measurements of covariates) in the ICE and IPW models: age (continuous with a linear term), sex (men/women), race (Black/White participant), education (nominal categorical, less than completed high school, high school/GED/vocational school, or any college), smoking status (ever or never smoker), number of chronic conditions (nominal categorical, 0, 1, or ≥2, including namely, hypertension, diabetes, coronary heart disease, heart failure, stroke, cancer, and chronic lung disease), BMI (continuous with a restricted cubic spline of 3 internal knots at quartiles), eGFR (continuous with a restricted cubic spline of 2 internal knots at tertiles), and total cholesterol (continuous with a linear term). MVPA minutes/week was modeled linearly in both methods. Protein level at the last visit was modeled using a restricted cubic spline with 3 internal knots at quartiles. The functional forms were selected based on residual plots and likelihood ratio tests (data not shown).

We estimated the 95% confidence intervals using 1000 samples from the non-parametric cluster bootstrap^42^ where individuals were sampled with replacement and all of their follow-up observations were retained. For the IPW method, to ensure satisfactory control of confounding by the confounders including MVPA minutes/week and protein level from the last visit, we checked the standardized mean differences (SMD) of these variables (1) between participants who achieved ≥150 minutes/week of MVPA and those who did not, respectively at Visit 1 and Visit 3 (conditional on MVPA ≥150 minutes/week at Visit 1 and no prior censoring) and (2) between participants who were censored and those who remained under study, respectively at Visit 1 and Visit 2 (conditional on MVPA ≥150 minutes/week at Visit 1 and no prior censoring). The SMDs were calculated before and after applying the final weights at respective visits.^43,44^ We used SMD within ±0.1 as good covariate balance and within ±0.25 as acceptable covariate balance.^45,46^ We also checked the distribution of the final weights at Visit 3 as recommended in previous literature.^47^ All protein levels used in this analysis were log 2-transformed and standardized using their standard deviation (SD) at Visit 3. The detailed procedures of the two methods, including the g-formula, the identifying assumptions, the model specifications, and the covariate measurements are described in Supplemental Methods.

### Sensitivity analyses

To check that residual confounding and selection bias by individual chronic conditions, drinking status, family income, and histories of binary and categorical confounders (excluded from the covariate list due to positivity violation) were reasonably controlled by the included covariates, we checked the SMDs across levels of adherence to the recommended MVPA and levels of censoring as described earlier.

Next, we performed the same ICE and IPW analyses after excluding participants who had major chronic conditions (i.e., coronary heart disease, heart failure, stroke, cancer, chronic lung disease, eGFR <30 ml/min/1.73 m^2^) or underweight (BMI <18.5 kg/m^2^) at baseline (n = 2,951, 19.8%, Figure S1). By restricting the study sample to a healthier subset of individuals, the variability of unmeasured confounders may be reduced and therefore better controlled.

Finally, we performed a tipping analysis^48^ (Supplemental Methods) at Visit 1 to assess how strongly an unmeasured confounder at or before Visit 1 must be associated with MVPA at Visit 1 (i.e., the exposure) and the protein levels at Visit 3 (i.e., the outcomes) to bring the effects of exposure on the outcomes to the null (i.e., the tipping point). This tipping analysis was not a direct assessment of the robustness to unmeasured confounding of *the effects of interest* in this paper. However, robustness to unmeasured confounding suggested by the tipping analysis may indicate the robustness of our effects of interest. To provide a context of plausible unmeasured confounders that would bring the effects to the null (e.g., what they could be and how strongly they are associated with the exposure and the outcome), we used the confounding of Visit 2 BMI, eGFR, and protein level on the effects of MVPA *at Visit 3* on protein levels at Visit 3 to compare with the tipping points for the effects of MVPA *at Visit 1* on protein levels at Visit 3. We discuss the rationale and the detailed procedures for the tipping analysis in the Supplemental Methods.

To address the unobserved outcome due to death, we performed another sensitivity analysis using the same ICE and IPW procedures after excluding all observations from participants who died before Visit 3 (n = 794, 5.3%, Figure S2). Death before Visit 3 was defined by not attending Visit 3 and having a date of death on or prior to February 5^th^, 1996, the last observed Visit 3 date. This approach can result in selection bias,^49^ but similar estimates to the main analysis can suggest that our findings were robust to death-related selection bias.

The 95% confidence intervals in all sensitivity analyses were obtained using the same non-parametric cluster bootstrap as in the main analysis. All analyses were performed in R (version 4.3.1).^50^

## Results

### Participant characteristics

The baseline (Visit 1) characteristics of the 14,898 participants are summarized in Table 3 by whether they met PA guidelines (≥150 minutes/week of MVPA) at baseline. Participants who met the guidelines had similar baseline age, cholesterol, and eGFR, but lower BMI, compared to those who did not meet the guidelines. Women, participants self-identified as Black, and participants who had less than completed high school education were less likely to meet PA guidelines. The prevalence of coronary heart disease, heart failure, stroke, cancer, and lung disease was low (<10%). Participants with more chronic conditions were less likely to meet PA guidelines.

**Table 3.** Baseline characteristics of participants who achieved ≥150 minutes/week of MVPA and who did not achieve ≥150 minutes/week of MVPA at baseline.

| Mean (SD) / N (%) | MVPA <150<br>minutes/week<br>n = 9208 | MVPA $\geq 150$<br>minutes/week<br>n = 5690 |
| --- | --- | --- |
| Age, years | 54.1 (5.7) | 54.4 (5.8) |
| Women | 5496 (59.7%) | 2689 (47.3%) |
| Black | 2989 (32.5%) | 900 (15.8%) |
| Education |  |  |
| Less than completed high school | 2623 (28.5%) | 874 (15.4%) |
| High school/GED/vocational school | 3852 (41.8%) | 2255 (39.6%) |
| Any College | 2733 (29.7%) | 2561 (45.0%) |
| Ever smokers | 5341 (58.0%) | 3380 (59.4%) |
| Total cholesterol, mmol/L | 5.6 (1.1) | 5.5 (1.0) |
| eGFR, ml/min/1.73 m<sup>2</sup> | 101.7 (14.0) | 101.3 (12.2) |
| BMI, kg/m<sup>2</sup> | 28.1 (5.7) | 26.9 (4.5) |
| Number of chronic conditions |  |  |
| 0 | 4475 (48.6%) | 3231 (56.8%) |
| 1 | 3134 (34.0%) | 1682 (29.6%) |
| $\geq 2$ | 1599 (17.4%) | 777 (13.7%) |
| Hypertension | 3476 (37.8%) | 1706 (30.0%) |
| Diabetes | 1237 (13.4%) | 549 (9.7%) |
| Coronary heart disease | 394 (4.3%) | 346 (6.1%) |
| Heart failure | 513 (5.6%) | 206 (3.6%) |
| Stroke | 185 (2.0%) | 85 (1.5%) |
| Cancer | 518 (5.6%) | 361 (6.3%) |
| Lung disease | 576 (6.3%) | 230 (4.0%) |
| MVPA time, minutes/week | 31.4 (47.5) | 335.0 (168.1) |

Notes. eGFR = estimated glomerular filtration rate. BMI = body mass index. MVPA = moderate-to-vigorous physical activity.

The transitions between meeting PA guidelines and censoring are summarized in Figure 1B. About one-third of the participants met PA guidelines at Visit 1. Of the participants who met PA guidelines at Visit 1, 42.1% maintained ≥150 minutes/week of MVPA at Visit 3, compared to 23.8% who decreased to <150 minutes/week at Visit 3. The remaining 34.1% of the participants were censored. Of the participants who did not meet the guidelines at Visit 1, 43.8% maintained <150 minutes/week of MVPA at Visit 1, compared to 16.7% who increased to ≥150 minutes/week at Visit 3. The remaining 39.5% of the participants were censored.

### Covariate balance for IPW method

Because the weight estimation was based on a different model for each outcome protein that included only the history of the outcome protein, not other proteins, this resulted in different weights for different outcome proteins. Therefore, the covariate balance was examined separately for each outcome protein. Figures 4 and 5 depict the balance before and after weighting for CACNA2D3 and HTRA1 as the outcomes. The balance for all other proteins can be found in Figures S3 and S4. For all the 45 protein outcomes, after applying the IPW final weights, good balance (SMD within ±0.1 SD) was achieved for all included confounders, MVPA minutes/week, and protein levels at all available visits (i) between participants who achieved ≥150 minutes/week of MVPA and those who did not, both at Visit 1 and at Visit 3 (conditional on MVPA ≥150 minutes/week at Visit 1 and no prior censoring) and (ii) between participants who were censored and those who remained under study, both at Visit 1 and at Visit 2 (conditional on MVPA ≥150 minutes/week at Visit 1 and no prior censoring). Moreover, the final weights at Visit 3 had a mean close to 1 for all protein outcomes (Table S1).

**Figure 4.**
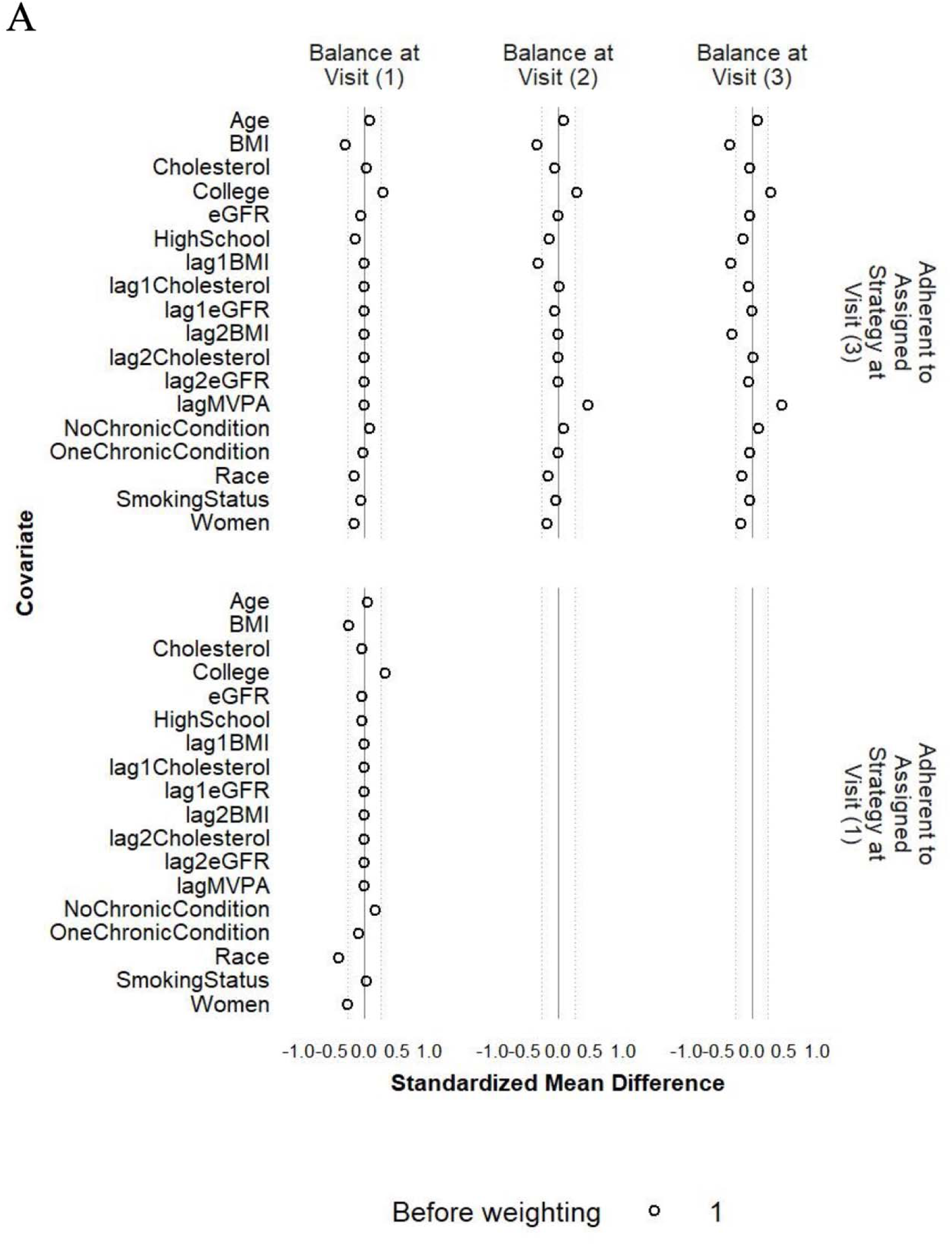
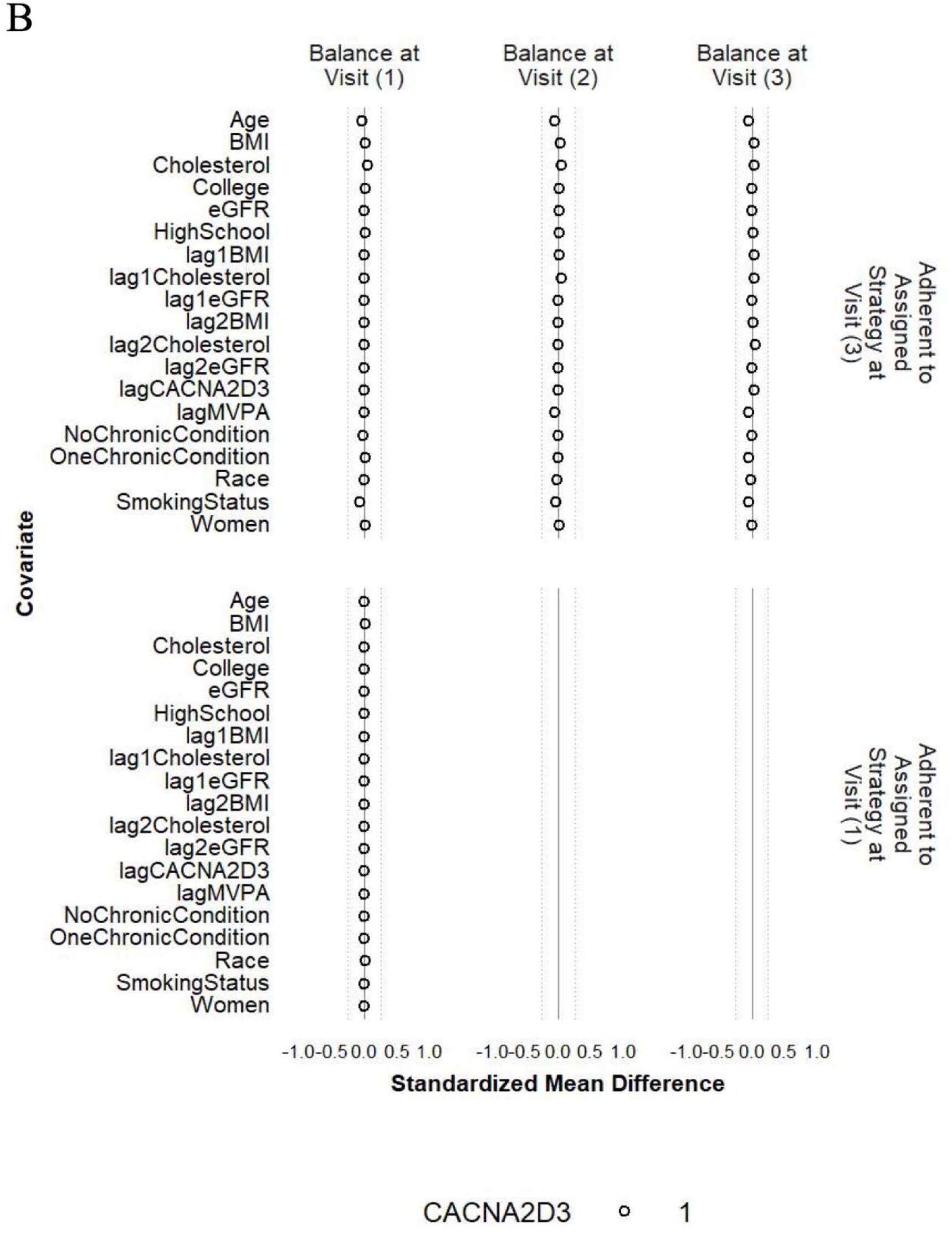
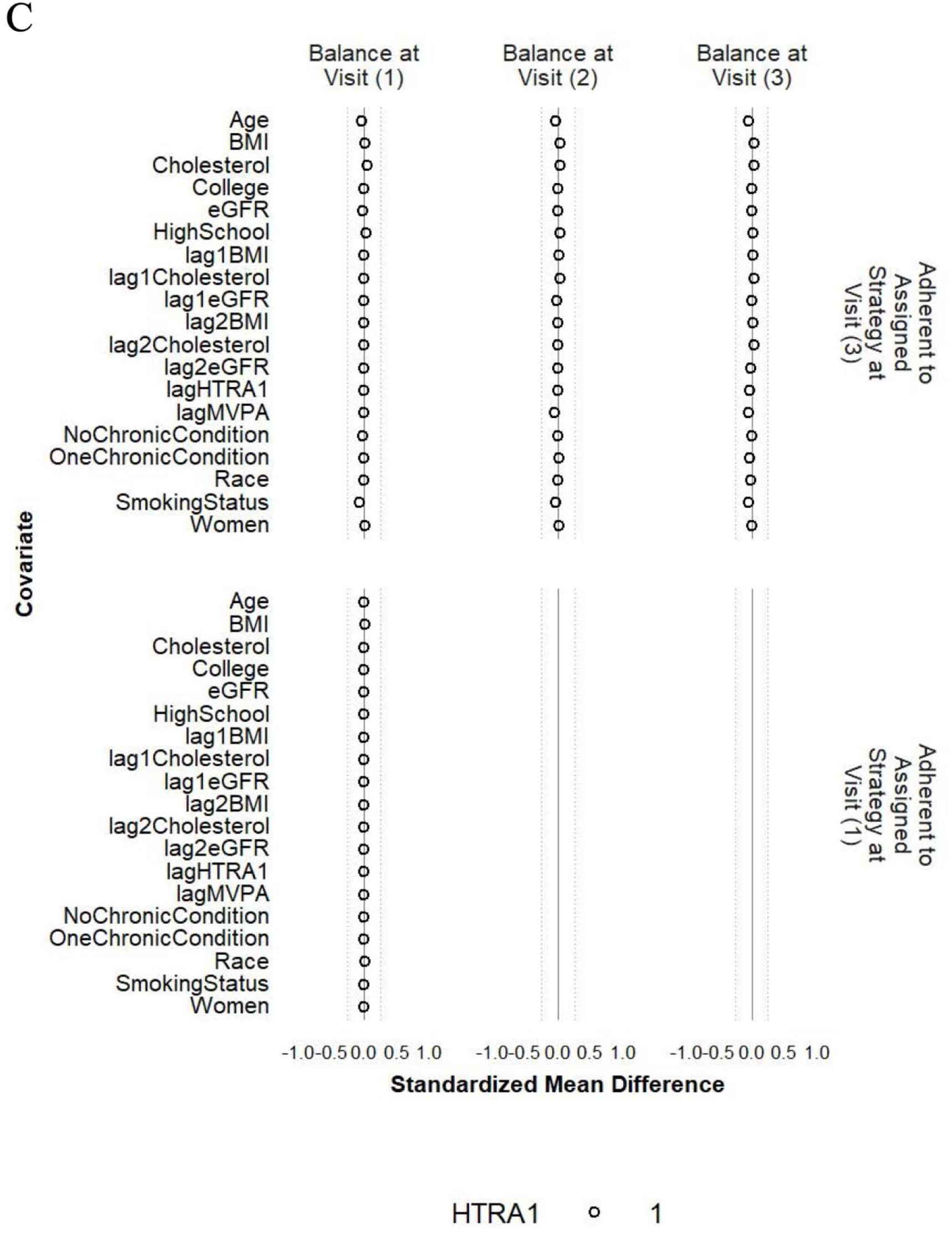
Balance of included covariates between participants who achieved ≥150 minutes/week of MVPA and those who did not at Visit 1 and Visit 3 before weighting (A), after weighting with CACNA2D3 as the outcome (B) and after weighting with HITRA1 as the outcome (C). Note. The circle represents the intervention strategy. The dashed lines denote ±0.25 SD.

**Figure 5.**
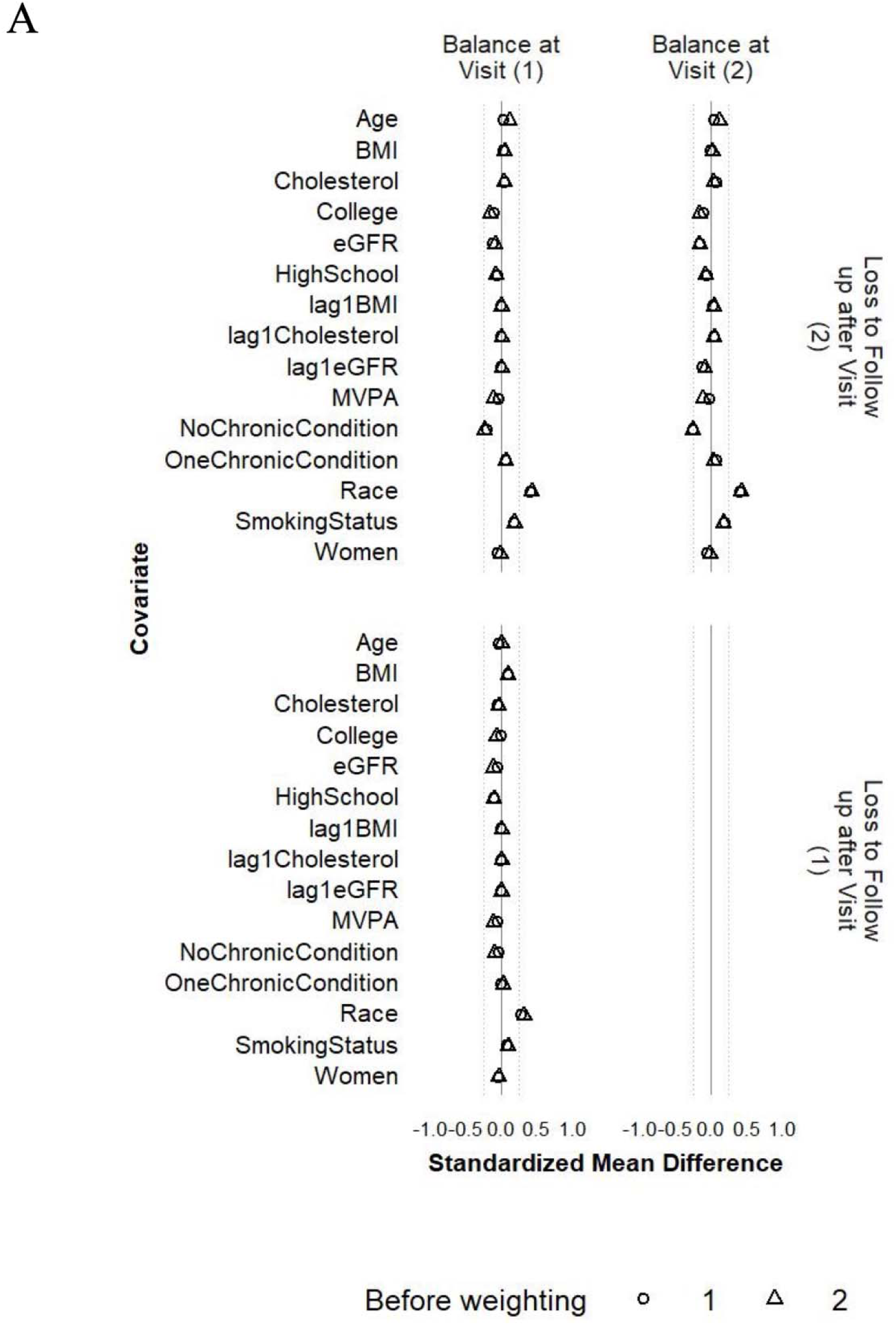
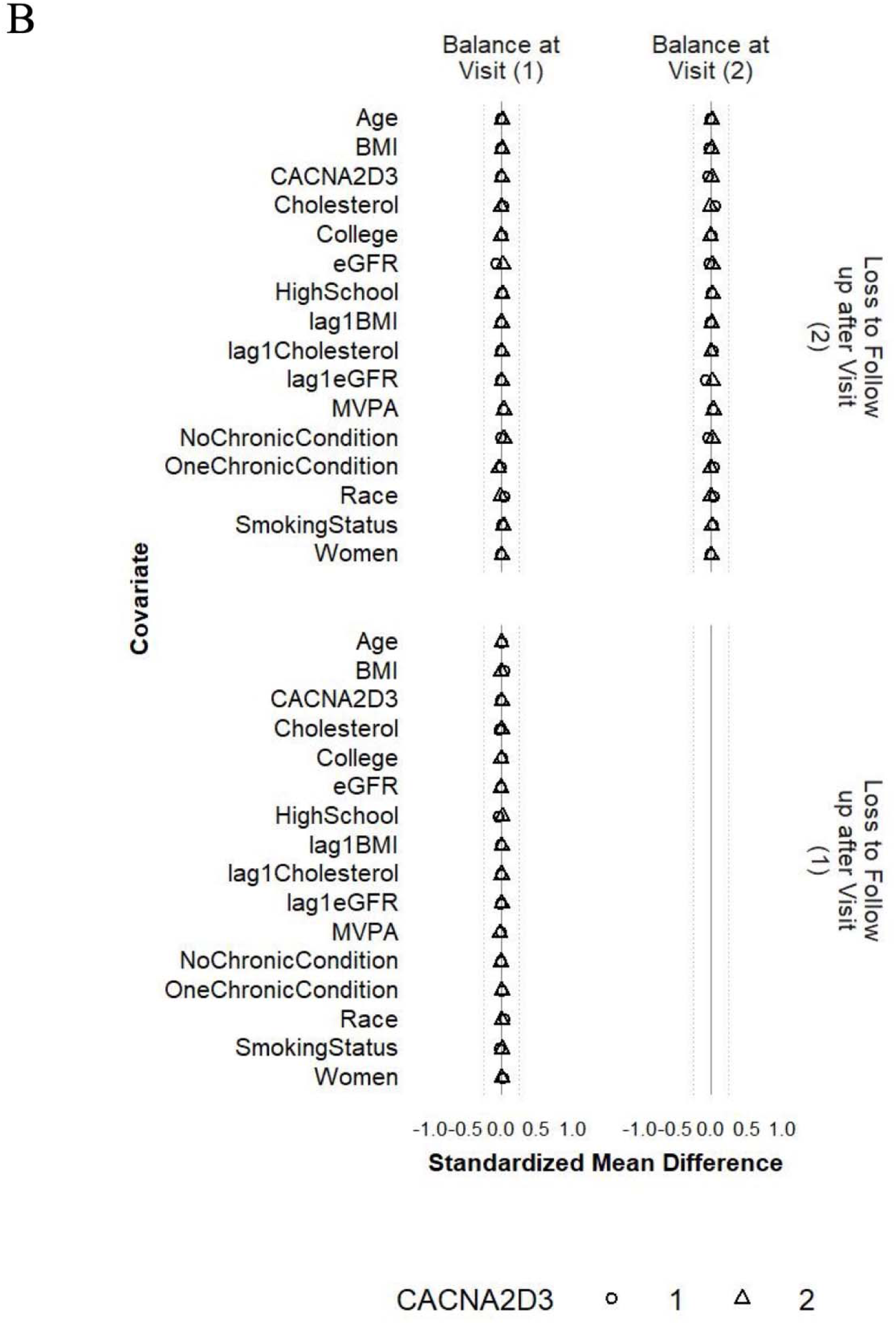
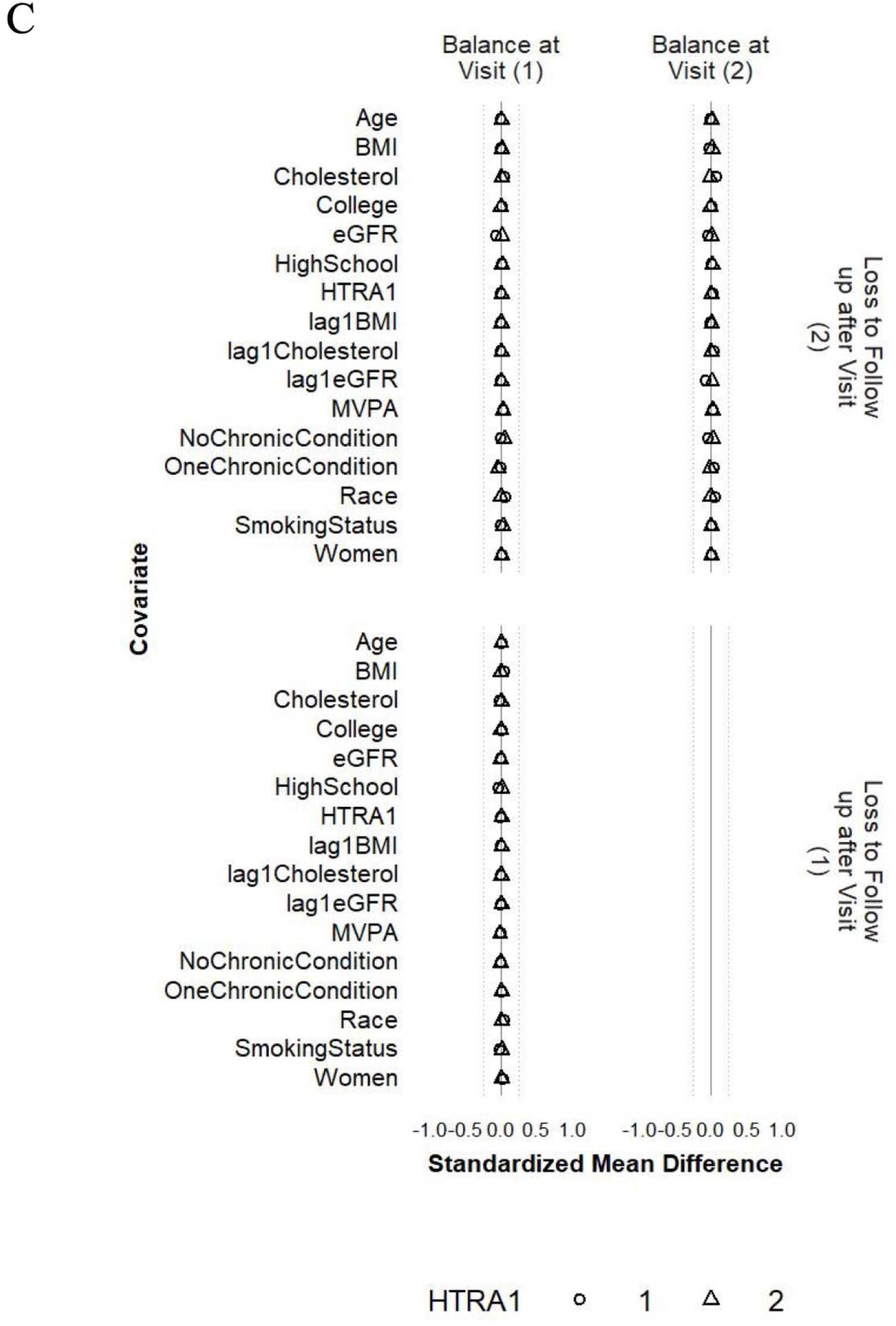
Balance of included covariates between participants who were lost and those who remained under study at Visits 1 and 2 before weighting (A), after weighting with CACNA2D3 as the outcome (B) and after weighting with HITRA1 as the outcome (C). Note. The circle represents the intervention strategy. The triangle represents the control strategy. The dashed lines denote ±0.25 SD.

### Effects of long-term adherence to the PA guidelines on frailty-associated proteins

For both the ICE and IPW methods, we found small differences in the population-level average for the 45 frailty-associated proteins comparing all participants achieving and maintaining ≥150 minutes/week of MVPA at Visits 1 and 3 to the “natural course” strategy (ranged from 0.04 to 0.11 SD, Figure 6 and Table S2). The largest increase in the population-level average was observed with voltage-dependent calcium channel subunit alpha-2/delta-3 (CACNA2D3; difference_ICE_ = 0.10 SD, 95% CI: 0.05, 0.14; difference_IPW_ = 0.11 SD, 95% CI: 0.05, 0.15). The largest decrease in the population-level average was observed with high-temperature requirement serine protease A1 (HTRA1; difference_ICE_ = −0.09 SD, 95% CI: −0.13, - 0.05; difference_IPW_ = −0.08, 95% CI: −0.12, −0.04). Other proteins with larger increased population-level averages included neurogenic locus notch homolog protein 1 (NOTCH1), neural cell adhesion molecule 1 (NCAM1), and contactin-1 (CNTN1). Other proteins with larger decreased population-level averages included C-reactive protein (CRP), transmembrane protein 132D (TMEM132D), and vesicle-fusing ATPase (NSF). All these proteins had 95% CIs not overlapping with zero.

**Figure 6.**
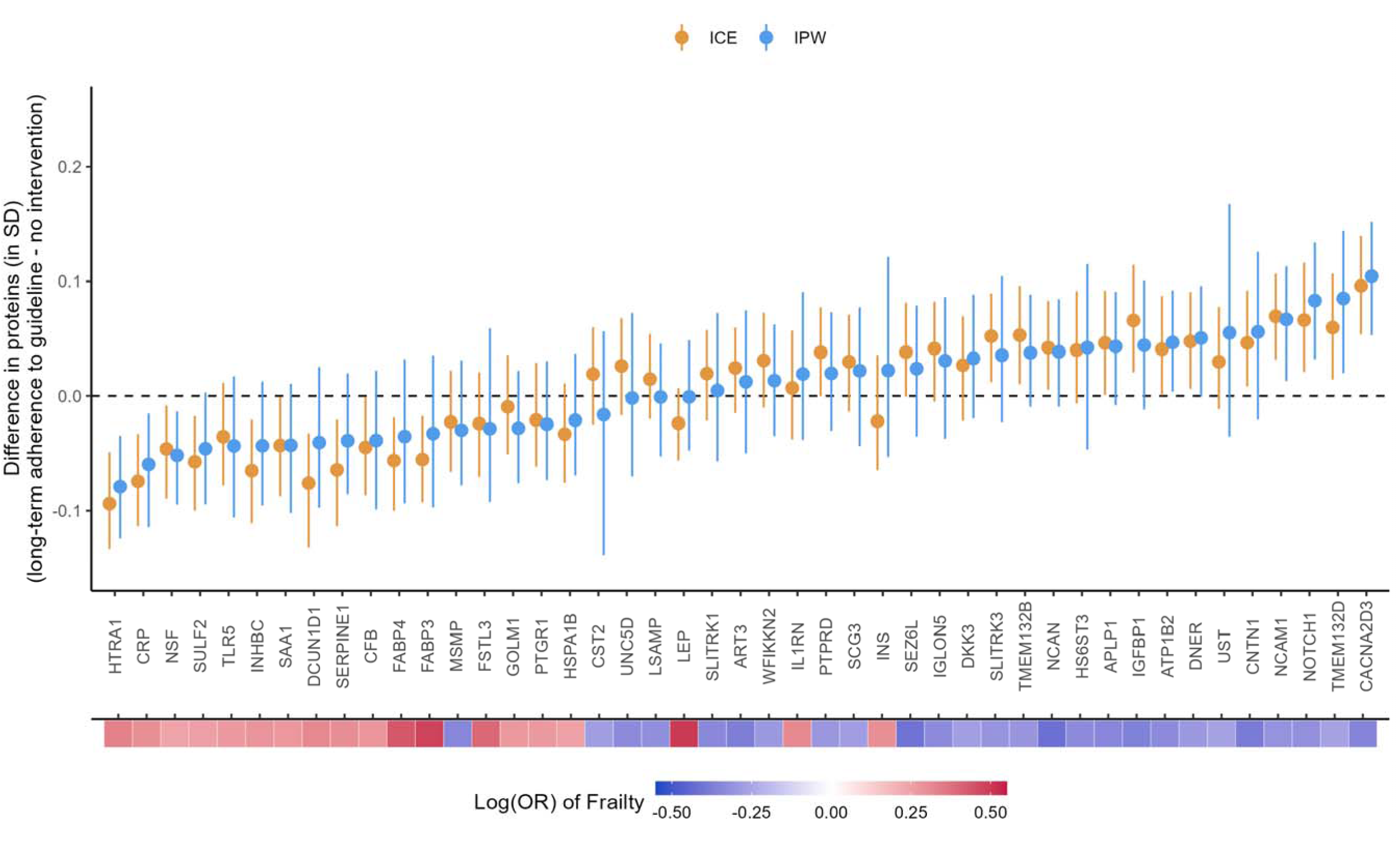
Causal effects of long-term adherence to physical activity guideline on proteins associated with frailty. Long-term adherence to physical activity guideline was measured by achieving ≥150 minutes/week of MVPA at Visit 1 and Visit 3. The heat map at the bottom depicts the strength of associations between the proteins and frailty.

The effect estimates from the ICE and IPW methods mostly agree with wider CIs from the IPW method. A few proteins with smaller effect sizes had larger discrepancies between the method but the CIs were all consistent with null effects (e.g., leptin [LEP, difference_IPW_ = −0.001, 95% CI: −0.05, 0.05; difference_ICE_ = −0.02, 95% CI: −0.06, 0.01]; insulin [INS; difference_IPW_ = 0.02, 95% CI: −0.05, 0.12; difference_ICE_ = −0.02, 95% CI: −0.06, 0.04); and cystatin-SA [CST2; difference_IPW_ = −0.02, 95% CI: −0.14, 0.06; difference_ICE_ = 0.02, 95% CI: −0.03, 0.06]).

The directions of the effects of PA on the frailty-associated proteins were mostly consistent with the known directions of associations between the proteins and frailty and the known benefits of PA on frailty. For example, a higher level of CACNA2D3 was associated with a lower risk of frailty in the previous study (i.e., protective, Figure 6), and meeting the PA guidelines at both Visits 1 and 3 increased the population level of CACNA2D3. The two exceptions were prostate-associated microseminoprotein (MSMP) and interleukin-1 receptor antagonist protein (IL1RN). However, both had 95% CI overlapping with zero, consistent with null effects.

### Sensitivity analysis: balance of individual chronic conditions, drinking status, family income, and histories of binary and categorical confound

The balance of individual chronic conditions, drinking status, family income, and lagged values of the binary and categorical confounder were improved after weighting, even though these variables were not included in the IPW models (Figures 7 and 8, S5, and S6). For all protein outcomes, after applying the IPW final weights, SMDs of most covariates were within ±0.1 SD (i) comparing participants who achieved ≥150 minutes/week of MVPA and those who did not, both at Visit 1 and at Visit 3 (conditional on MVPA ≥150 minutes/week at Visit 1 and no prior censoring) and (ii) comparing participants who were censored to those who remained under study, both at Visit 1 and at Visit 2 (conditional on MVPA ≥150 minutes/week at Visit 1 and no prior censoring). The exceptions were drinking status and family income for (i) and family income, stroke, and lung disease for (ii), but all had SMS within ±0.2 SD.

**Figure 7.**
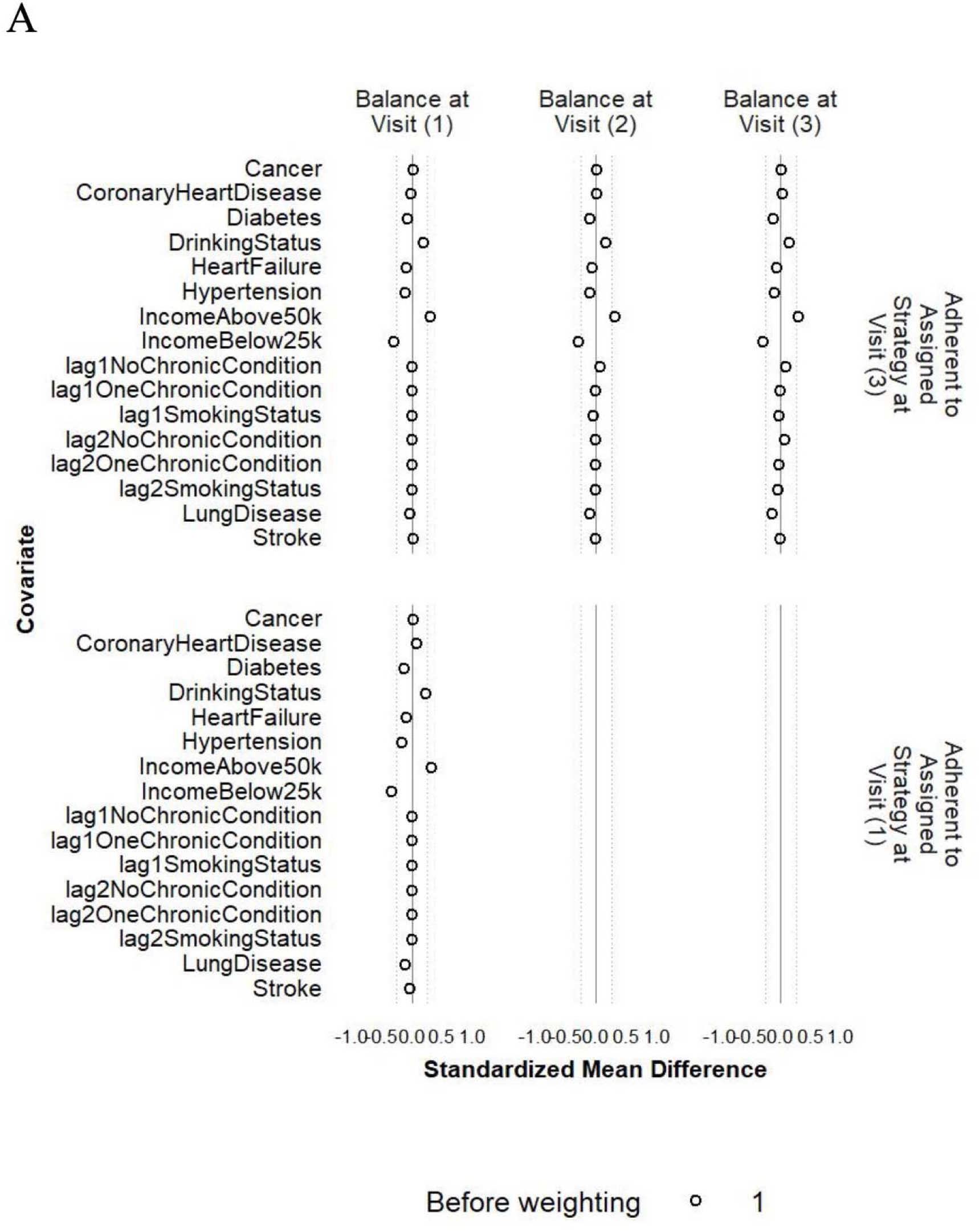
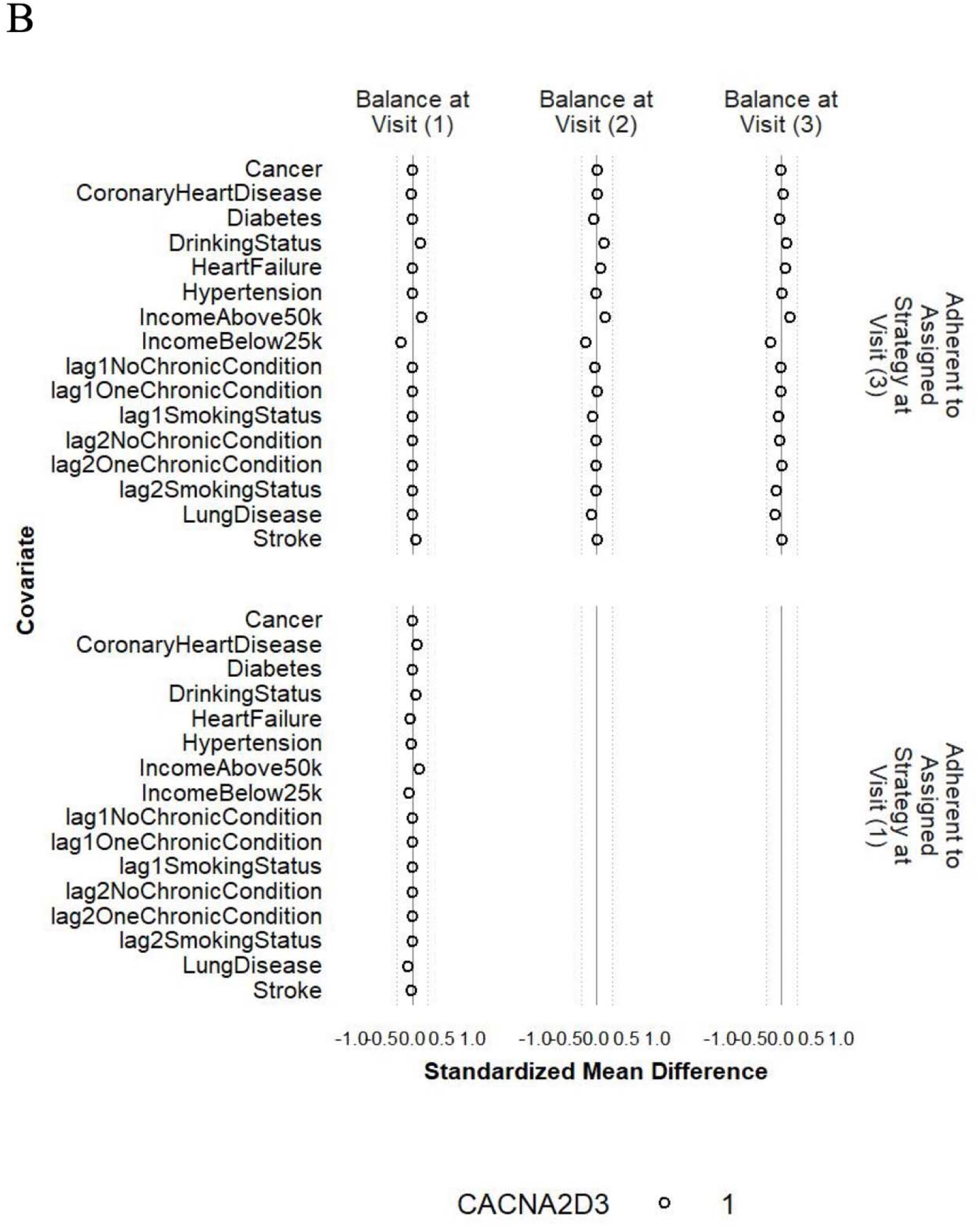
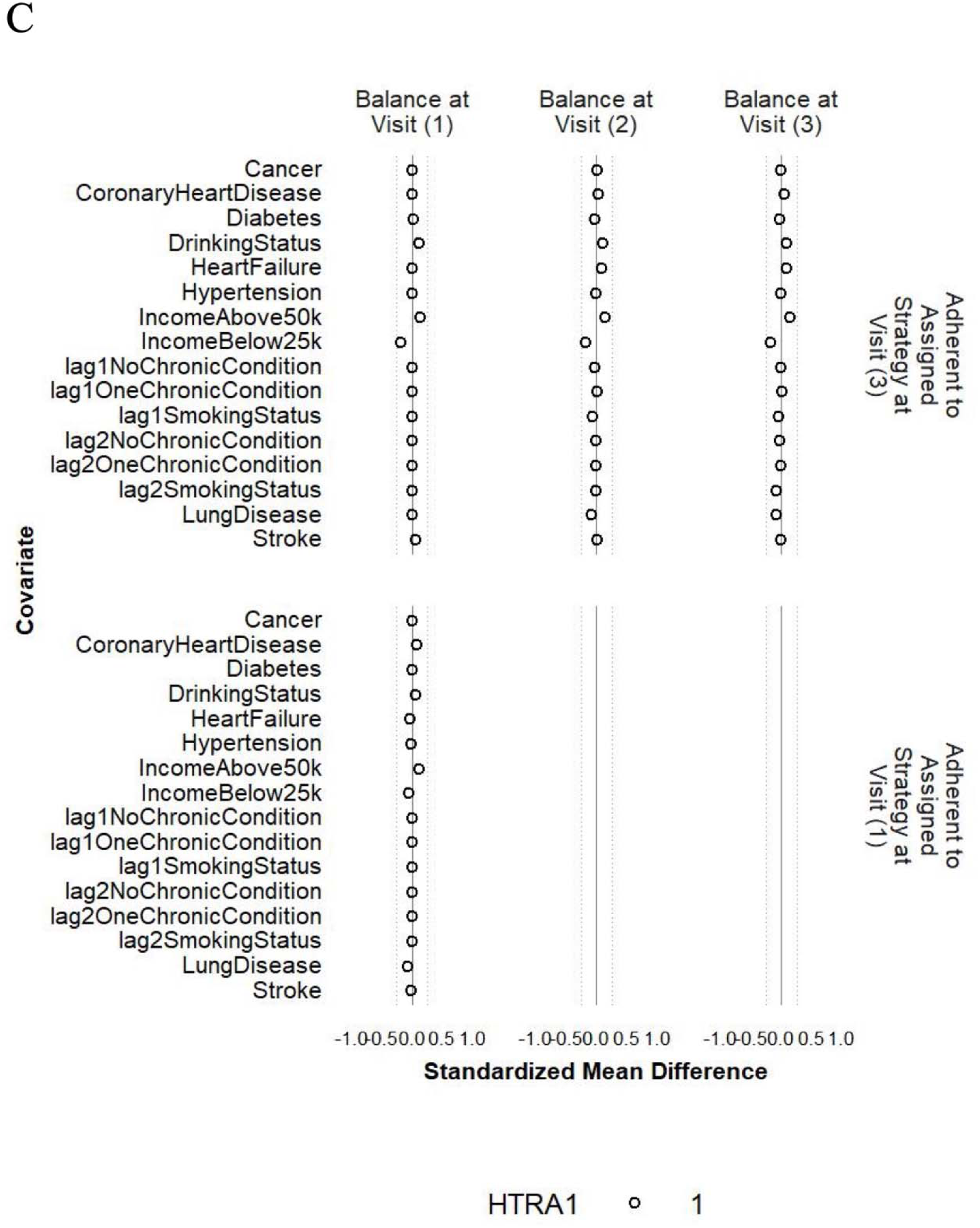
Balance of individual chronic conditions, drinking status, family income, and histories of smoking status and number of chronic conditions between participants who achieved ≥150 minutes/week of MVPA and those who did not at Visit 1 and Visit 3 before weighting (A), after weighting with CACNA2D3 as the outcome (B) and after weighting with HITRA1 as the outcome (C). Note. The circle represents the intervention strategy. The dashed lines denote 0.25 SD.

**Figure 8.**
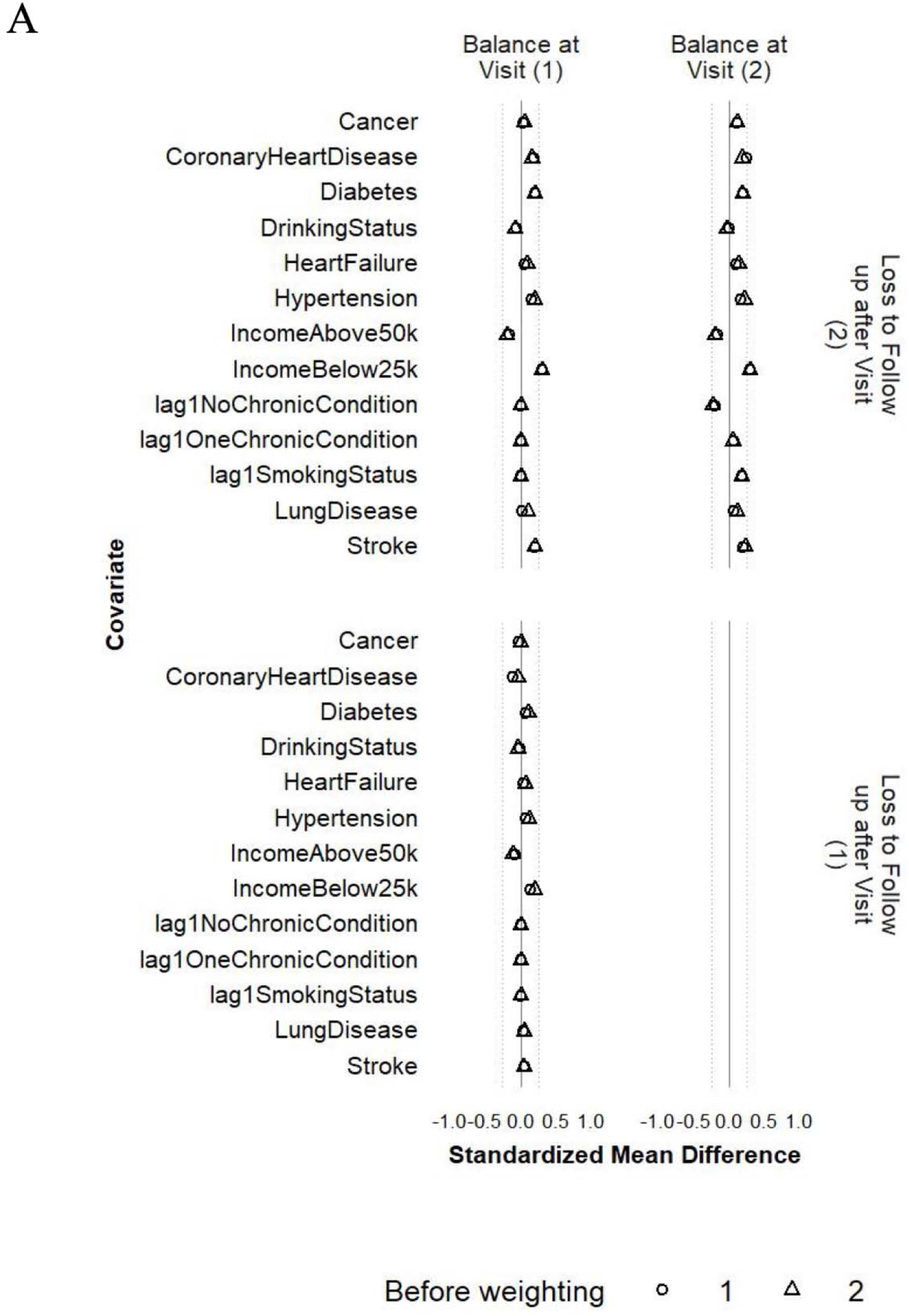
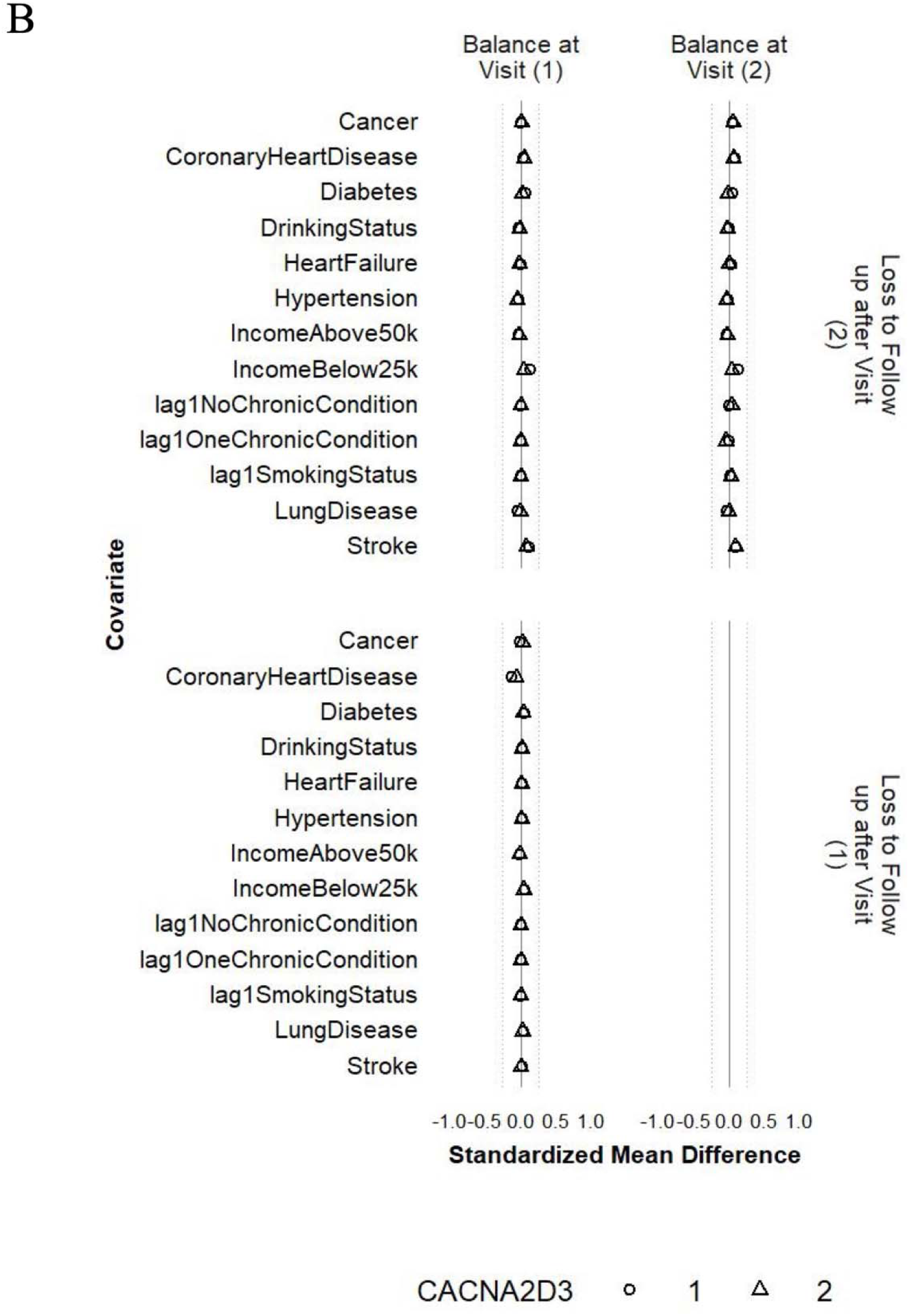
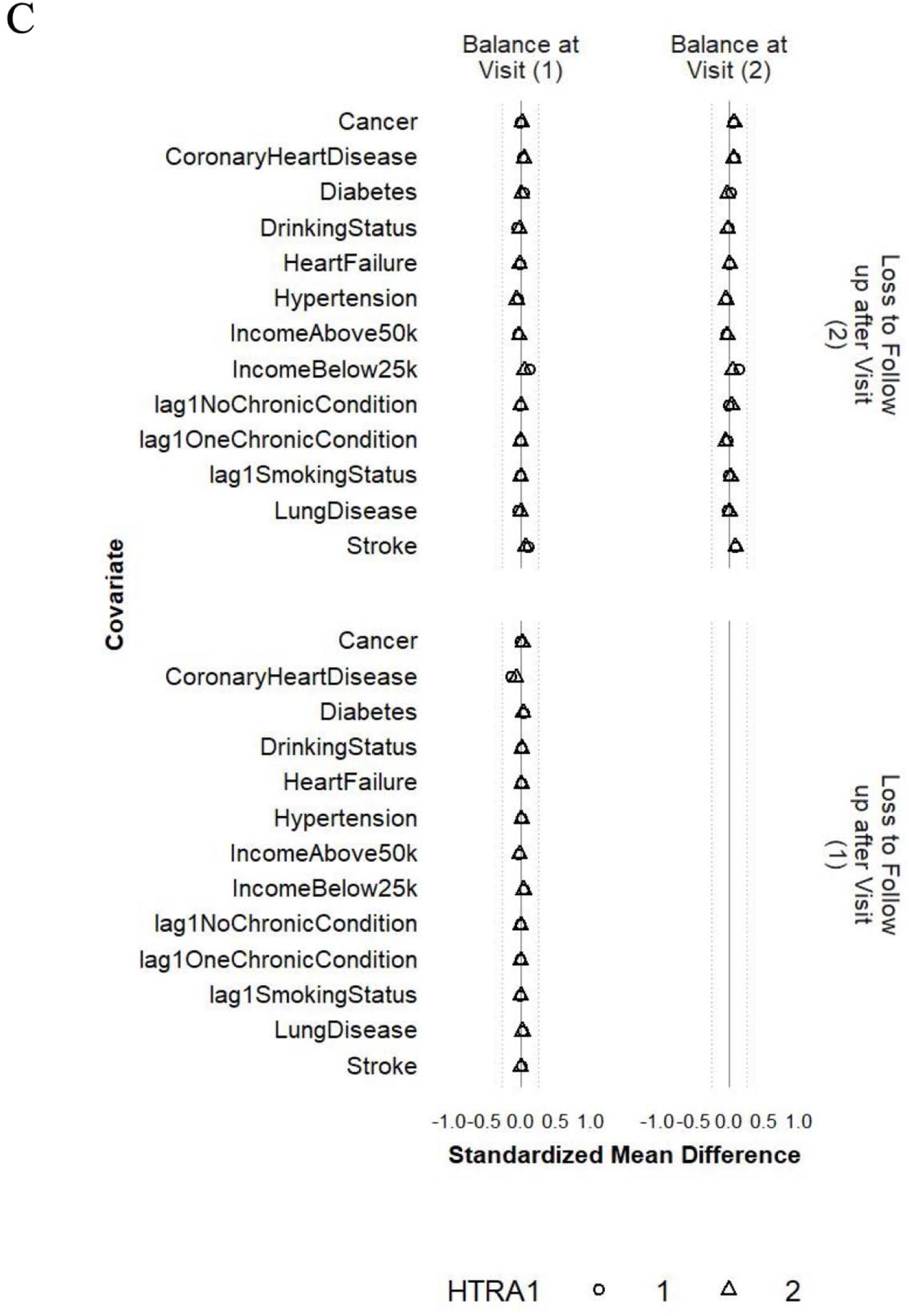
Balance of individual chronic conditions, drinking status, family income, and histories of smoking status and number of chronic conditions between participants who were lost and those who remained under study at Visits 1 and 2 before weighting (A), after weighting with CACNA2D3 as the outcome (B) and after weighting with HITRA1 as the outcome (C). The circle represents the intervention strategy. Note. The triangle represents the control strategy. The dashed lines denote ±0.25 SD.

### Sensitivity analysis: excluding participants with major chronic conditions at Visit 1

Excluding participants with major chronic conditions at Visit 1 produced mostly consistent results with the main results using either method (Figure 9). The effects for some proteins were slightly attenuated, e.g., CACNA2D3 and HTRA1. However, the effects for other proteins were strengthened using the IPW method, e.g., insulin-like growth factor-binding protein 1 (IGFBP1), amyloid-like protein 1 (APLP1), transmembrane protein 132B (TMEM132B), and delta and Notch-like epidermal growth factor-related receptor (DNER). CST2 and INS now had more similar effect estimates and the same effect directions between the ICE and IPW methods. For the IPW method, good balance (SMDs within ±0.1 SD) was achieved for all the included covariates for all protein outcomes (Figures S7-S8 for CACNA2D3 and HTRA1; plots for other proteins not shown). Most excluded covariates achieved good balance (SMDs within ±0.1 SD) in the sensitivity analysis sample. Other excluded covariates (drinking status, family income, lung disease, stroke, coronary heart disease, and cancer) achieved satisfactory balance (SMDs within ±0.25 SD, Figures S9-S10 for CACNA2D3 and HTRA1; plots for other proteins not shown).

**Figure 9.**
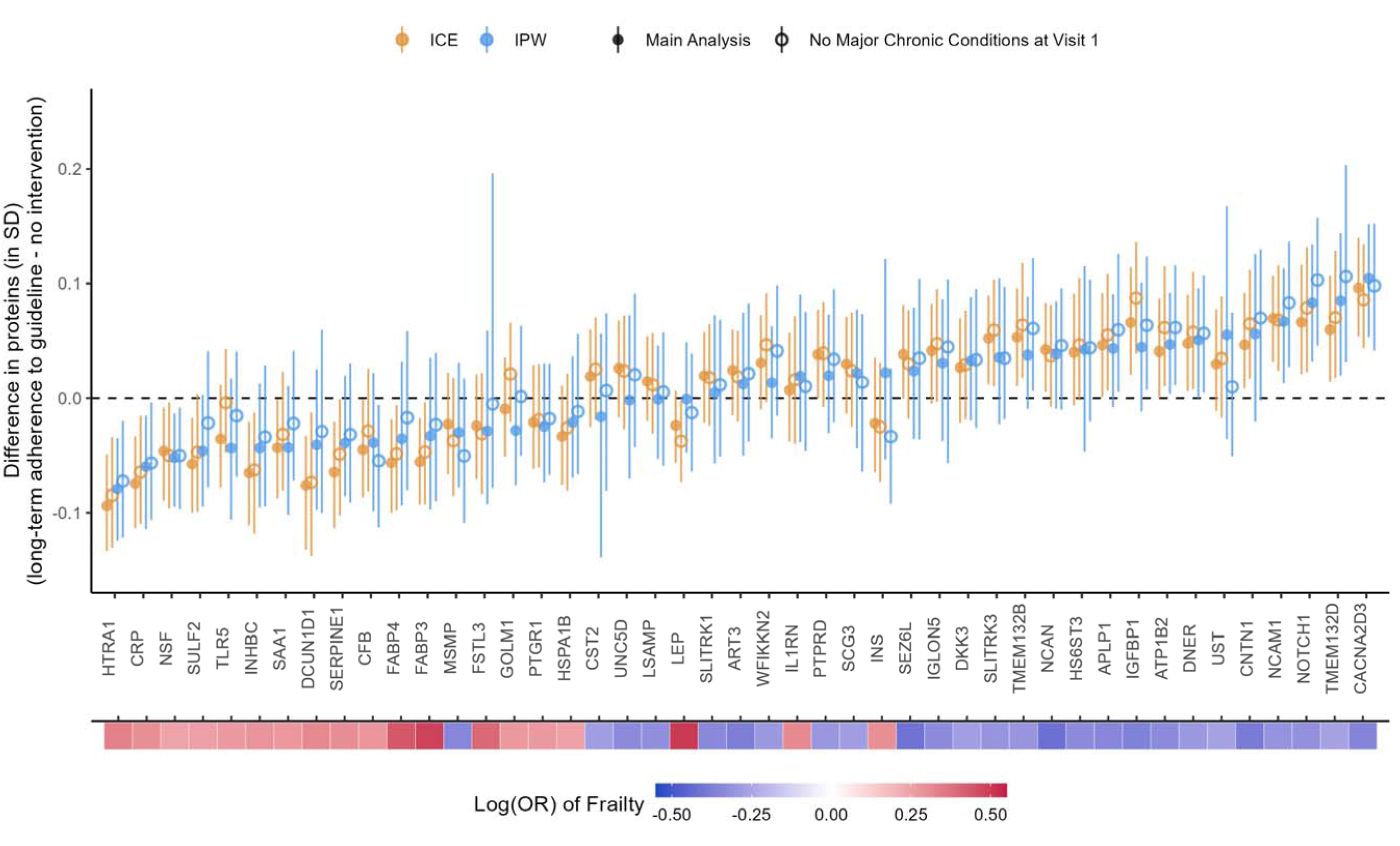
Comparison between main analysis results and sensitivity analysis excluding participants with major chronic conditions at Visit 1. The heat map at the bottom depicts the strength of associations between the proteins and frailty.

### Sensitivity analysis: tipping analysis for the effects of Visit 1 MVPA on Visit 3 proteins

The tipping points for the effect of MVPA minutes/week at Visit 1 on the levels of CNCNA2D3 and HTRA1 at Visit 3 are presented in Figure 10. An unmeasured confounder, with similar confounding strength as Visit 2 measures of BMI or eGFR on *Visit 3* MVPA and Visit 3 CNCNA2D3/HTRA1, would *not* bring the effect of *Visit 1* MVPA on these two proteins at Visit 3 to the null. However, an unmeasured confounder, with similar confounding strength as a Visit 2 measure of CNCNA2D3 on *Visit 3* MVPA and Visit 3 CNCNA2D3 level, would bring the effect of *Visit 1* MVPA on Visit 3 CNCNA2D3 to the null. An unmeasured confounder, with similar confounding strength as a Visit 2 measure of HTRA1 on Visit 3 MVPA and Visit 3 HTRA1 level, would bring the effect of MVPA minutes/week at Visit 1 on HTRA1 at Visit 3 to the null. Similar patterns were observed for all other proteins (Figure S11).

**Figure 10.**
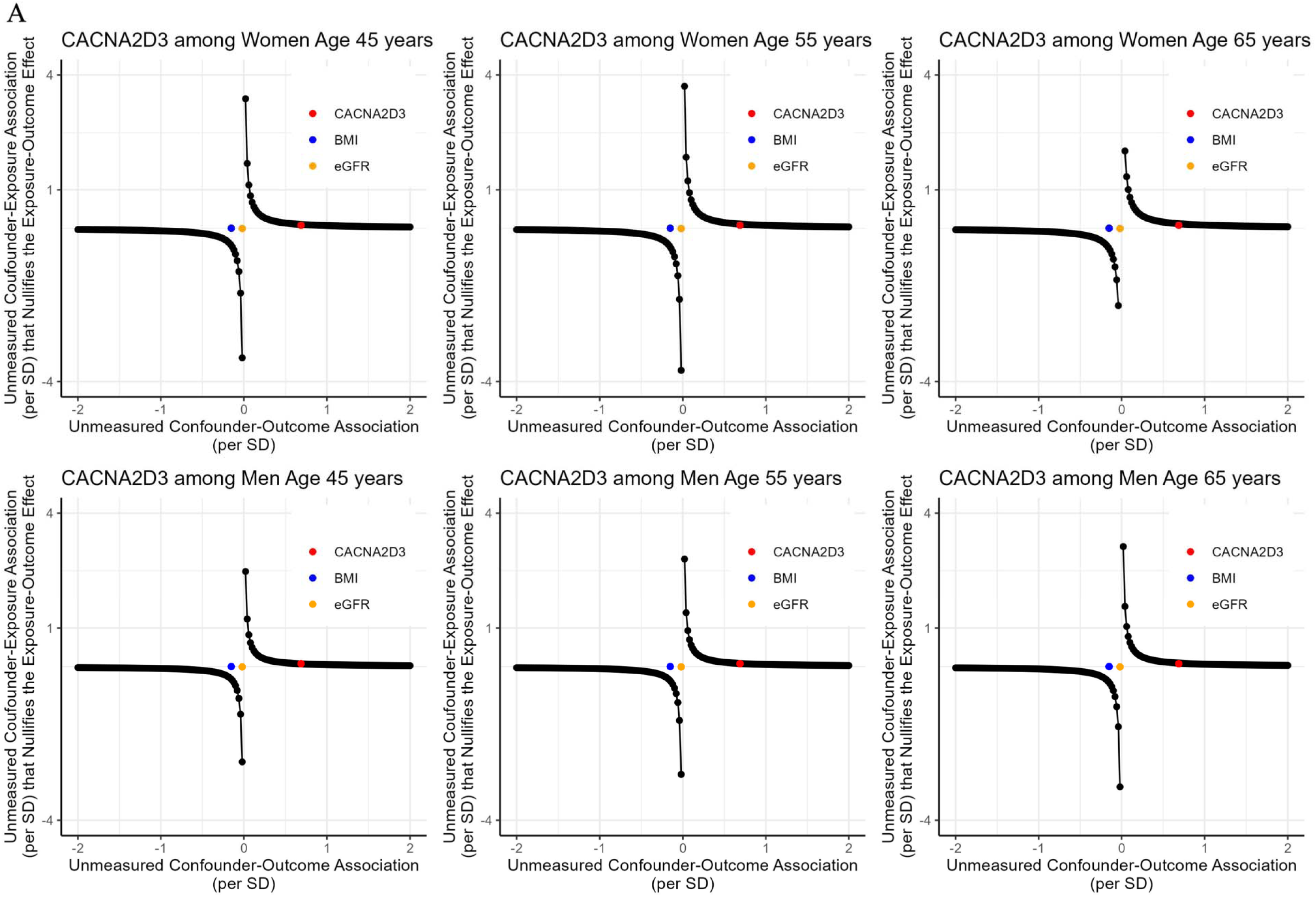
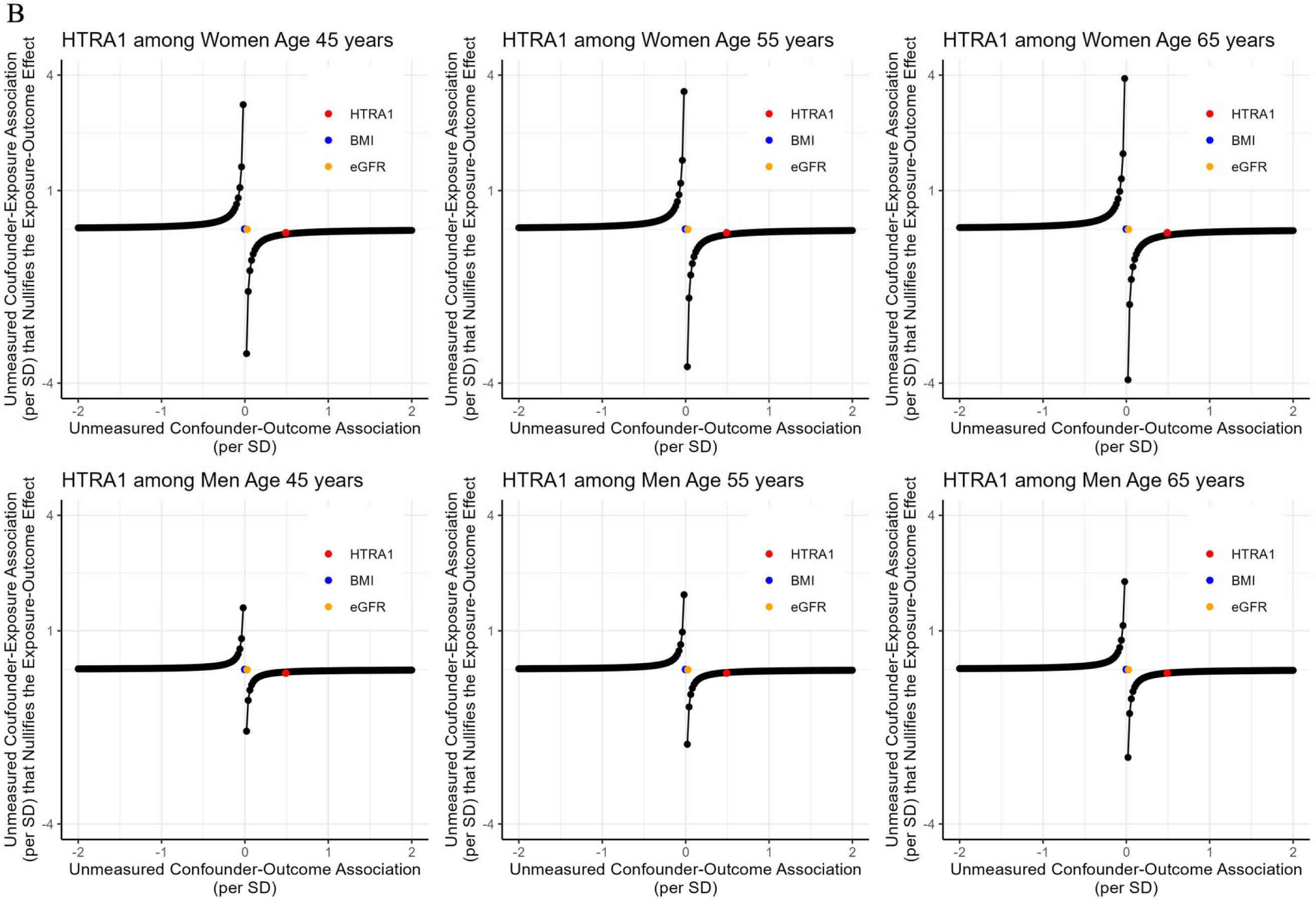
The tipping points of an unmeasured confounder for the effect of MVPA at Visit 1 on CNCNA2D3 (A) and HTRA1 (B) at Visit 3 Notes. CNCNA2D3 = voltage-dependent calcium channel subunit alpha-2/delta-3; HTRA1 = high-temperature requirement serine protease A1; BMI = body mass index, eGFR = estimated glomerular filtration rate

### Sensitivity analysis: excluding participants who died before Visit 3

The effect estimates after excluding participants who died before Visit 3 were close to the main estimates with small attenuations for some proteins e.g., CACNA3D3 and CRP (Figure 11). For the IPW method, good balance (SMDs within ±0.1 SD) was achieved for the included covariates for all protein outcomes. Good balance (SMDs within ±0.1 SD) was also achieved for all excluded individual chronic conditions except those for coronary heart disease. Coronary heart disease, drinking status, and family income had SMDs within ±0.2 SD (Figures S12-S15 for CACNA2D3 and HTRA1; plots for other proteins not shown).

**Figure 11.**
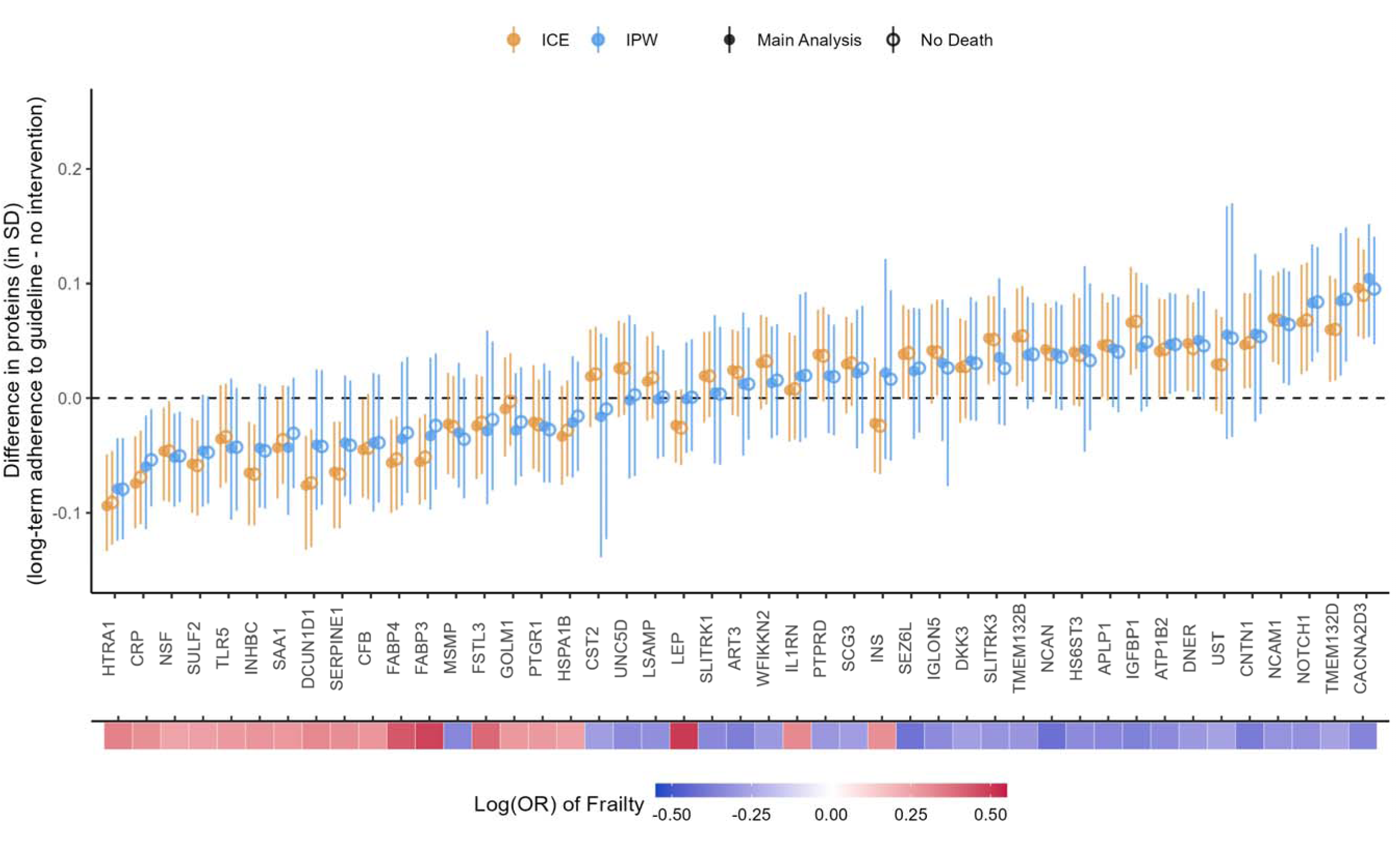
Comparison between main analysis results and sensitivity analysis excluding participants who died before Visit 3. The heat map at the bottom depicts the strength of associations between the proteins and frailty.

## Discussion

We used a target trial emulation design and the ICE and IPW methods to estimate the effects of adhering to the US CDC PA guidelines (i.e., ≥150 minutes/week of MVPA) in midlife on 45 plasma proteins previously shown to be associated with frailty in later life.^27^ We found that achieving and maintaining ≥150 minutes/week of MVPA at Visit 1 and Visit 3 in ARIC, which we assumed to represent long-term adherence to the recommended PA level in midlife, had beneficial effects on the population-level averages of many frailty-associated proteins. Our findings expand on previous studies, which focused on either the effects of short-term structured exercise programs on proteins or cross-sectional associations between MVPA and proteins, by showing that long-term, less structured MVPA during midlife may improve health on a molecular level by altering a segment of the circulating proteome. Specifically, our results suggest that MVPA during midlife alters the abundance of a set of proteins and biological pathways that may play an early role in subsequent frailty development.^27^

Our findings on some proteins were consistent with changes after short-term structured exercise programs documented in previous work. For example, IGFBP1, a protein that enhances glucose uptake in the periphery tissues^51^, showed in our analysis a higher population level under the long-term adherence to PA guidelines in our analysis. In previous studies, IGFBP1 was also significantly increased after 20 weeks of endurance exercise,^16^ and higher leisure-time PA was cross-sectionally associated with a higher level of IGFBP1.^22^ We also found that the population level of contactin-1 (CNTN1), a cell adhesion molecule that protects muscle strength and mobility,^52,53^ was higher under the long-term adherence to PA guidelines, consistent with the increased level after a 3-month aerobic exercise program (though not statistically significant).^18^ Our results enhance previous findings by providing longitudinal evidence in an unstructured setting more reflective of free-living PA.

Previous studies had mixed findings of the effects of structured exercise programs on CRP, a pro-inflammatory cytokine, the sustained high level of which in mid-to-late life has been associated with slower gait speed in late life,^54^ and NCAM1, the deficiency of which has been implicated in in memory and cognitive impairment.^55^ Robbins and colleagues found that the level of CRP increased, and the level of NCAM1 decreased in plasma after 20 weeks of endurance exercise.^16^ However, a 3-month aerobic exercise program saw decreased CRP and increased NCAM1 in plasma.^18^ Our findings on these two proteins were consistent with the second study. Such discrepancies could reflect the diverging effects of different PA/exercise types in which people chose to engage. It may also reflect the diverging effects of acute exercise and regular exercise, as acute exercise elevates CRP with a peak increase after up to 28 hours,^56^ but regular exercise reduces CRP.^57^ More research on exercise type, intensity, and duration is needed to further disentangle these results.

FABP3 and FABP4, fatty-acid proteins of which higher circulation levels have been linked to muscular dystrophy, metabolic conditions, and cardiovascular disease,^22,58,59^ were surprisingly found to be elevated in plasma after 20 weeks of endurance exercise.^16^ Additionally, FABP3 was also elevated in plasma after 3 months of aerobic exercise and in thigh muscle after 20 weeks of resistance training among older adults (but reduced in young adults [25.0±1.1 years]).^17,18^ In our analysis, both proteins were reduced with long-term adherence to PA guidelines, which were more consistent with the current knowledge of circulating FABP3 and FABP4 with health and diseases. Particularly, FABP3 is known to be released into circulation as a result of tissue injury.^58,59^ Therefore, the elevated plasma level of FABP3 after exercise programs found in previous studies could reflect exercise-induced muscle injury.^60^ Our findings on FABP4 are supported by one observational study that reported reduced FABP4 in plasma with increased PA over 1-year follow-up.^61^ Taken together, these discrepant findings highlight the need for studies on habitual PA beyond structured exercise programs.

We found beneficial effects of long-term adherence to PA guidelines on several novel proteins that have not been examined in previous studies, including the two proteins with the largest effect sizes, CACNA2D3 and HTRA1. CACNA2D3 is a protein in the voltage-dependent calcium channel that modulates synaptic transmission.^62^ Disruption in the *CACNA2D3* gene has been implicated in Alzheimer’s disease.^62^ In previous work, we have found that lower levels of CACNA2D3 measured in middle and older age were associated with a higher risk of frailty.^25,27^ HTRA1 is a serine protease that breaks down extracellular matrix proteins and inhibits the anti-inflammatory function of transforming growth factor-β (TGF-β) proteins.^63,64^ Higher HTRA1 has been shown to be associated with frailty and poor musculoskeletal health.^63,64^ The higher level of CACNA2D3 and lower level of HTRA1 with long-term adherence to PA guidelines found in this analysis were congruent with the current understanding of the functions of these two proteins. Supporting our findings, one study reported a lower methylation of the *CACNA2D3* gene (which can result in higher expression of CACNA2D3 protein) was found with more PA.^65^ Future studies are needed to replicate our results and further investigate the effects of PA on these proteins.

The ICE and IPW produced largely consistent effect directions except for a few proteins with small effect sizes and 95% CI consistent with null effects. Moreover, covariates balance was good after weighting in the IPW method, suggesting good model specifications. Taken together, these results provide confidence to our findings. Proteins that were not substantially altered by long-term adherence to PA guidelines in this analysis may be targets for other types of interventions. Combining these other interventions with PA in daily living may enhance our ability to reduce frailty risk in later life.

Our sensitivity analyses also suggested robustness of our findings to residual confounding. We found good or satisfactory balances of the covariates not included in the ICE and IPW models. We also found similar effects after excluding participants with major chronic conditions. The tipping analysis suggested that only an unmeasured confounding as strong as Visit 2 protein levels on Visit 3 MVPA and Visit 3 protein levels would explain away the effects we found in the main analysis. However, such an unmeasured confounder may not be plausible. Even for protein levels at or prior to Visit 1, their associations with the outcome, i.e., protein levels at Visit 3, would be weaker than the associations between protein levels at Visit 2 and the outcome because of a longer time between the two Visit 1 and Visit 3 than between Visit 2 and Visit 3.

Our findings were robust to the left truncation by death as suggested by the similar estimates from the sensitivity analysis excluding participants who died before Visit 3. This was likely due to the small number of deaths in our sample (n = 794, 5.3%). Future studies should examine the robustness of our findings using more advanced methods discussed in Supplemental Methods.

This study has several limitations, the first set of which are results of the ARIC study design: (1) we used only two measurements of PA to determine the adherence to PA guidelines over 6 years of follow-up, which may not have fully captured the true adherence; (2) with the scarce measurements, we did not lag the confounders and PA, but instead assumed that the measurements of confounders preceded PA and proteins measurements at the same visit; (3) we included different depths of histories for continuous confounders at different visits (e.g., BMI at Visits 1-3 was included in the ICE model at Visit 3, but only BMI at Visit 1 was included in the baseline model). Future studies should use an alternative dataset to address these limitations. Second, the self-reported PA in ARIC presents major challenges with measurement error. Regular participation in MVPA is easier to recall and sporadic MVPA may not be captured as accurately, leading to imperfect measurement of MVPA.^66^ Moreover, the weekly level was estimated using recalled frequency and duration which may not be accurate. Accelerometer-measured PA can address these challenges because it captures a more complete assessment of daily movement. However, accelerometers usually do not specify the types of PA. Therefore, an ideal study to replicate our results would combine accelerometer data with information about the types of PA (e.g., from a PA diary). Lastly, we focused on MVPA, which is consistent with the physical activity guidelines. However, evidence exists that light and non-exercise PA have health benefits.^67^ Even for MVPA, other contrasts besides adherence to guidelines, e.g., dose-response effects of increasing minutes/week of MVPA, may be relevant to clinical practice. Future studies are needed to investigate the biological implications of different strategies that target different aspects of PA to more comprehensively understand the health benefits of PA.

We also identified several directions for future research both in the area of PA and biological markers. First, we focused on the effects of PA on individual frailty-associated proteins. Future studies examining the cumulative effects of multiple proteins may further advance our understanding of the benefits of PA on biological pathways relevant to frailty prevention or treatment. Second, clinical meaningful differences for proteins measured by the SomaScan platform have not been established. With increasing research using this platform and other proteomic platforms (e.g., Olink), understanding the meaningful clinical differences will greatly contribute to future research that investigates interventional strategies to improve biological processes underlying age-related conditions. Third, our effect estimates allowed individuals to have different MVPA minutes/week within the range of the strategies. Future studies may explore other aspects of PA, such as the type of MVPA (e.g., jogging [aerobic] versus weightlifting [resistance]) or how the above-guideline MVPA was achieved (e.g., 30 minutes/session for 5 sessions per week versus shorter durations per session and more frequent sessions) together with the duration to provide a more comprehensive PA profile of an individual. Lastly, though we performed extensive checks and sensitivity analyses to support the assumptions of exchangeability, positivity, and no model misspecification, violations of these assumptions in observational data are still possible. Future clinical trials on free-living PA that collect blood-based proteins are needed to further strengthen the evidence of the biological basis of free-living PA as an invention strategy for frailty.

Despite the limitations, our study provides a first line of evidence that achieving and maintaining MVPA levels above the recommended threshold (150 minutes/week) throughout midlife has beneficial effects on the population average of multiple proteins previously associated with frailty risk in later life. Many of these proteins had not been examined in previous studies of exercise programs or self-report MVPA and could be novel targets for interventions. Specifically, our results highlighted several proteins involved in the nervous system (CACNA2D3, CNTN1, and NCAM1) and inflammation (HTRA1 and CRP) that should be further investigated to understand their role in mediating the beneficial effects of PA on frailty. Moreover, proteins that were minimally affected by long-term adherence to PA guidelines in our study (e.g., LEP) inform future studies to explore other forms of intervention (e.g., dietary intervention) that can affect these proteins. Such interventions can be integrated with PA to improve the treatment and prevention of frailty. Lastly, as we gain further knowledge on the meaningful differences of the proteins, our results will inform whether more resources should be spent to encourage long-term adherence to PA guidelines in the middle-aged population.

## Declarations

### Author contributions

F.L, J.A.S, and J.W.J contributed to the study conception and design. F.L., K.A.W, and P.P contributed to the data acquisition. F.L. analyzes the data. F.L, J.A.S, and J.W.J contributed to the interpretation of the results. All co-authors provided substantial revisions to the manuscript.

### Ethics approval

ARIC study protocols were approved by institutional review boards at each participating center: University of North Carolina at Chapel Hill, Chapel Hill, NC; Wake Forest University, Winston-Salem, NC; Johns Hopkins University, Baltimore, MD; University of Minnesota, Minneapolis, MN; and University of Mississippi Medical Center, Jackson, MS.

### Consent to participate

All participants gave written informed consent at each study visit and proxies provided consent for participants who were judged to lack capacity.

### Data availability statement

Pre-existing data access policies for the ARIC study specify that research data requests can be submitted to each steering committee; these will be promptly reviewed for confidentiality or intellectual property restrictions and will not unreasonably be refused. Please refer to the data sharing policies of these studies. Individual level patient or protein data may further be restricted by consent, confidentiality or privacy laws/considerations. These policies apply to both clinical and proteomic data.

