## Supplemental Methods for "The effect of long-term adherence to physical activity recommendations in midlife on plasma proteins associated with frailty in the Atherosclerosis Risk in Communities (ARIC) study"

*Deviations of the emulated trials from the target trial*

The emulated trial had three major deviations from the target trial. First, functional limitations potentially preventing PA were not directly measured. Though some studies have used diseases such as myocardial infarction and stroke and difficulty climbing stairs or walking blocks as a surrogate of these exclusion criteria,^1^ the CDC guidelines do not exclude individuals with chronic conditions or mobility disability from their recommendations.^2^ Therefore, we allowed everyone in the study sample to be eligible for the PA intervention. Second, though the target trial would not exclude any participants based on race, we excluded participants who were neither Black nor White due to their extremely small number (n = 49, Figure 1A), to avoid introducing data sparsity bias (e.g., non-positivity).^3,4^ Lastly, although a target trial would probably measure MVPA frequently (e.g., self-report weekly) or even continuously (e.g. via accelerometers), MVPA was measured in ARIC using self-report at baseline (Visit 1) and once during the follow-up (Visit 3). With the unavailable measurements of MVPA between Visit 1 and Visit 3, we assumed that participants who met the guideline at both visits also met the guideline during the intervening period.

*Notations*

We use the following notations for the g-formula and the IPW and ICE procedures. We use *t* = 1, 2, or 3 to denote the study visit. At each visit, *K_t_* denotes the minutes of moderate-to-vigorous physical activity (MVPA). *A_t_ =* 1 indicates that the recommended ≥150 minutes/week of MVPA is achieved, and 0 otherwise. *A_t_* is a coarsening variable of *K_t_*. *Y_t_* denotes the plasma protein level. *L_t_* denotes the confounders including smoking status, number of chronic conditions (0, 1, or ≥2), body mass index (BMI), estimated glomerular filtration rate (eGFR), and total cholesterol. *L_1_* also includes time-fixed confounding of age, sex, race, and education. *C_t_* denotes censoring after visit *t*. We use the overbar to denote the history of a variable up to a *t*, e.g., $\bar{A}_{t}=\left\{ A_{1},\ldots, A_{t} \right\}$. By definition, variables with subscript of −1 are empty. We use the superscripts to denote potential values of variables that may be contrary to the observed values. For example, $Y_{3}^{\bar{K}_{3}=\bar{k}_{3}, \bar{A}_{3}=\bar{1},\bar{C}_{2}=\bar{0}}$ is the potential outcome of the protein level at Visit 3 if a participant had met the recommended ≥150 minutes/week of MVPA at all visits (i.e., $\bar{A}_{3}=\bar{1}$) with specific minutes of MVPA at each visit (i.e., $\bar{K}_{3}=\bar{k}_{3}$) and had not been censored during follow-up (i.e., $\bar{C}_{2}=\bar{0}$).

*The identifying assumptions and the g-formula*

Based on Figure 2, the exchangeability assumptions for identifying (i.e., consistently estimating) the causal effect of adhering to the recommended ≥150 minutes/week of MVPA at Visit 1 and Visit 3 on the protein level at Visit 3 are^5^:

$Y_{3}^{\bar{K}_{3}=\bar{k}_{3}, \bar{A}_{3}=\bar{1},\bar{C}_{2}=\bar{0}}\perp K_{t}|\bar{L}_{t}=\bar{l}_{t}, \bar{K}_{t-1}=\bar{k}_{t-1},\bar{A}_{t-1}=\bar{1}, \bar{C}_{t-1}=\bar{0}, \bar{Y}_{t-1}=\bar{y}_{t-1}$ for$t=1, 3$

$Y_{3}^{\bar{K}_{3}=\bar{k}_{3}, \bar{A}_{3}=\bar{1},\bar{C}_{2}=\bar{0}}\perp C_{t}|\bar{L}_{t}=\bar{l}_{t},\bar{K}_{t}=\bar{k}_{t}, \bar{A}_{t}=\bar{1}, \bar{C}_{t-1}=\bar{0},\bar{Y}_{t}=\bar{y}_{t})$ for $t=1, 2$

*t* only takes the value of 1 or 3 for the first expression because MVPA was only measured at those two visits. Here, we assumed that the measurements of MVPA (*K_t_*) preceded protein measurements at the same visit (*Y_t_*) because *K_t_* was measured by a physical activity questionnaire that asked participants to report their sports and exercise in the past year at a study visit, whereas proteins were measured using blood samples collected at study visits. For simplicity, we further assumed that the measurements of confounders (*L_t_*) preceded MVPA measurement at the same visit (*K_t_*). Though some confounders were derived solely using measurements at the study visits, e.g., hypertension, diabetes, BMI, eGFR, and cholesterol, these measurements may represent long-term health conditions that preceded the MVPA measurement. All these variables were selected because they have been shown to be associated with protein levels^6^ , and they may affect MVPA levels.

The positivity assumptions are:

$f\left( K_{t} | \bar{L}_{t}=\bar{l}_{t}, \bar{K}_{t-1}=\bar{k}_{t-1},\bar{A}_{t}=\bar{1}, \bar{C}_{t-1}=\bar{0}, \bar{Y}_{t-1}=\bar{y}_{t-1} \right)>0$w.p.1 for $t=1, 3$

$P\left( C_{t}=0 | \bar{L}_{t}=\bar{l}_{t},\bar{K}_{t}=\bar{k}_{t}, \bar{A}_{t}=\bar{1}, \bar{C}_{t-1}=\bar{0},\bar{Y}_{t}=\bar{y}_{t} \right)>0$ w.p.1 for $t=1, 2$

where w.p.1 means with probability of 1.^5^ We checked the positivity assumption among categorical covariates using the UpSet plots (Appendix Figures 1-4). As shown in the plots, for participants who did not meet the guidelines at each visit (i.e., the “nonadherent” dot is solid in Figures 1-2) or who were lost to follow-up (i.e., the “censored” dot is solid in Figures 3-4), there were always participants who shared the same values of the categorical covariates and met the guidelines or remained in the study (i.e., the next column has the same dot pattern for the covariates but the “nonadherent”/”censored” dot is blank whereas the “adherent”/” uncensored” dot is solid).

The consistency assumption is: if observed $\bar{K}_{t}=\bar{k}_{t}$that satisfied $\bar{A}_{t}= \bar{1}$, then $Y_{t}=Y_{t}^{\bar{K}_{t}=\bar{k}_{t}, \bar{A}_{t}=\bar{1}}$ and $L_{t+1}=L_{t+1}^{\bar{K}_{t}=\bar{k}_{t}, \bar{A}_{t}=\bar{1}}.$ The multiple versions of achieving and maintaining the recommended MVPA through different MVPA minutes/week were addressed by using the stochastic exposure.^5,7^

Assuming the three assumptions hold, the protein level at Visit 3 under the intervention strategy (i.e., all participants achieved ≥150 minutes/week of MVPA at Visits 1 and 3) can be identified using the following g-formula. We used summation for easier presentation.

$$E\left[ Y_{3}^{\bar{K}_{3}=\bar{k}_{3}, \bar{A}_{3}=\bar{1},\bar{C}_{2}=\bar{0}} \right]$$

$$=\sum_{k_{3}} \sum_{l_{3}} \sum_{y_{2}} \sum_{l_{2}} \sum_{k_{1}} \sum_{l_{1}} E\left[ Y_{3} | K_{1},K_{3},L_{1},L_{2},L_{3},Y_{2},C_{1}=C_{2}=0, A_{1}=1,A_{3}=1 \right]$$

$$\times f\left( K_{3} | K_{1},L_{1},L_{2},L_{3},Y_{2},C_{1}=C_{2}=0,A_{1}=1,A_{3}=1 \right)$$

$$\times f\left( L_{3} \right|K_{1},L_{1},L_{2},Y_{2},C_{1}=C_{2}=0,A_{1}=1)$$

$$\times f\left( Y_{2} \right|K_{1},L_{1},L_{2},C_{1}=0,A_{1}=1)$$

$$\times f\left( L_{2} \right|K_{1},L_{1},C_{1}=0,A_{1}=1)$$

$$\times f\left( K_{1} \right|L_{1},A_{1}=1)$$

$$\times f(L_{1})$$

*IPW procedure and model specification*

We applied “artificial censoring” to the data, respectively for the intervention strategy and the control strategy. That is, observations when participants deviated from their assigned strategy and thereafter were deleted. To do so, we made two copies of the data (one for the intervention strategy and the other for the control strategy), applied the “artificial censoring” separately in each copy, and then combined them into one dataset in *long format* for analysis. We created an indicator of “being artificially censored”, *S_t_*, with 1 indicating the participant was censored (i.e., deviated from the assigned strategy) and 0 otherwise. Under the control strategy, everyone had a value of 0 for this indicator. Because this “artificial censoring” indicator is a one-to-one transformation of whether a participant achieved ≥150 minutes/week of MVPA conditioning on achieving ≥150 minutes/week in the previous visit, the inverse probability of “artificial censoring” weights are equivalent to the inverse probability of treatment weight (IPTW) typically used to control for confounding.^8^ These weights are distinct from the inverse probability of censoring weights (IPCW) that addresses the selection bias due to loss to follow-up and missing data. For simplicity, we use IPTW to denote the inverse probability of “artificial censoring” weights.

Because all participants in the control copy had 0 for the “artificial censoring indicator”, they did not contribute to the estimation of the IPTW. Moreover, as MVPA was not measured at Visit 2, observations at Visit 2 in the intervention copy did not contribute the IPTW estimation either. We excluded these observations by assigning weight of 0 when fitting the IPTW model. All other observations were assigned a weight of 1. The denominator of the IPTW was estimated using a weighted pooled logistic model with the “artificial censoring” indicator as the dependent variable using these weights. The IPTW denominator model included the main terms of the time-fixed confounders (age, sex, race, and education), the categorical time-varying confounders at the current visit (smoking status and number of chronic conditions), the continuous time-varying confounders (BMI, eGFR, and total cholesterol) at the current visit and lagged values from the previous two visits, and the lagged MVPA minutes/week and the lagged protein level from the last available visit. When lagged values were unavailable, e.g., no measurement prior to Visit 1 for all confounders, a value of 0 was used. Based on partial residual plots (not shown), we used restricted cubic spline terms for the current and lagged values of BMI (3 internal knots at 25^th^, 50^th^, and 75^th^ percentiles), eGFR (2 internal knots at 33^rd^ and 67^th^ percentiles) and protein level at Visit 2 (3 internal knots at 25^th^, 50^th^, and 75^th^ percentiles). We included a visit indicator with 1 indicating Visit 3 and 0 indicating Visit 1. We also included in the model three sets of interaction terms: (i) age interactions with the current values of all the time-varying confounders, other time-fixed confounders, the lagged MVPA minutes/week, and the lagged protein level, (ii) sex interactions with the current values of all the time-varying confounders, the time-fixed confounders, the lagged MVPA minutes/week, and the lagged protein level, and (iii) interactions between the visit indicator and the current values of the time-varying confounder and the time-fixed confounders except for age.

The IPCW was estimated in the combined dataset of the intervention copy and the control copy. Since censoring only occurred after Visit 1 and Visit 2 as protein at Visit 3 was the outcome of interest, observations at Visit 3 were excluded from model estimation by assigning a weight of 0 to those observations. The denominator of the IPCW was estimated using a weighted pooled logistic model with the indicator of not returning to the next study visit as the dependent variable. The IPCW denominator model included the main terms of the time-fixed confounders, the categorical time-varying confounders at the current visit, the continuous time-varying confounders at the current visit and the lagged values from the previous visit, and the current MVPA minutes/week and the protein level. The same restricted cubic splines were used for BMI, eGFR and protein levels as in the IPTW model. We included a visit indicator with 1 indicating Visit 2 and 0 indicating Visit 1 and a copy indicator with 1 indicating the intervention copy and 2 indicating the control copy. We also included in the model four sets of interaction terms: (i) age interactions with the current values of all the time-varying confounders, other time-fixed confounders, the current MVPA minutes/week, and the current protein level, (ii) sex interactions with the current values of all the time-varying confounders, the time-fixed confounders, the current MVPA minutes/week, and the current protein level, (iii) interactions between the visit indicator and the current values of the time-varying confounder and the time-fixed confounders except for age, and (iv) interactions between the visit indicator and the copy indicator.

The numerator of the IPTW was estimated using a pooled logistic regression model with only the visit indicator (Visit 1 or 3) as the independent variable. The numerator of the IPCW was estimated using a pooled logistic model that included the visit indicator (Visit 1 or 2), the copy indicator, and the interaction term between the two. The same weights were used to exclude observations not contributing the estimation of the weights.

The probability of having the observed “artificial censoring” status was predicted using the IPTW numerator model and denominator model. The ratio of the two predicted probabilities was the visit-specific IPTW. For example, for a participant who achieved ≥150 minutes/week of MVPA, the IPTW was the probability of “not being artificially censored” predicted from the numerator model divided by the probability of “not being artificially censored” predicted from the denominator model. Observations excluded from the IPTW models (i.e., the control copy and Visit 2 of the intervention copy) were given an IPTW of 1. The visit-specific IPCW was calculated the same way using the IPCW numerator and denominator models. Observations from Visit 3 were given an IPCW of 1.

The final weights were calculated by taking the product of the time-specific IPTW and IPCW across all visits up to each visit. The final weights at Visit 3 were used to fit the marginal structural model (MSM). The MSM included Visit 3 observations from participants who had achieved ≥150 minutes/week of MVPA at Visits 1 and Visit 3. The MSM used a weighted linear regression model with the observed protein level at Visit 3 as the dependent variable and the copy indicator (i.e., indicating the strategy) as the only independent variable. The 95% confidence interval was estimated using 1000 samples from the non-parametric cluster bootstrap^9^ where individuals were sampled with replacement and all of their follow-up observations were retained.

Since we examined 45 frailty-associated proteins as the outcome, the entire procedure was repeated 45 times, one for each protein. For each protein, only the level of the same protein at an earlier available visit, not other proteins, was used in the IPTW and IPCW models.

*ICE procedure and model specification*

Different from the IPW procedure, *wide format* data was used for the ICE procedure. We also created two copies of data for the intervention strategy and the control strategy but did not combine the copies. Instead, the following steps were performed in each copy.

Step 1a: we fitted a linear regression model of the observed protein level at Visit 3 among participants who were “not artificially censored” by Visit 3 and not censored (i.e., lost to follow-up or had missing data) before Visit 3. The model included the main terms of the time-fixed confounders, the categorical time-varying confounders at Visit 3, the continuous time-varying confounders at Visits 1-3, the MVPA minutes/week at Visits 1 and 3, and the protein level at Visit 2. The same number of internal knots used in the IPW procedure were used in the restricted cubic splines for BMI, eGFR, and protein level at each visit. The model also included two sets of interaction terms: (i) age interactions with other time-fixed confounders, the time-varying founders at Visit 3, the MVPA minutes/week at Visits 1 and 3, and the protein level at Visit 3, and (ii) sex interactions with the same variables.

Step 1b: using the model from Step1a, we predicted the outcome protein level among participants who were “not artificially censored” at Visit 3 and were not censored before Visit 3. This prediction was mainly for participants who had complete confounder measurements and MVPA measures at all visits but had missing protein measurements at Visit 3. If there had been no such participants, Steps 1a and 1b could have been omitted.

Step 2a: using the predicted values from Step 1b, we fitted a linear regression model among participants who were “not artificially censored” at Visit 3 and were not censored before Visit 3. The model included all the same terms as the Step 1a model except that all the terms involving the MVPA minutes/week at Visit 3 were removed from the model.

Step 2b: using the model from Step 2a, we predicted the outcome protein level among participants who were “not artificially censored” at Visit 2 and were not censored before Visit 3. For the intervention copy, this prediction included participants who achieved ≥150 minutes/week of MVPA at Visit 1 but did not achieve ≥150 minutes/week of MVPA at Visit 3. This step implicitly assigned MVPA minutes/week at Visit 3 for these participants based on their confounder histories.

Step 3a: using the predicted values from Step 2b, we fitted a linear regression model among participants who were “not artificially censored” at Visit 1 and were not censored before Visit 3. The model included all the same terms as the Step 2a model except that all the terms involving the time-varying confounders at Visit 3 were removed.

Step 3b: using the model from Step 3a, we predicted the outcome protein level among participants who were “not artificially censored” at Visit 1 and were not censored before Visit 2. This prediction included participants who were censored after Visit 2. This step averaged the outcome over the empirical distributions of the confounders at Visit 3.

Step 4a: using the predicted values from Step 3b, we fitted a linear regression model among participants who were “not artificially censored” at Visit 1 and were not censored before Visit 2. The model included all the same terms as the Step 3a model except that all the terms involving the time-varying confounders and the protein level at Visit 2 were removed.

Step 4b: using the model from Step 4b, we precited the outcome protein level among participants who were “not artificially censored” at Visit 1. This prediction included participants who were censored after Visit 1. This step averaged the outcome over the empirical distributions of the confounders and the protein level at Visit 2.

Step 5a: using the predicted values from Step 4b, we fitted a linear regression model among participants who were “not artificially censored” at Visit 1. The model included all the same terms as the Step 4a model except that all the terms involving the MVPA minutes/week at Visit 1 were removed. At this stage, only the confounders at Visit 1 remained in the model.

Step 5b: using the model from Step 5b, we predicted the outcome protein level among all participants in the sample. In the intervention copy, this step implicitly assigned MVPA minutes/week at Visit 1 for participants who did not achieve ≥150 minutes/week of MVPA based on their baseline confounders. This step also averaged the outcome over the empirical distributions of the confounders at Visit 1.

Step 6: we took the mean of predicted values from Step 5b.

The effect of adhering to the recommended ≥150 minutes/week of MVPA at Visit 1 and Visit 3 on the protein level at Visit 3 was estimated by taking the difference between the two means from Step 6 in each copy. The 95% confidence interval was estimated using 1000 samples from the non-parametric cluster bootstrap^9^ where individuals were sampled with replacement and all of their follow-up observations were retained. We repeated the entire procedure 45 times, one for each protein outcome of interest. For each protein, only the level of the same protein at an earlier available visit, not other proteins, was used in the relevant model fitting steps.

*Tipping analysis for unmeasured confounding at Visit 1*

We used the tipping analysis described in this review^10^ to assess the robustness to unmeasured confounding for the effects of MVPA at Visit 1 (i.e., exposure) on protein levels at Visit 3 (i.e., outcomes). The tipping analysis uses the known/assumed unmeasured confounder-outcome association and exposure-outcome effect to calculate how strong the unmeasured confounder-exposure association must be to bring the exposure-outcome effect to the null (i.e., the tipping point). This tipping analysis was not a direct assessment on the robustness to unmeasured confounding of the effects of interest in this paper (i.e., the effects of MVPA ≥150 minutes/week at both Visit 1 and Visit 3 on protein levels at Visit 3), which was a combination of (i) the direct effects of MVPA at Visit 1 on protein levels at Visit 3 (i.e., not through MVPA at Visit 3) and (ii) the effects of MVPA at Visit 3 on protein levels at Visit 3 conditional on Visit 1. In contrast, the tipping analysis was performed on the total effects of MVPA at Visit 1 on protein levels at Visit 3 (direct effects [i] plus the effects through MVPA Visit 3). The tipping analysis has not been extended to direct effect or the time-varying exposure setting (i.e., our effects of interest). However, the total effects robust to unmeasured confounding suggested by the tipping analysis may indicate the robustness of the direct effects (i) to unmeasured confounding. Moreover, we controlled for more covariates (e.g., protein levels at Visit 2) and the prior values of covariates (e.g., BMI) for effects (ii), compared to direct effects (i). Therefore, effects (ii) were less likely subject to unmeasured confounding. As a result, total effects robustness to unmeasured confounding may indicate the robustness of our effects of interest (the combination of [i] and [ii]) to unmeasured confounding. Other sensitivity analysis methods for unmeasured confounding for time-varying exposure exist,^11^ but do not easily allow for comparing the potential unmeasured confounders to measured confounders, which is helpful to provide a context of the plausible unmeasured confounders when little is known about them (e.g., what they could be and how strongly they are associated with the exposure and the outcome).

We first estimated the effects of MVPA at Visit 1 on protein levels at Visit 3 using linear regression models. The models included the time-fixed confounders, the categorical and continuous time-varying confounders, and the MVPA minutes/week at Visit 1. Restricted cubic splines with the same number of internal knots used in IPW and ICE methods were used for continuous confounders. Age and sex interactions with other confounders and MVPA minutes/week were also included. This model was the same as the model in Step 4a of the ICE method. Because MVPA minutes/week were included in the interaction terms with age and sex, we calculated the effects of MVPA at Visit 1 on protein levels at Visit 3 in 6 age-sex strata (at 45, 55, and 65 years for men and women, respectively). The effects were calculated using the linear combinations of the coefficients from the main term of MVPA minute/week and the interaction terms.

The tipping analysis was performed in each stratum. For benchmarking with observed covariates (described in the next paragraph), we standardized all continuous covariates (except for age) and MVPA minutes/week using their respective standard deviations (SD) at Visit 1. We assumed a range from -2 to +2 SD with 0.02 SD increments for the associations between an unmeasured confounder and protein levels at Visit 3 (i.e., outcomes). Then, for each value, we calculated the tipping points, i.e., the associations between the unmeasured confounder and MVPA minutes/week at Visit 1 (i.e., exposure) that would bring the effects of MVPA at Visit 1 on protein levels at Visit 3 to the null for each age-sex stratum and each protein.

To provide context on whether there exists a plausible unmeasured confounder that can exceed the tipping point, we fitted the following two models at Visit 3. The first one was a model of protein levels at Visit 3 as a function of the *Visit 3* MVPA*,* the covariates at Visit 3, and one of the three *Visit 2* covariates (i.e., BMI, eGFR, and protein level [same protein as the outcome]). The *Visit 3* covariates had the same non-linear and interaction terms as in the tipping analysis. The *Visit 2* covariate was modeled using a linear term. The second model was a model of *Visit 3* MVPA as a function of the covariates at Visit 3 and one of the three *Visit 2* covariates, with the same linear/non-linear/interaction terms as the first model. We estimated the observed associations of the Visit 2 covariates, one at a time, with protein levels at Visit 3 and MVPA at Visit 3, respectively, from the first and the second model. These two associations for the three *Visit 2* covariates were plotted in the same coordinates with the tipping points. If the point for the *Visit 2* covariate was not between the two curves formed by the tipping points, then it indicates that an unmeasured confounder with confounding strength as *Visit 2* covariate on *Visit 3* MVPA and Visit 3 protein level would bring the effect of *Visit 1* MVPA on these two proteins at Visit 3 to the null. We chose BMI, eGFR, and protein level as benchmarks because they were the strongest confounders in our analysis.

We performed the benchmarking with observed covariates this way because we could not use the associations of observed covariates at Visit 1 (e.g., BMI at Visit 1) with MVPA at Visit 1 and protein levels at Visit 3 for this comparison. This is because the associations of the unmeasured confounders with MVPA at Visit 1 and protein levels at Visit 3 should be conditional on all observed covariates (including BMI at Visit 1). The coefficients of an observed covariate at Visit 1 (e.g., BMI at Visit 1) were only conditional on the remaining covariates. Cinelli and Hazlett circumvented this issue by using the observed partial *R^2^* of an observed confounder to obtain a bound on the partial *R^2^* of the unmeasured confounder as strong as the observed confounder and then used the bound to compare to the tipping points.^12,13^ We could not use the partial *R^2^* due to the presence of interaction terms with MVPA minutes/week.

*Other methods for left truncation by death not used in this study*

Besides the exclusion of participants who died before the outcome was measured (our sensitivity analysis), a few other advanced methods exist but were not used in this study. These methods have different limitations. The principal stratification method deals with left truncation by death by estimating the effect in a subpopulation of participants who would have survived with or without intervention (“principal stratum”).^14^ It ignores other participants in the population, e.g., those who did not survive without the intervention but would have survived with the intervention. Also, we do not know who is in the principal stratum as we do not observe survival under intervention versus no intervention for the same person simultaneously (i.e., one of the scenarios is counterfactual).^15^ The separable effects method handles truncation by death by separating an intervention strategy into two components: the first component exerts an effect on the outcome of interest not through death, and the second component exerts an effect on the outcome of interest through death. The separable effects estimate a direct effect of the first component of the intervention on the outcome of interest while setting the second component of the intervention to a fixed value for everyone.^16^ The hypothetical decomposition of the intervention strategy may not be plausible in real life. Therefore, the approach may also have challenges for real-life interpretation.

*Covariate measurements*

Time-fixed covariates include age (years), sex (male or female), race (black or white), and education (less than high school, high school/GED/vocational school, and any college) were self-reported at Visit 1. Smoking status (current, former, and never smoker) was also self-reported. Current and former smoking were combined into one category for positivity. Body mass index (BMI, kg/m^2^) was calculated using weight and height measured at each visit. Estimated glomerular filtration rate (eGFR, mL/min/1.73 m^2^) at Visits and 3 was calculated using serum creatinine, serum cystatin C, age, and sex using the CKD-EPI 2021 Equation.^17^ At Visit 1, eGFR estimation excluded serum cystatin C because it was not measured at the time.^17^ Total cholesterol was measured using the enzymatic method.^18^ The number of chronic conditions (0, 1 or ≥2) included the following diseases: hypertension, diabetes, coronary heart disease (CHD), heart failure, stroke, cancer, and chronic lung disease. Hypertension was defined as a systolic blood pressure >140 mmHg, a diastolic blood pressure >90 mmHg, or use of hypertensive medication. Diabetes was defined as a fasting glucose ≥126 mg/dL, non-fasting glucose ≥200 mg/dL, current use of diabetes medication, or self-reported physician diagnosis. CHD was defined as a self-reported history at Visit 1 and adjudicated events at subsequent visits based on medical record evidence of myocardial infarction, coronary artery bypass graft or angioplasty and myocardial infarction determined by ECG. Heart failure was identified by self-reported medication use for heart failure or meeting the Gothenburg Criteria^19^ at Visit 1. At Visits 2 and 3, heart failure was identified as having a heart failure at Visit 1 or having a hospitalization with an ICD discharge diagnosis code indicating heart failure in any position before the date of the visit. Stroke was defined as self-report physician diagnosis at Visit 1. At Visits 2 and 3, stroke was defined by having stroke at Visit 1 or having adjudicated event after Visit 1 but before Visit 2 or 3. Cancer was ascertained by linkage with cancer registry, medical records and hospital discharge summaries, and death certificates.^20^ Chronic lung disease was defined as self-reported physician diagnoses.

*References*

*Appendix Figure 1. Positivity checks across levels of adherence to recommended MVPA at Visit 1*

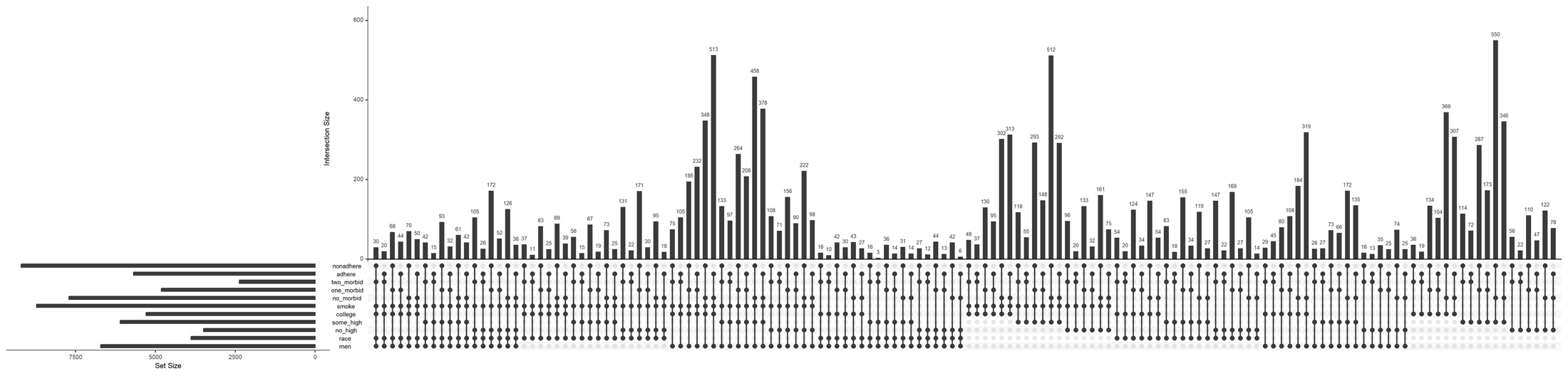

*Appendix Figure 2. Positivity checks across levels of adherence to recommended MVPA at Visit 3 conditional on adherence at Visit 1 and no prior censoring*

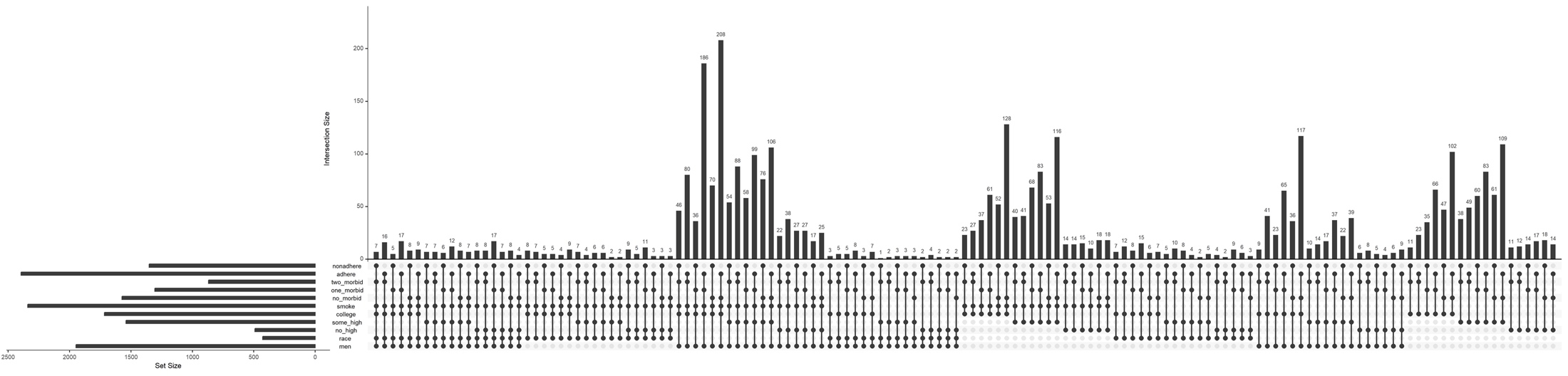

*Appendix Figure 3. Positivity checks across levels of censoring at Visit 1*

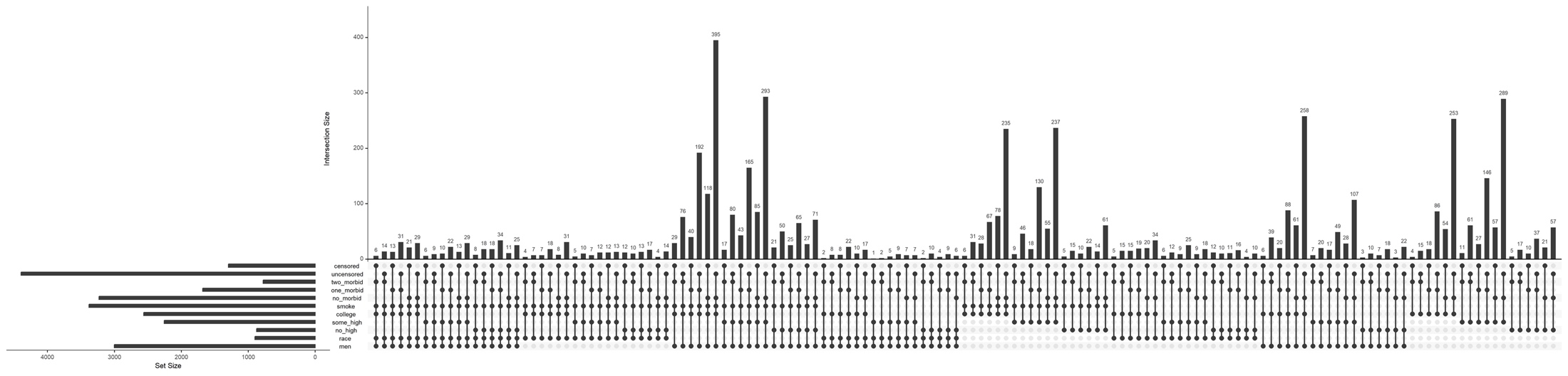

*Appendix Figure 4. Positivity checks across levels of censoring at Visit 2 conditional on adherence at Visit 1 and no prior censoring*

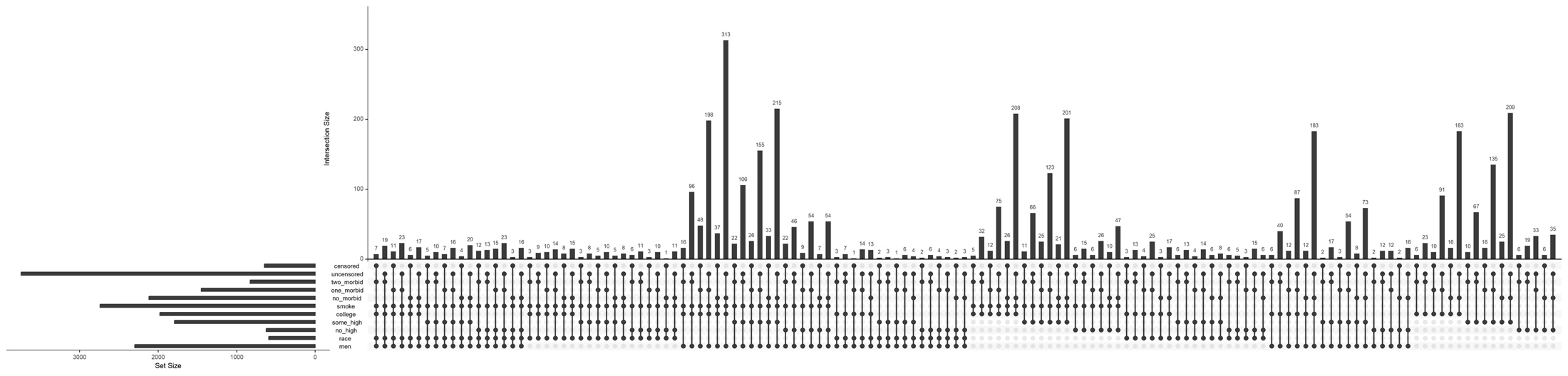
