## Supplemental Tables and Figures for "The effect of long-term adherence to physical activity recommendations in midlife on plasma proteins associated with frailty in the Atherosclerosis Risk in Communities (ARIC) study"

**Figure S1.** Sample selection excluding participants with major chronic conditions at baseline and transitions between achieving and not achieving ≥150 minutes/week of moderate-to-vigorous physical activity (MVPA) during the follow-up. Major chronic conditions included coronary heart disease, heart failure, stroke, cancer, and chronic lung disease.

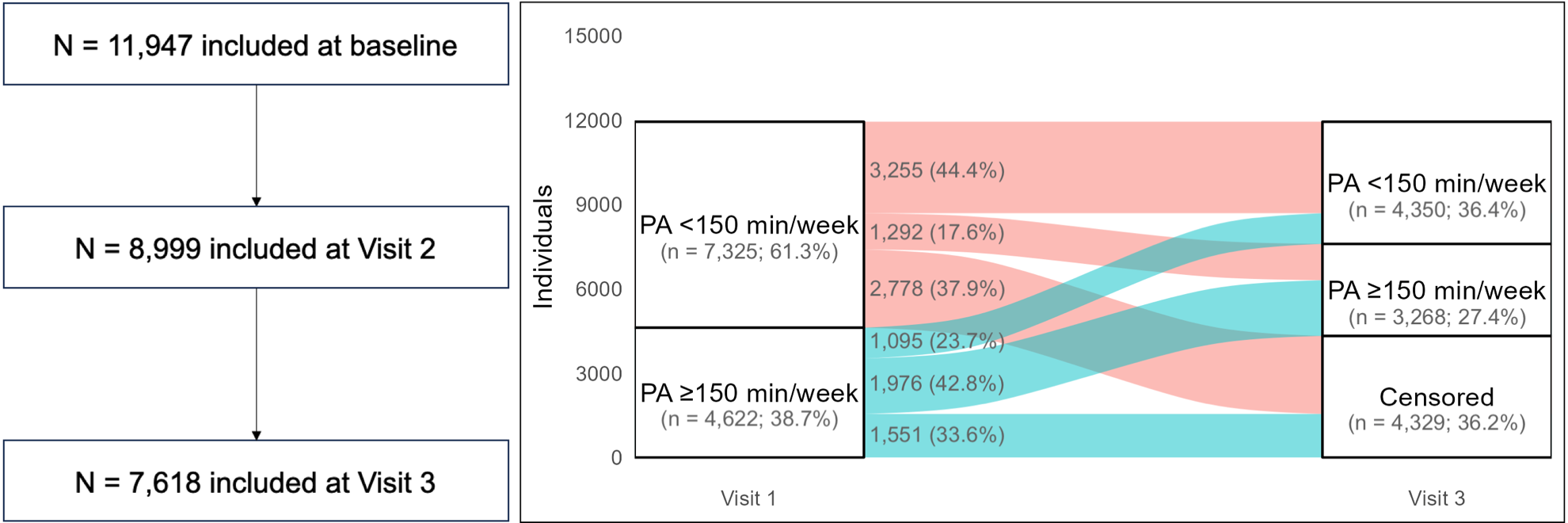

**Figure S2.** Sample selection excluding participants who died before Visit 3 and transitions between achieving and not achieving ≥150 minutes/week of moderate-to-vigorous physical activity (MVPA) during the follow-up. Death before Visit 3 was defined as date of death on or prior to February 5^th^, 1996.

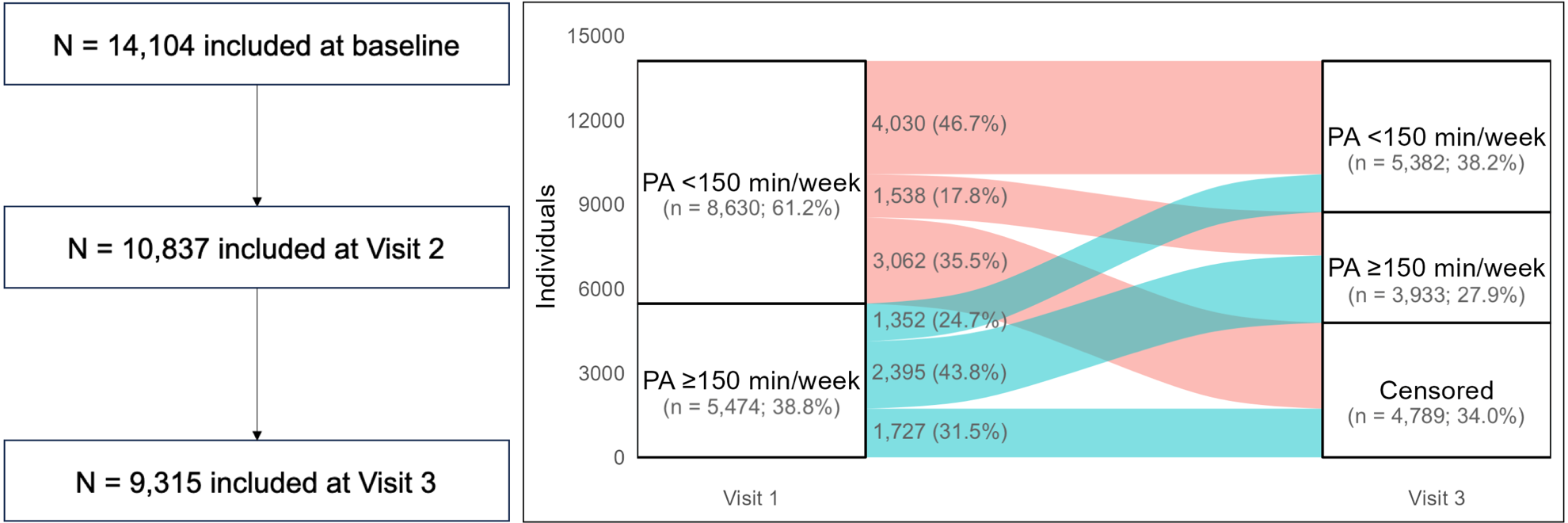

**Table S1.** Final weight distribution at Visit 3 for IPW method

| Protein Gene Symbol | Intervention strategy | Control strategy | Protein Gene Symbol | Intervention strategy | Control strategy |
| --- | --- | --- | --- | --- | --- |
|  | Mean (min, max) | Mean (min, max) |  | Mean (min, max) | Mean (min, max) |
| NCAN | 1.00 (0.30, 19.99) | 1.00 (0.74, 6.68) | SCG3 | 1.01 (0.30, 18.97) | 1.00 (0.74, 9.78) |
| SEZ6L | 1.01 (0.30, 24.03) | 1.00 (0.73, 8.79) | TMEM132D | 1.01 (0.30, 23.83) | 1.00 (0.73, 7.30) |
| IGFBP1 | 1.00 (0.31, 23.05) | 1.00 (0.73, 9.30) | UST | 1.01 (0.30, 21.99) | 1.00 (0.73, 6.99) |
| CNTN1 | 1.00 (0.30, 21.50) | 1.00 (0.73, 14.06) | FABP3 | 1.01 (0.30, 22.16) | 1.00 (0.72, 8.01) |
| MSMP | 1.00 (0.30, 25.76) | 1.00 (0.73, 8.17) | LEP | 1.00 (0.30, 24.61) | 1.00 (0.73, 7.29) |
| ATP1B2 | 1.00 (0.30, 22.21) | 1.00 (0.73, 7.79) | FABP4 | 1.01 (0.30, 22.99) | 1.00 (0.73, 7.79) |
| ART3 | 1.00 (0.31, 22.37) | 1.00 (0.73, 17.47) | FSTL3 | 1.00 (0.30, 19.56) | 1.00 (0.73, 10.36) |
| APLP1 | 1.01 (0.30, 27.07) | 1.00 (0.72, 7.12) | HTRA1 | 1.01 (0.30, 20.34) | 1.00 (0.72, 9.55) |
| CACNA2D3 | 1.00 (0.30, 27.46) | 1.00 (0.72, 5.63) | DCUN1D1 | 1.01 (0.30, 26.42) | 1.00 (0.73, 7.71) |
| IGLON5 | 1.01 (0.30, 25.75) | 1.00 (0.73, 7.31) | IL1RN | 1.01 (0.30, 23.22) | 1.00 (0.73, 8.61) |
| NOTCH1 | 1.00 (0.30, 23.67) | 1.00 (0.73, 5.92) | SERPINE1 | 1.01 (0.30, 23.49) | 1.00 (0.72, 8.01) |
| NCAM1 | 1.00 (0.30, 25.07) | 1.00 (0.72, 6.27) | CRP | 1.00 (0.30, 21.18) | 1.00 (0.74, 6.03) |
| UNC5D | 1.00 (0.30, 22.39) | 1.00 (0.73, 8.46) | INHBC | 1.01 (0.30, 22.73) | 1.00 (0.73, 7.44) |
| SLITRK1 | 1.00 (0.30, 16.84) | 1.00 (0.73, 8.05) | SAA1 | 1.01 (0.30, 26.36) | 1.00 (0.73, 7.69) |
| SLITRK3 | 1.00 (0.31, 22.81) | 1.00 (0.73, 7.99) | INS | 1.01 (0.30, 22.96) | 1.00 (0.72, 8.55) |
| PTPRD | 1.01 (0.30, 26.52) | 1.00 (0.74, 6.41) | TLR5 | 1.01 (0.30, 19.37) | 1.00 (0.73, 8.83) |
| TMEM132B | 1.01 (0.30, 20.87) | 1.00 (0.73, 8.12) | SULF2 | 1.01 (0.30, 21.76) | 1.00 (0.73, 7.58) |
| WFIKKN2 | 1.01 (0.30, 24.36) | 1.00 (0.73, 8.34) | CFB | 1.00 (0.30, 22.29) | 1.00 (0.73, 6.73) |
| HS6ST3 | 1.00 (0.30, 23.62) | 1.00 (0.73, 6.60) | GOLM1 | 1.00 (0.30, 23.42) | 1.00 (0.73, 7.47) |
| LSAMP | 1.01 (0.30, 23.22) | 1.00 (0.73, 8.31) | NSF | 1.01 (0.30, 23.72) | 1.00 (0.73, 8.40) |
| CST2 | 1.01 (0.30, 22.85) | 1.00 (0.73, 8.50) | PTGR1 | 1.00 (0.30, 21.69) | 1.00 (0.72, 8.53) |
| DKK3 | 1.01 (0.30, 23.42) | 1.00 (0.73, 8.08) | HSPA1B | 1.01 (0.30, 23.69) | 1.00 (0.72, 7.84) |
| DNER | 1.00 (0.30, 21.33) | 1.00 (0.72, 5.94) |  |  |  |

**Figure S3.** Balance of **included covariates** between participants who achieved ≥150 minutes/week of MVPA and those who did not at Visits 1 and 3 after weighting with each of the proteins (except for CACNA2D3 and HITRA1) as the outcome. The circle represents the intervention strategy. The dashed lines denote ±0.25 SD

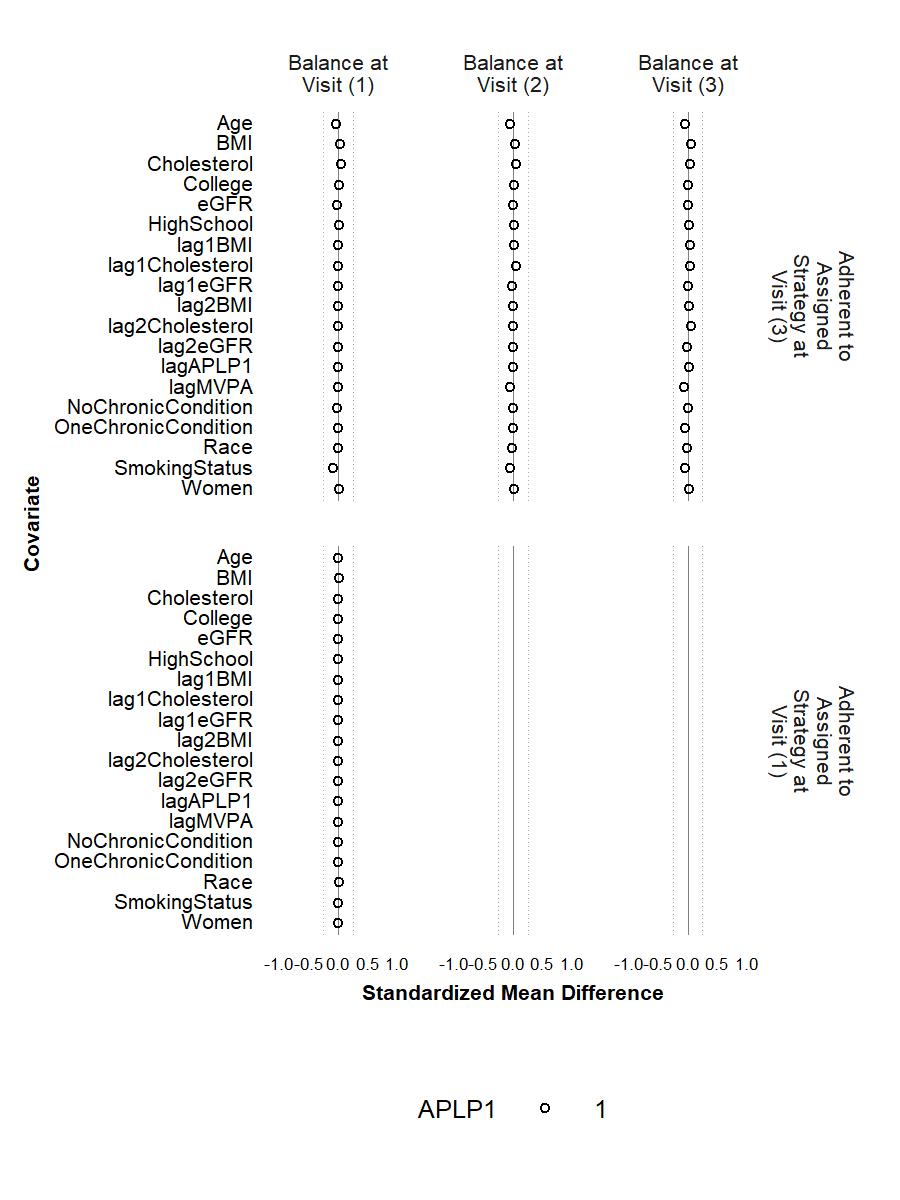

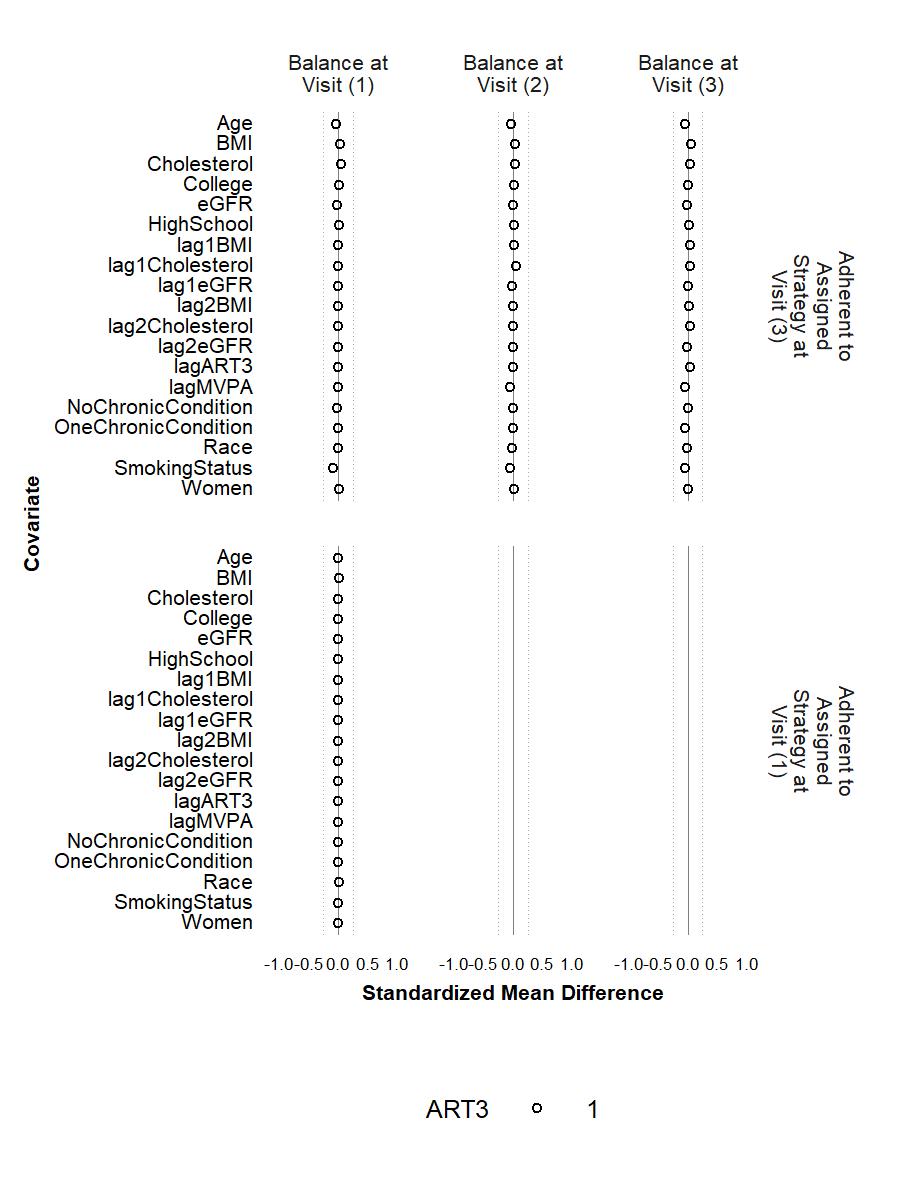

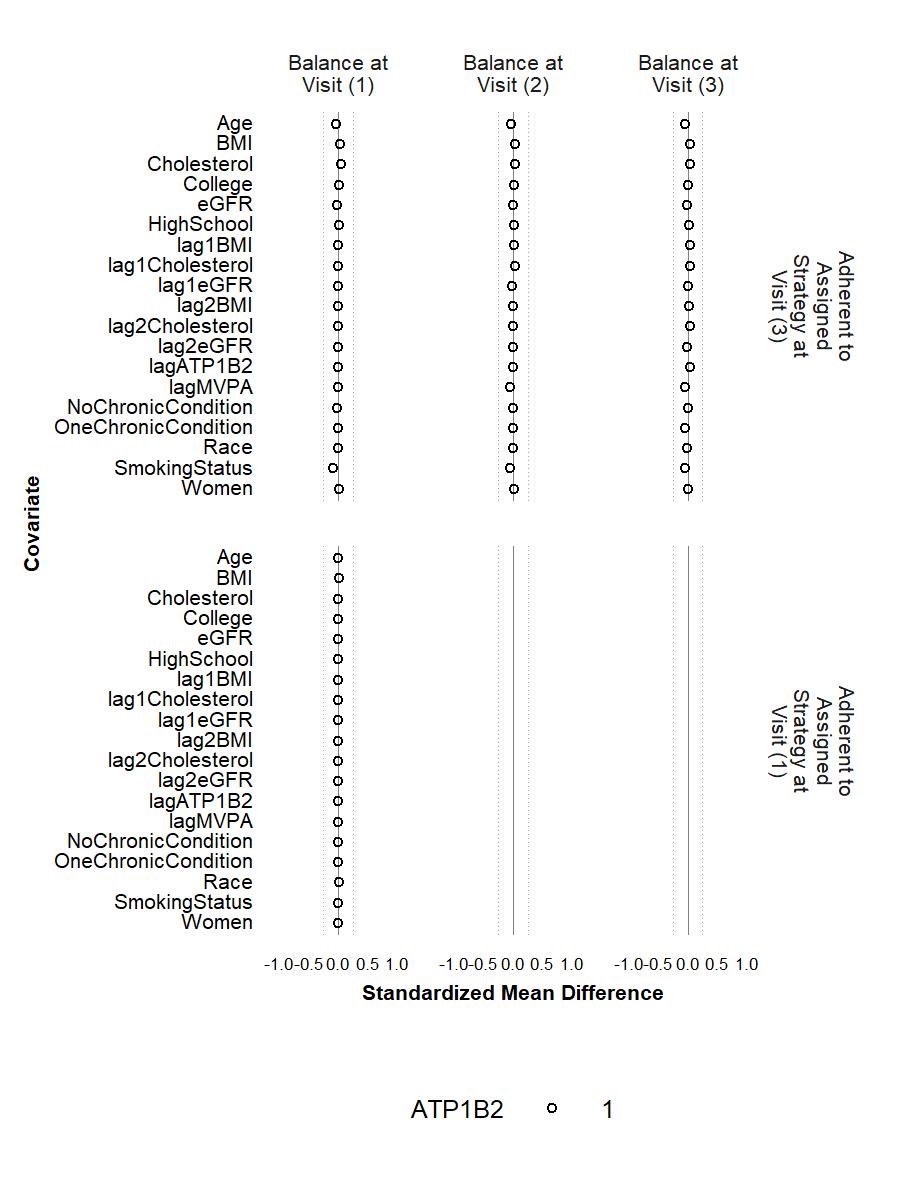

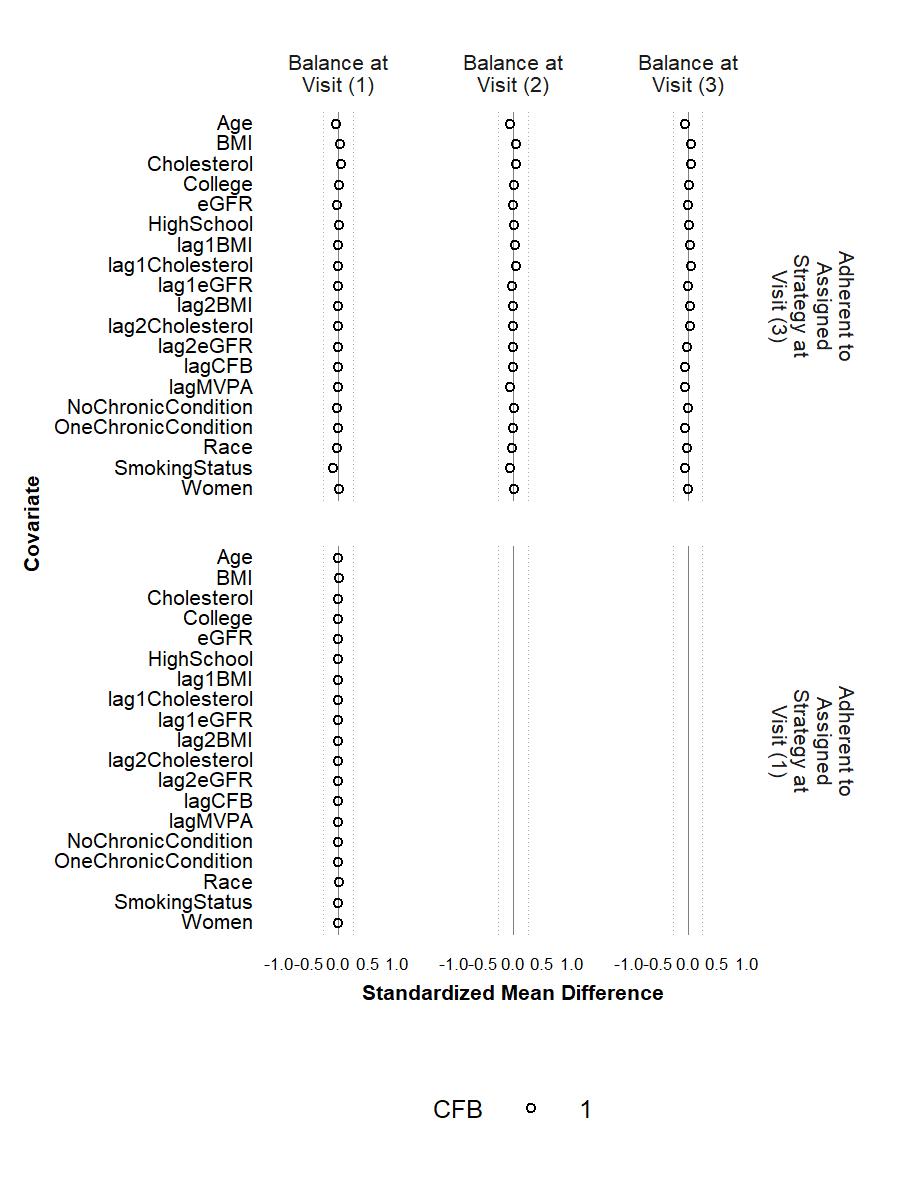

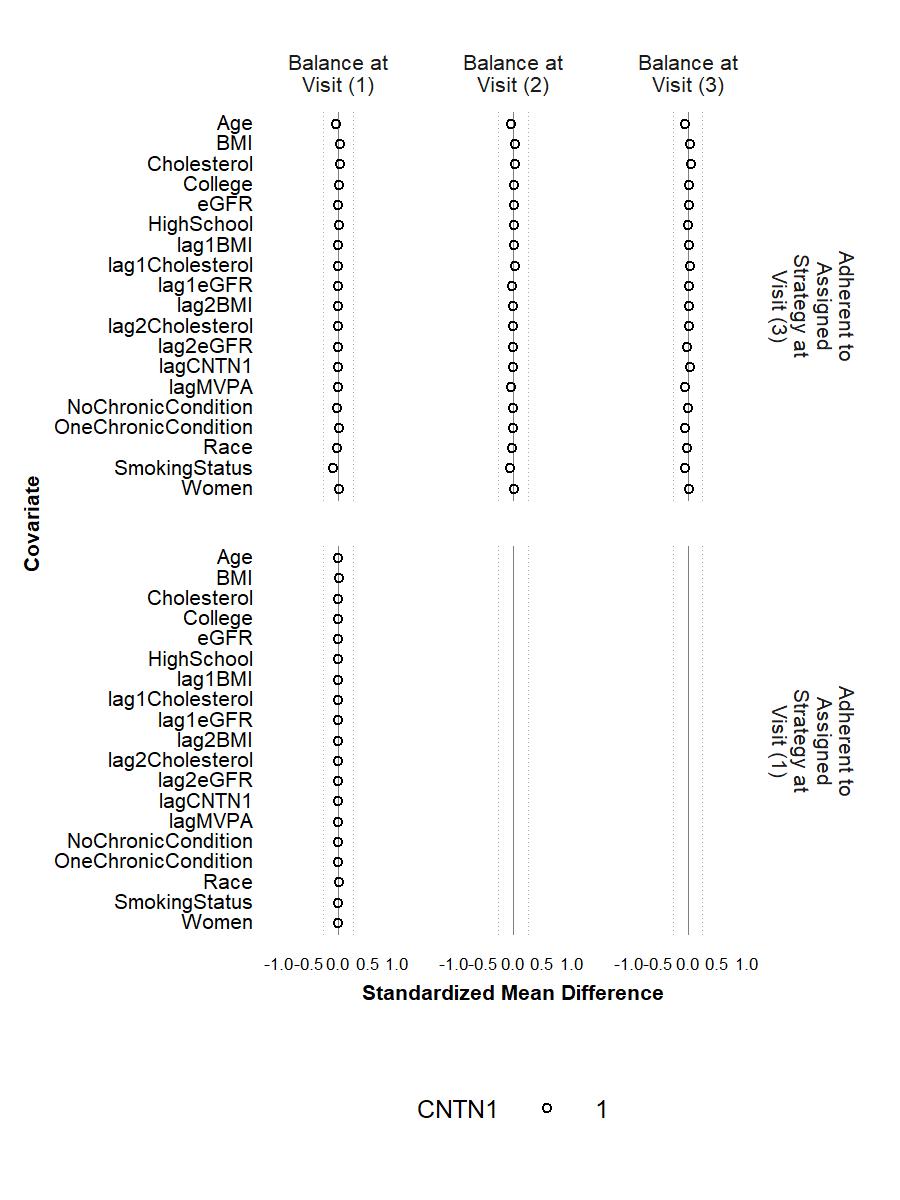

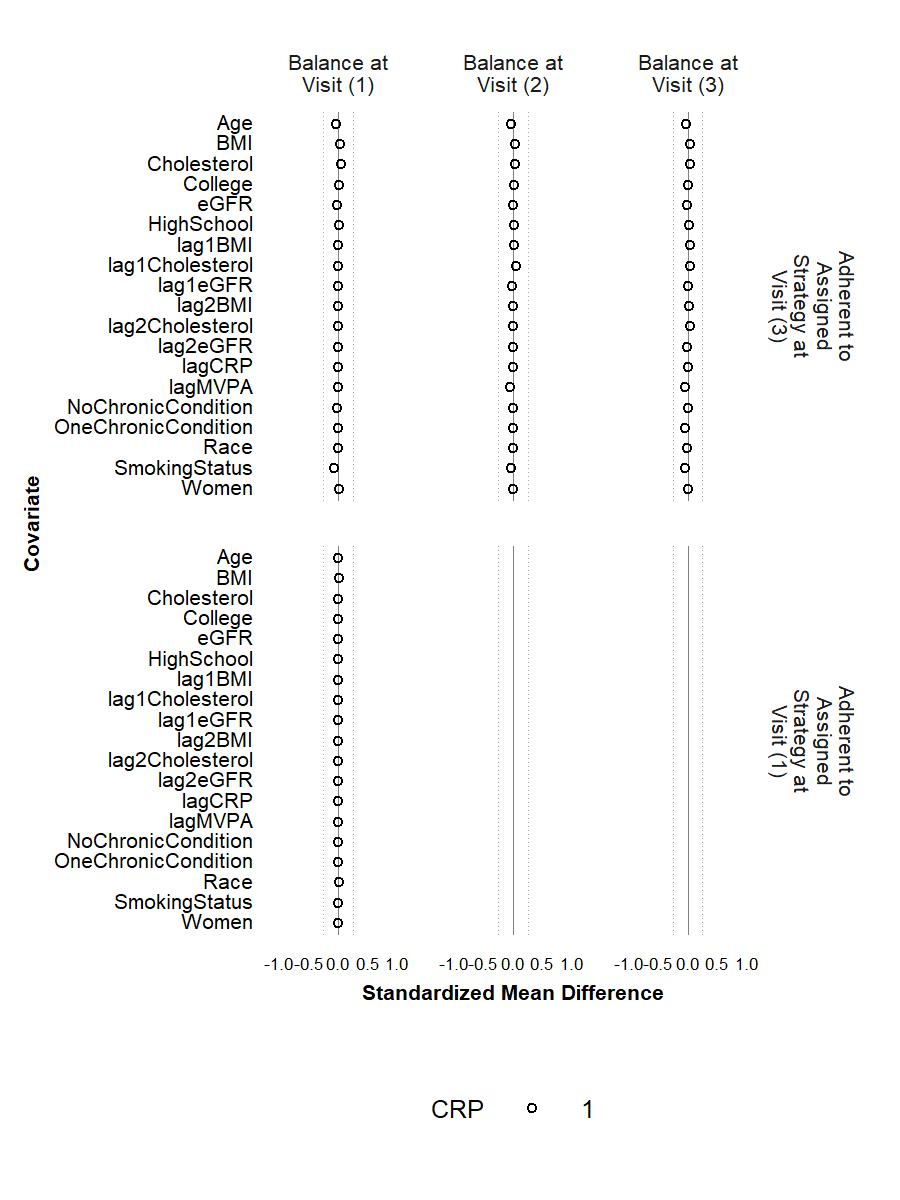

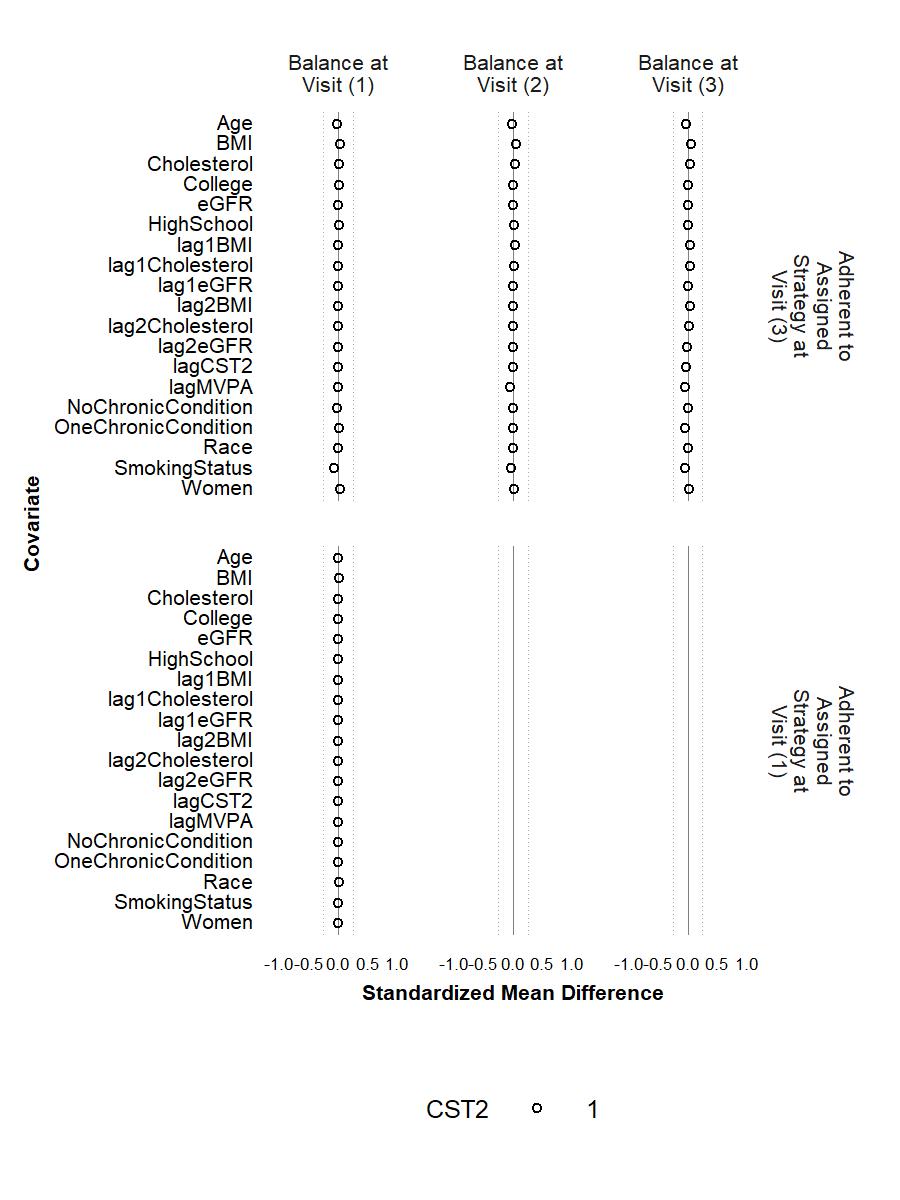

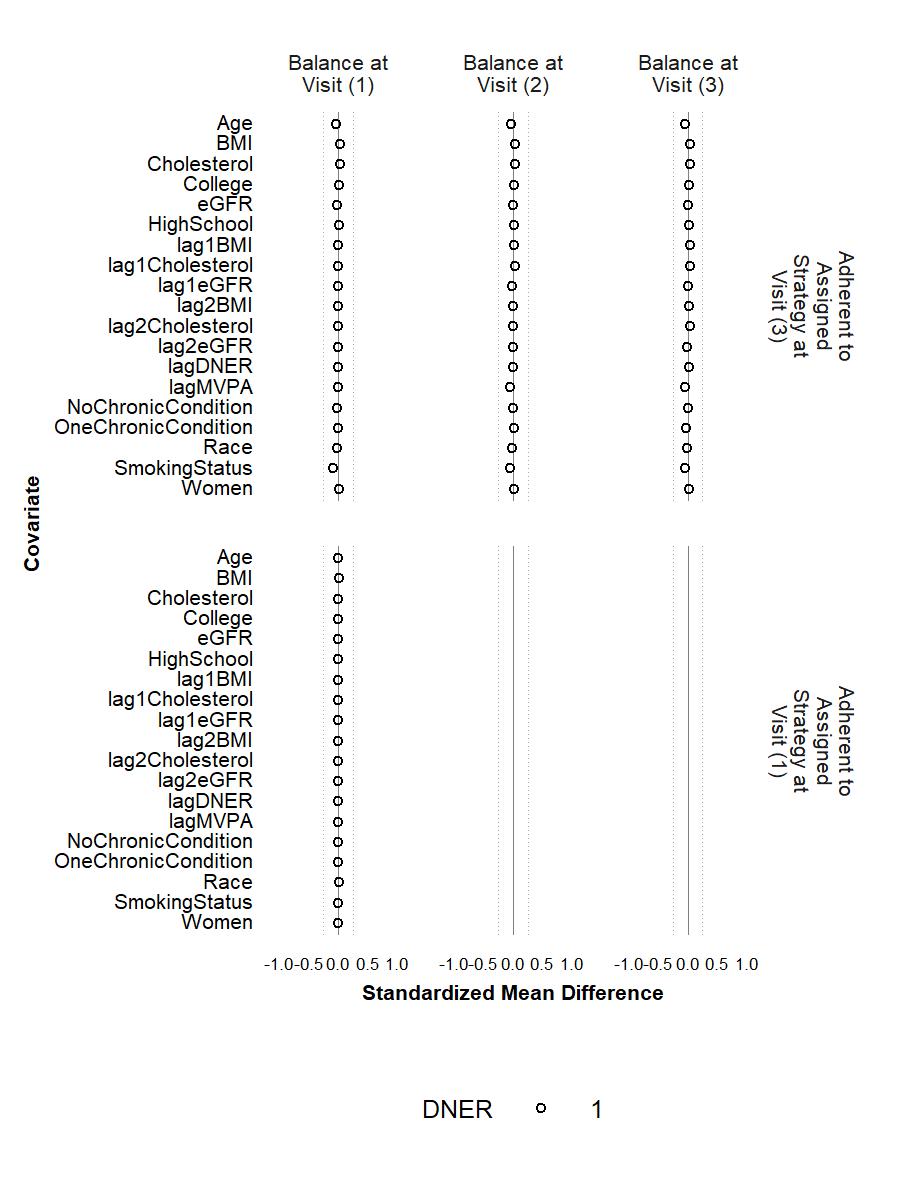

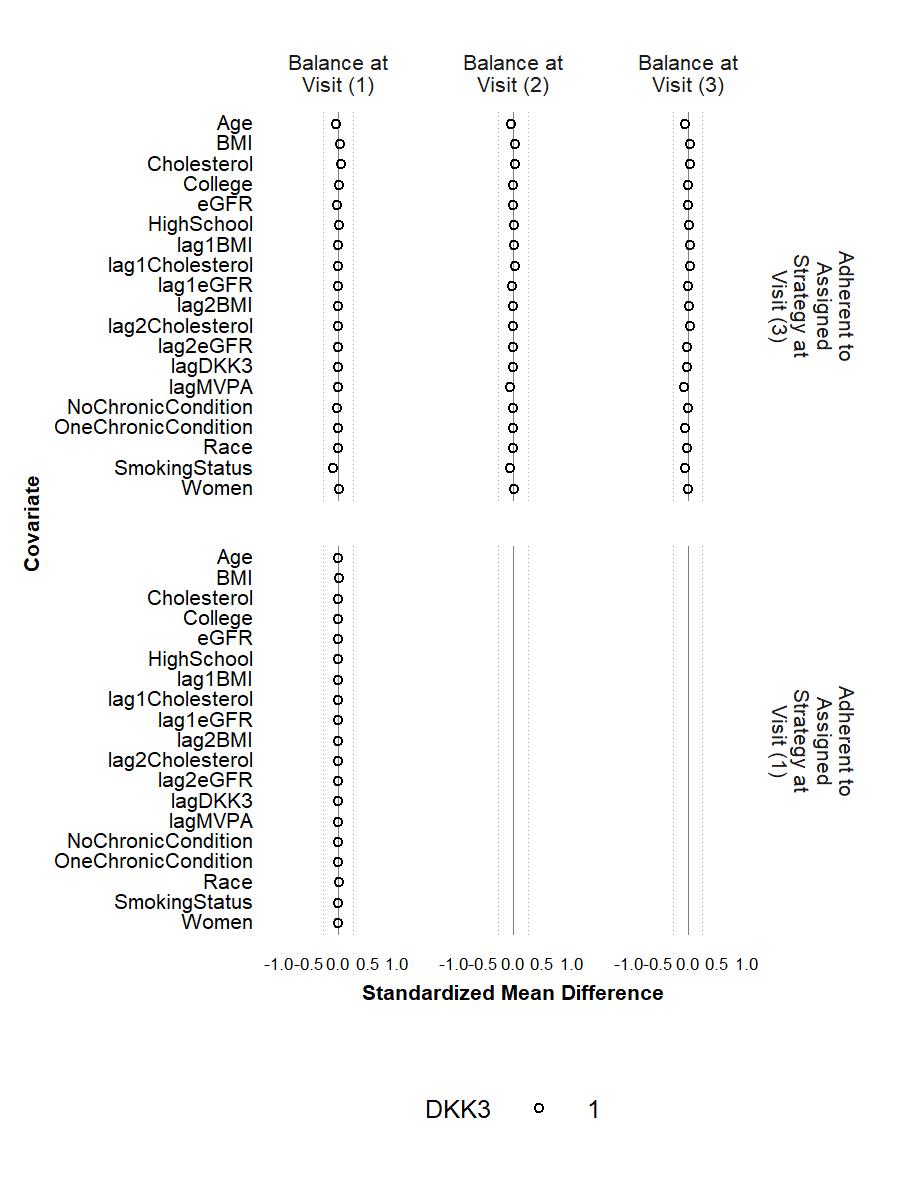

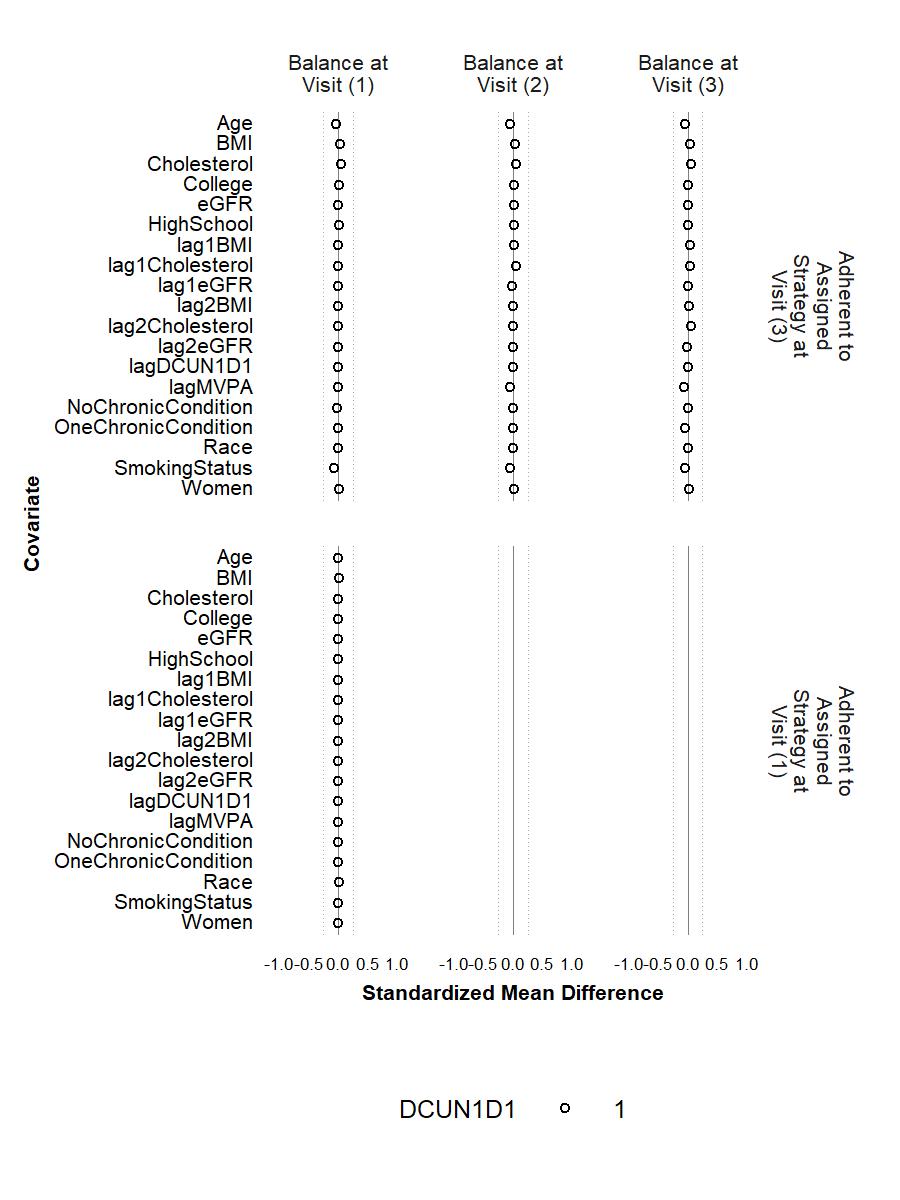

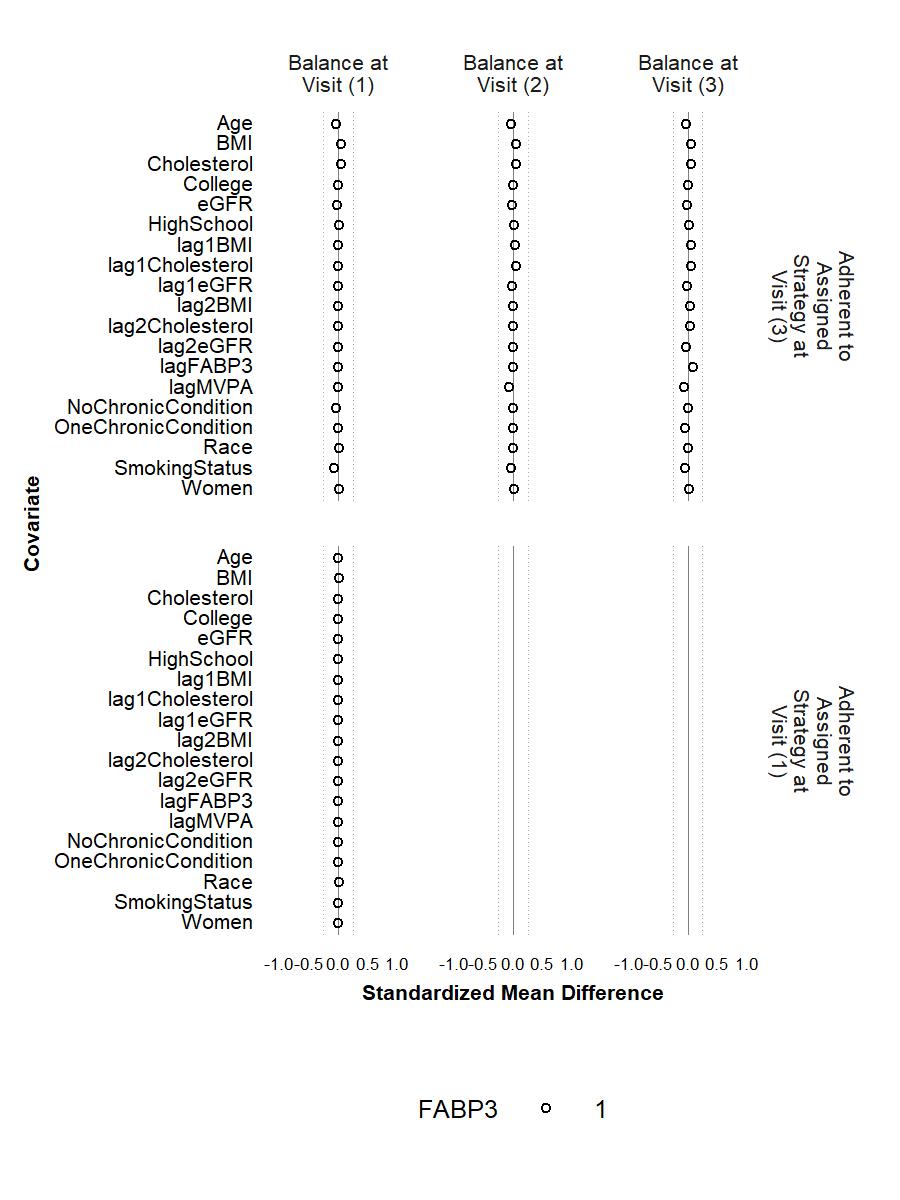

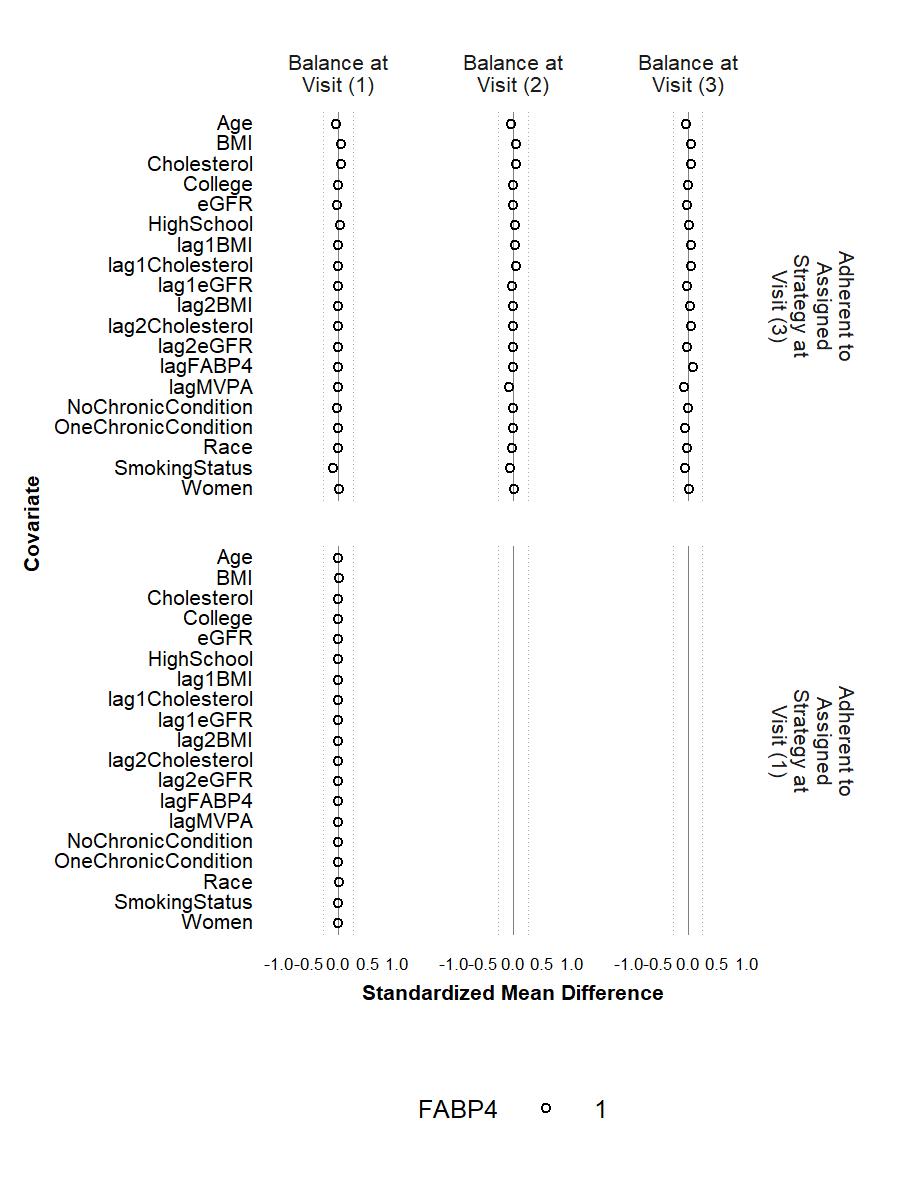

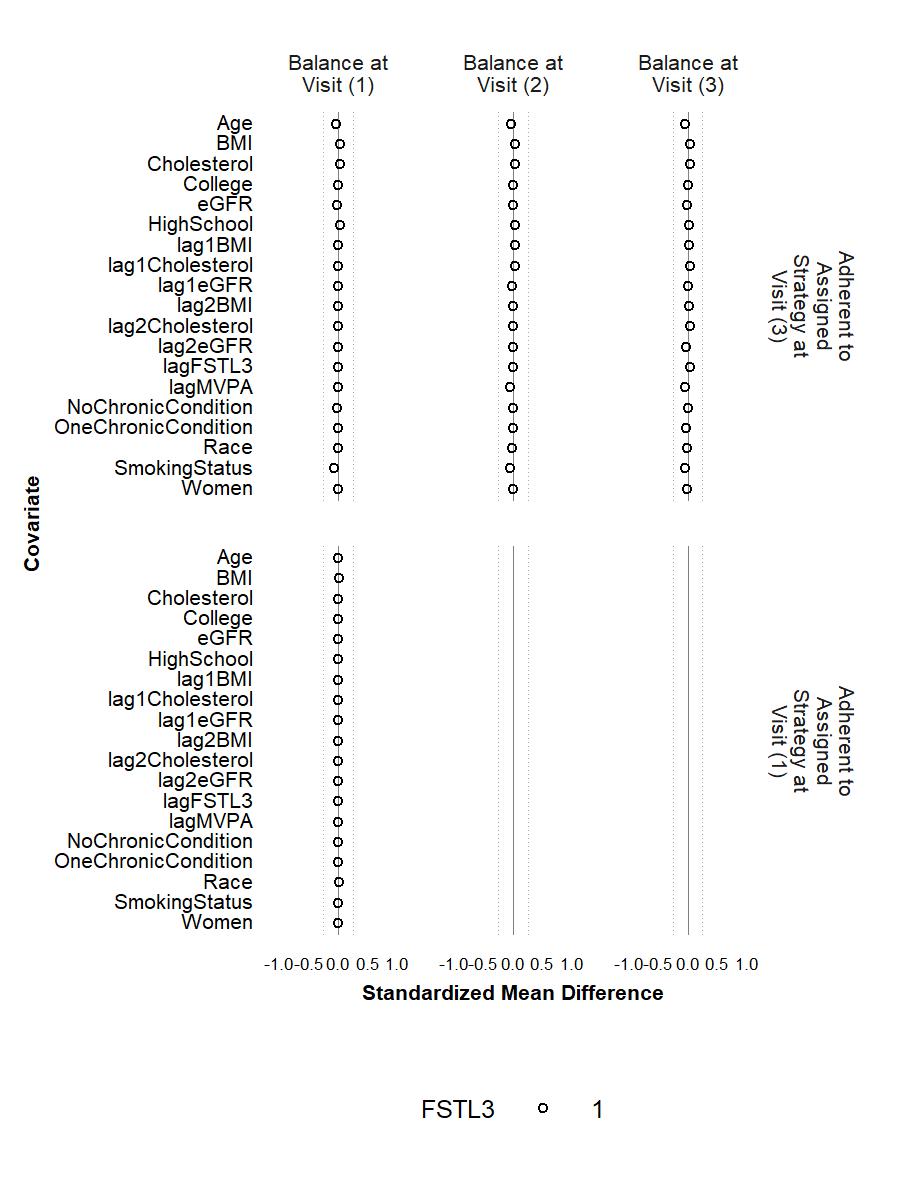

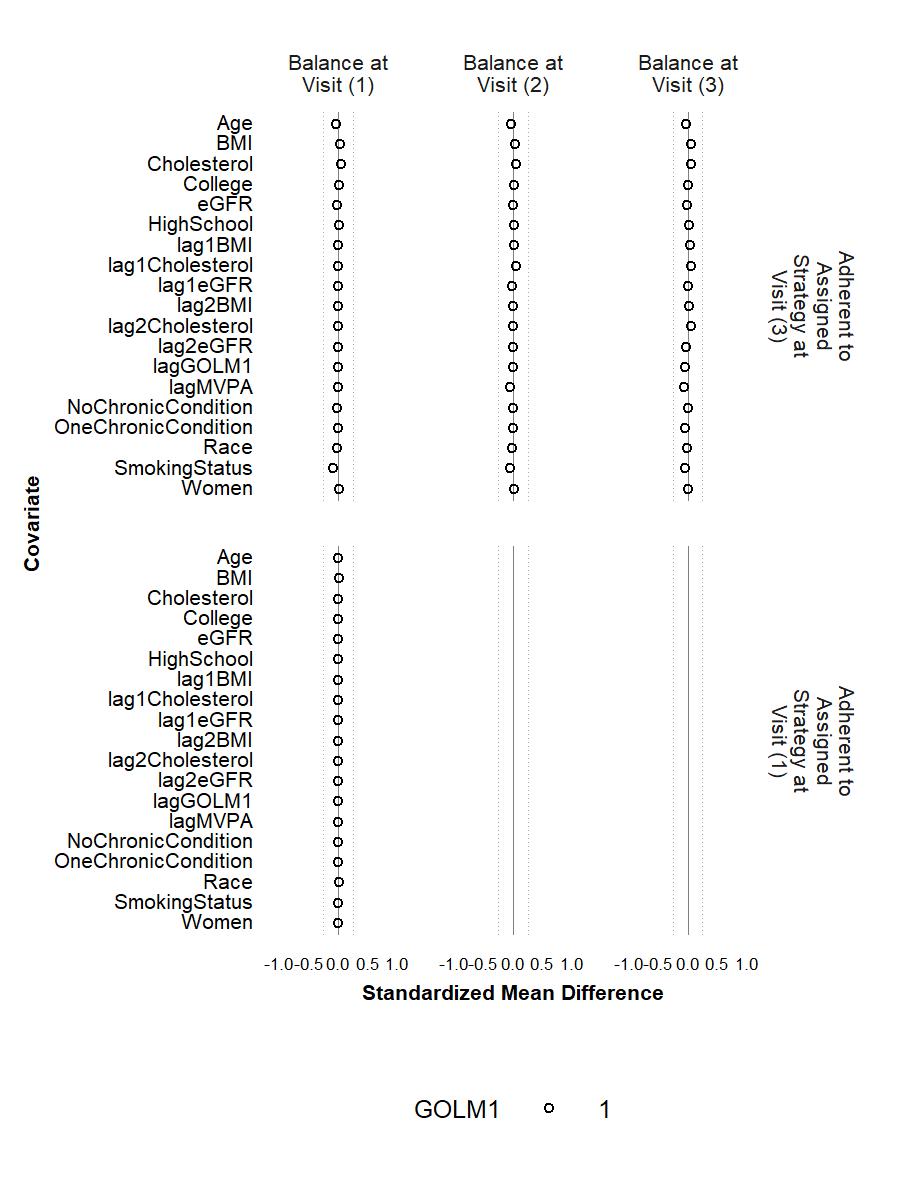

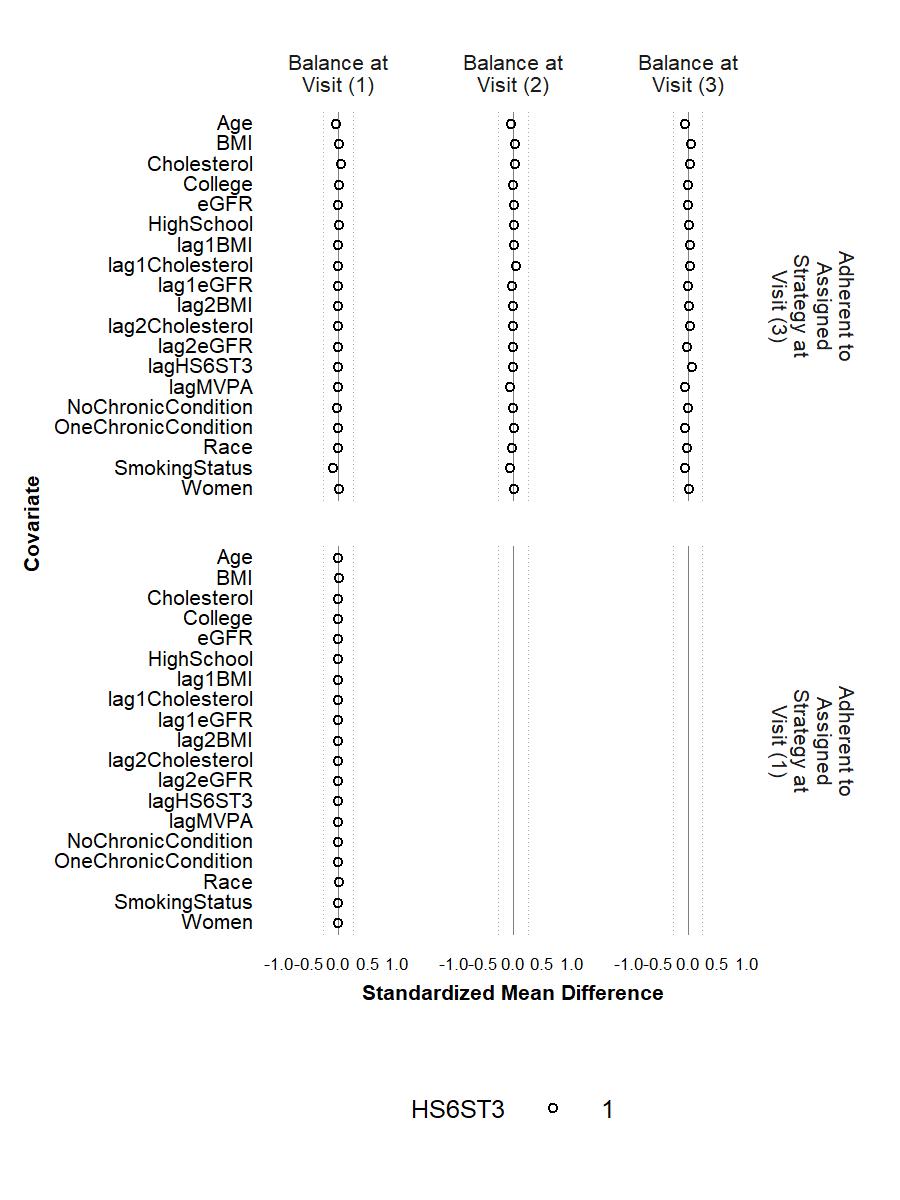

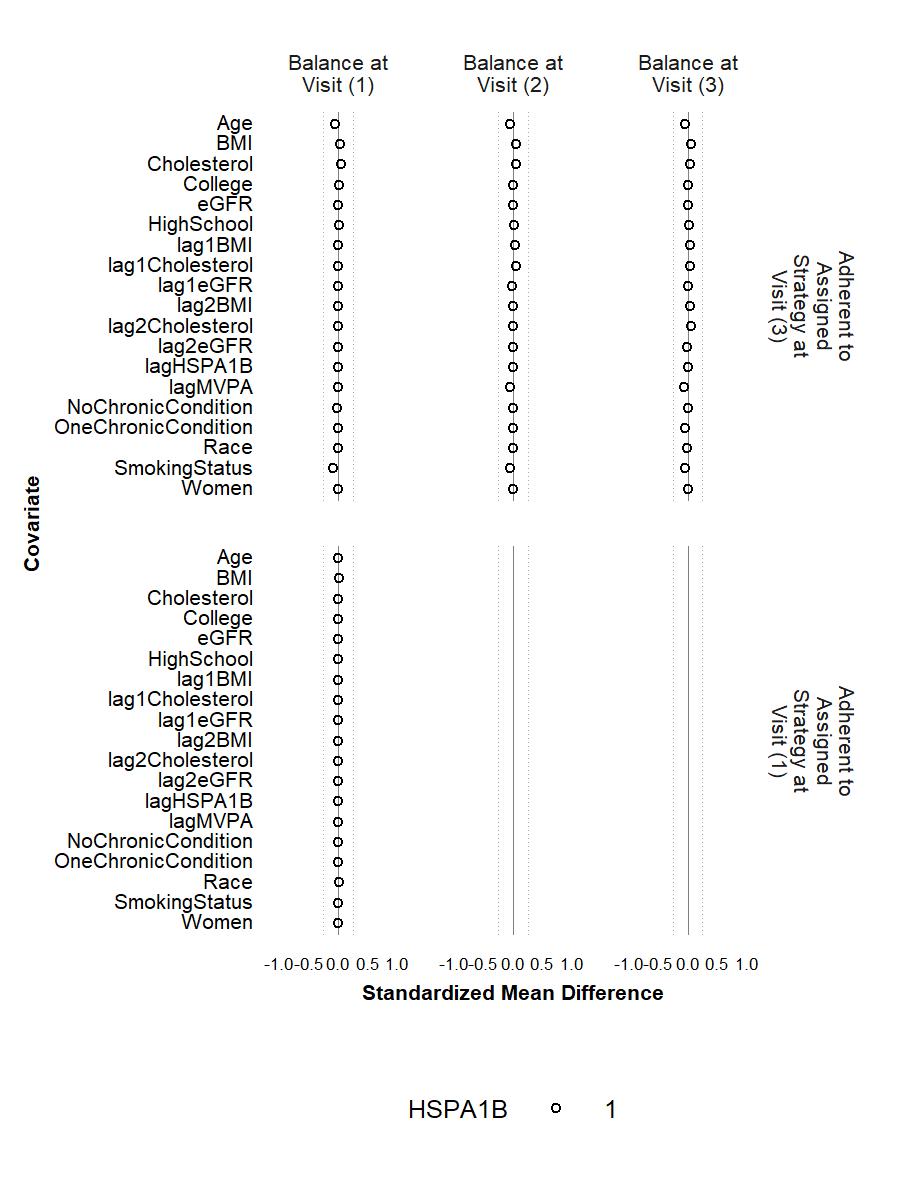

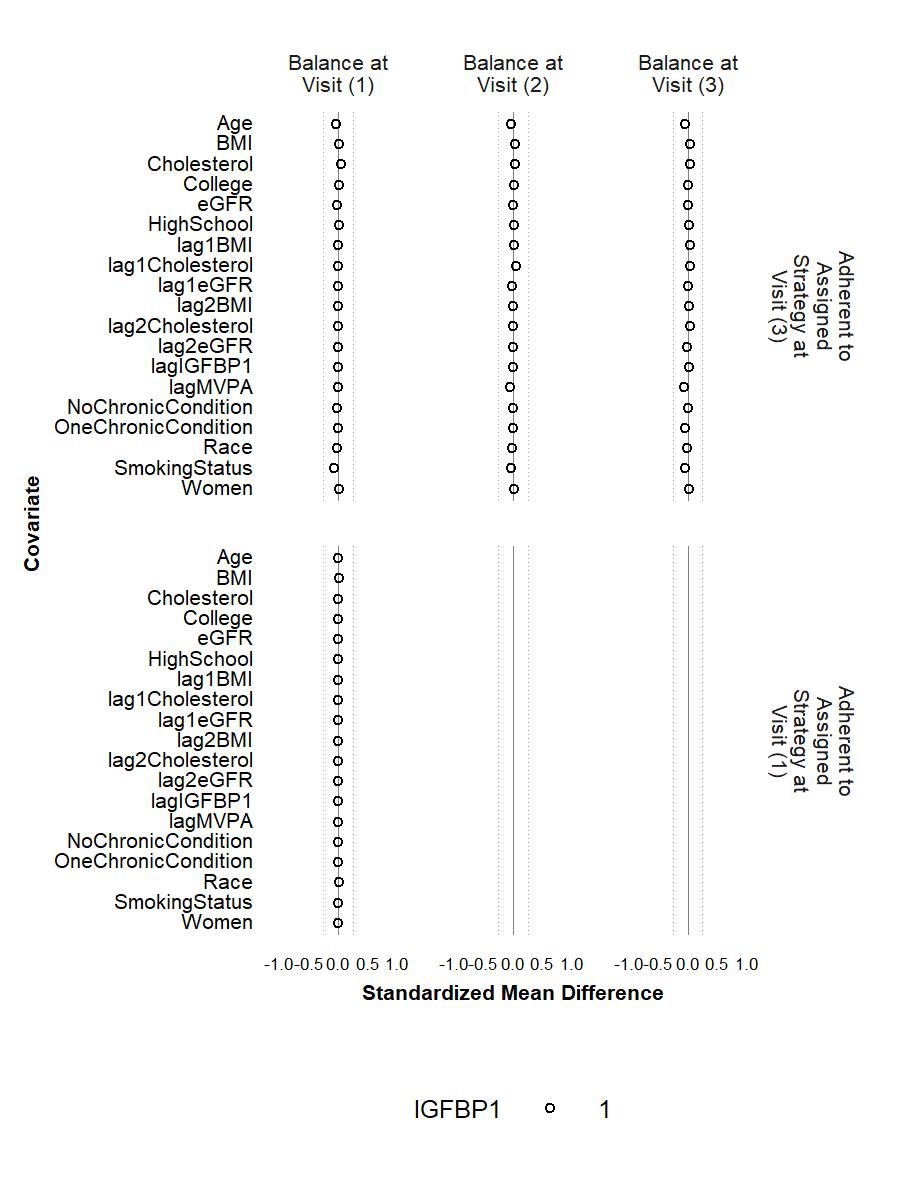

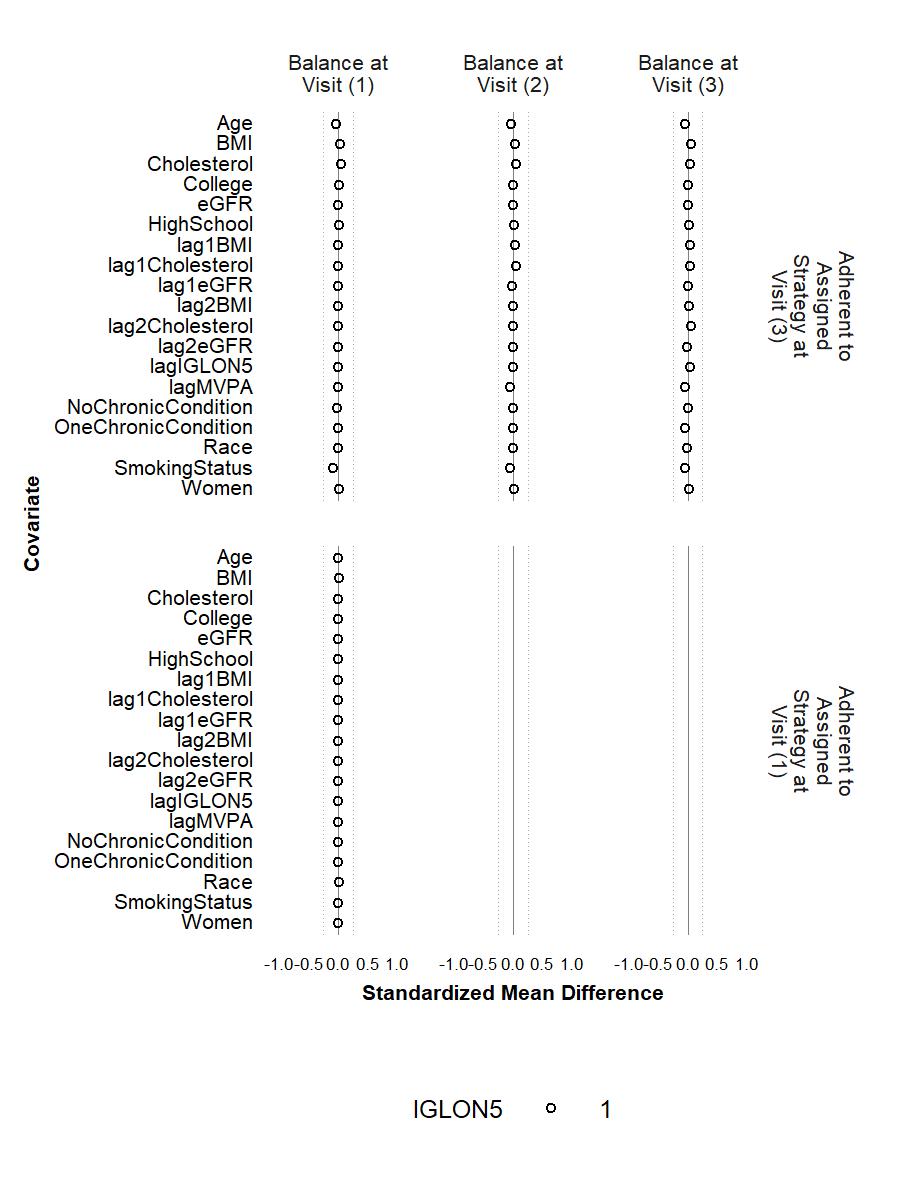

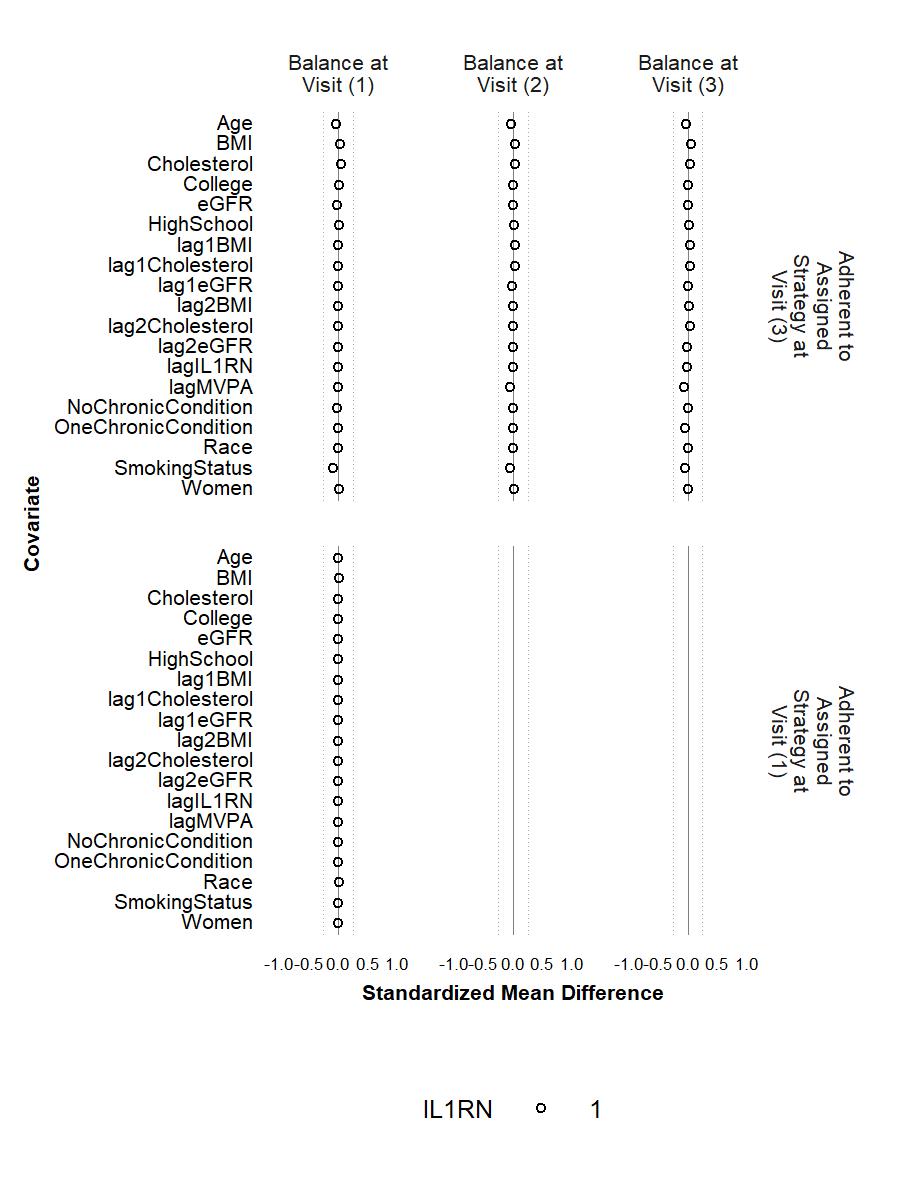

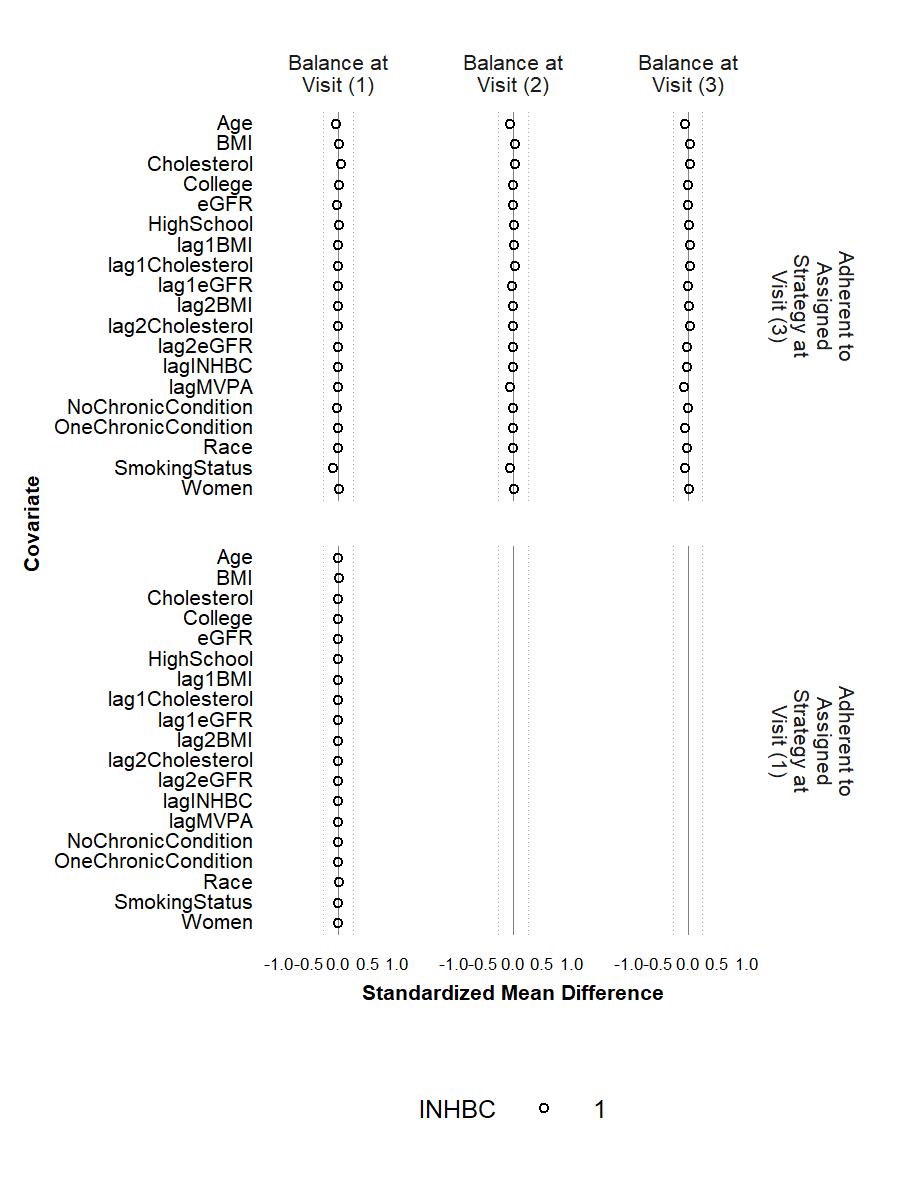

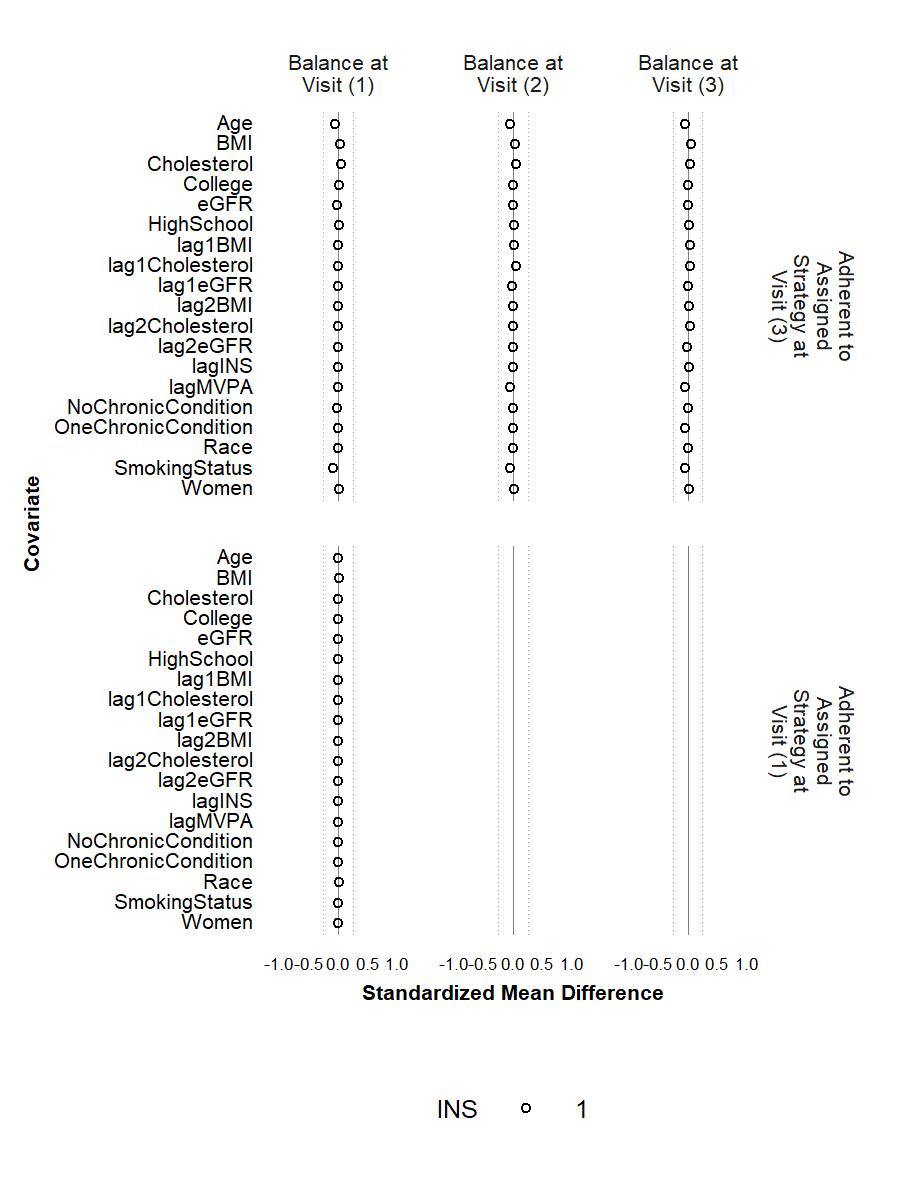

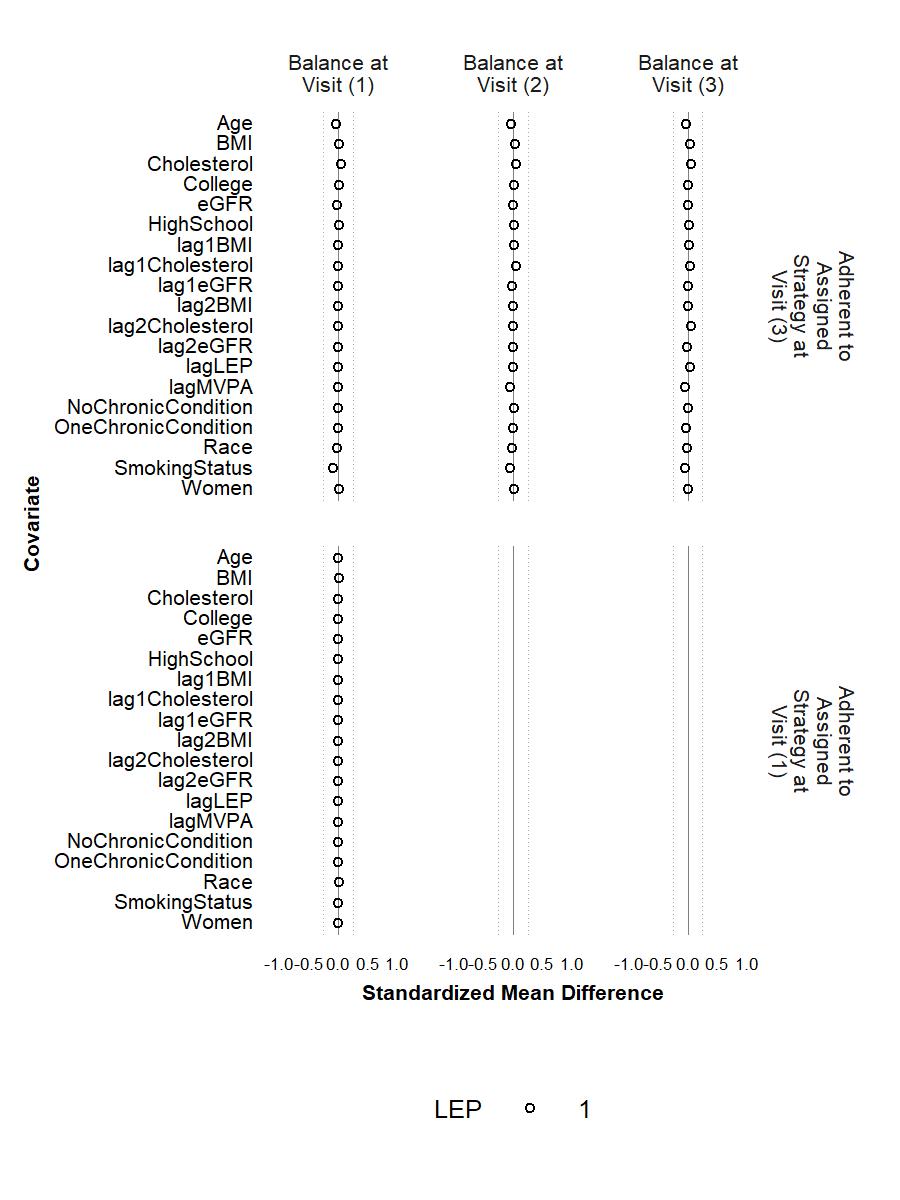

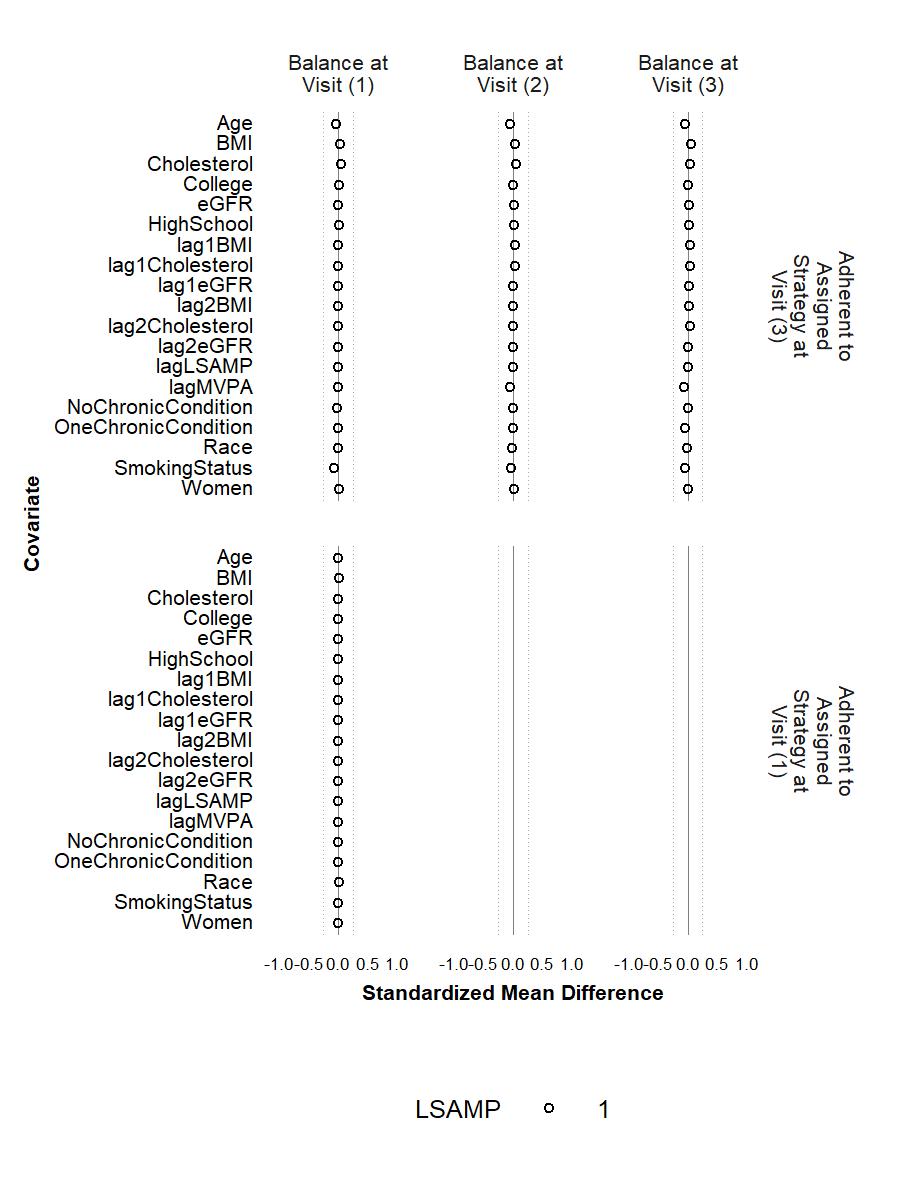

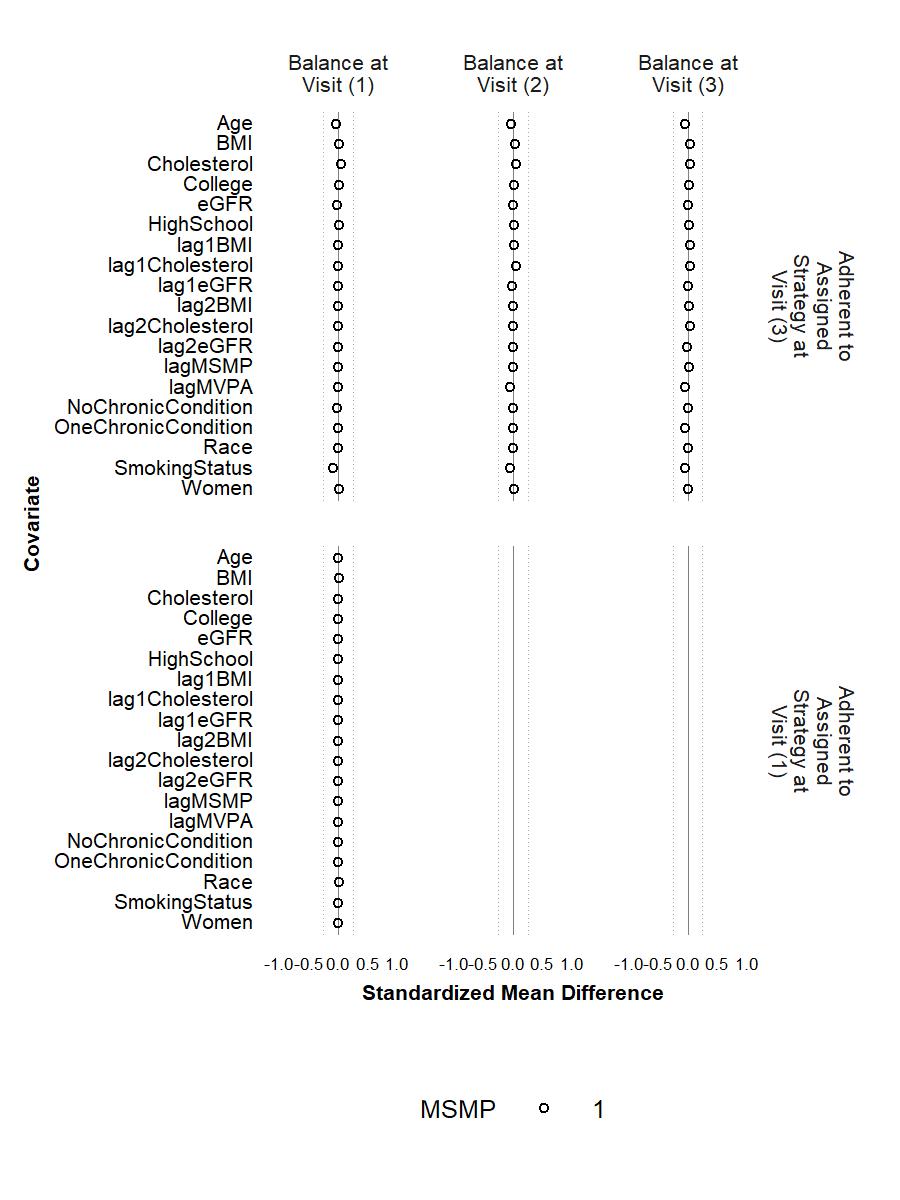

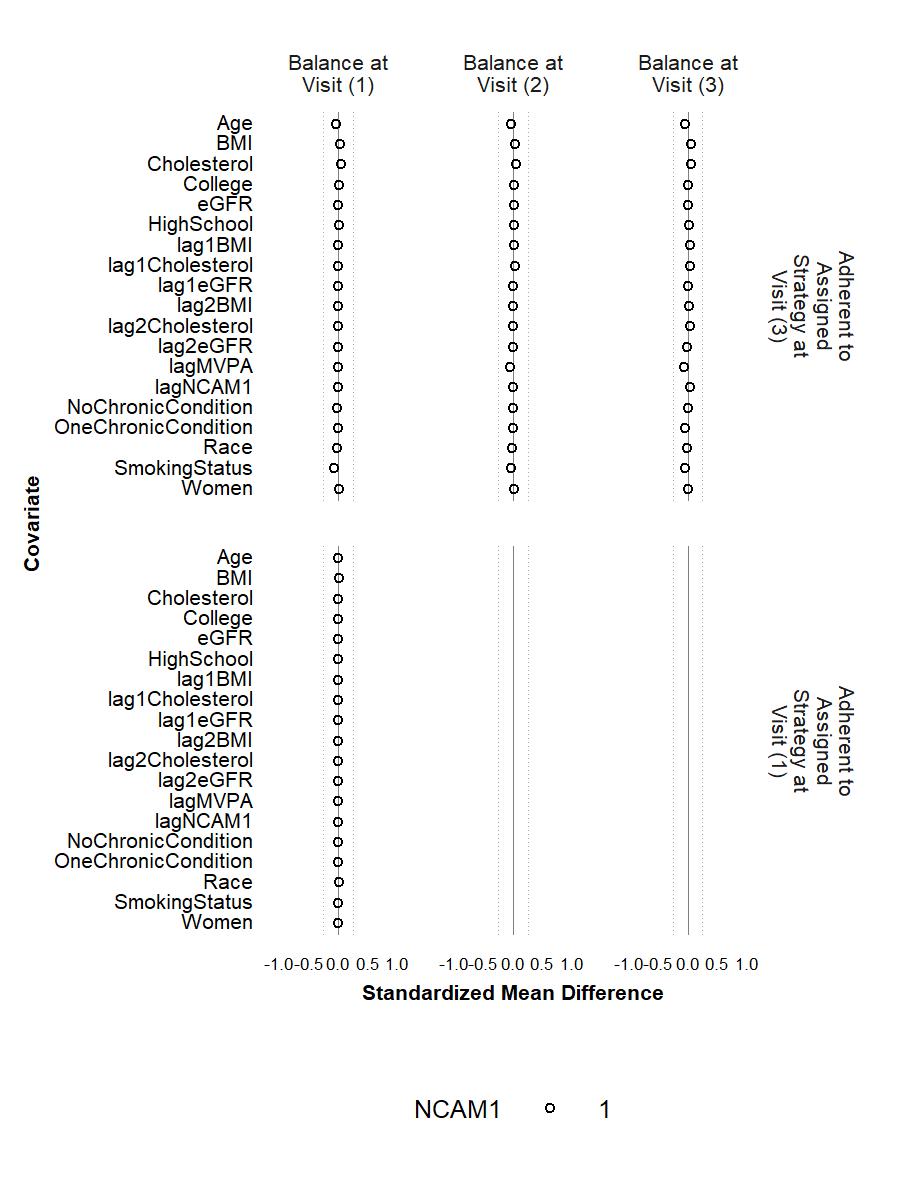

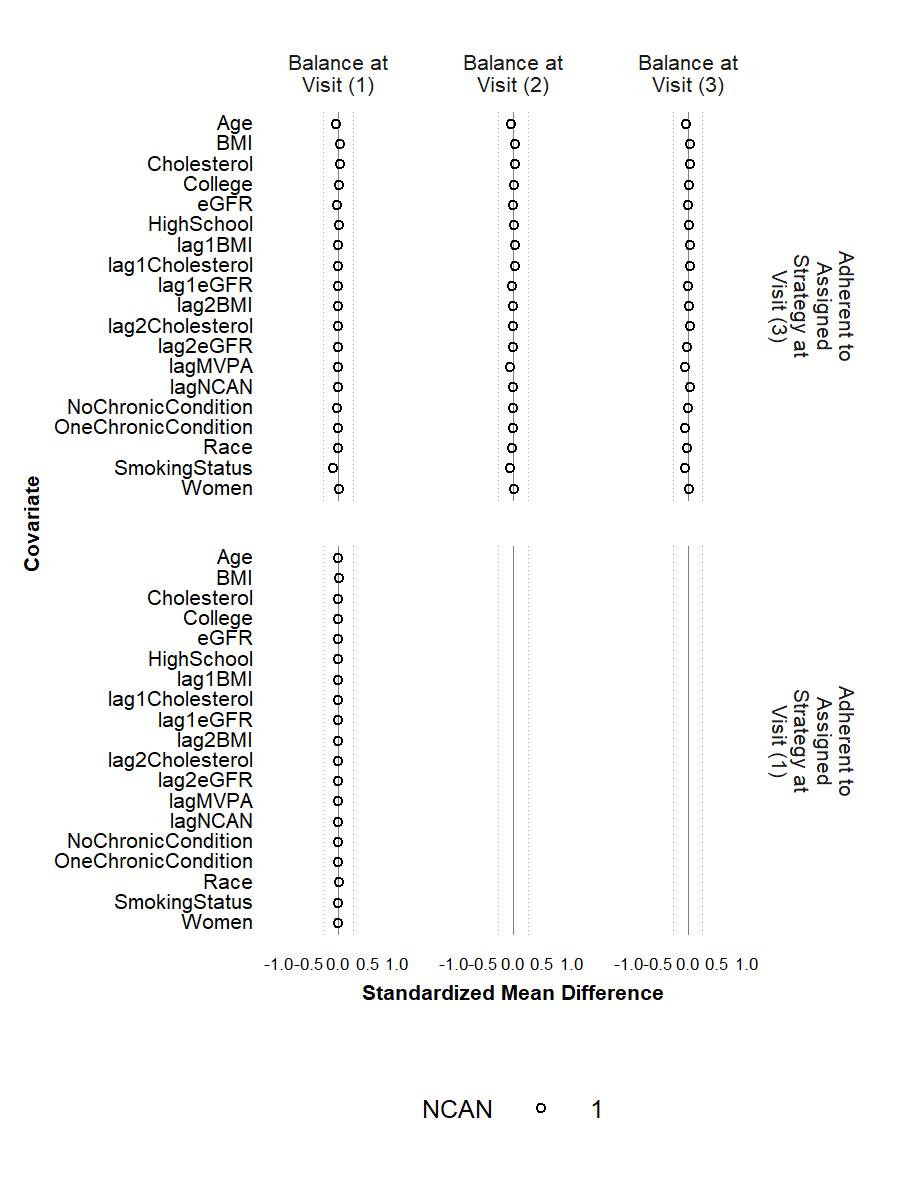

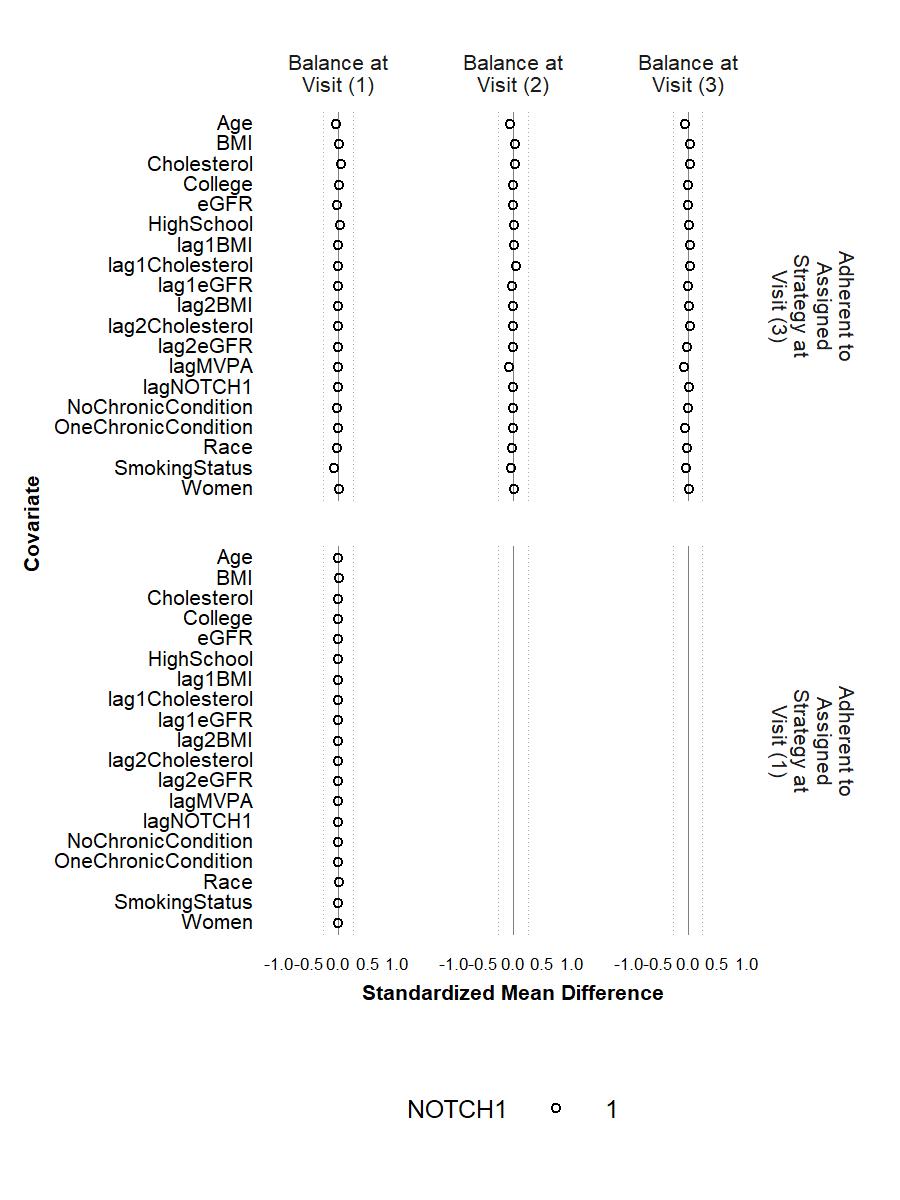

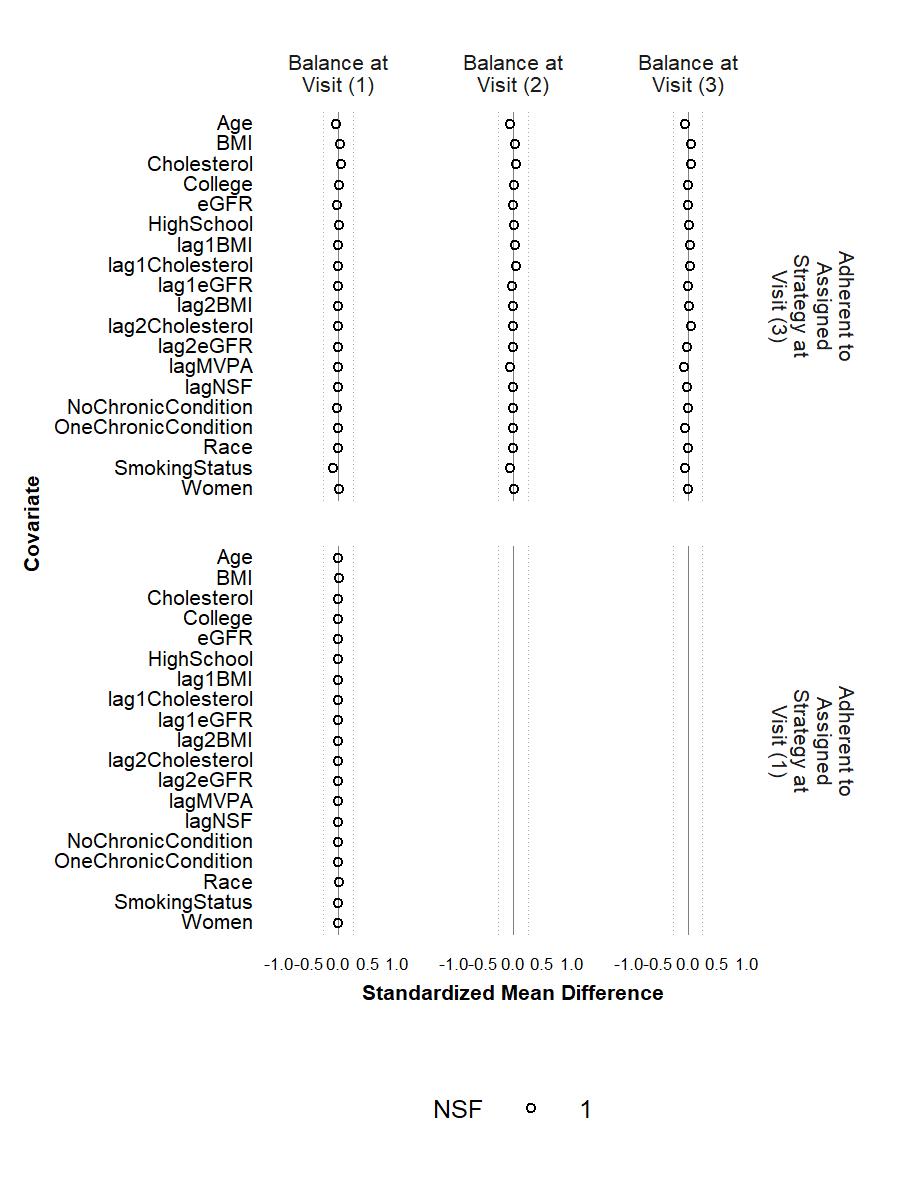

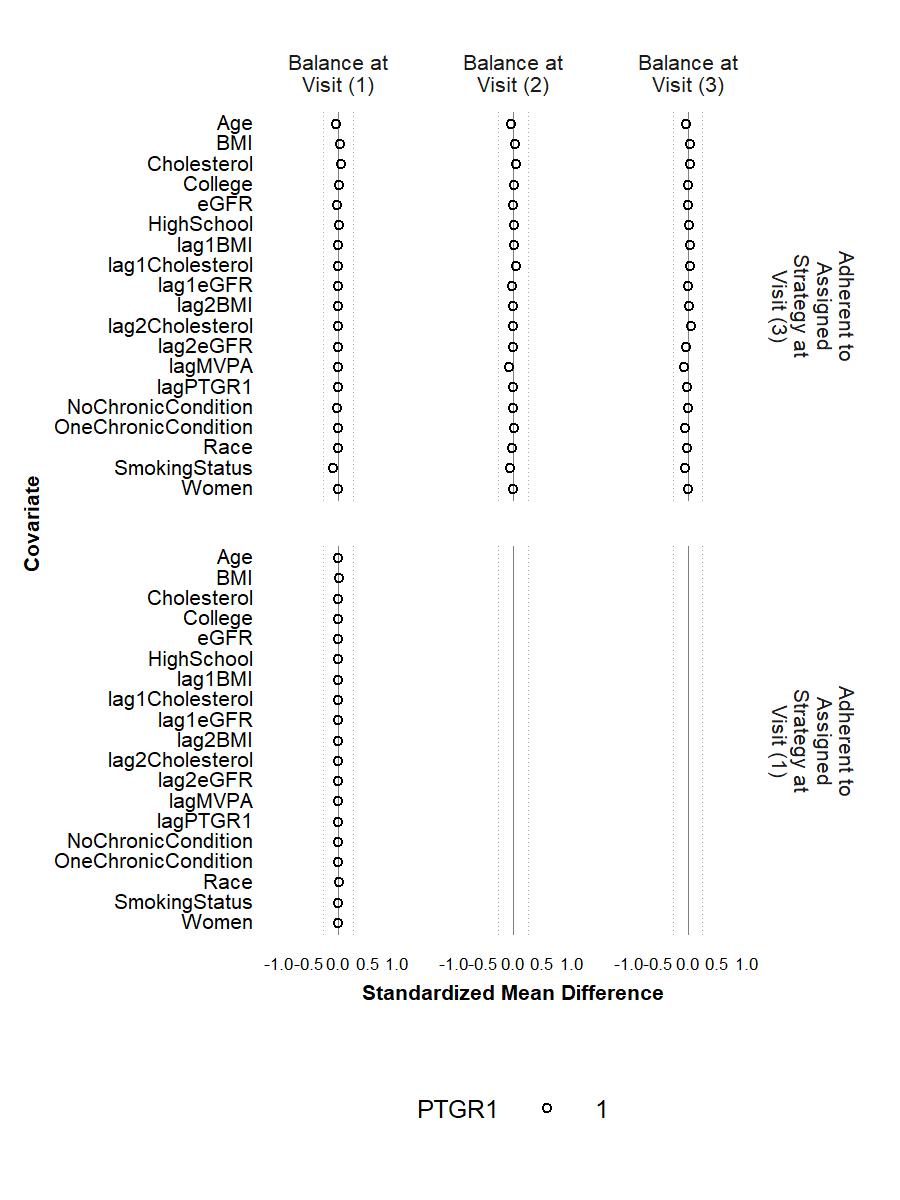

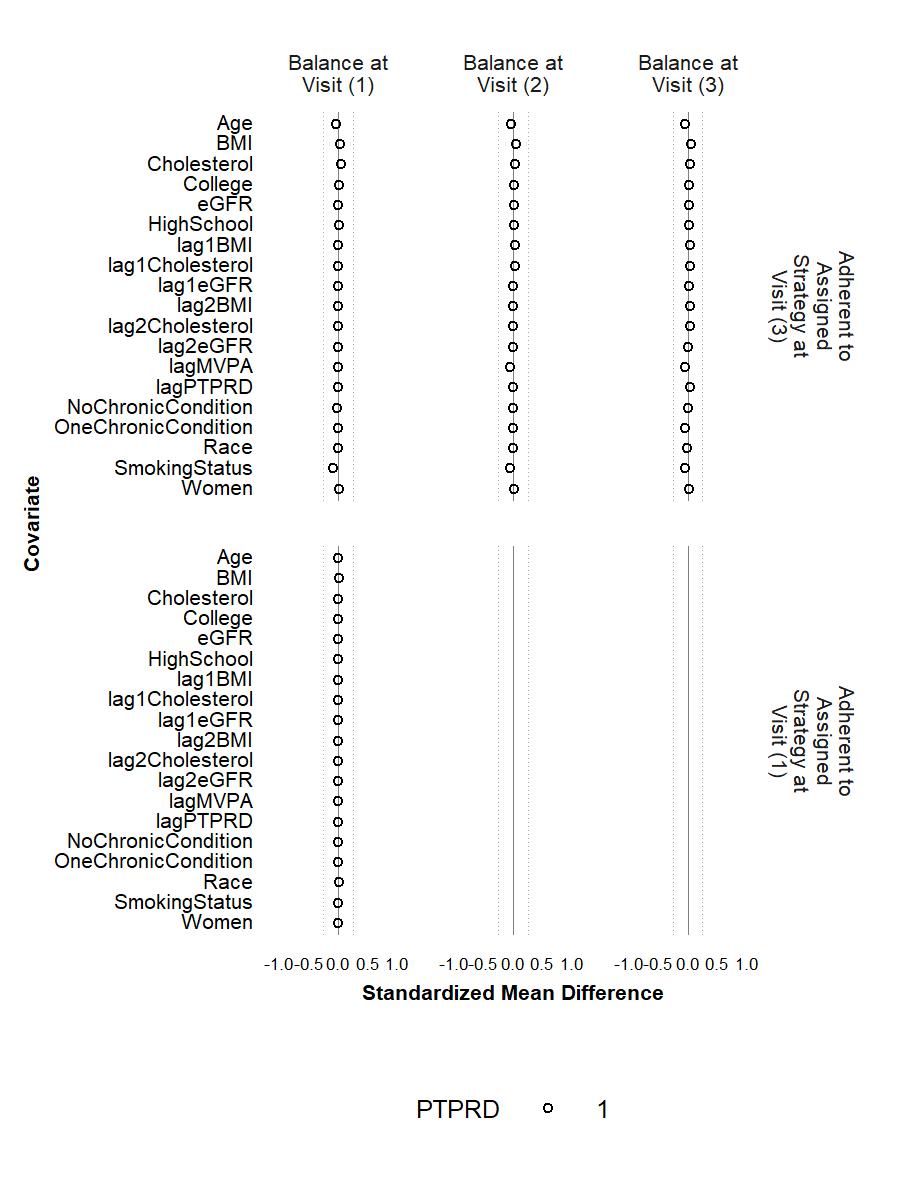

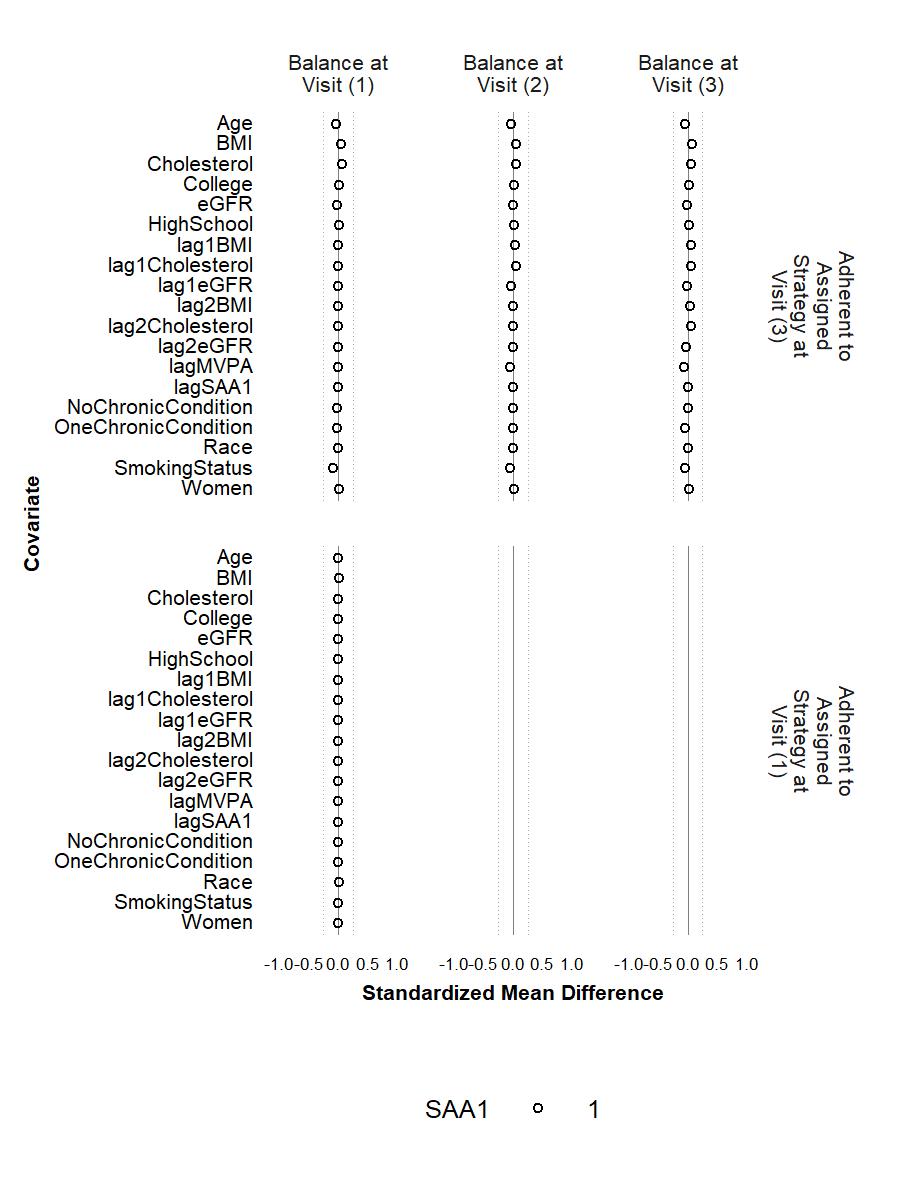

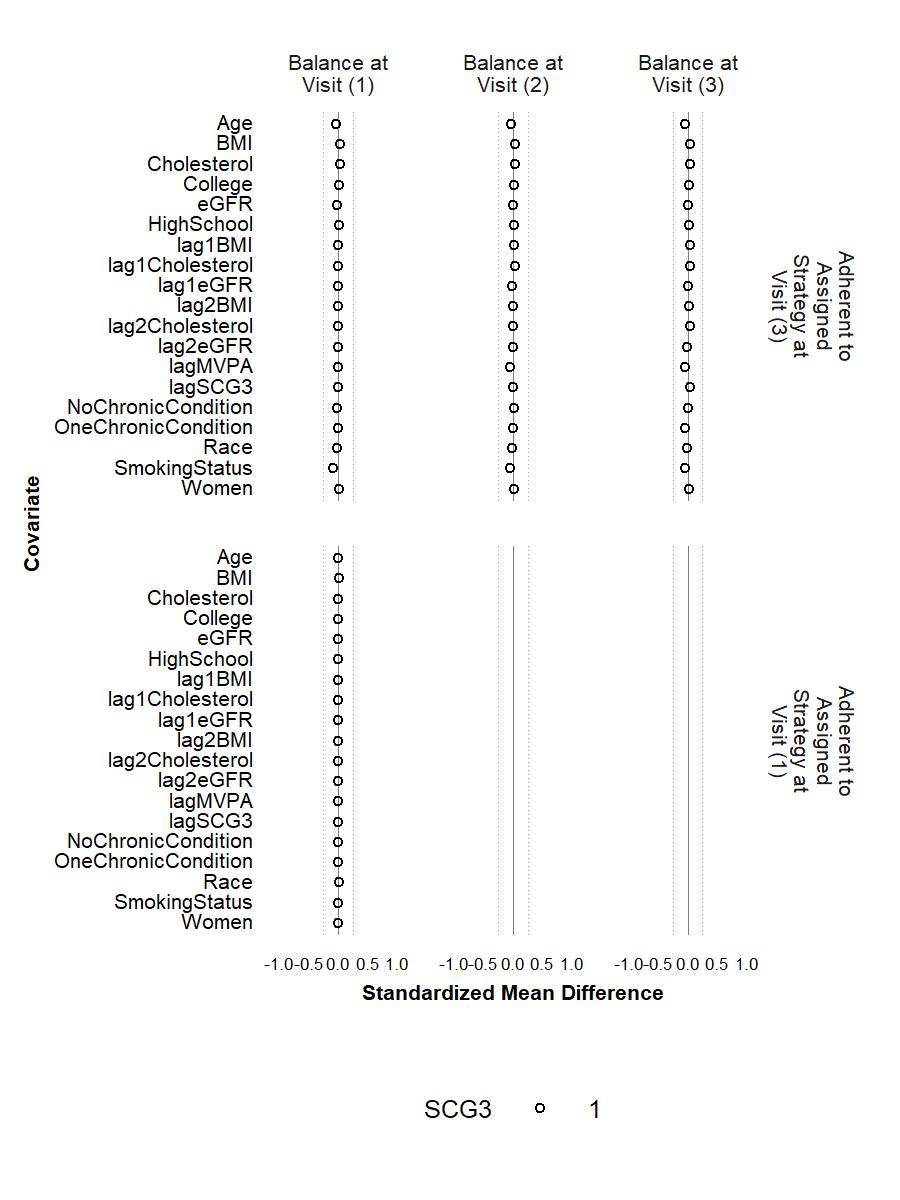

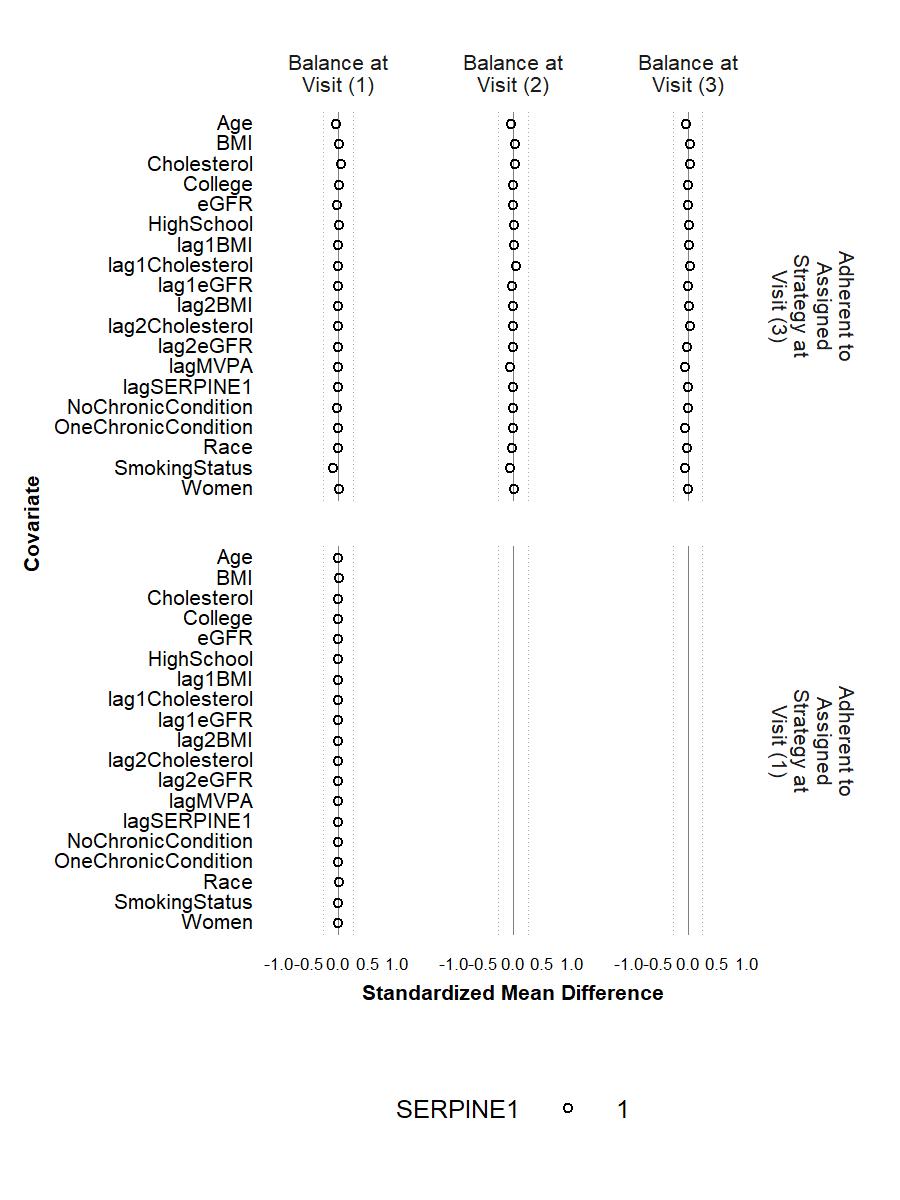

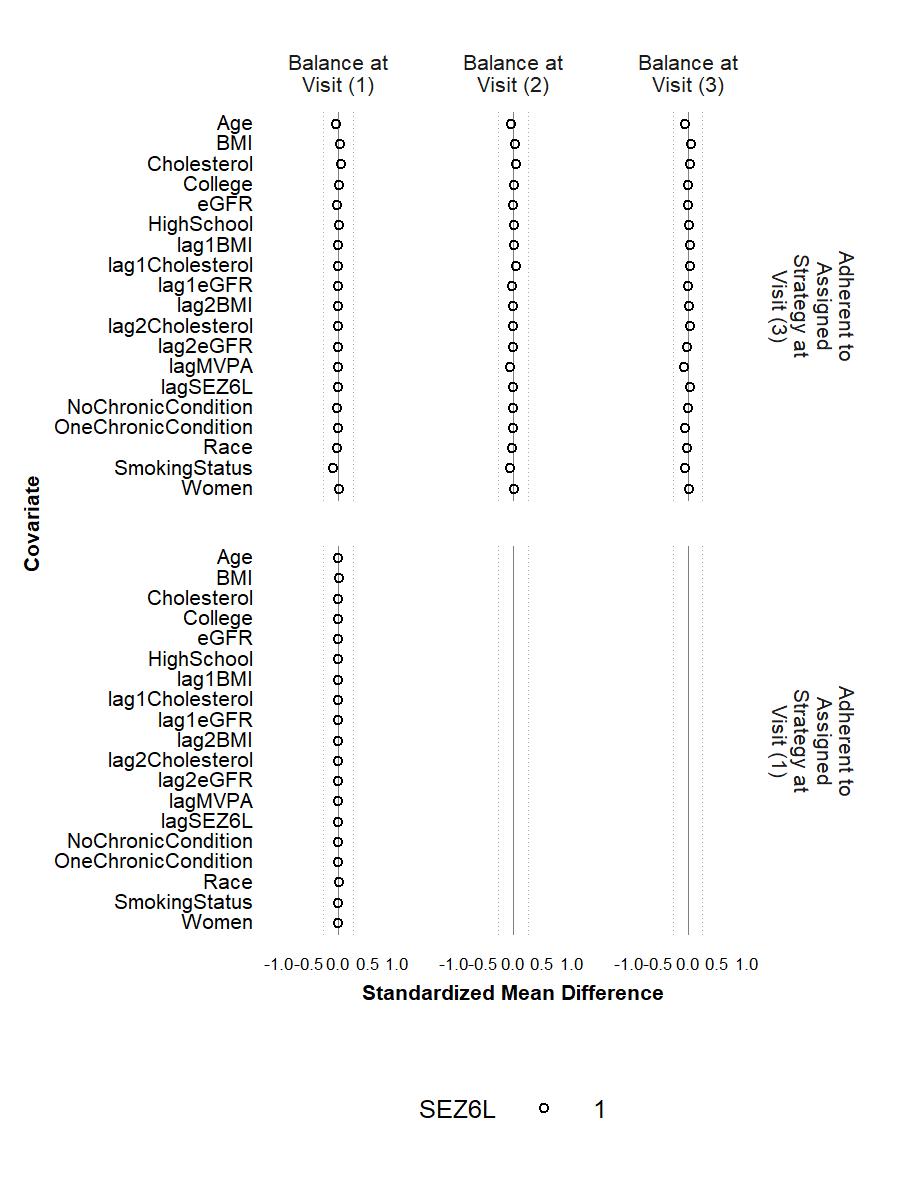

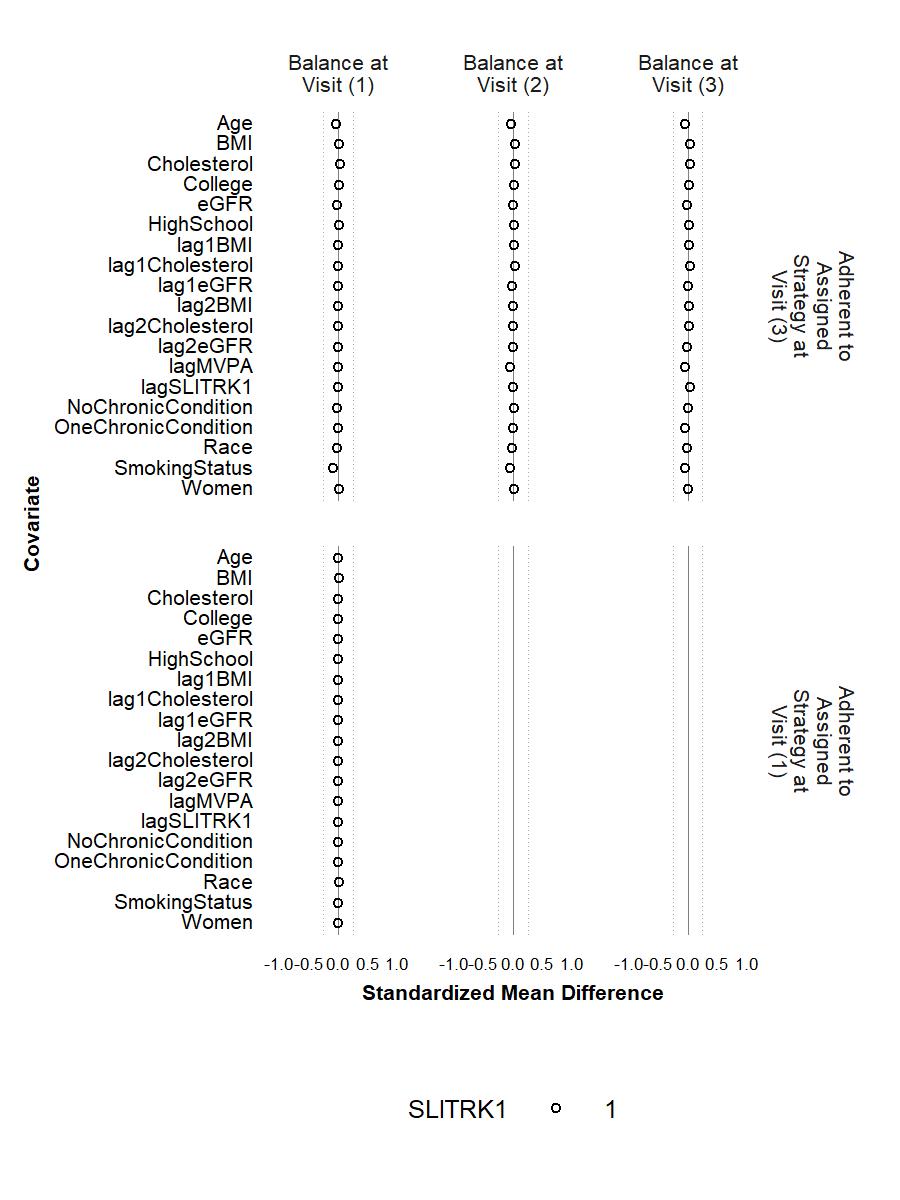

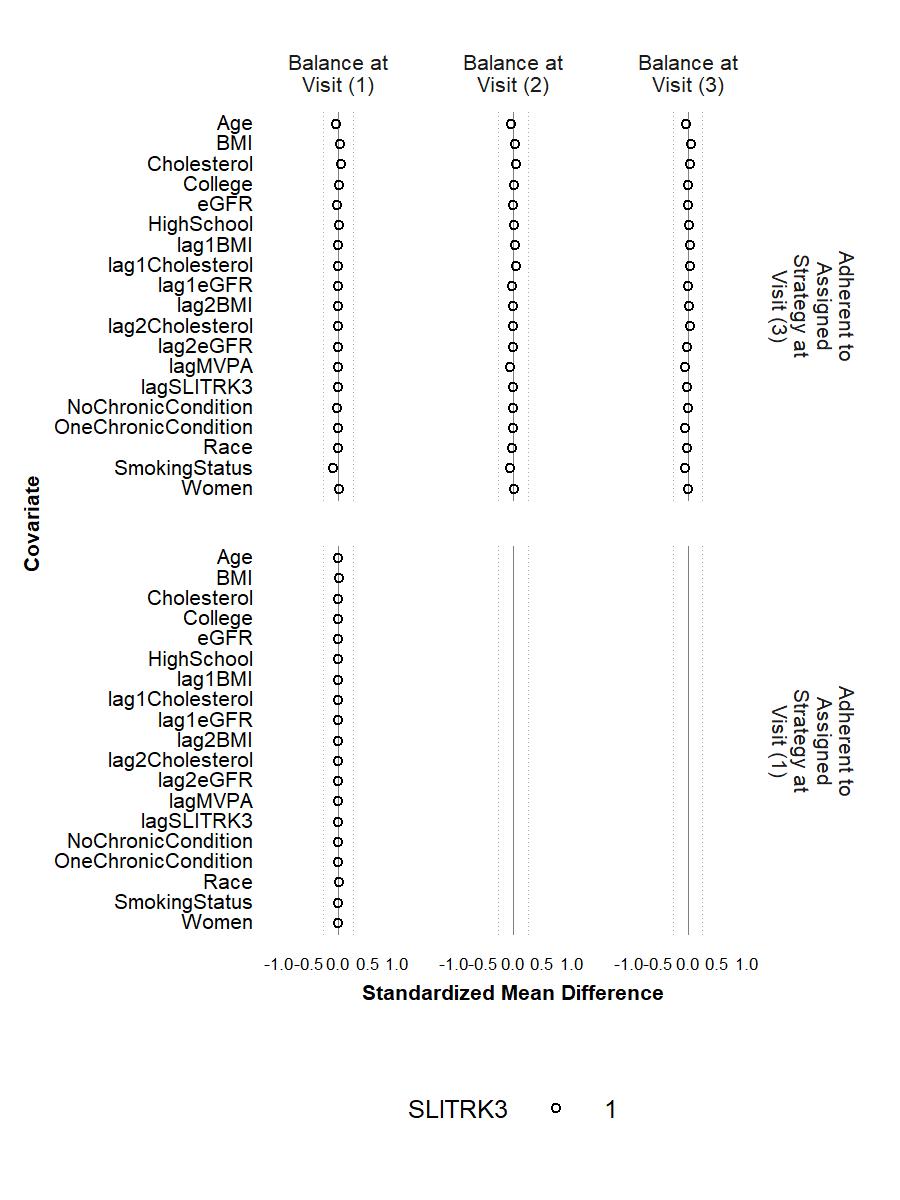

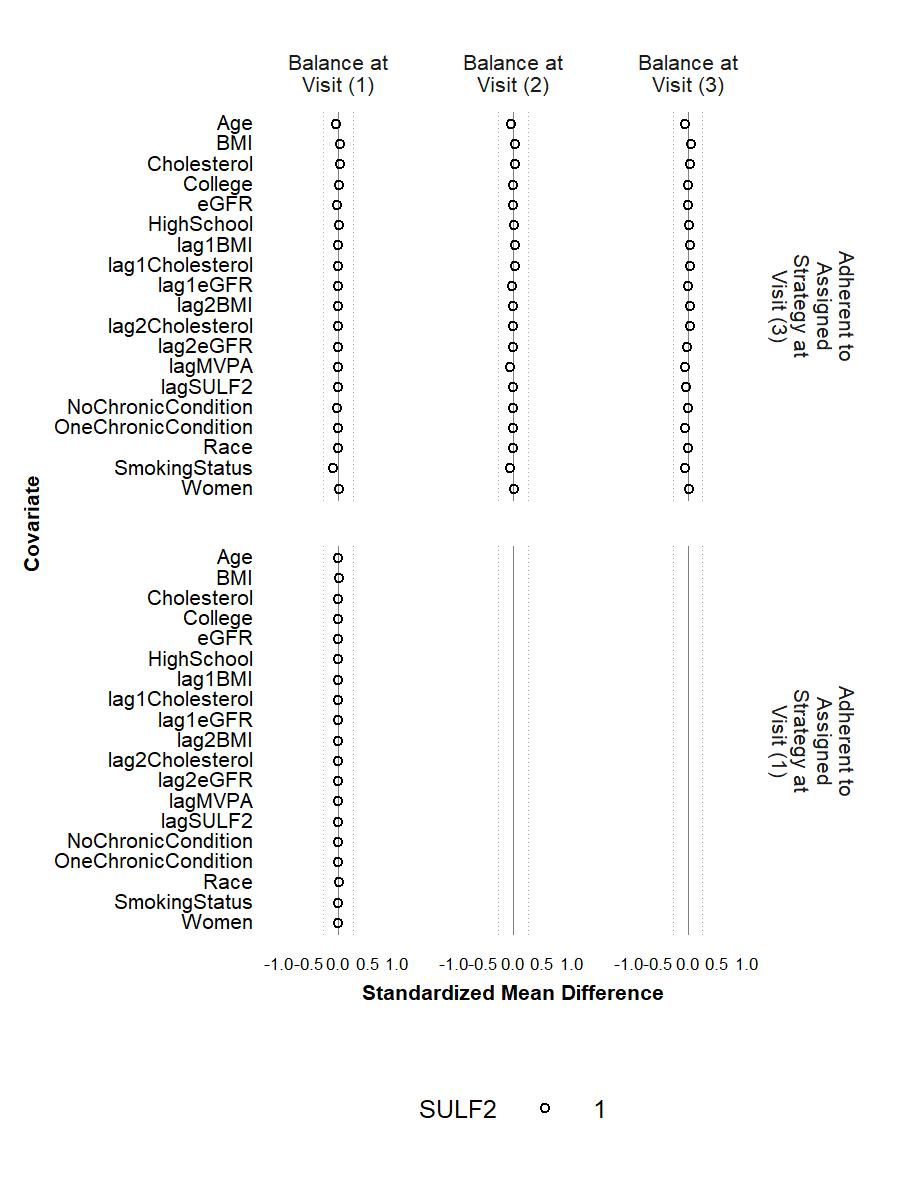

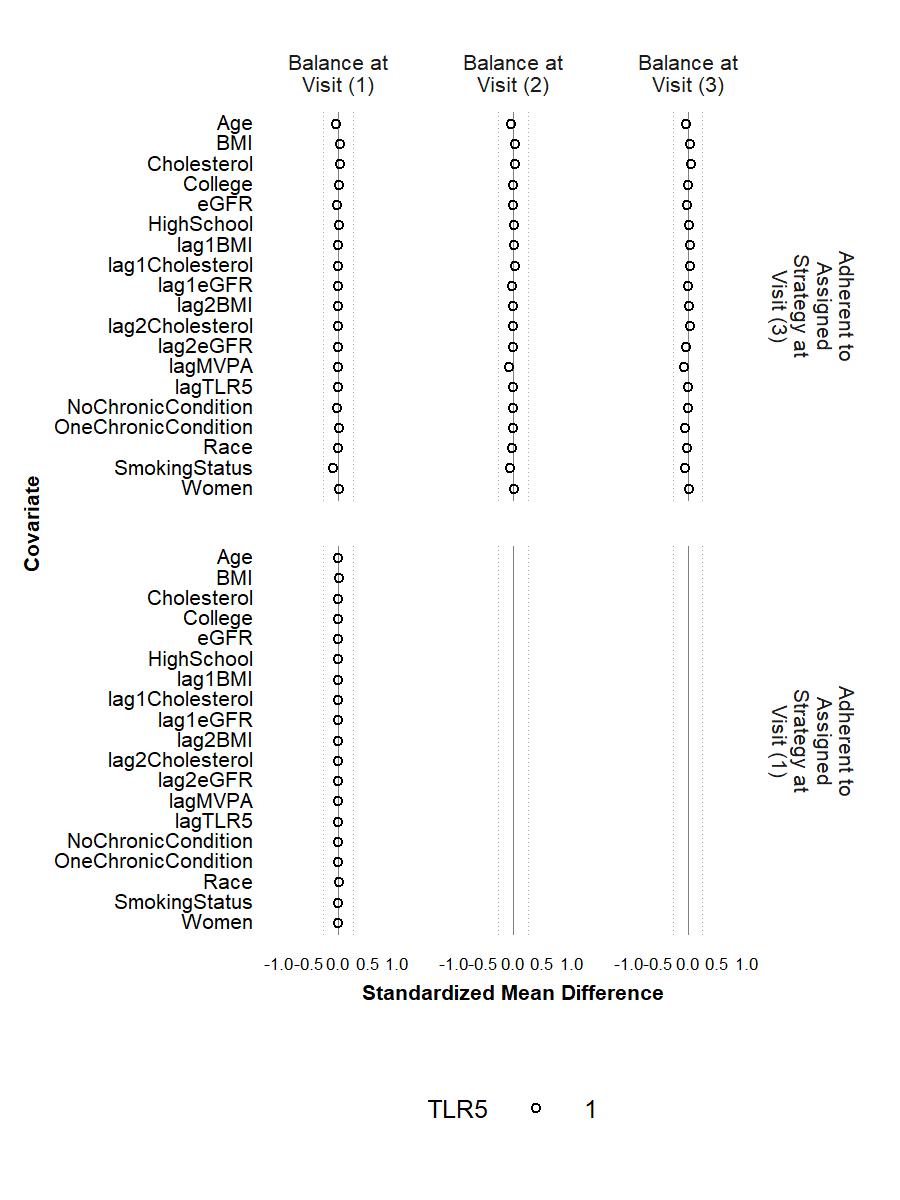

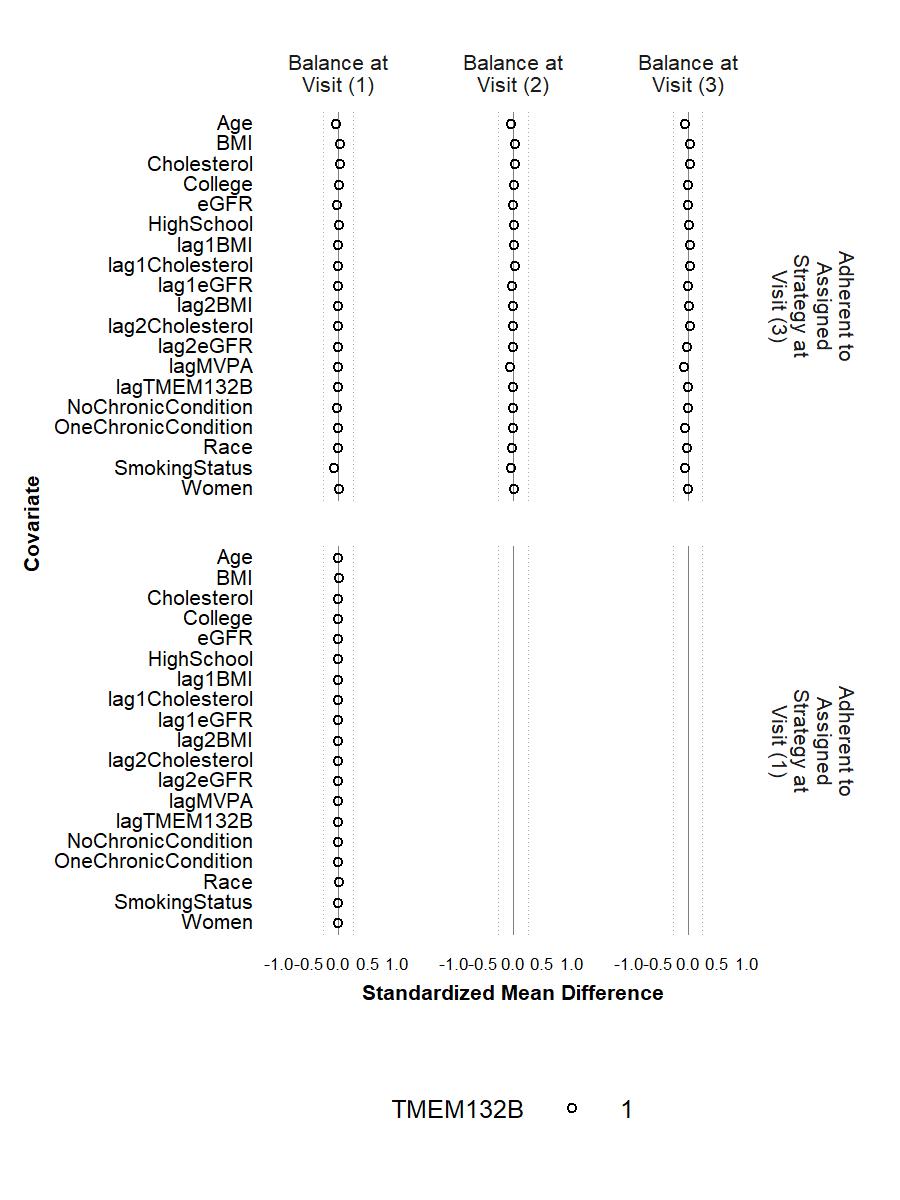

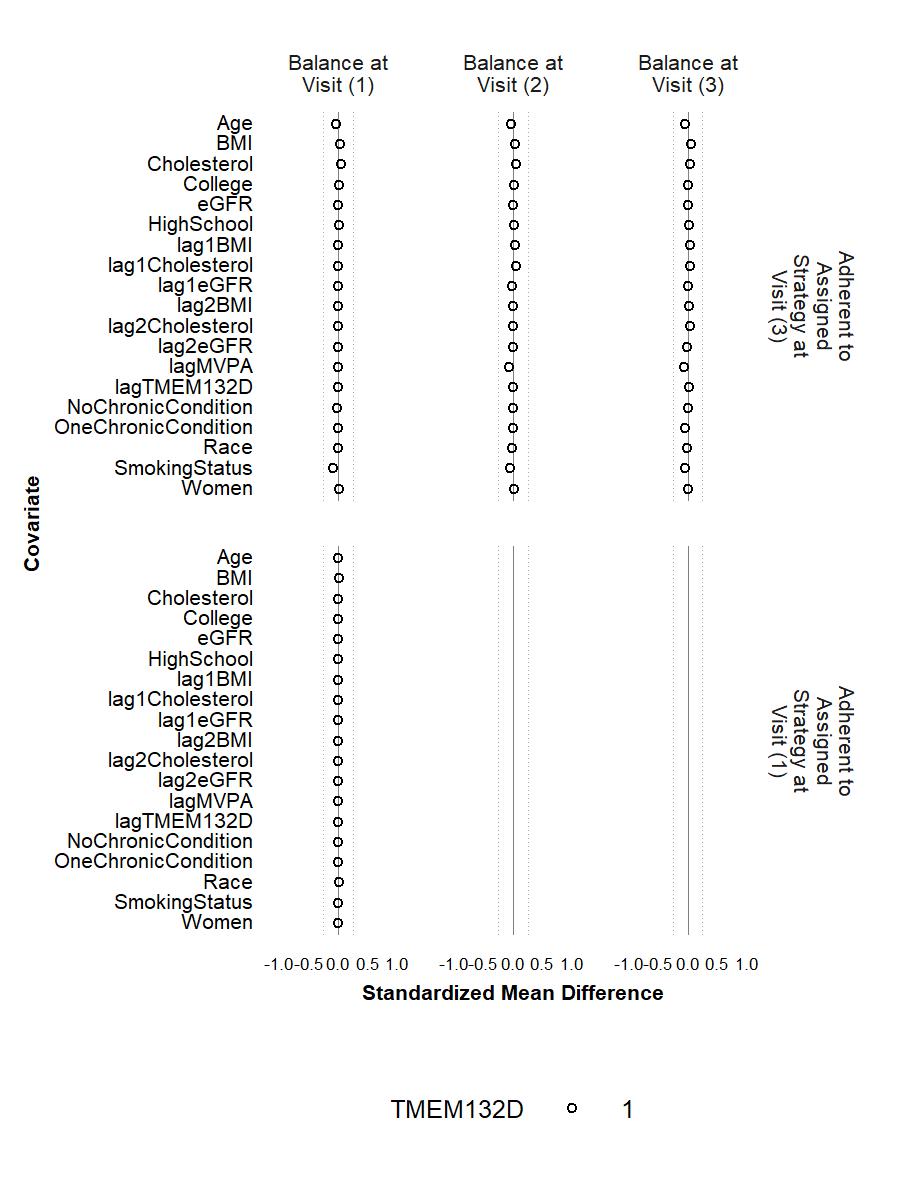

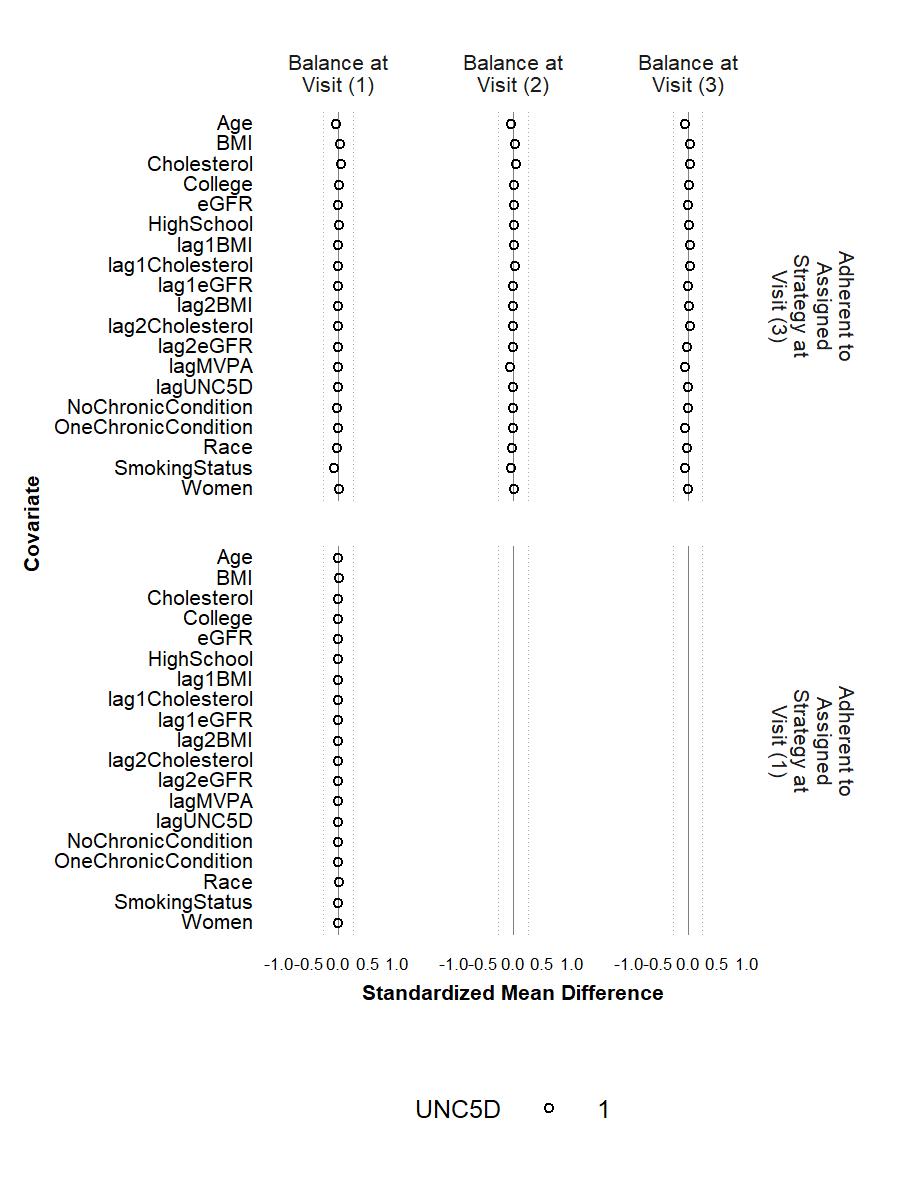

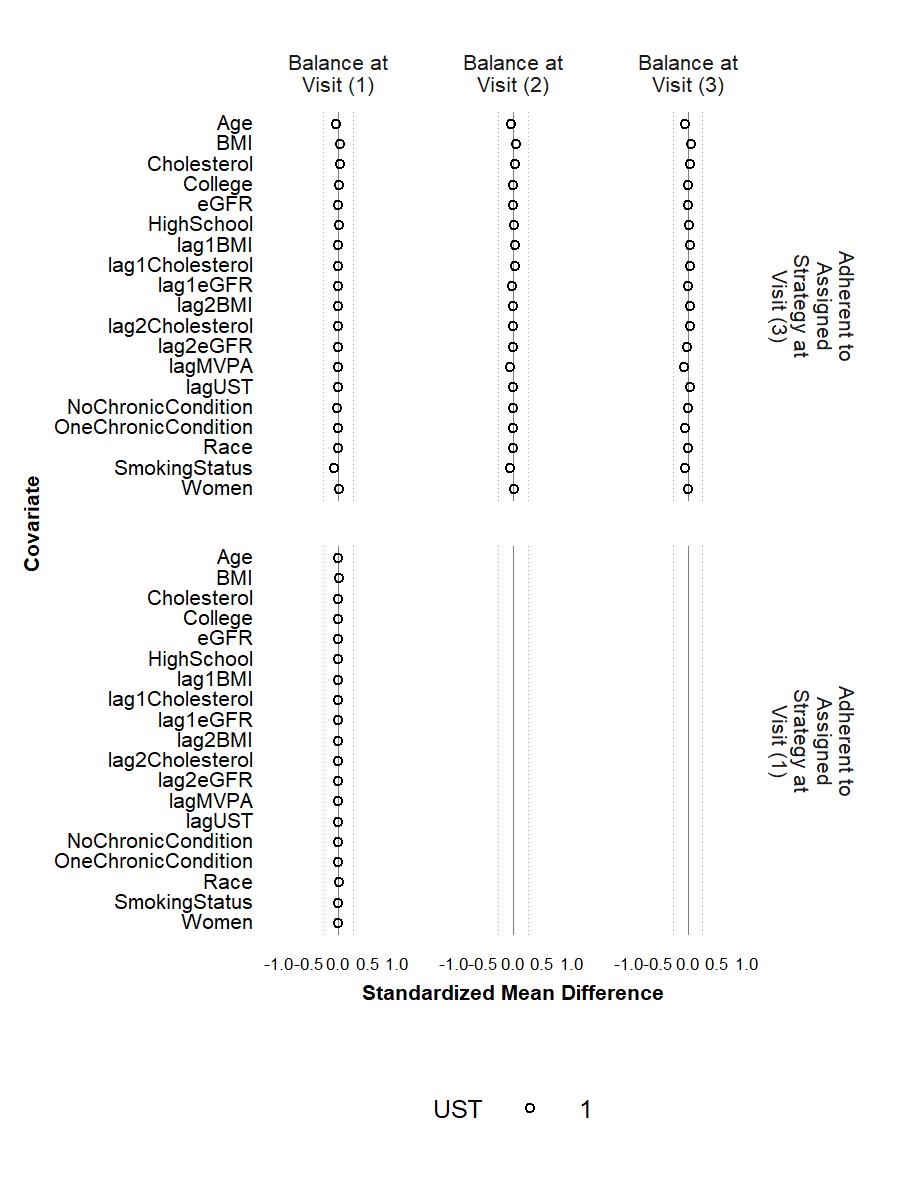

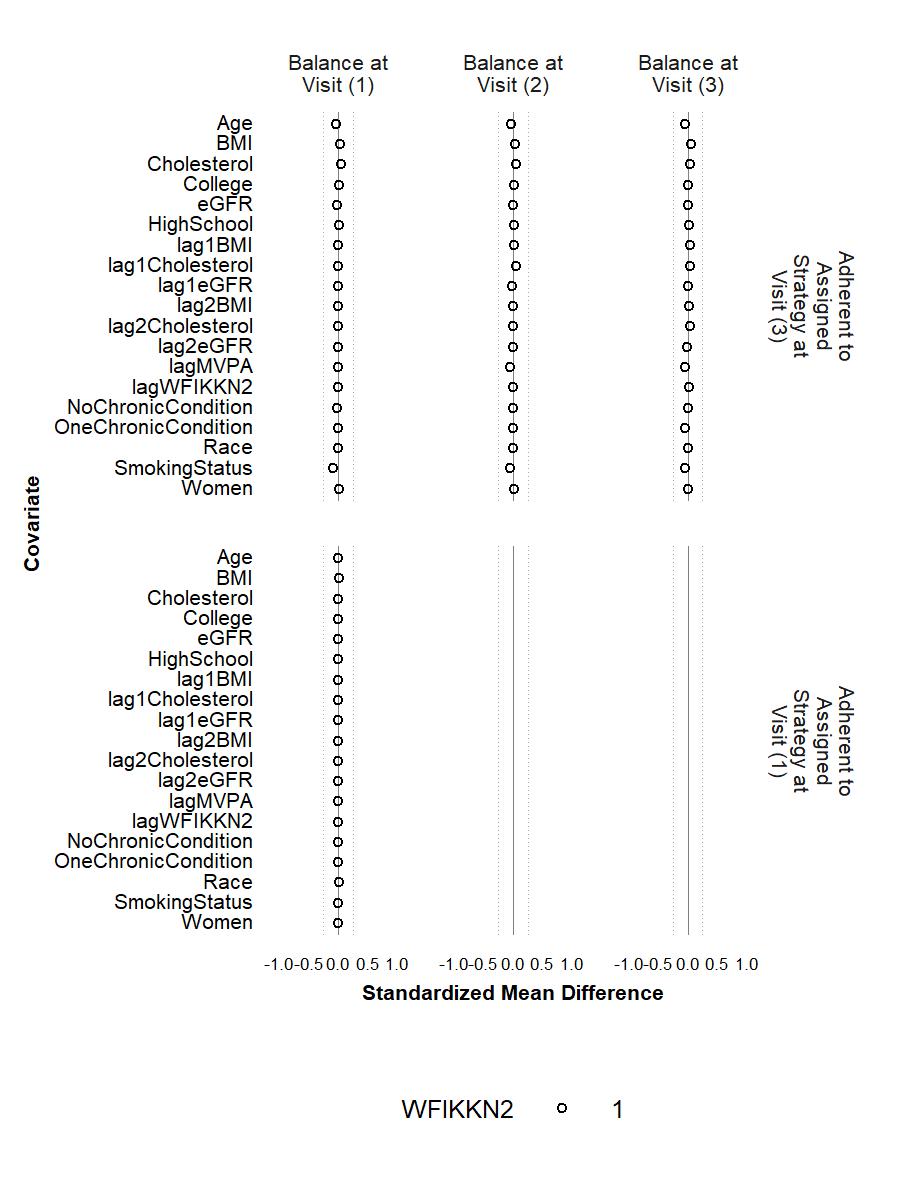

**Figure S4.** Balance of **included covariates** between participants who were lost and those who remained under study at Visits 1 and 2 after weighting with each of the proteins (except for CACNA2D3 and HITRA1) as the outcome. The circle represents the intervention strategy. The triangle represents the control strategy. The dashed lines denote ±0.25 SD.

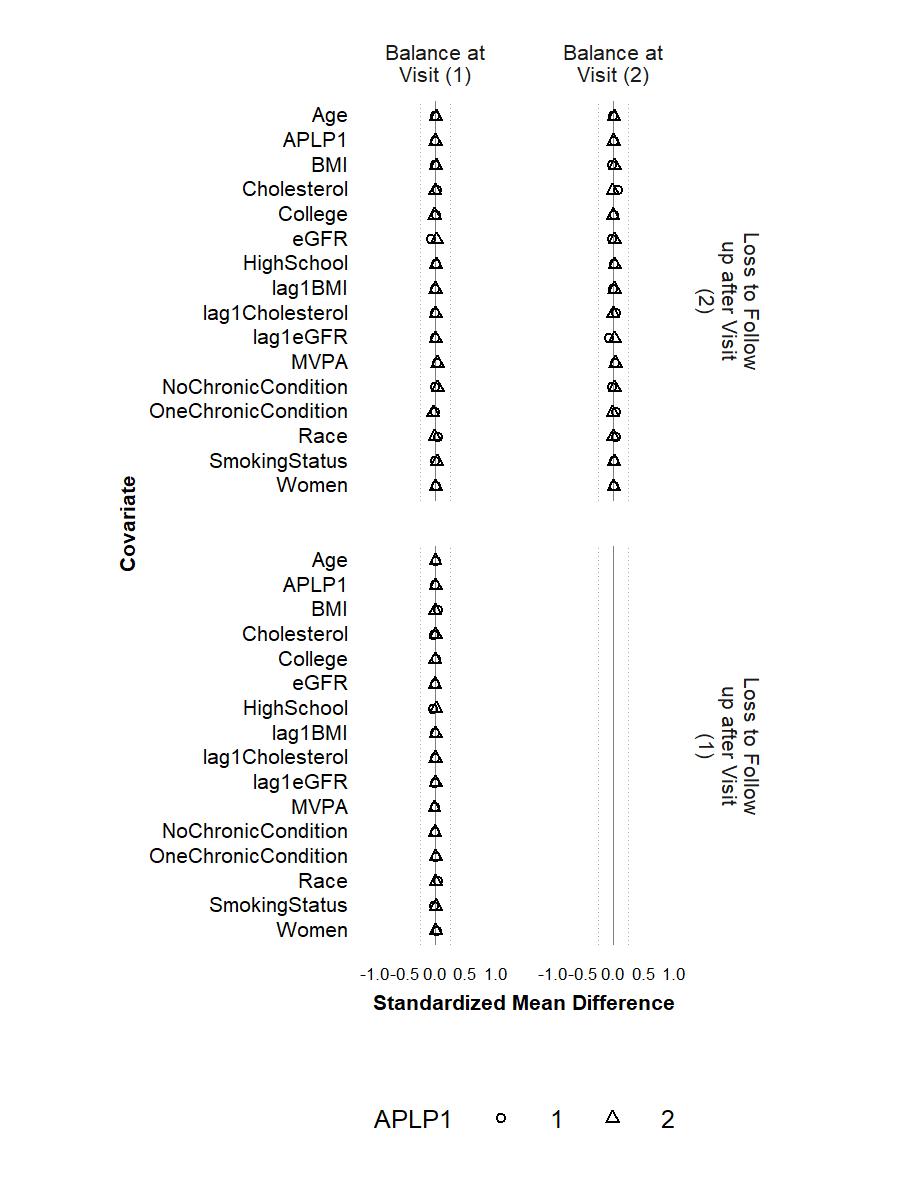

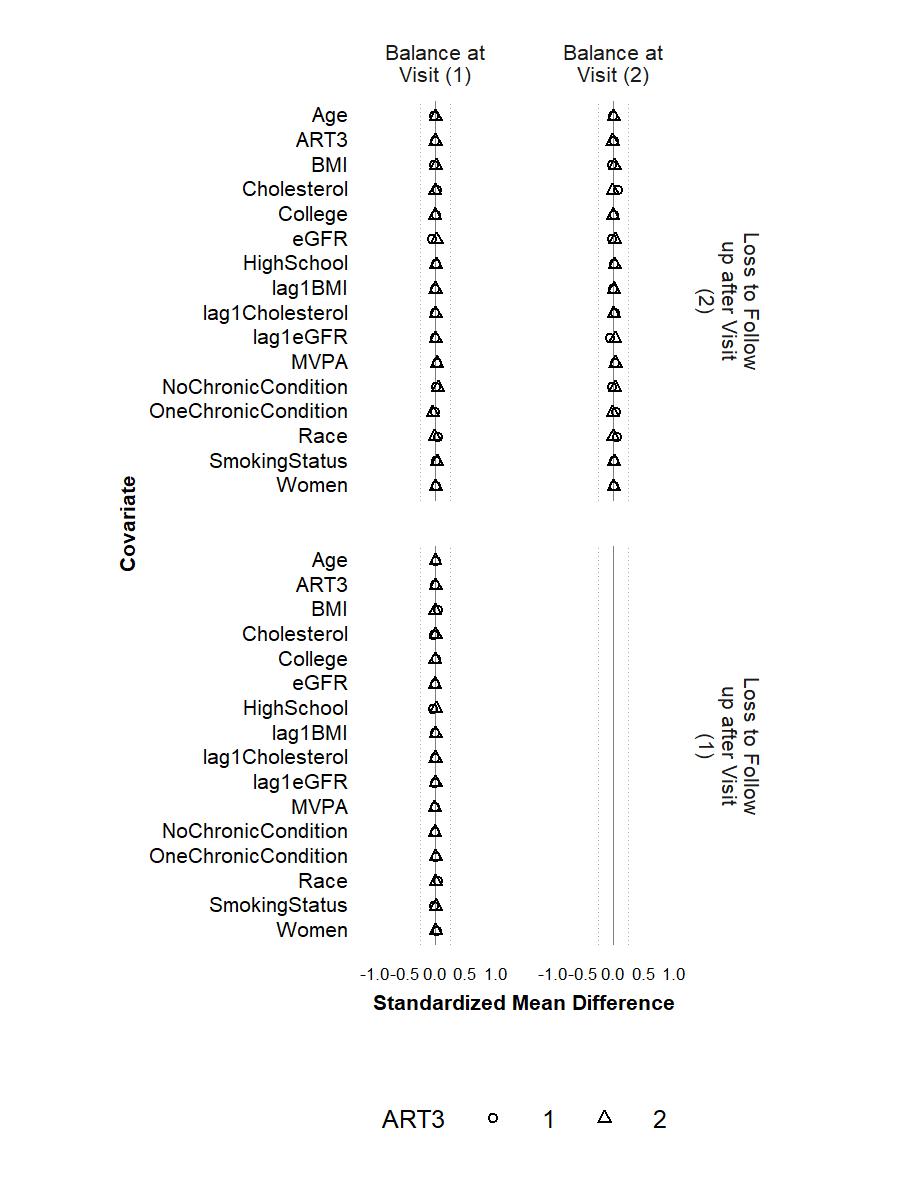

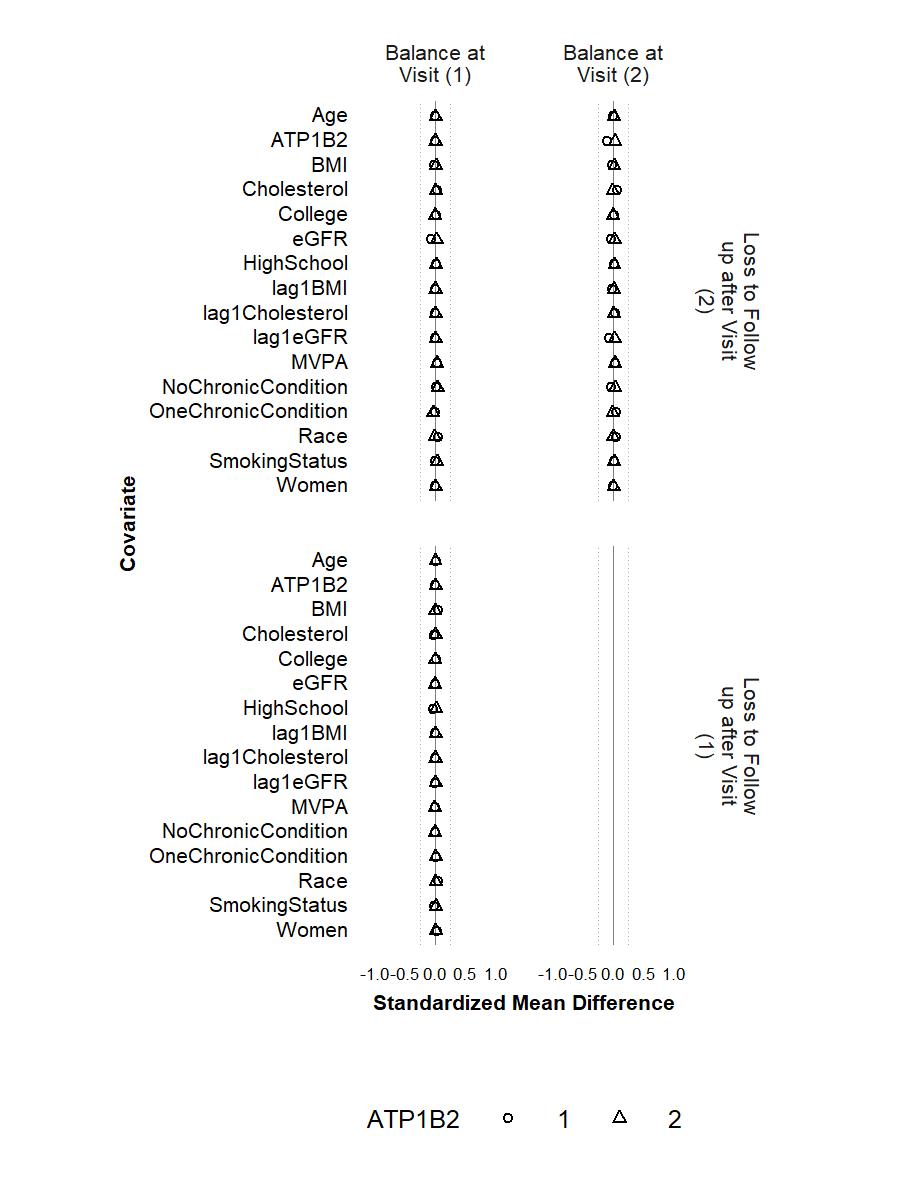

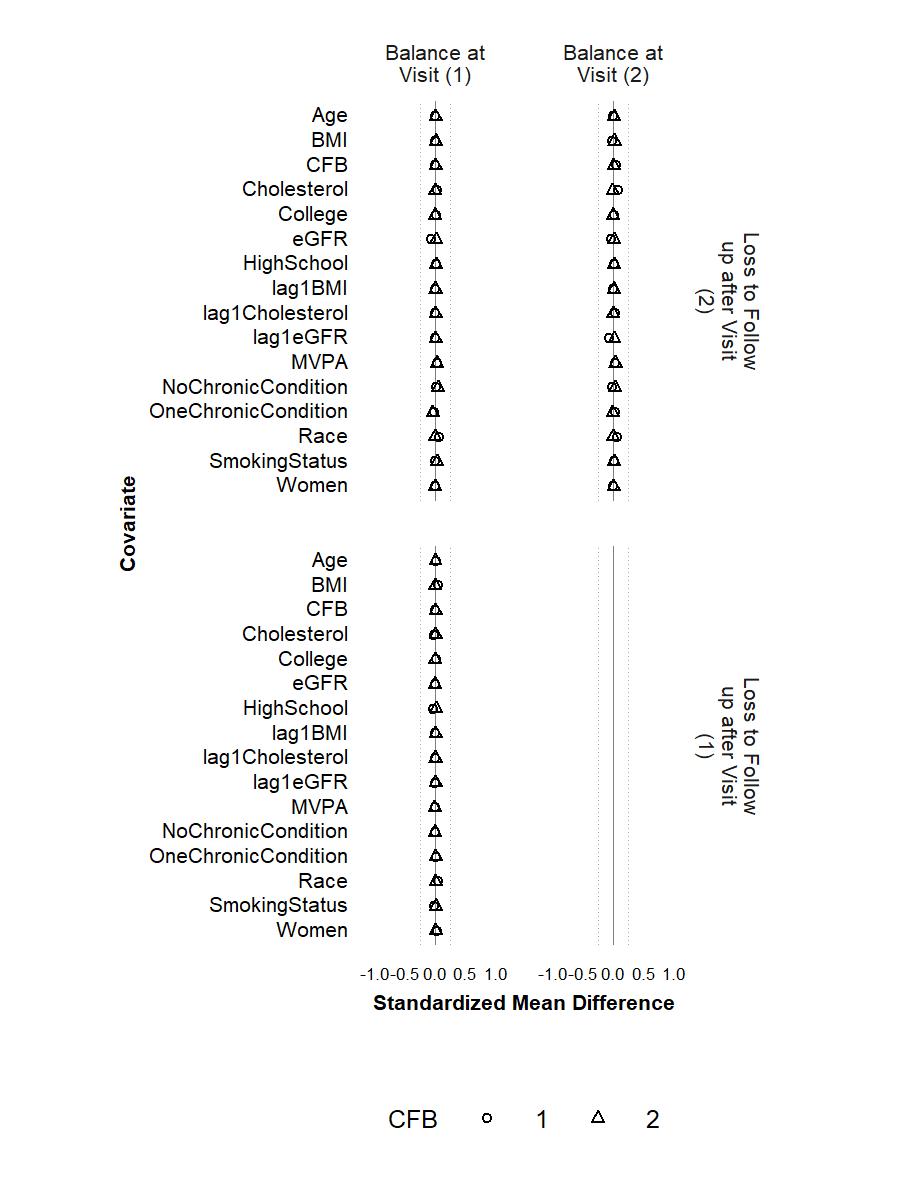

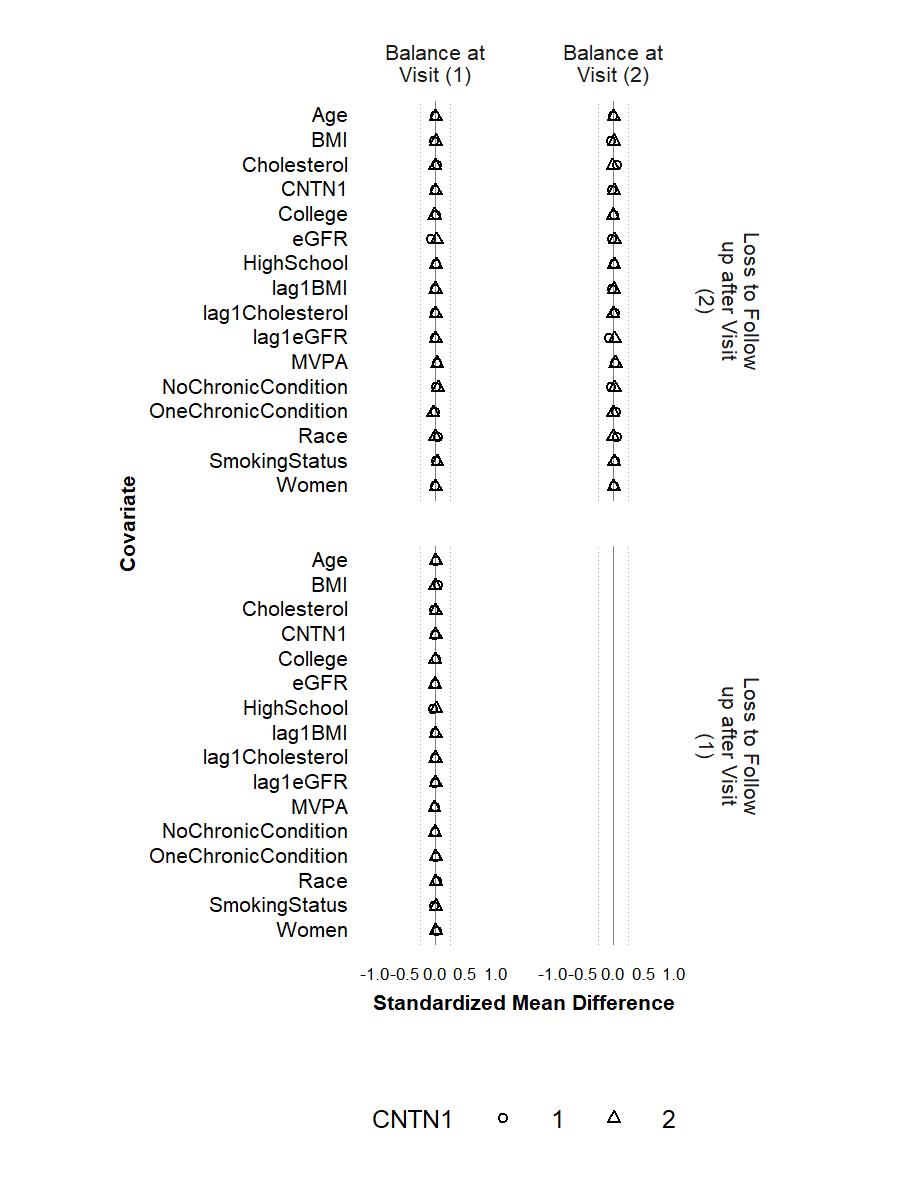

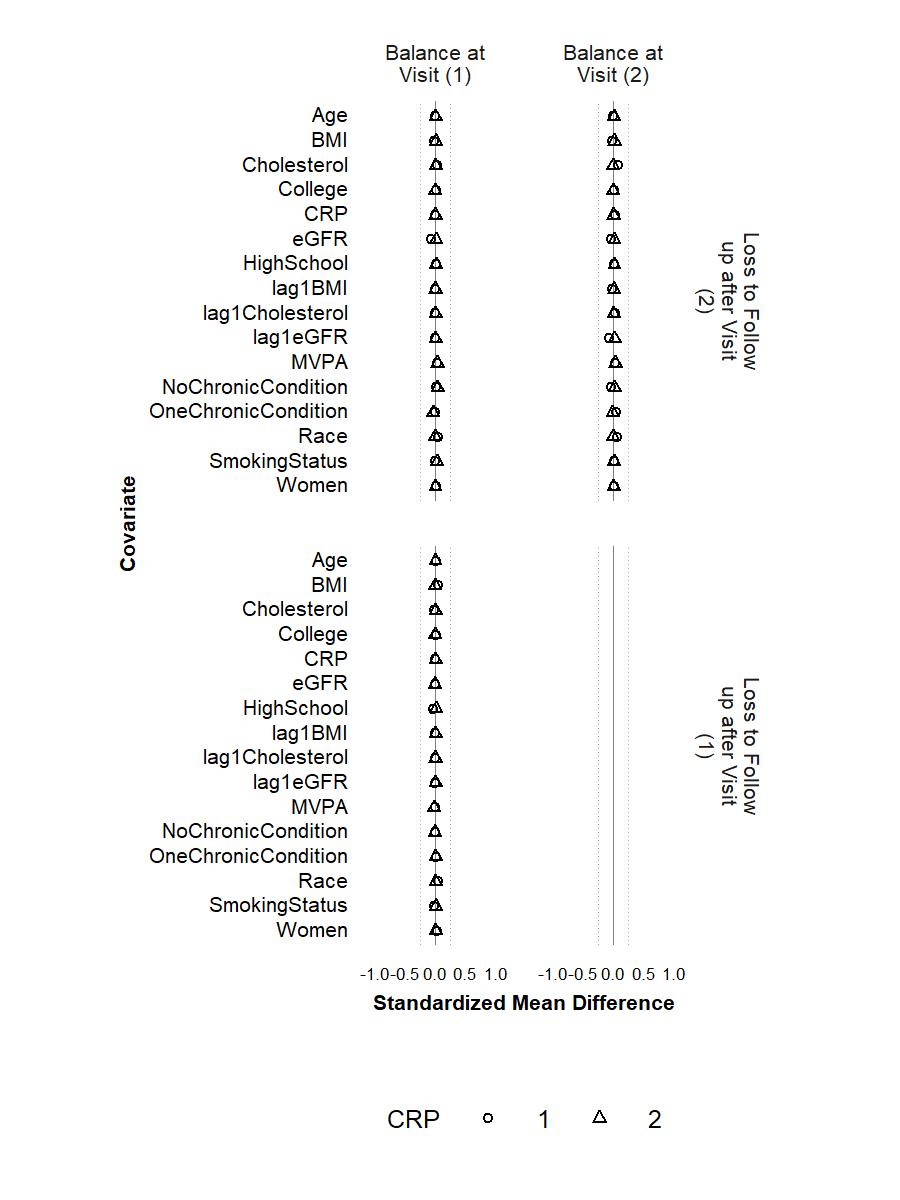

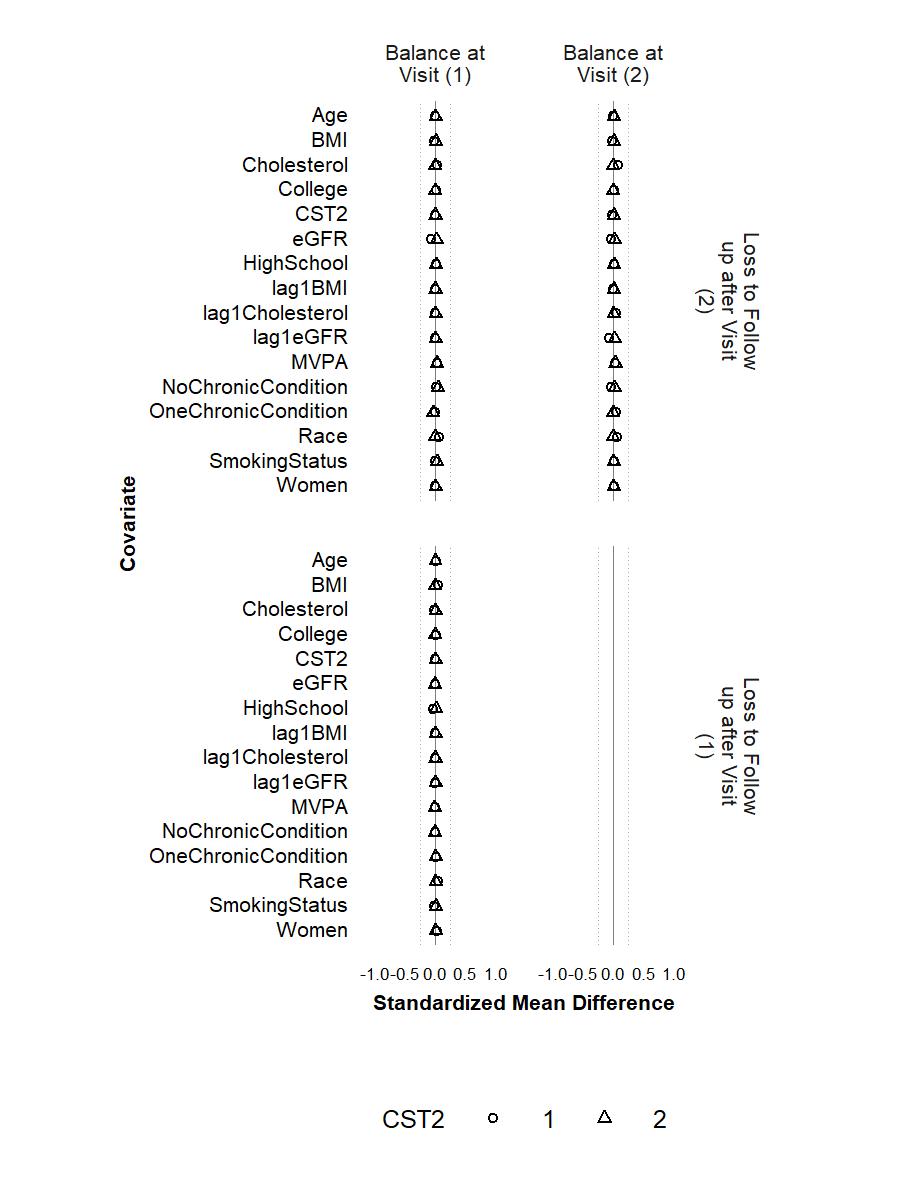

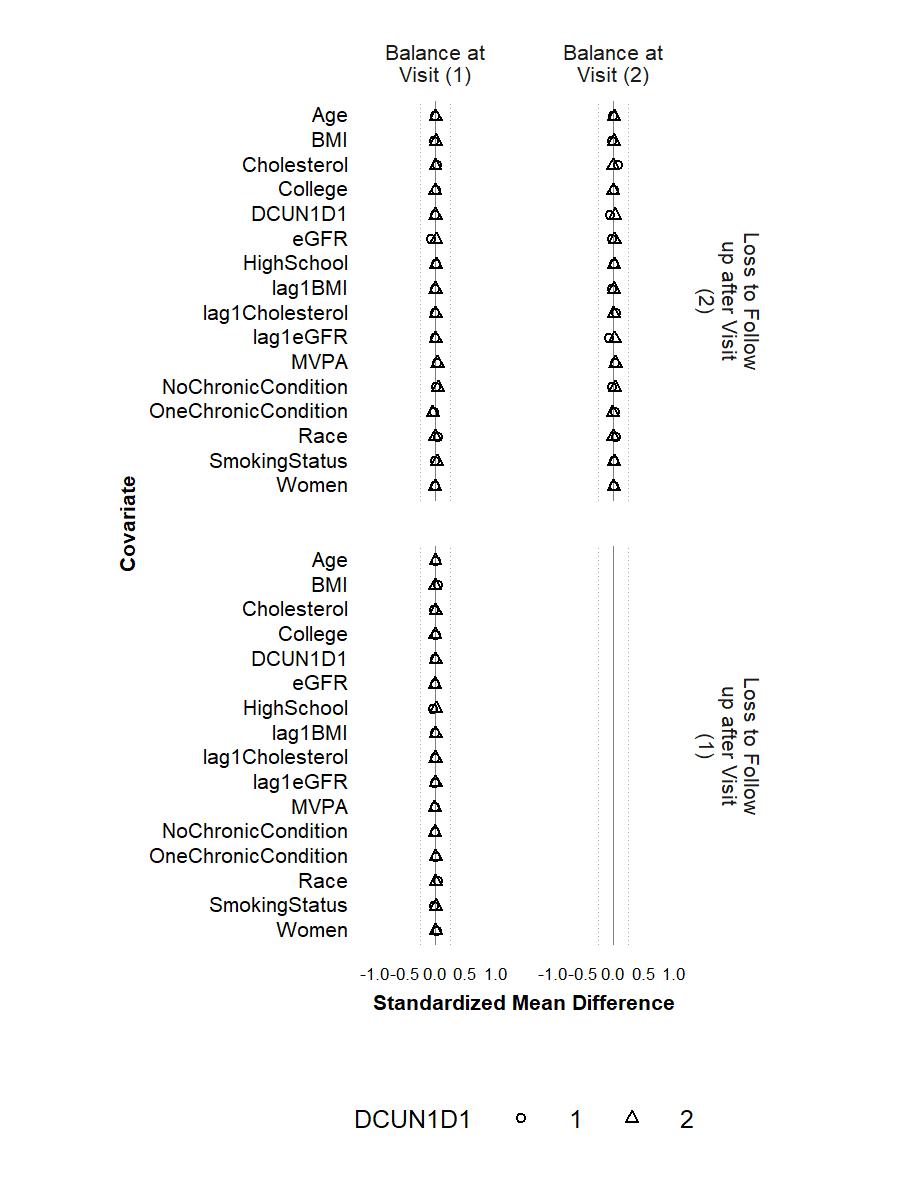

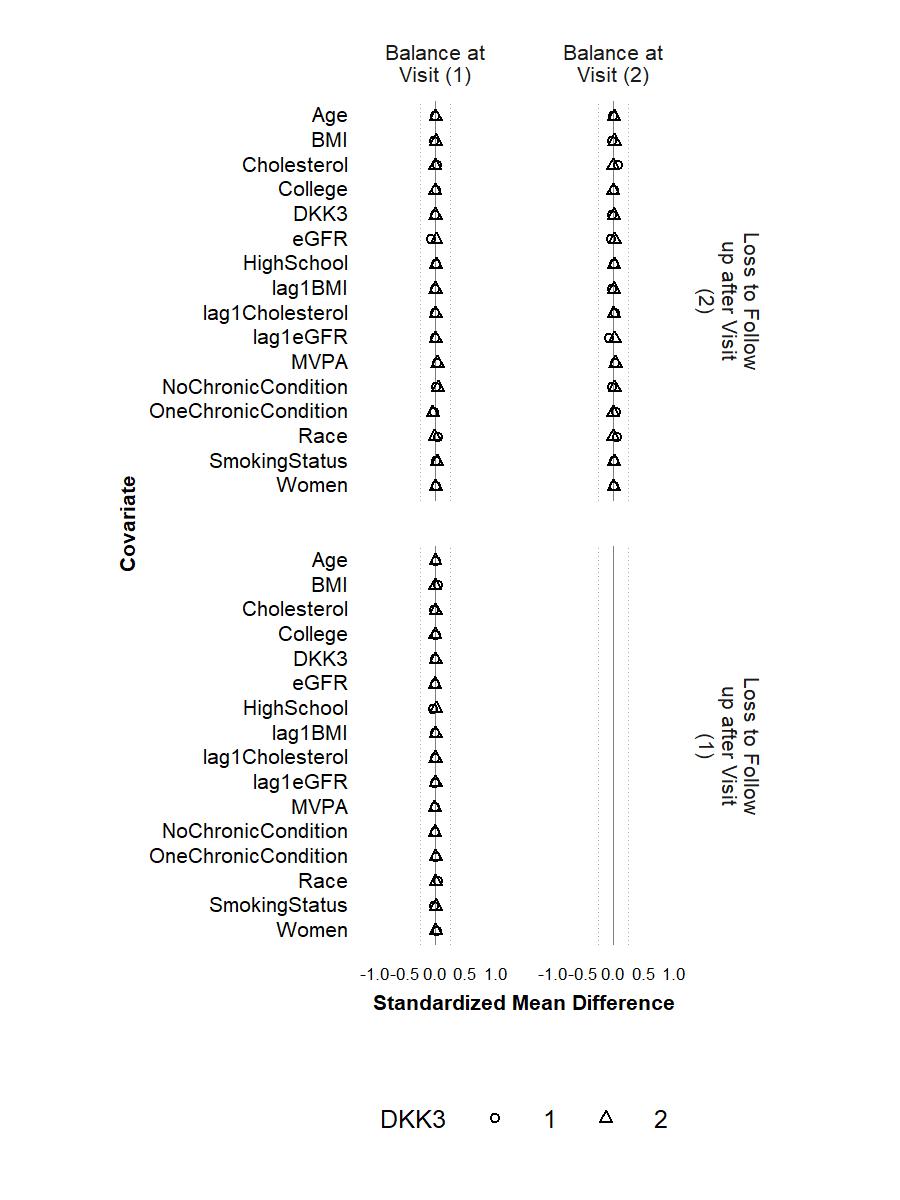

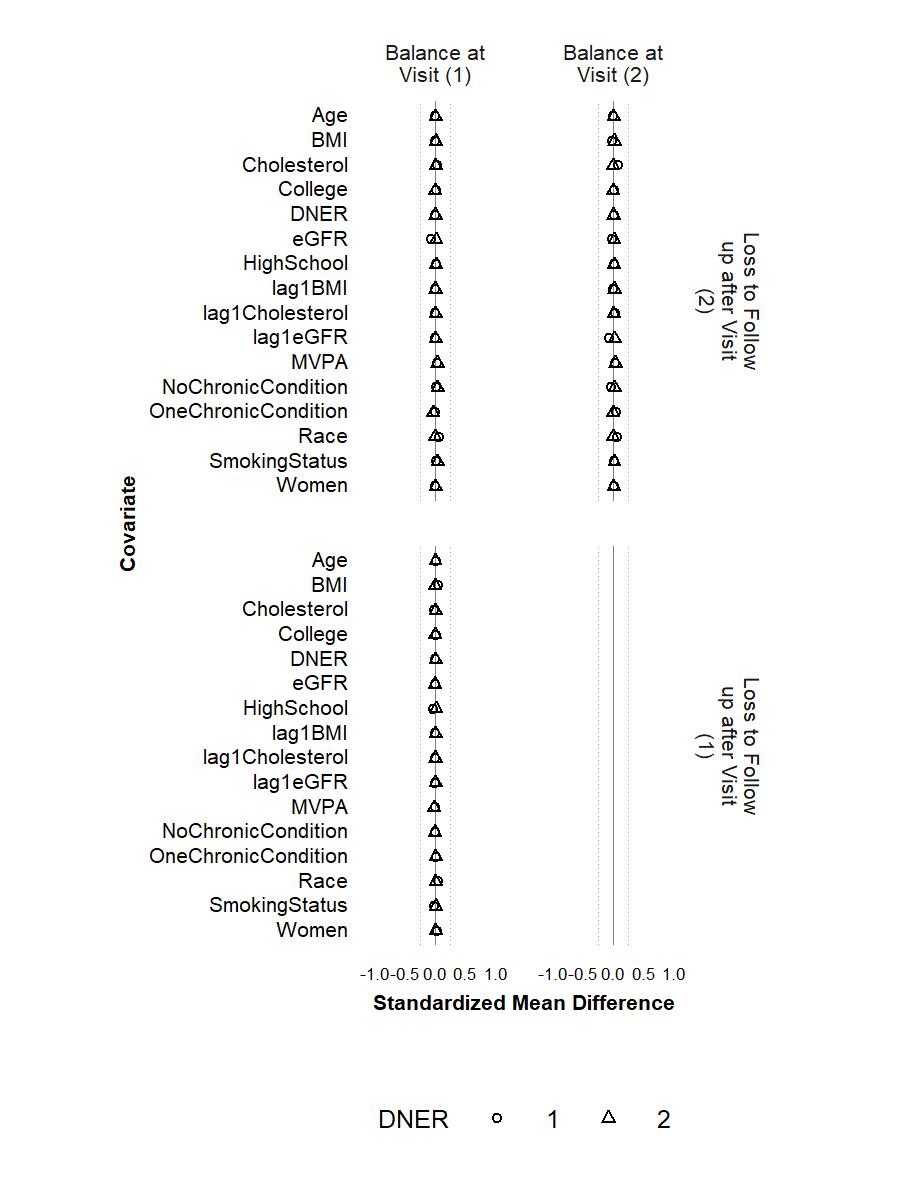

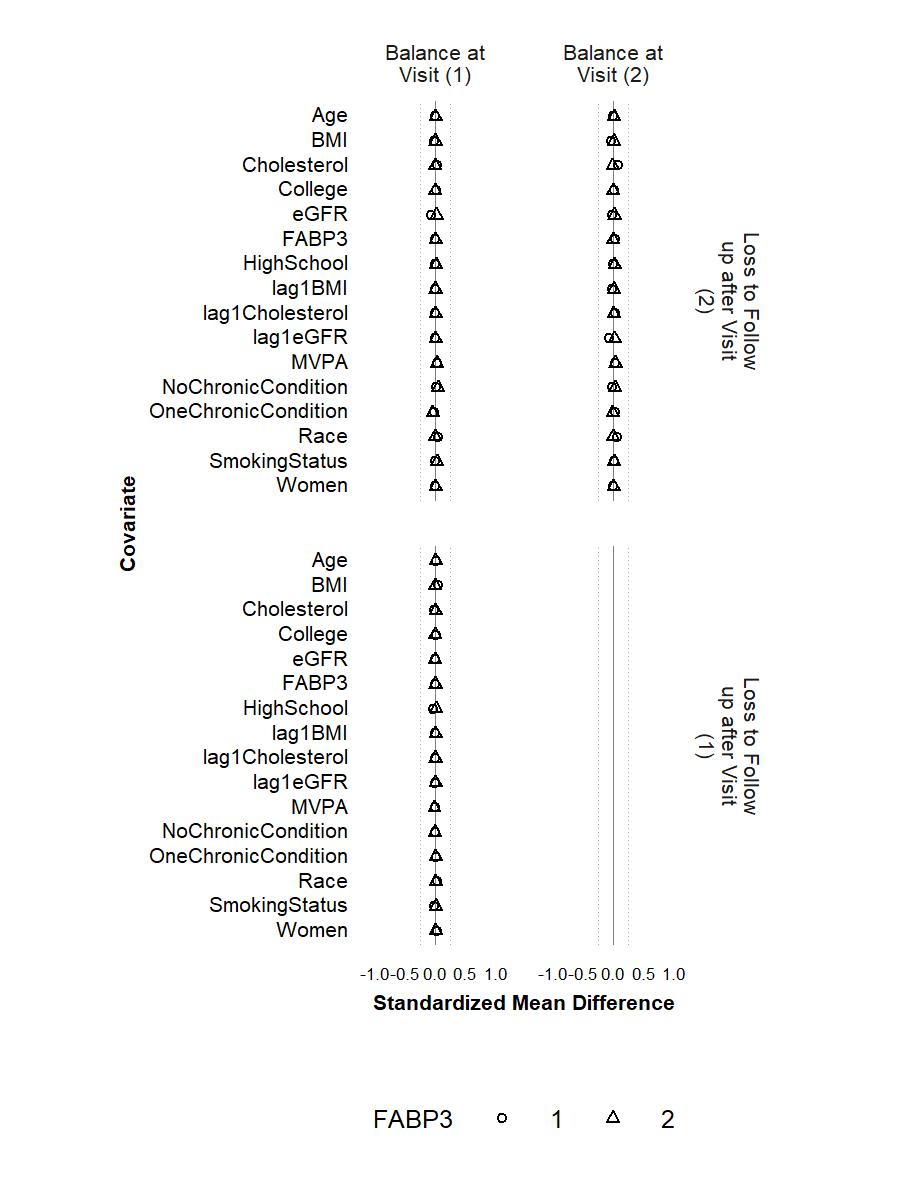

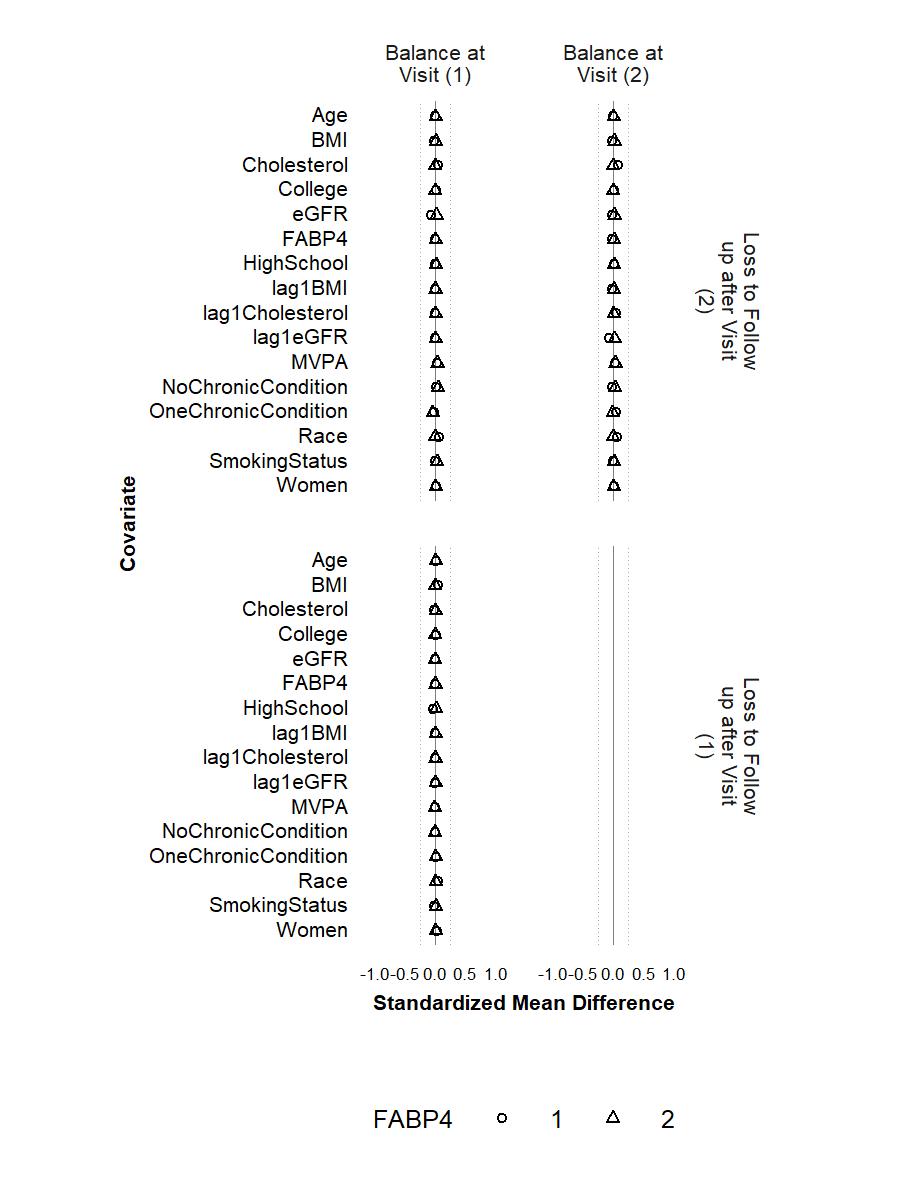

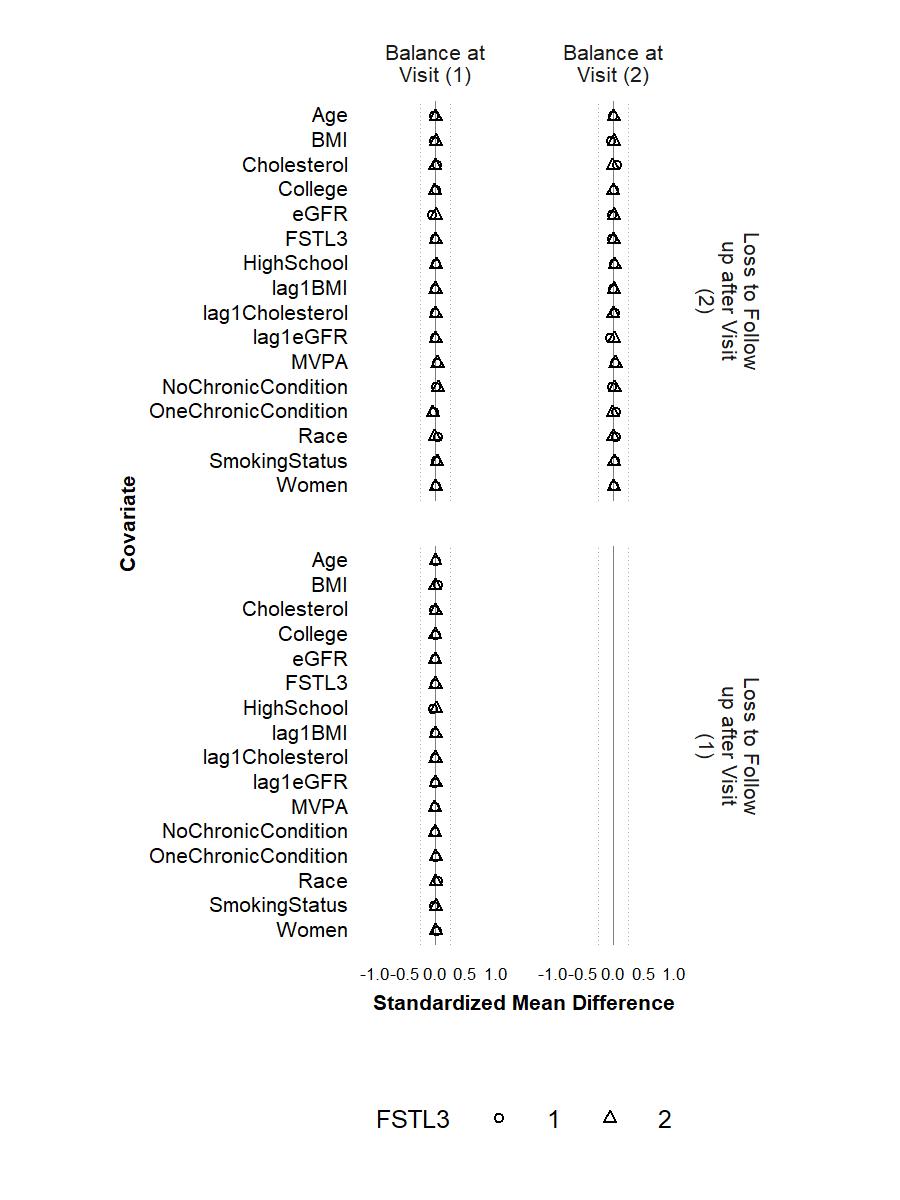

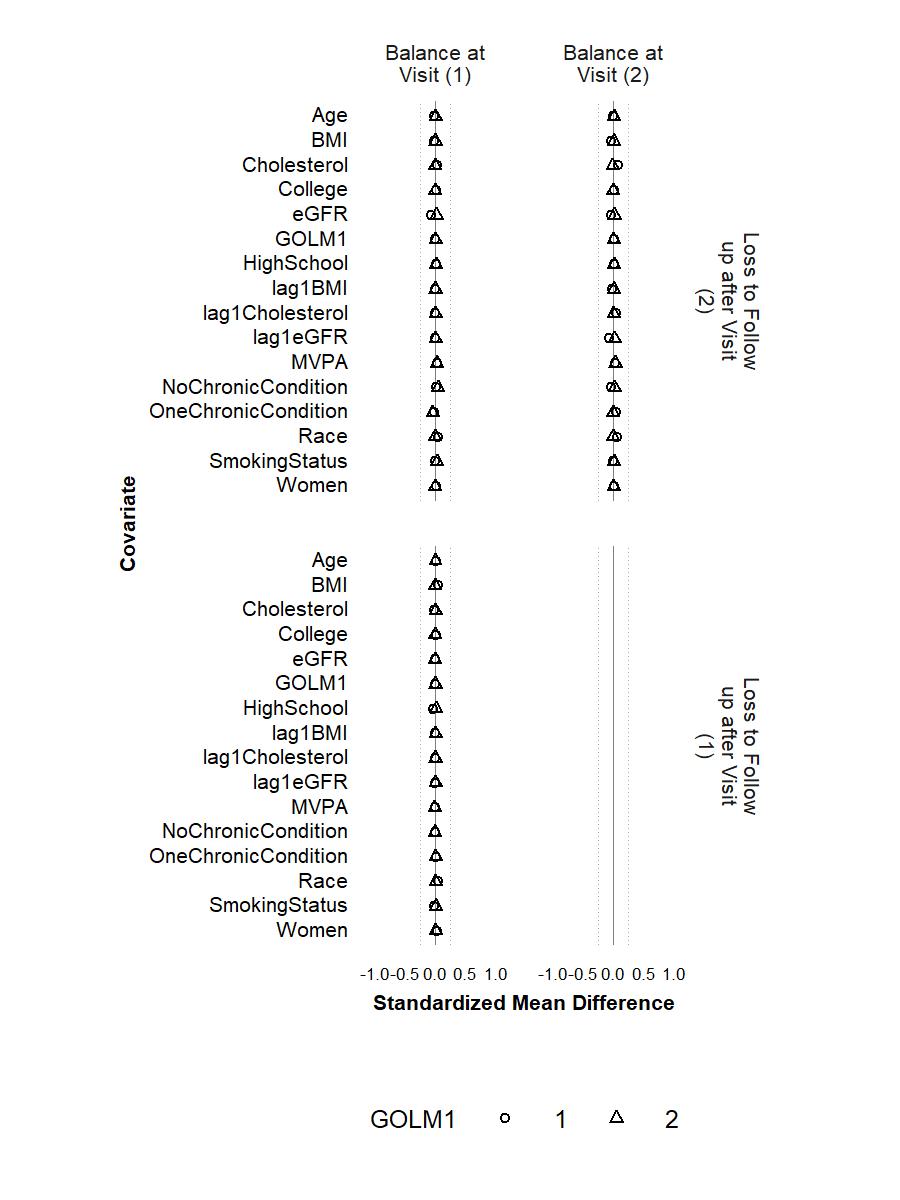

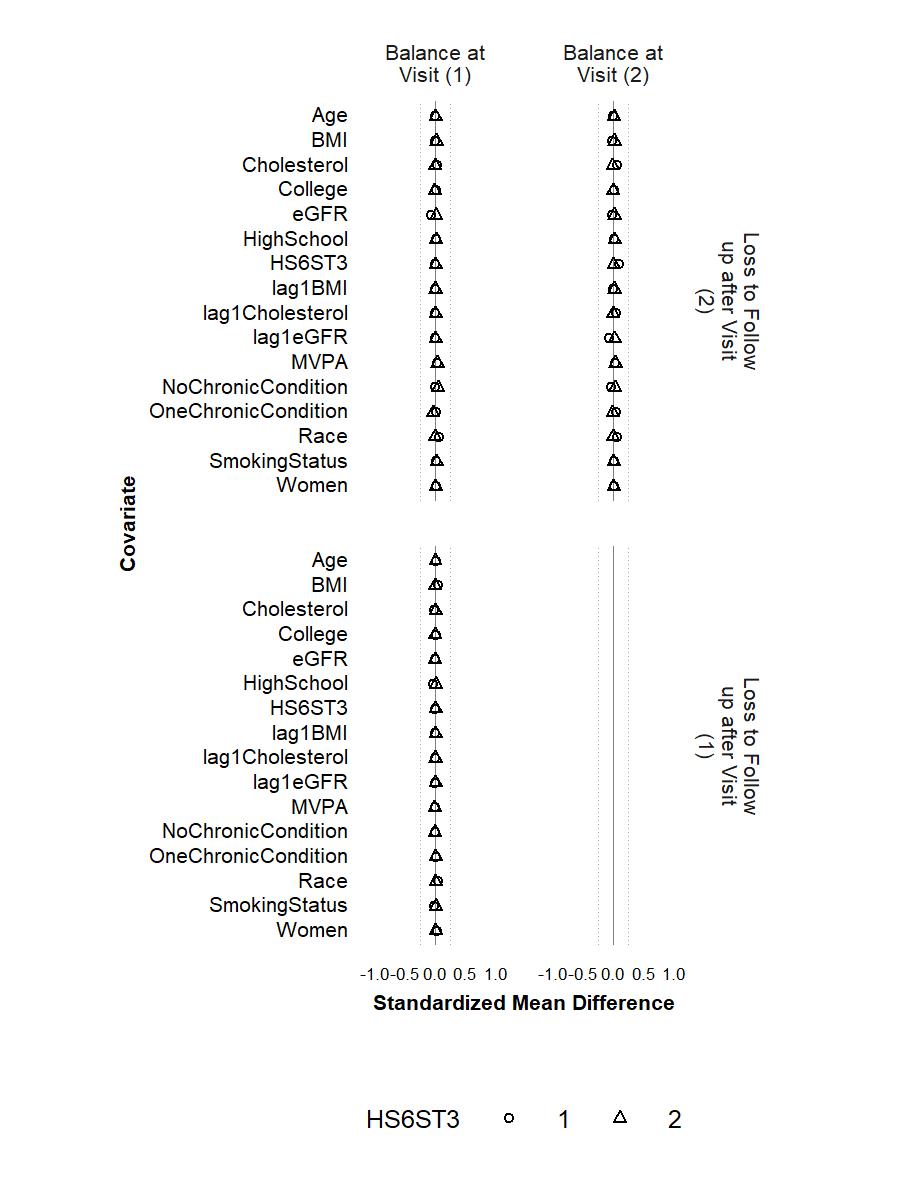

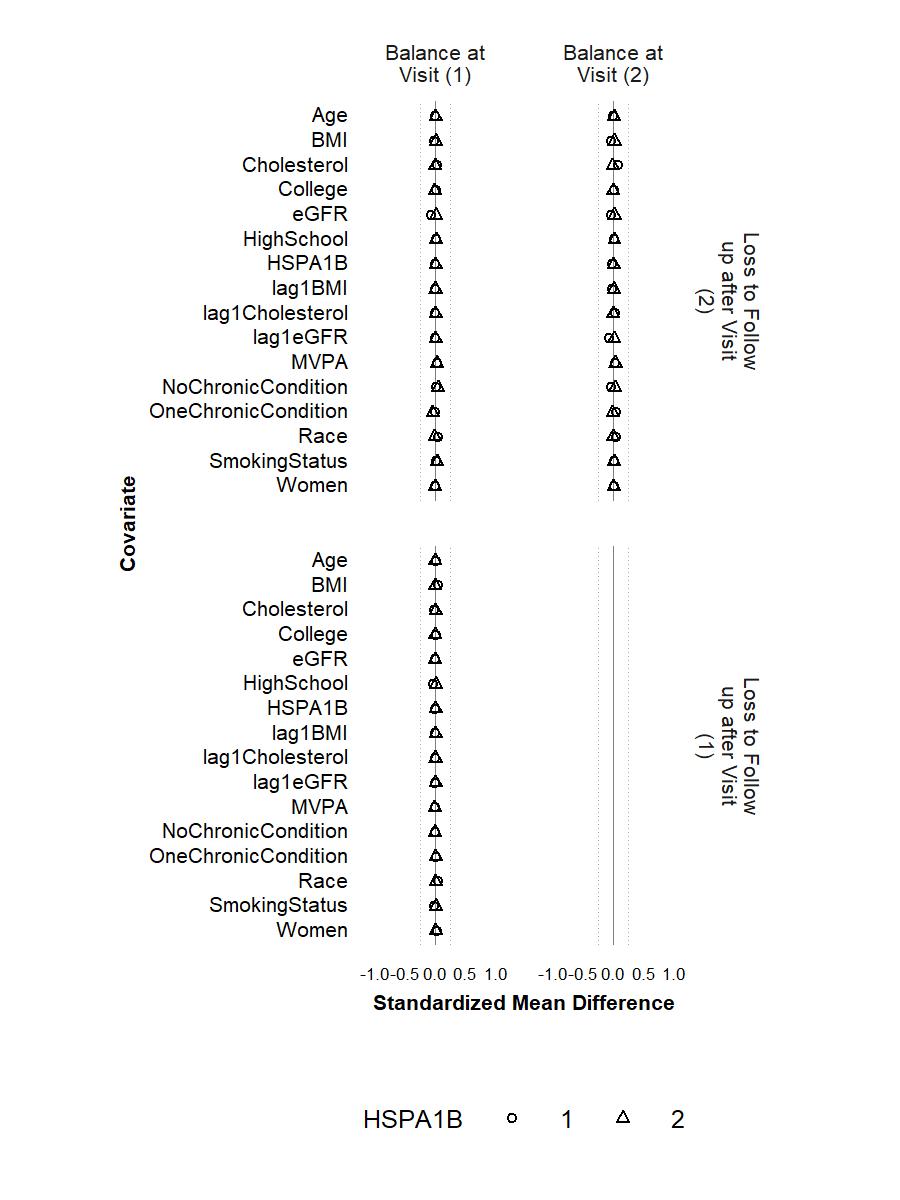

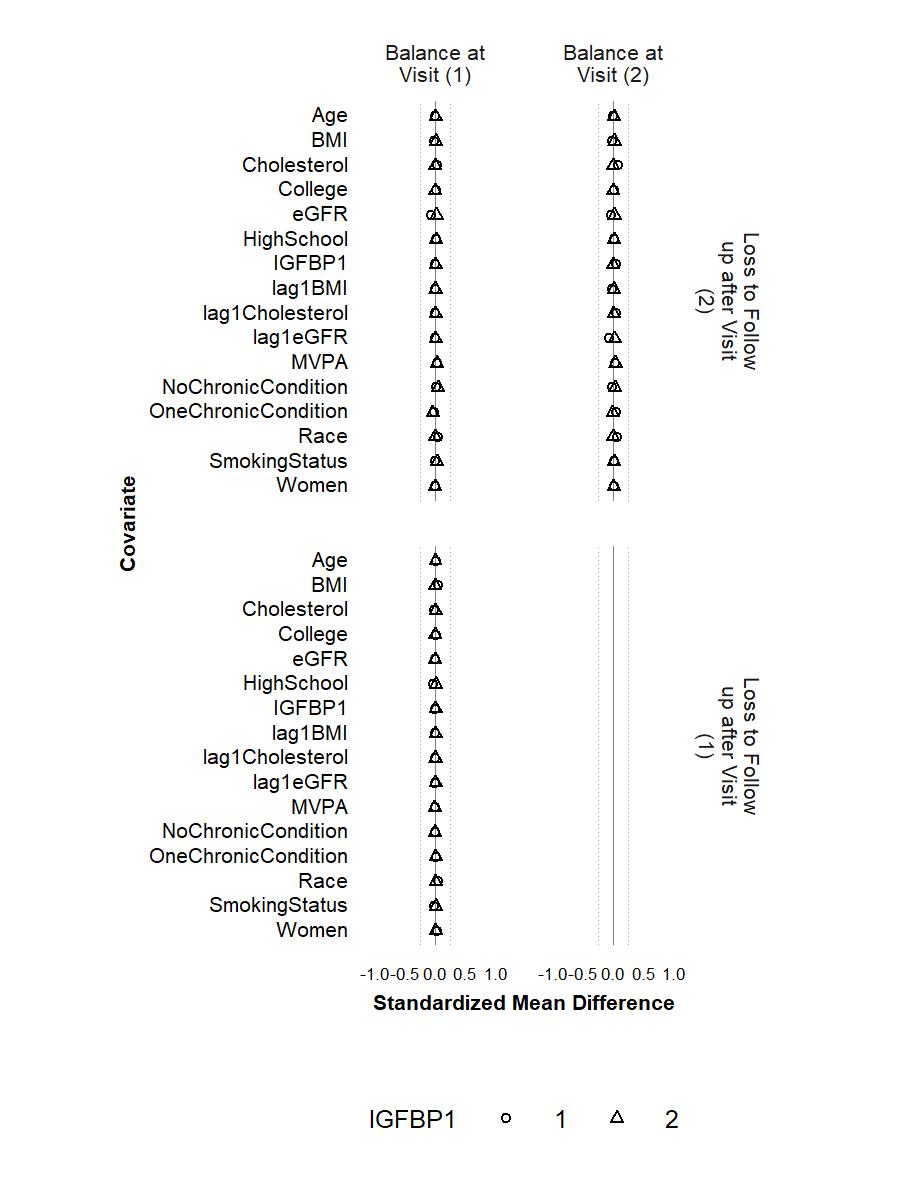

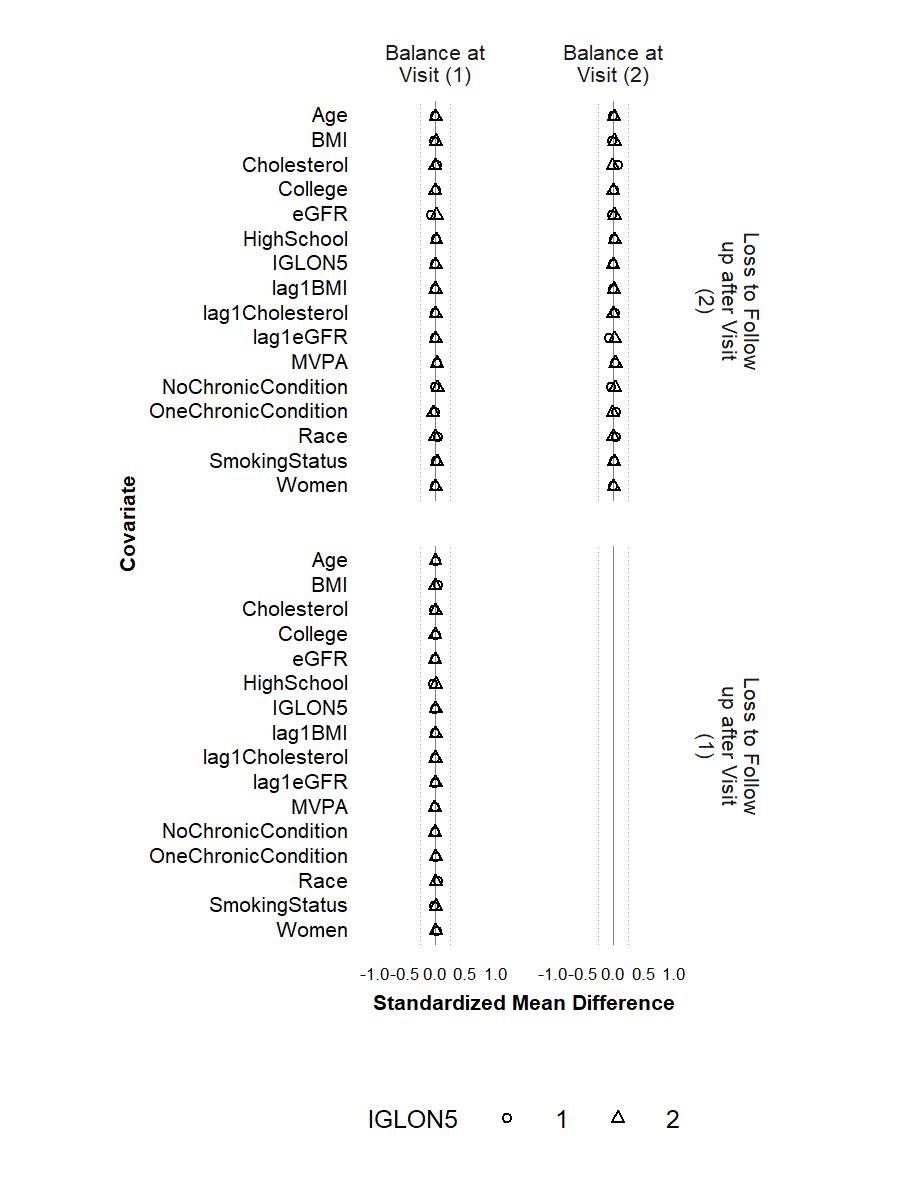

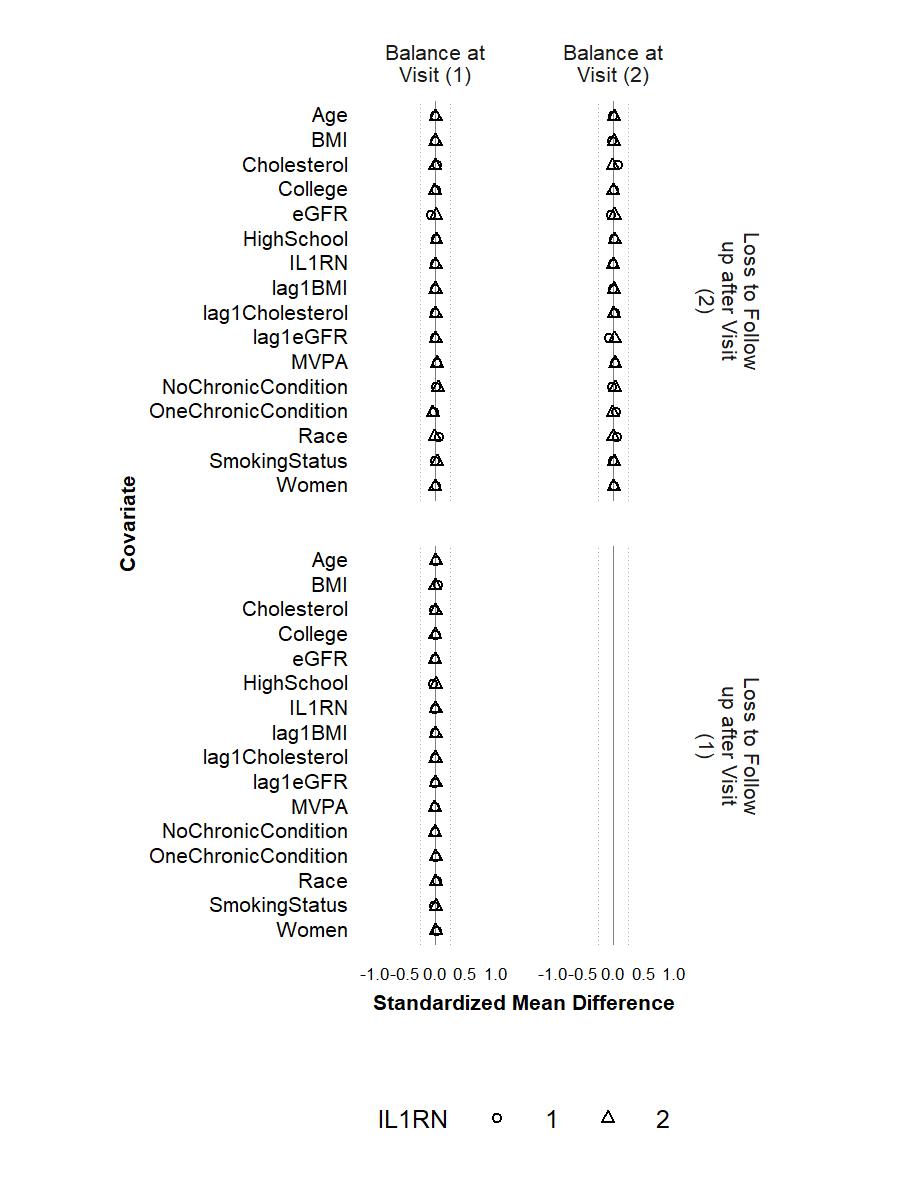

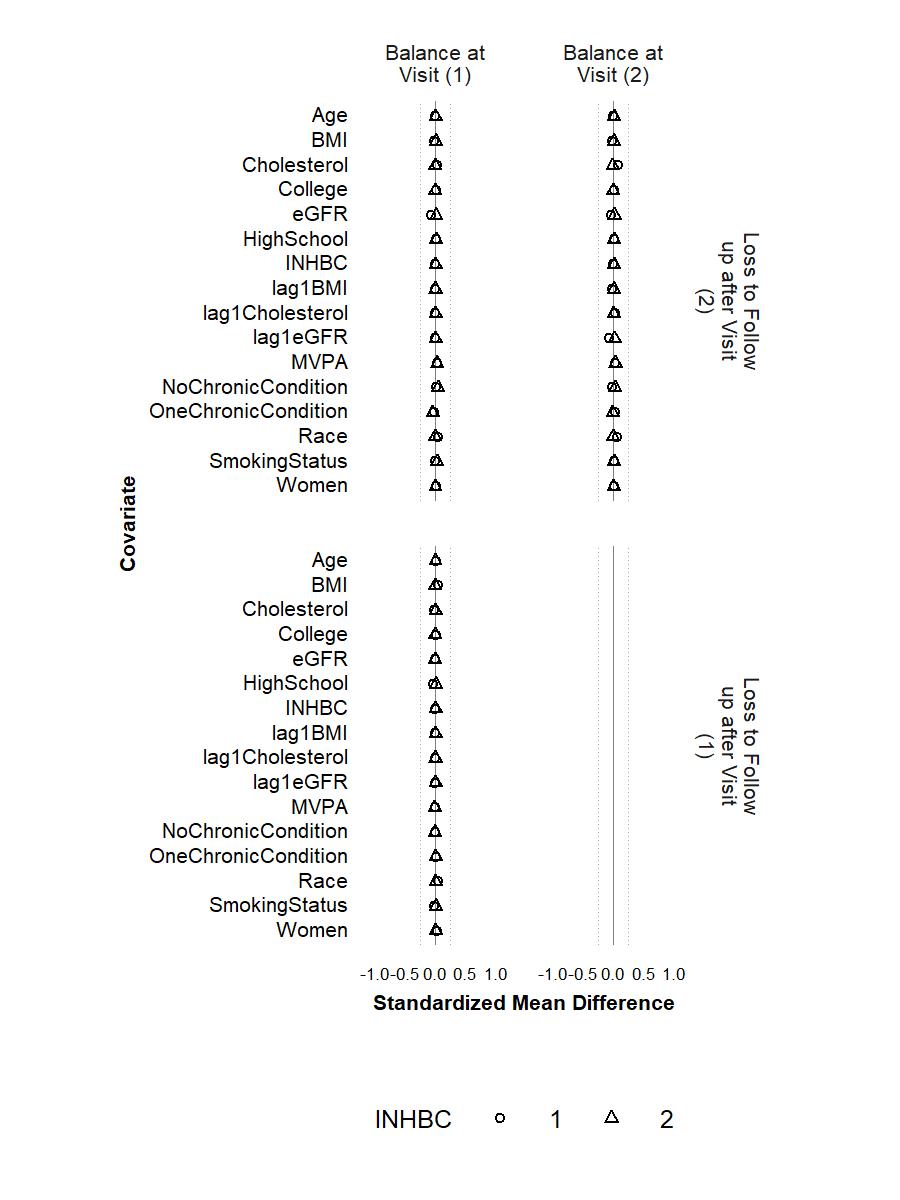

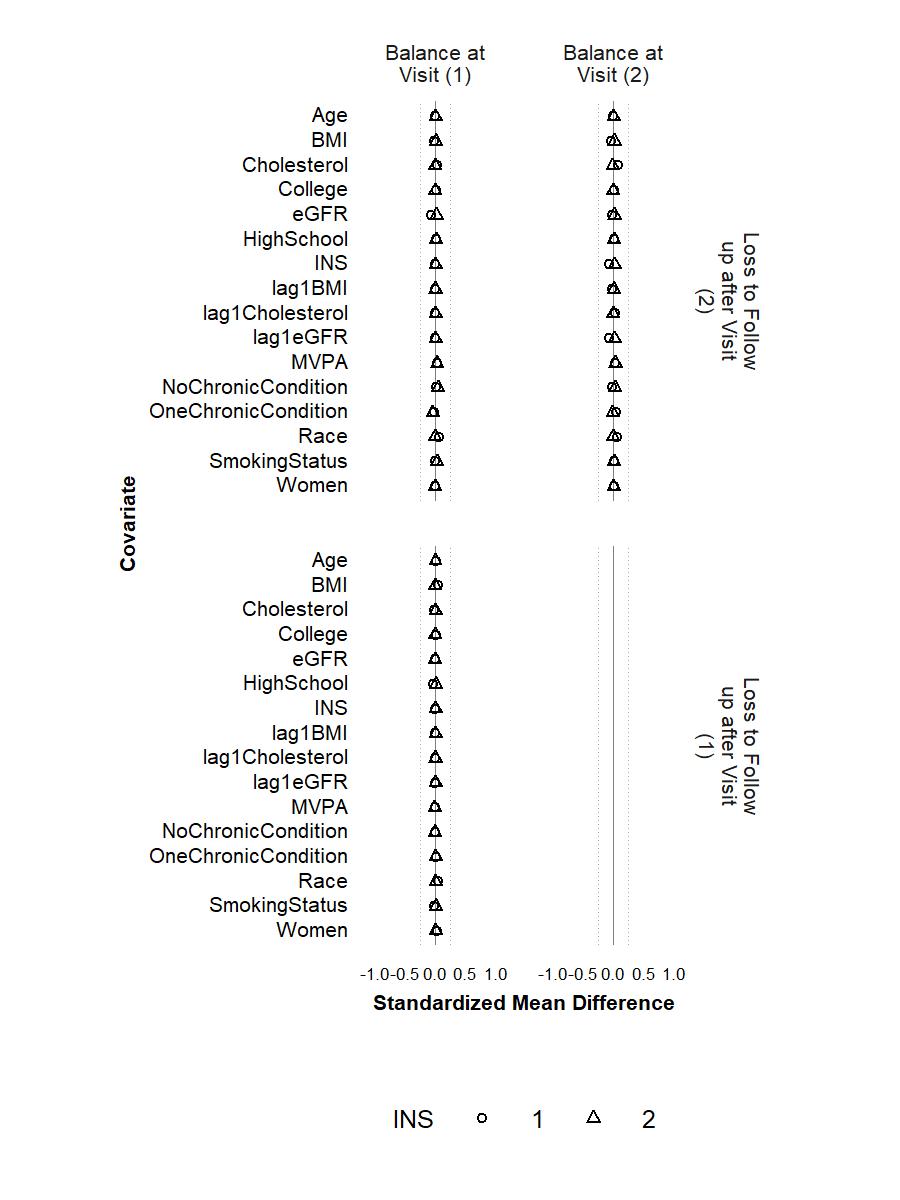

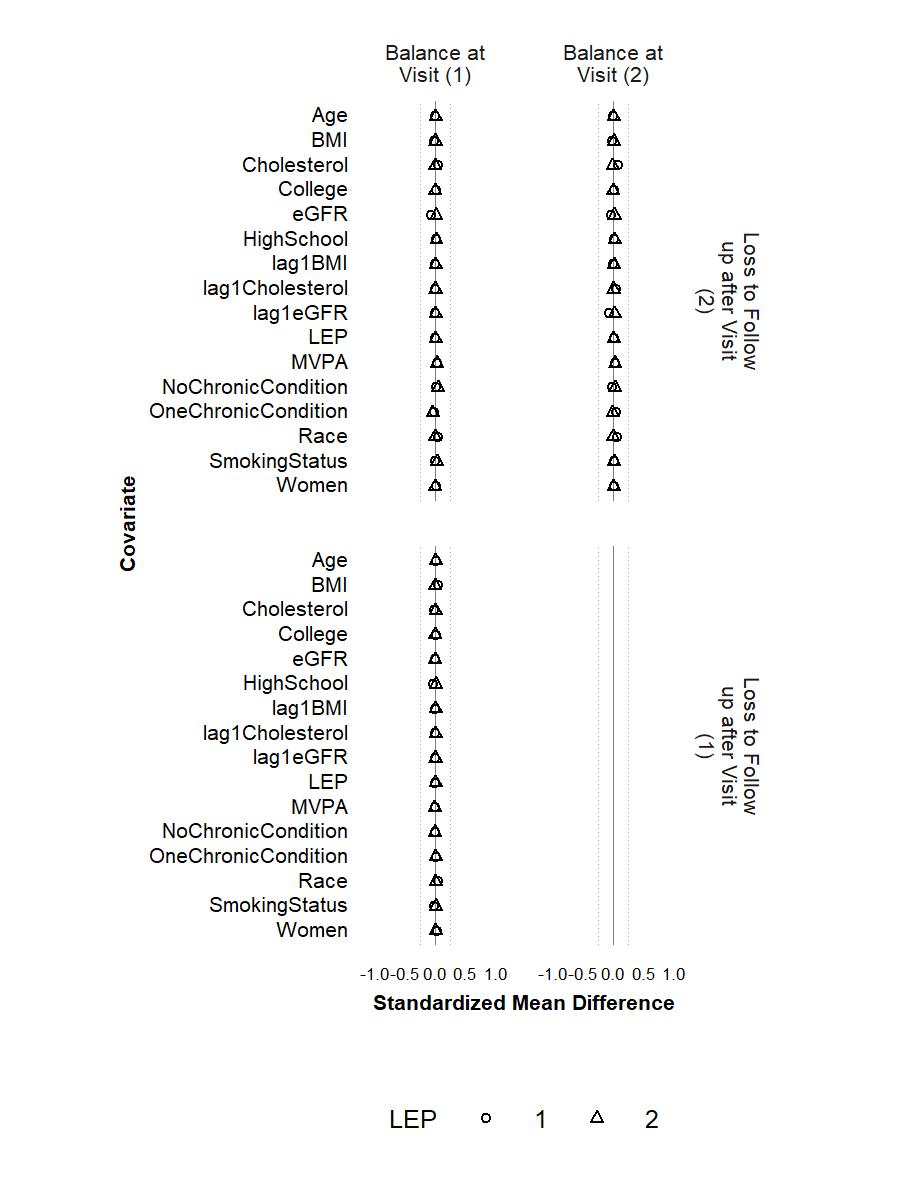

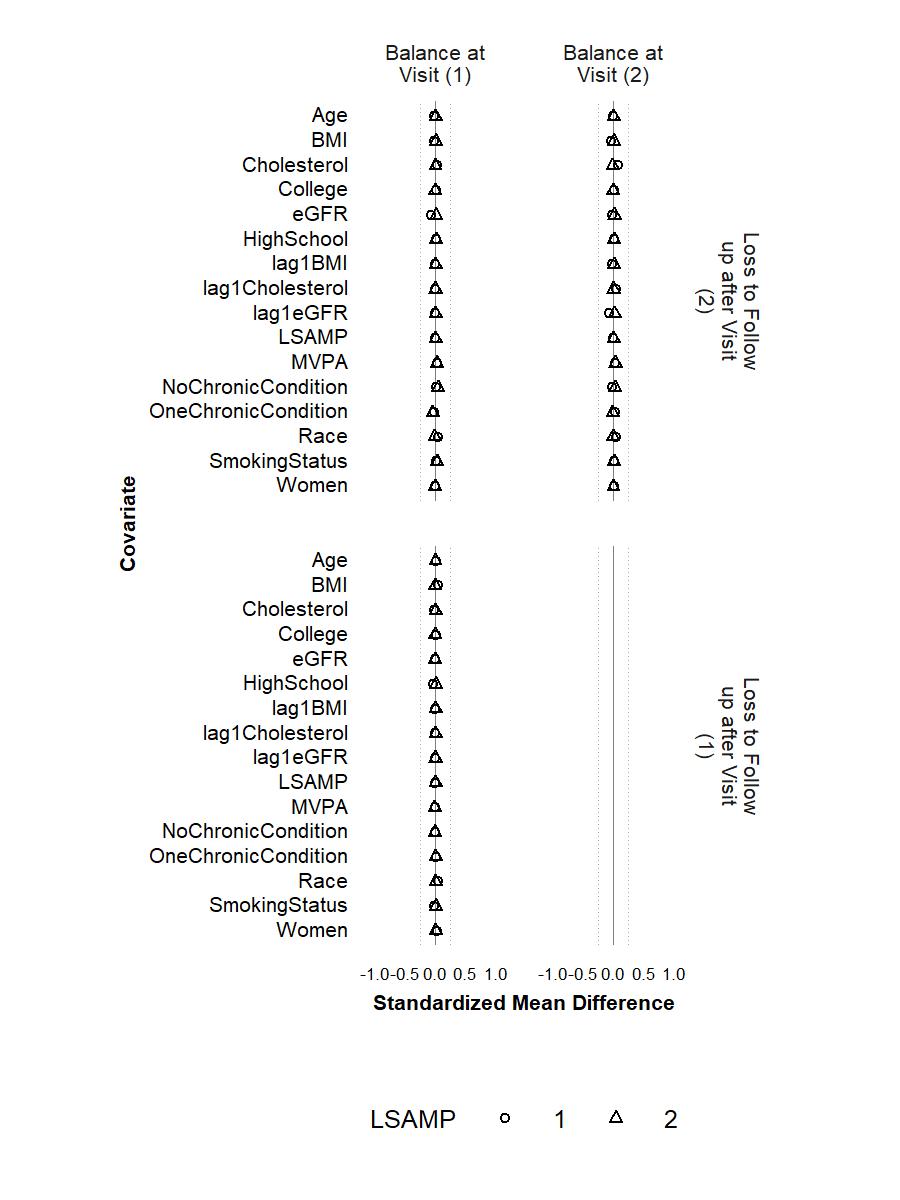

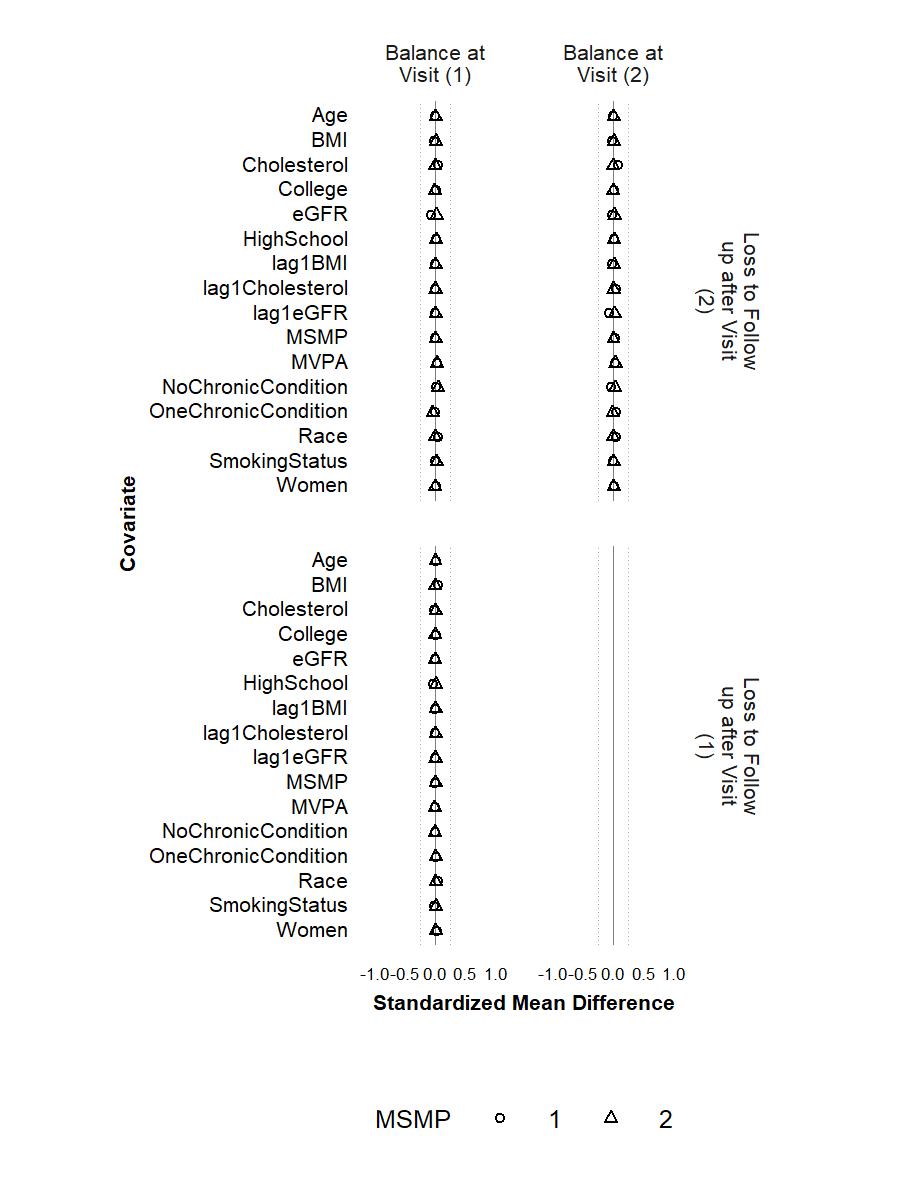

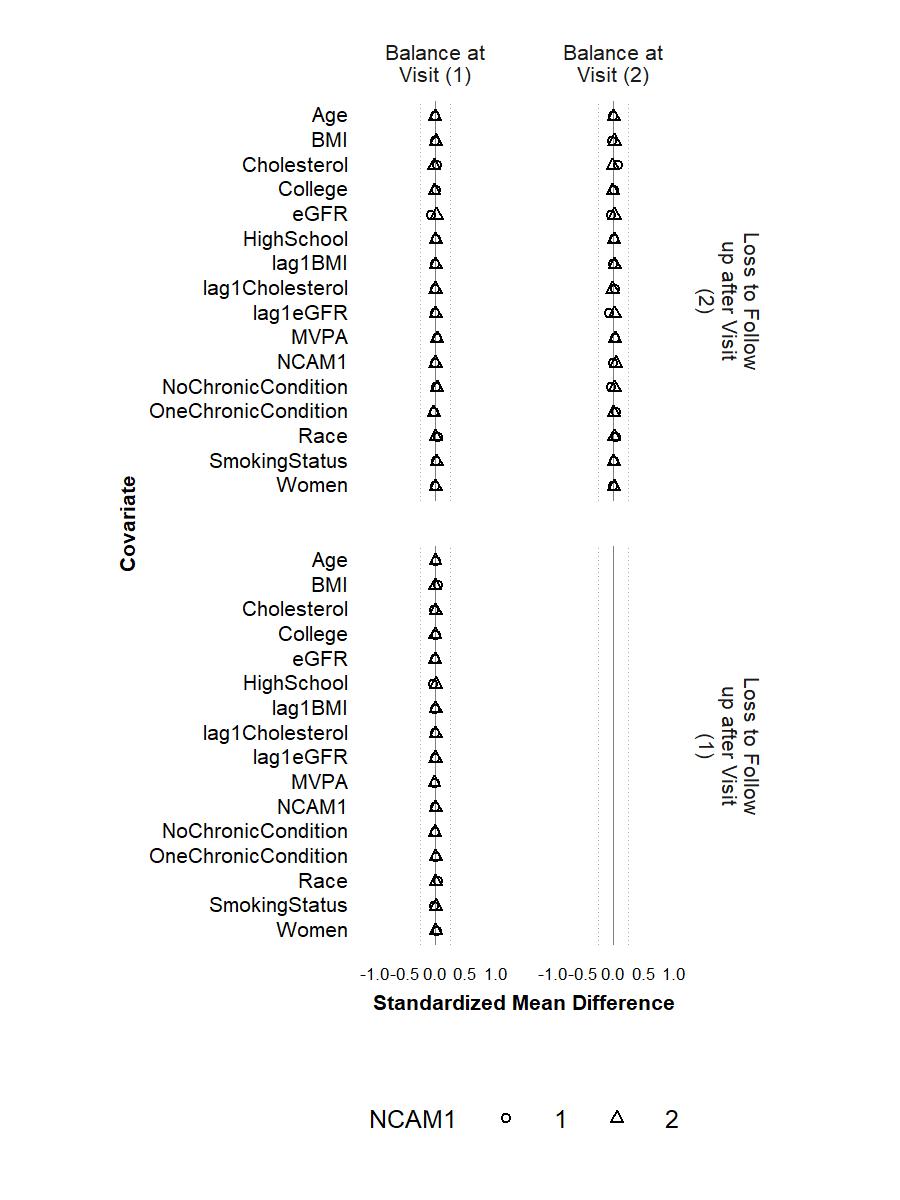

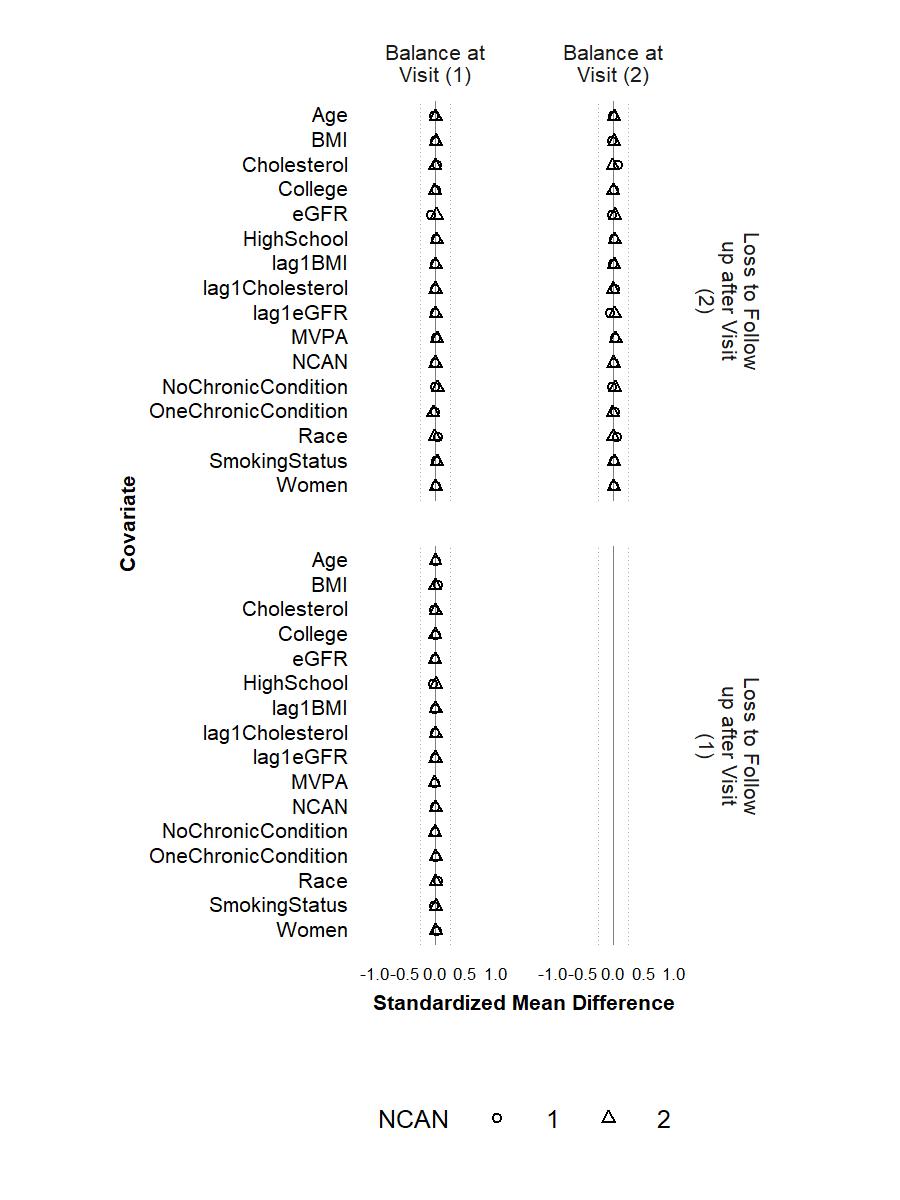

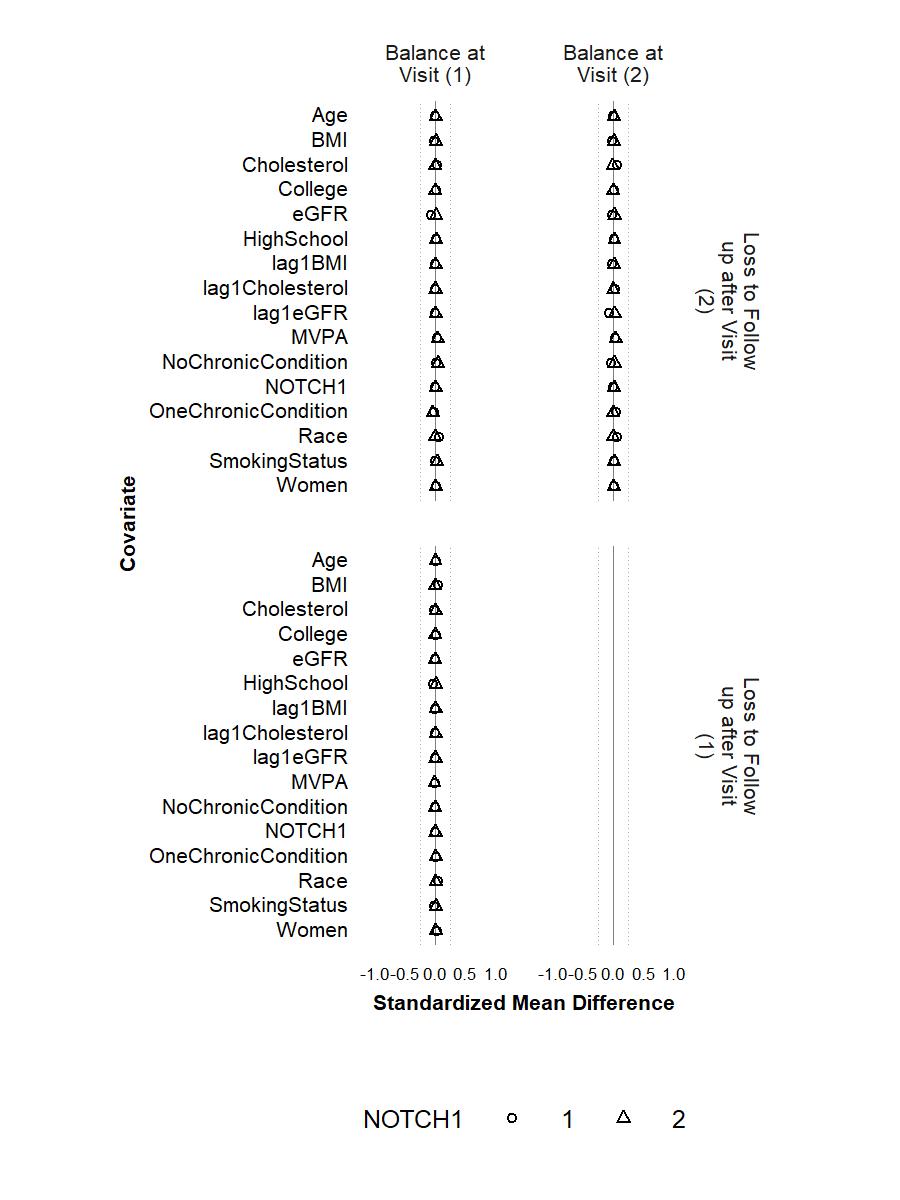

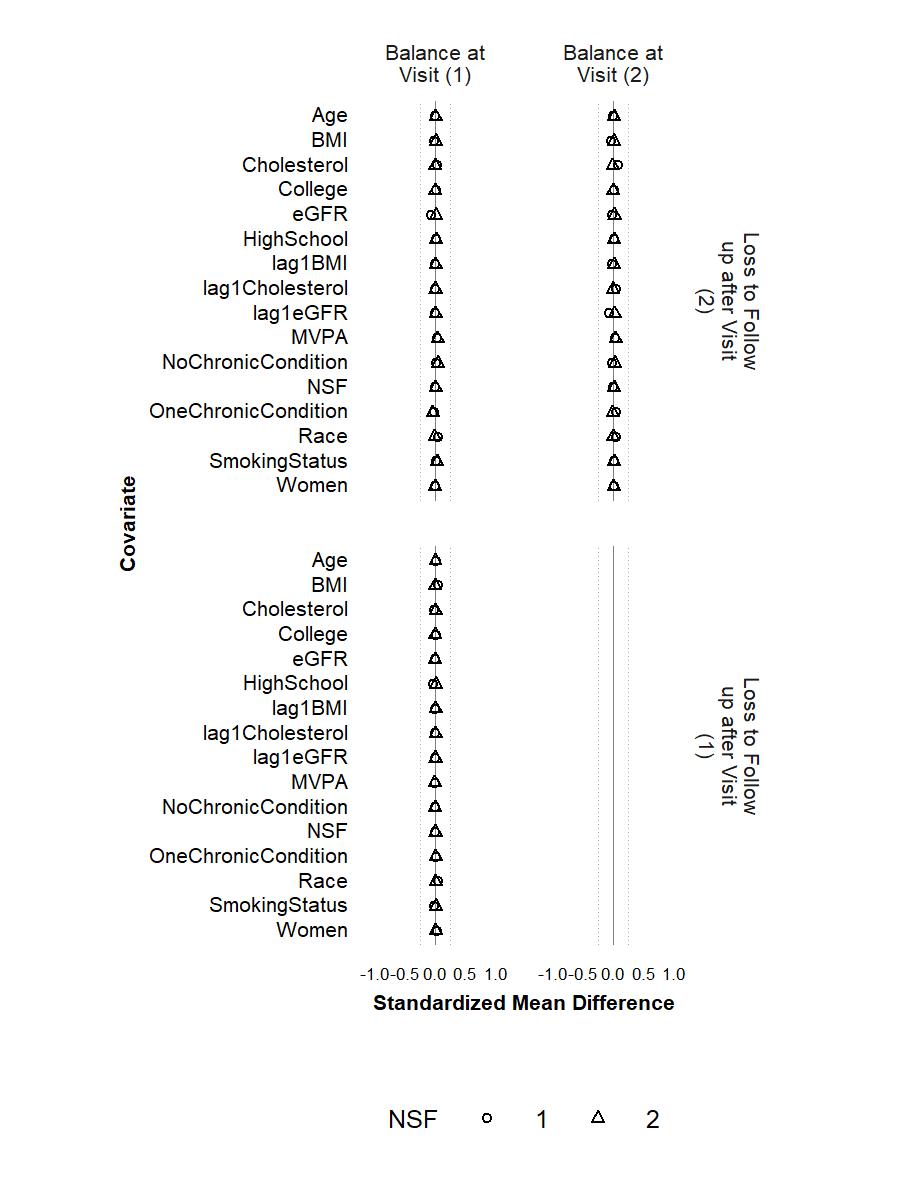

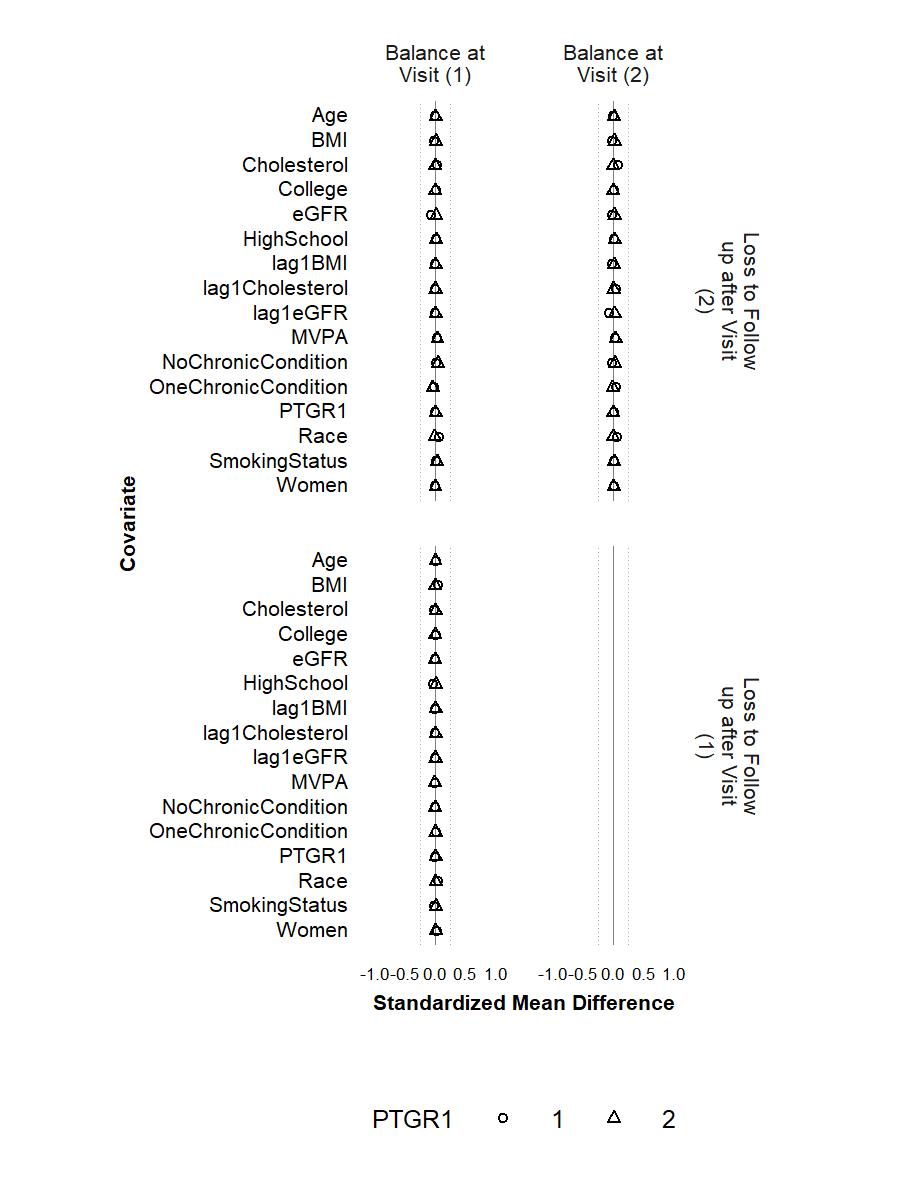

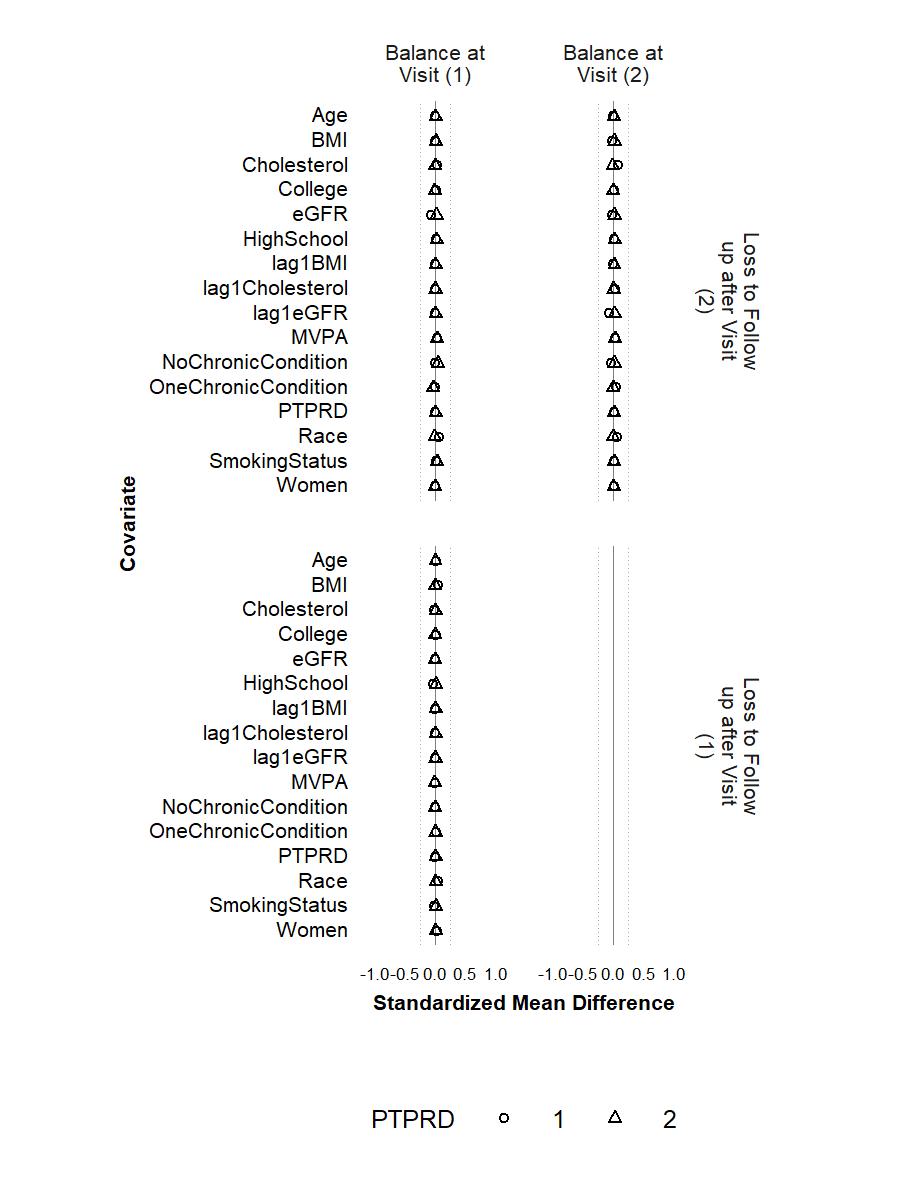

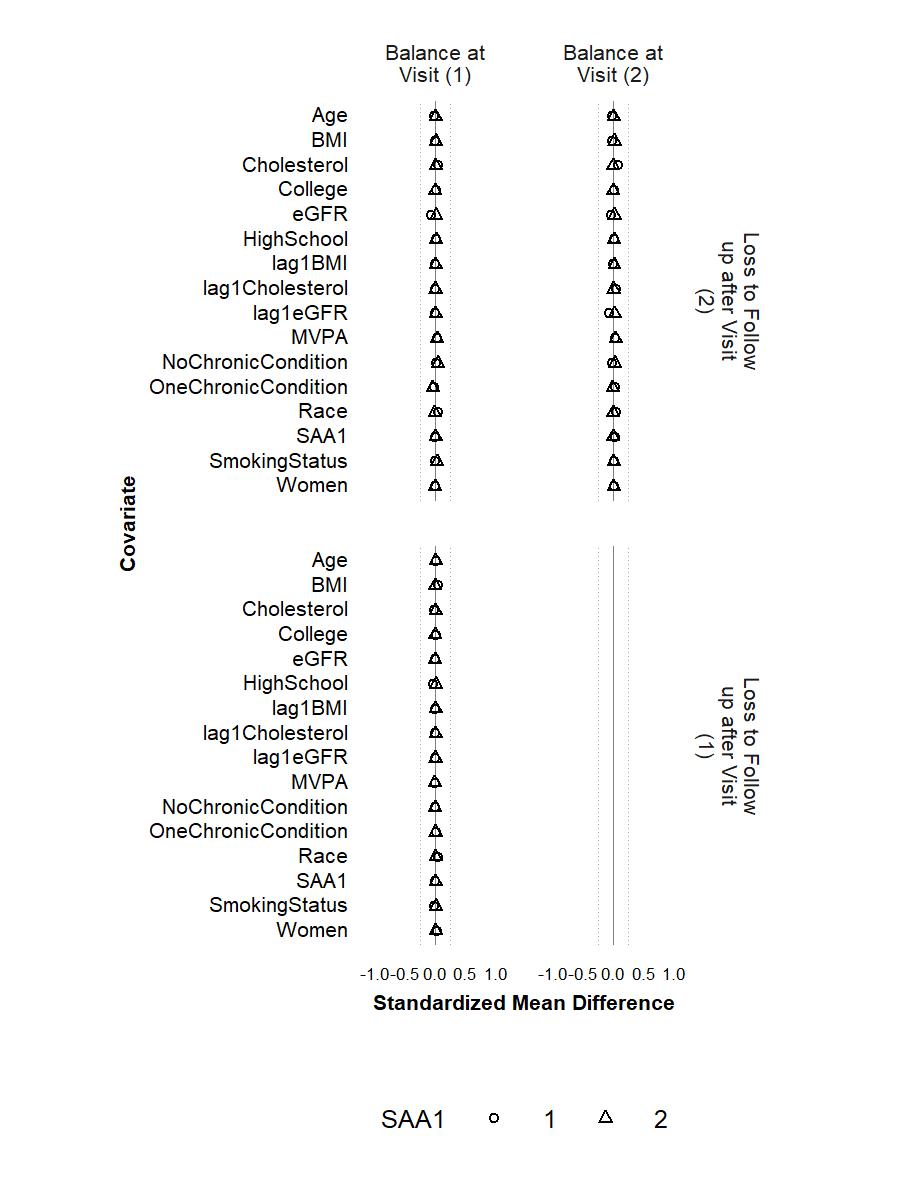

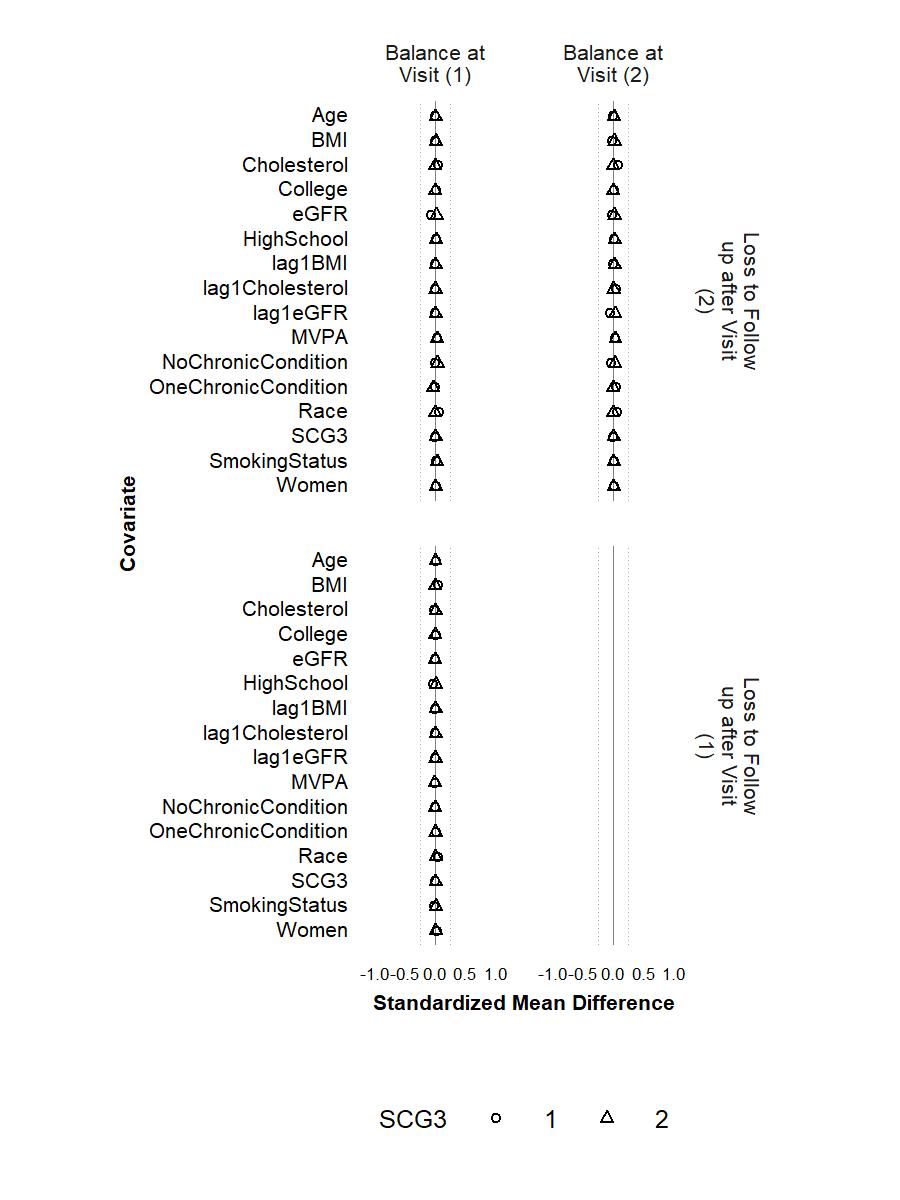

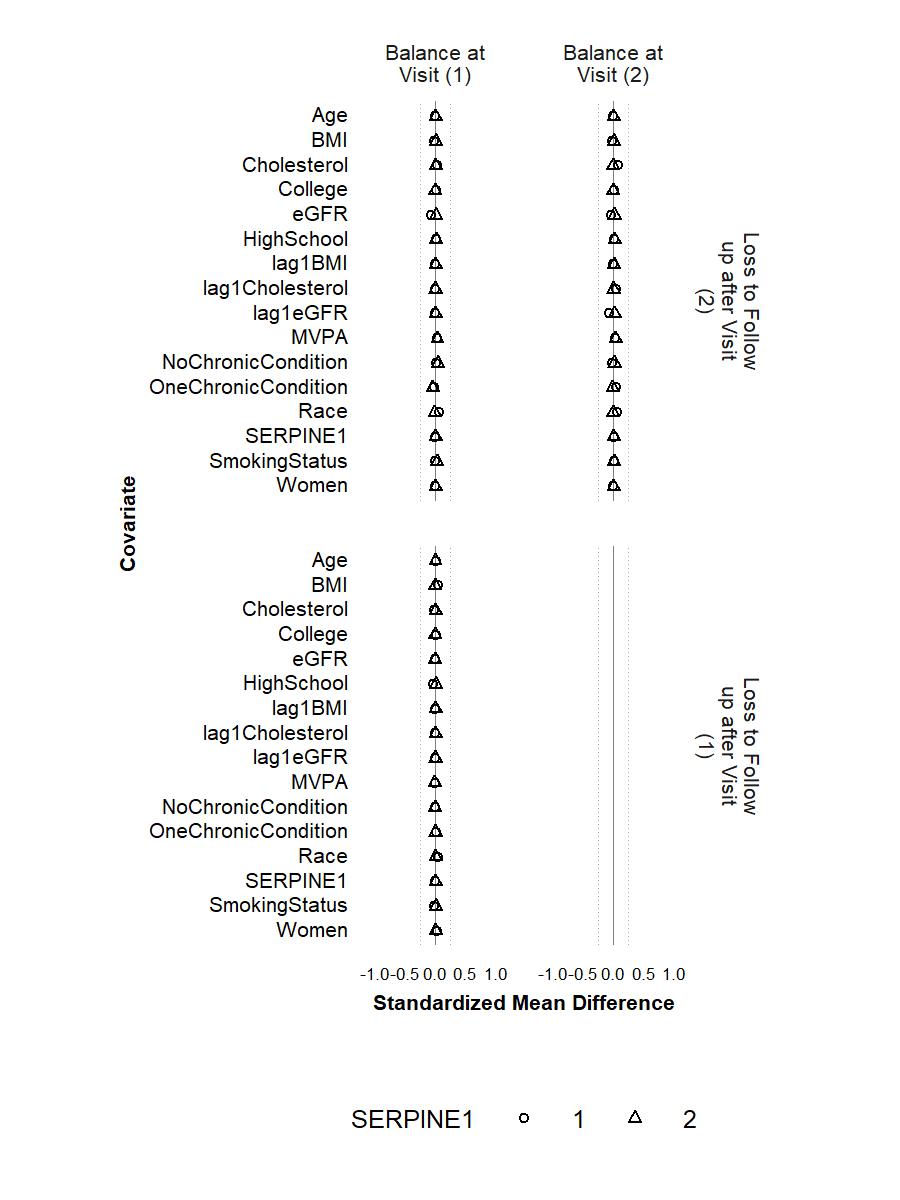

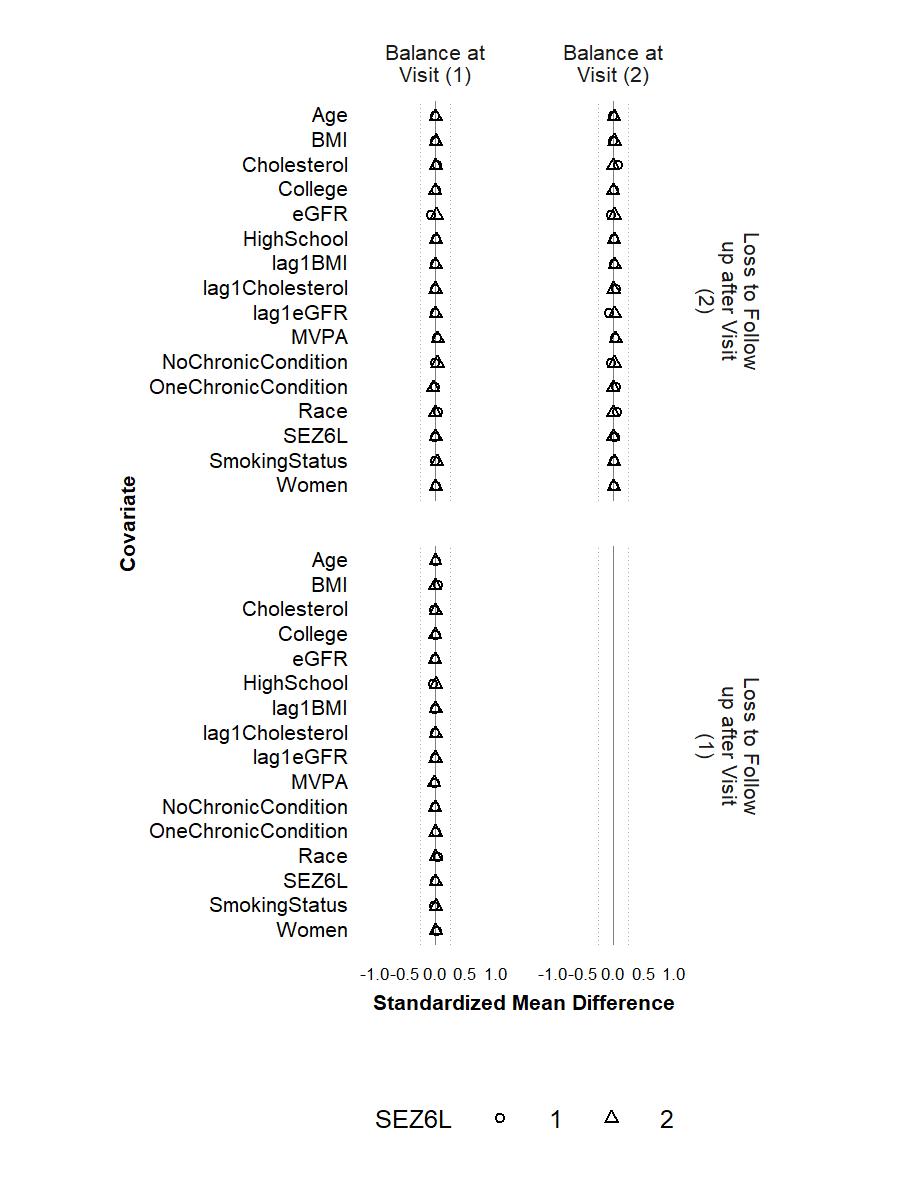

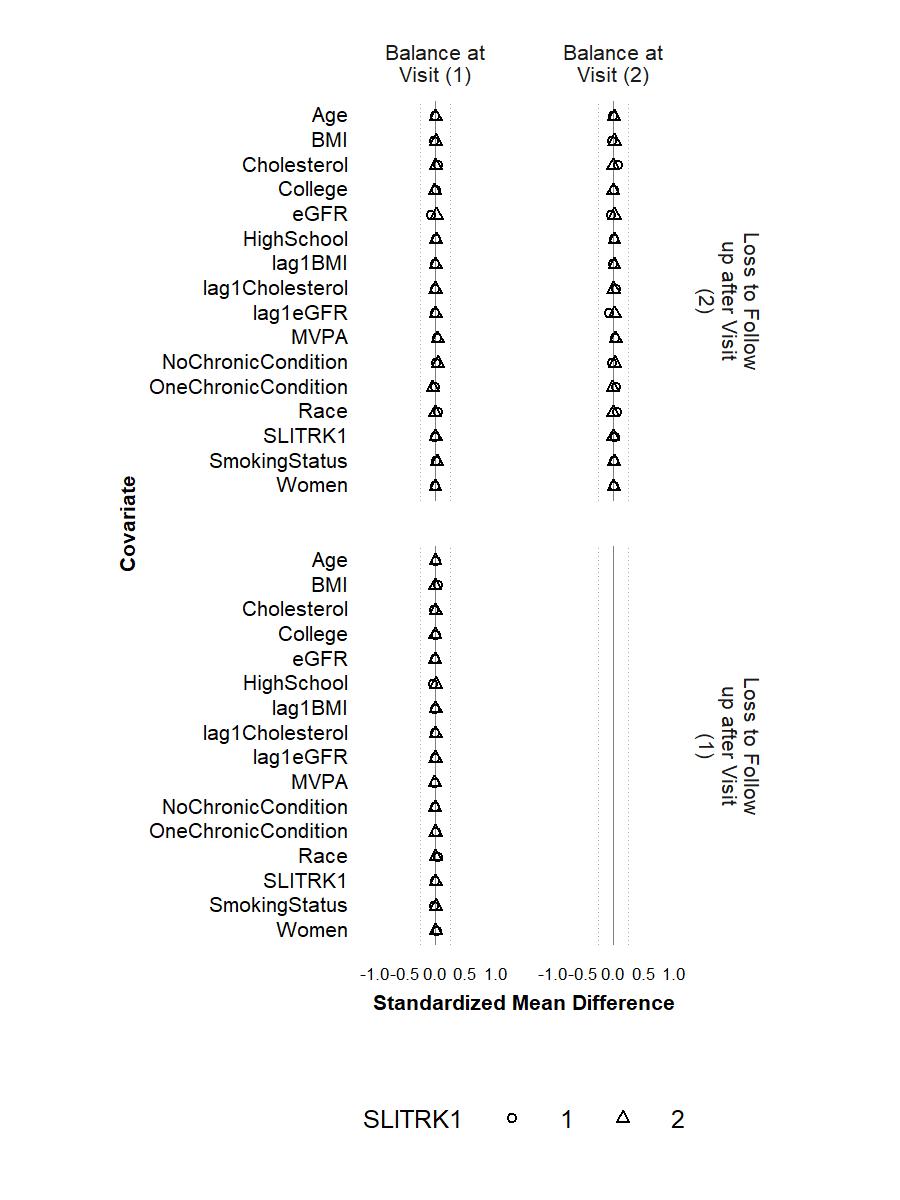

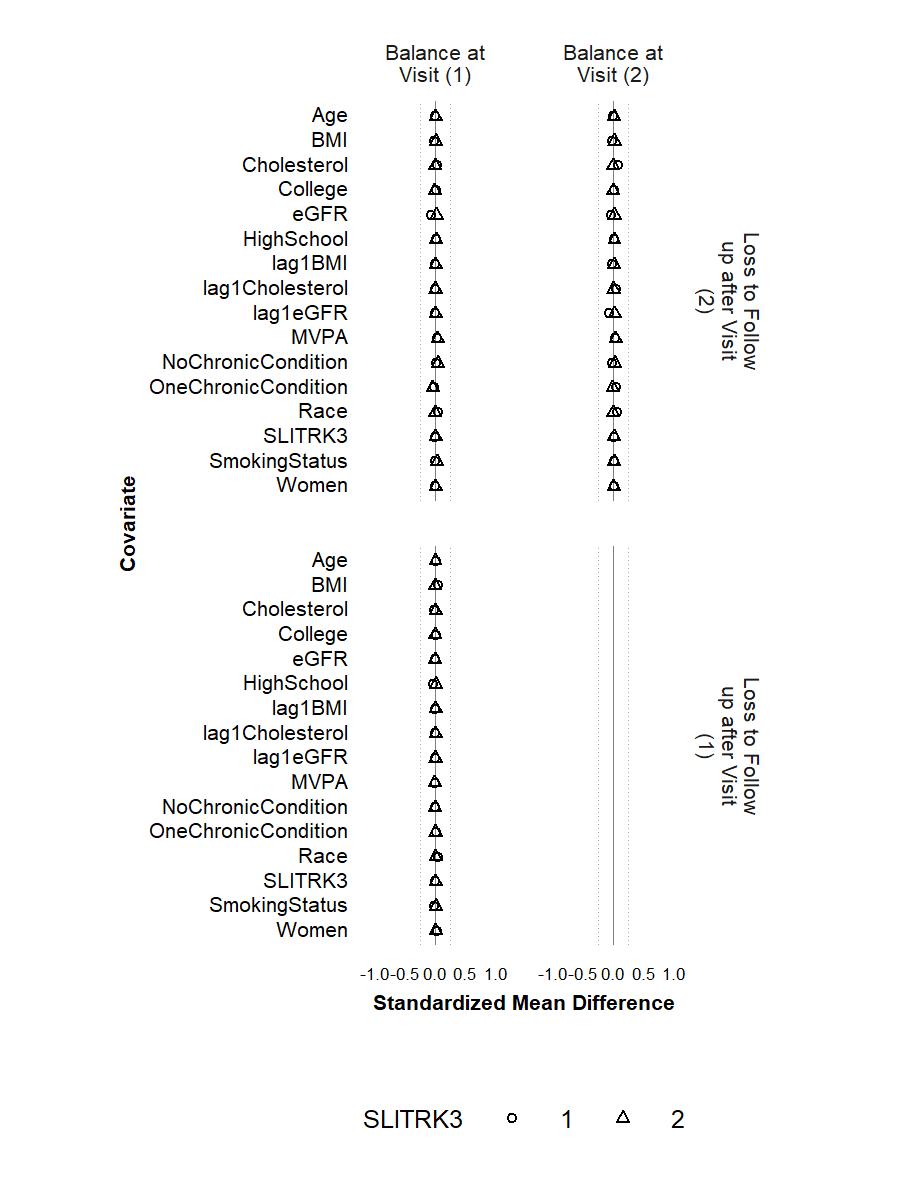

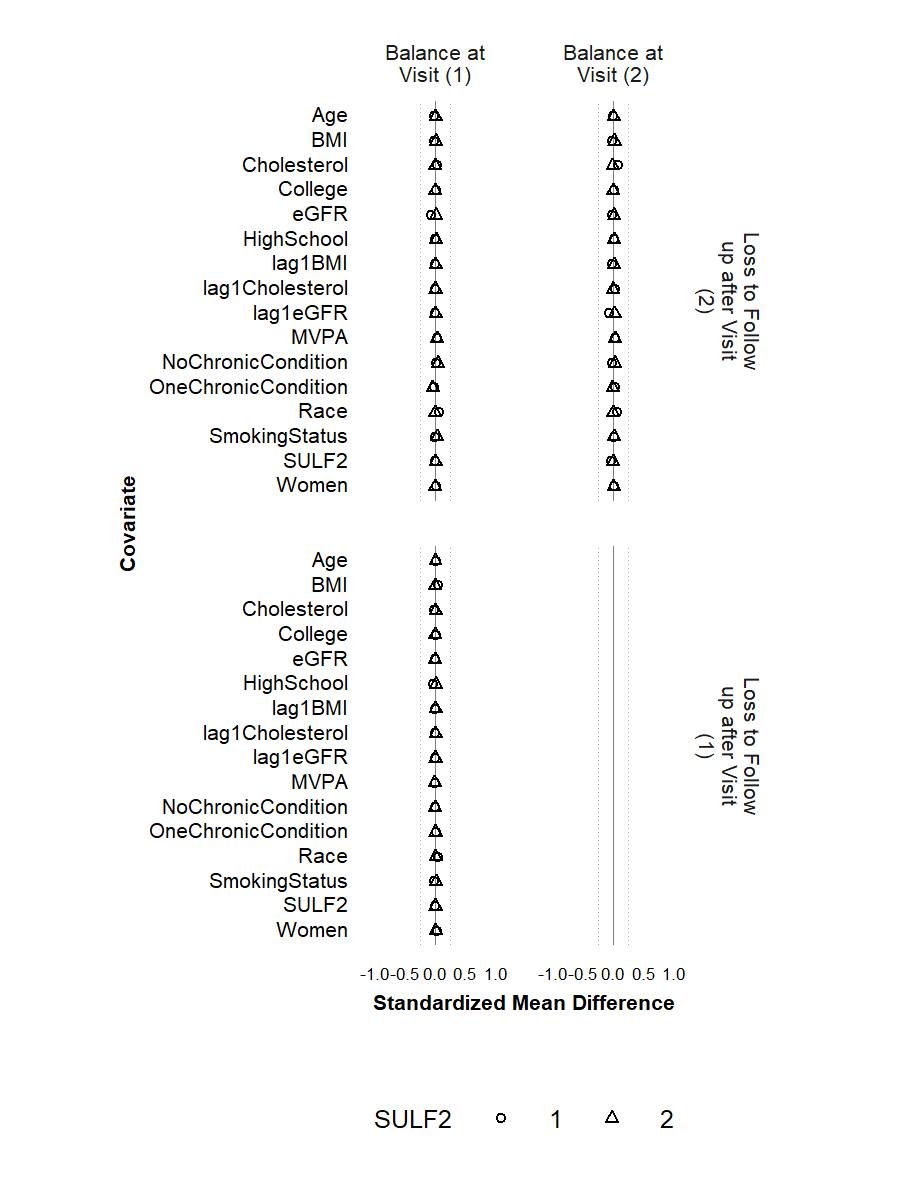

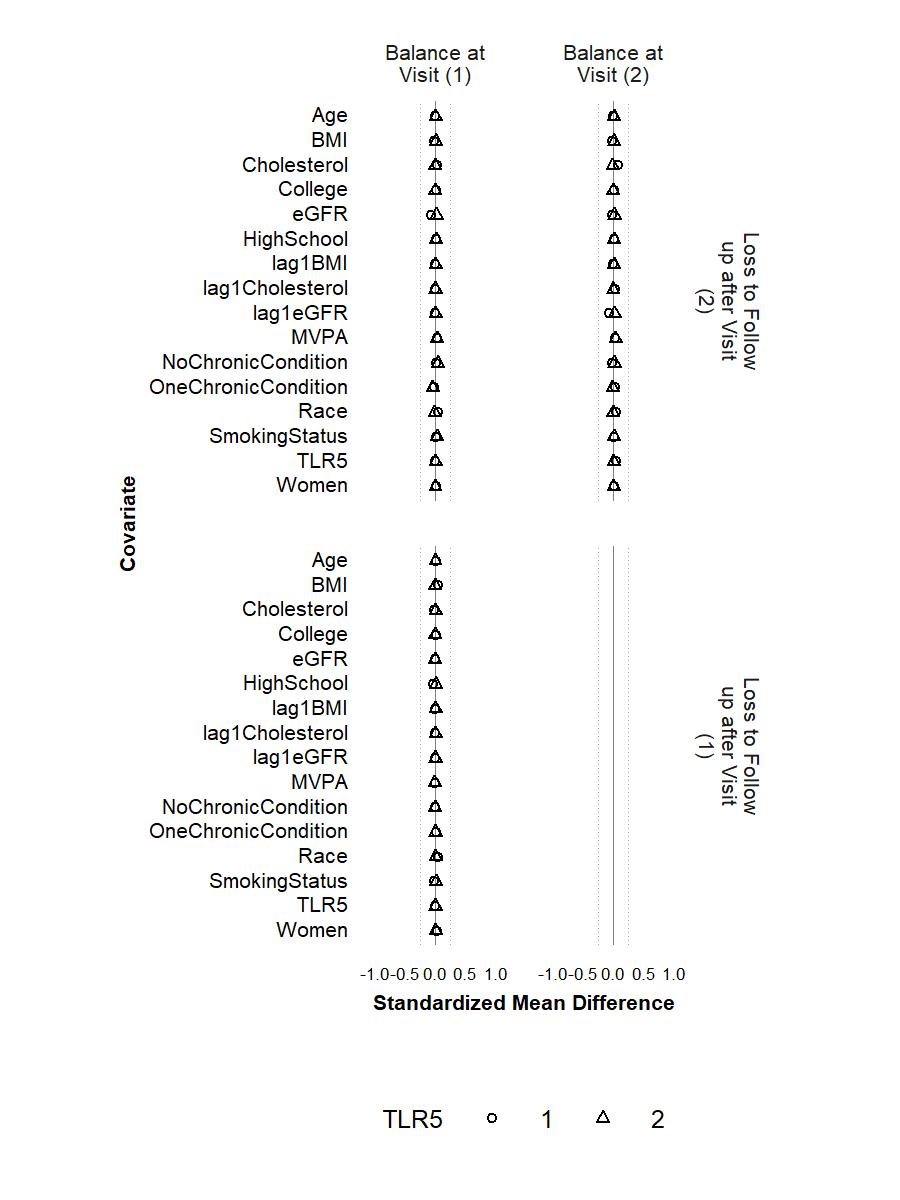

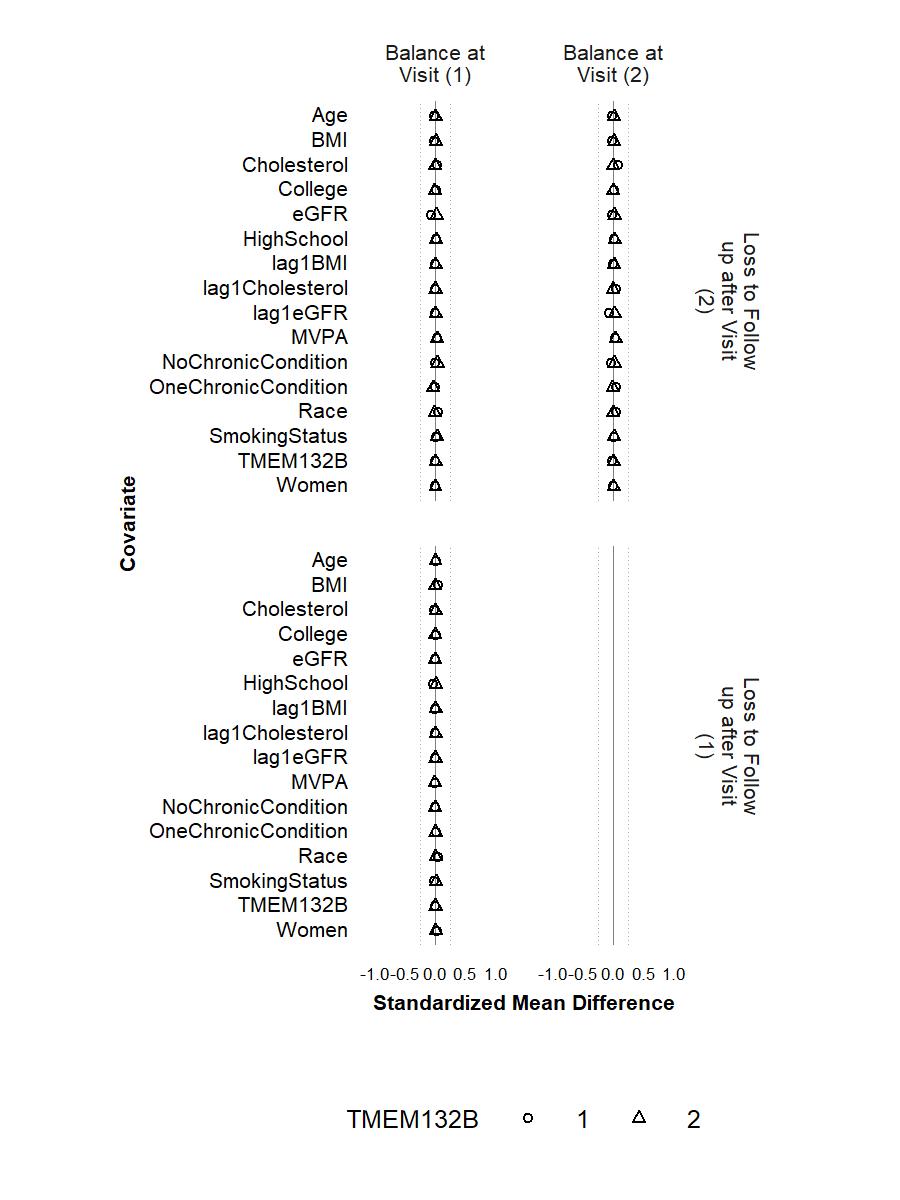

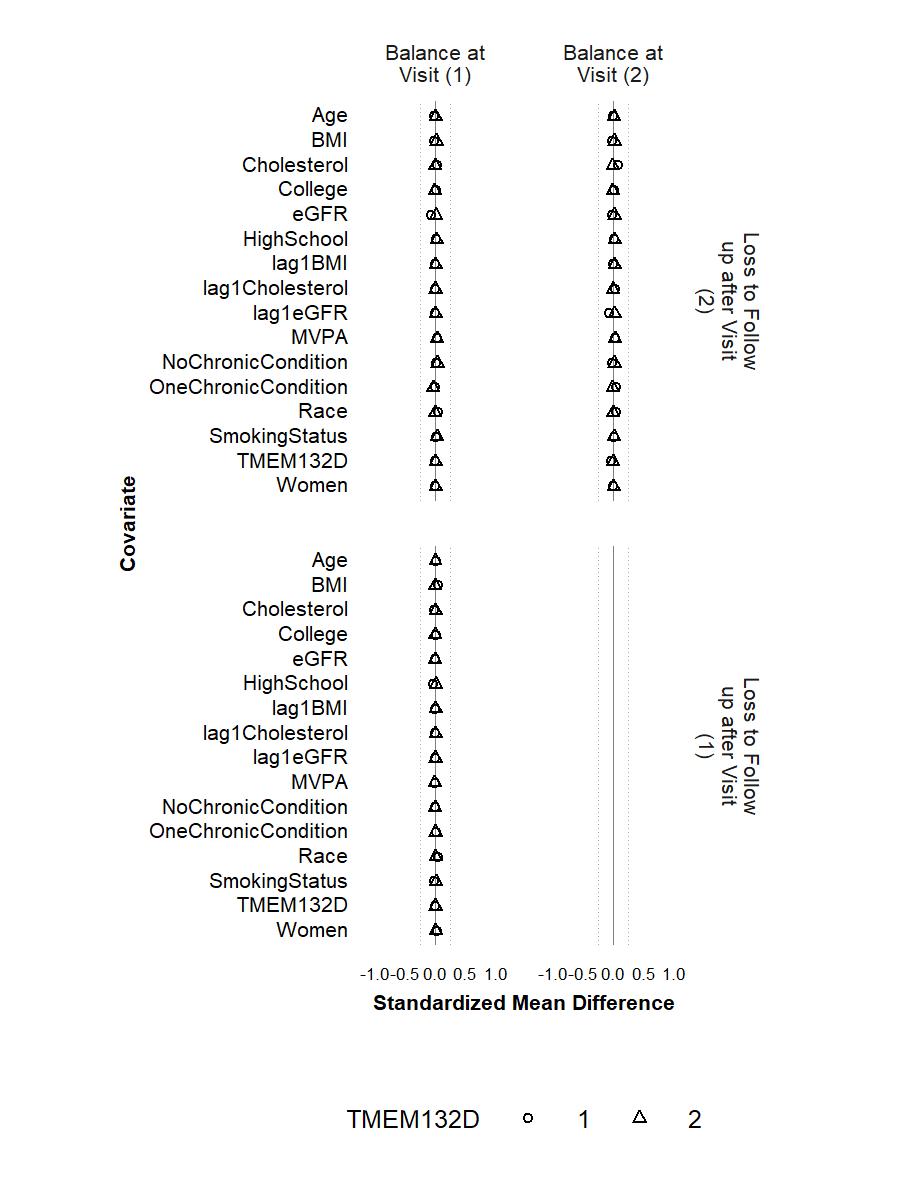

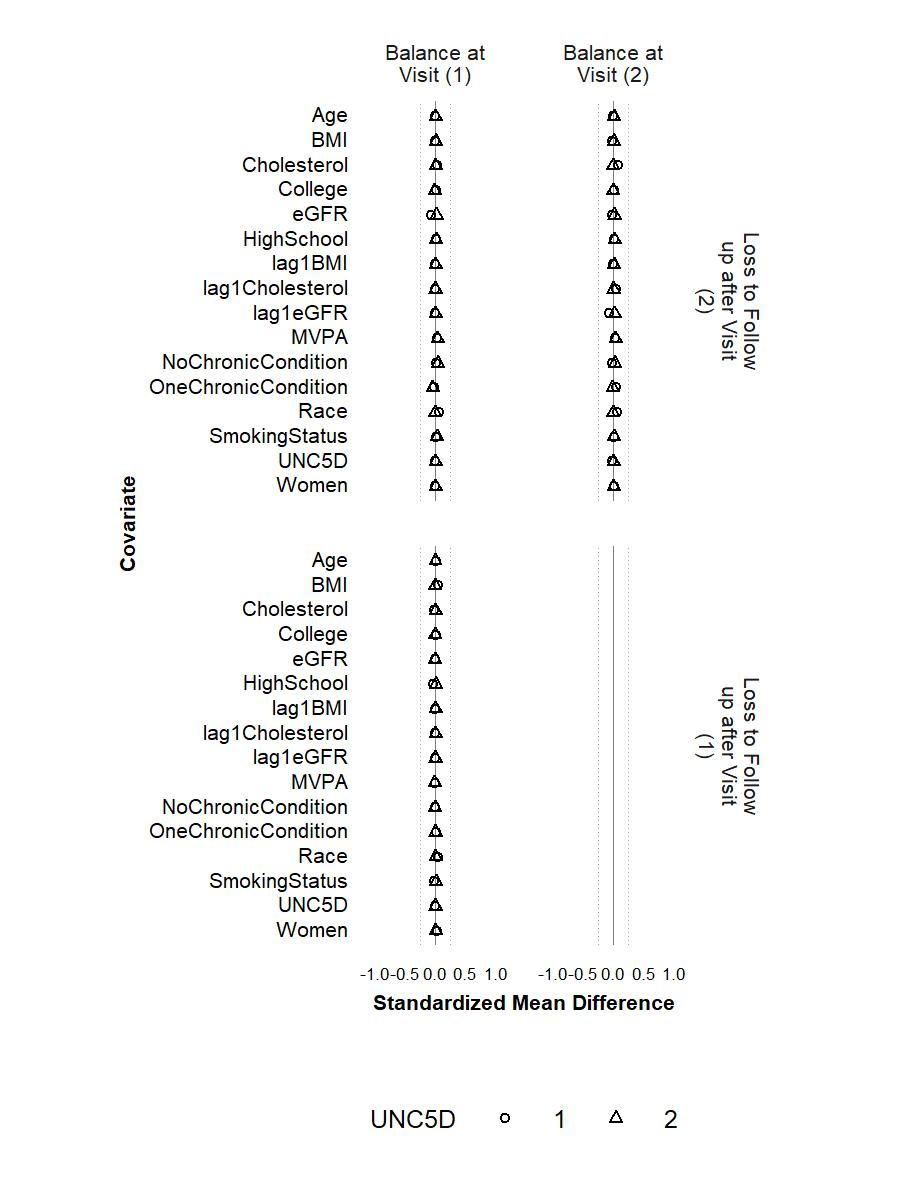

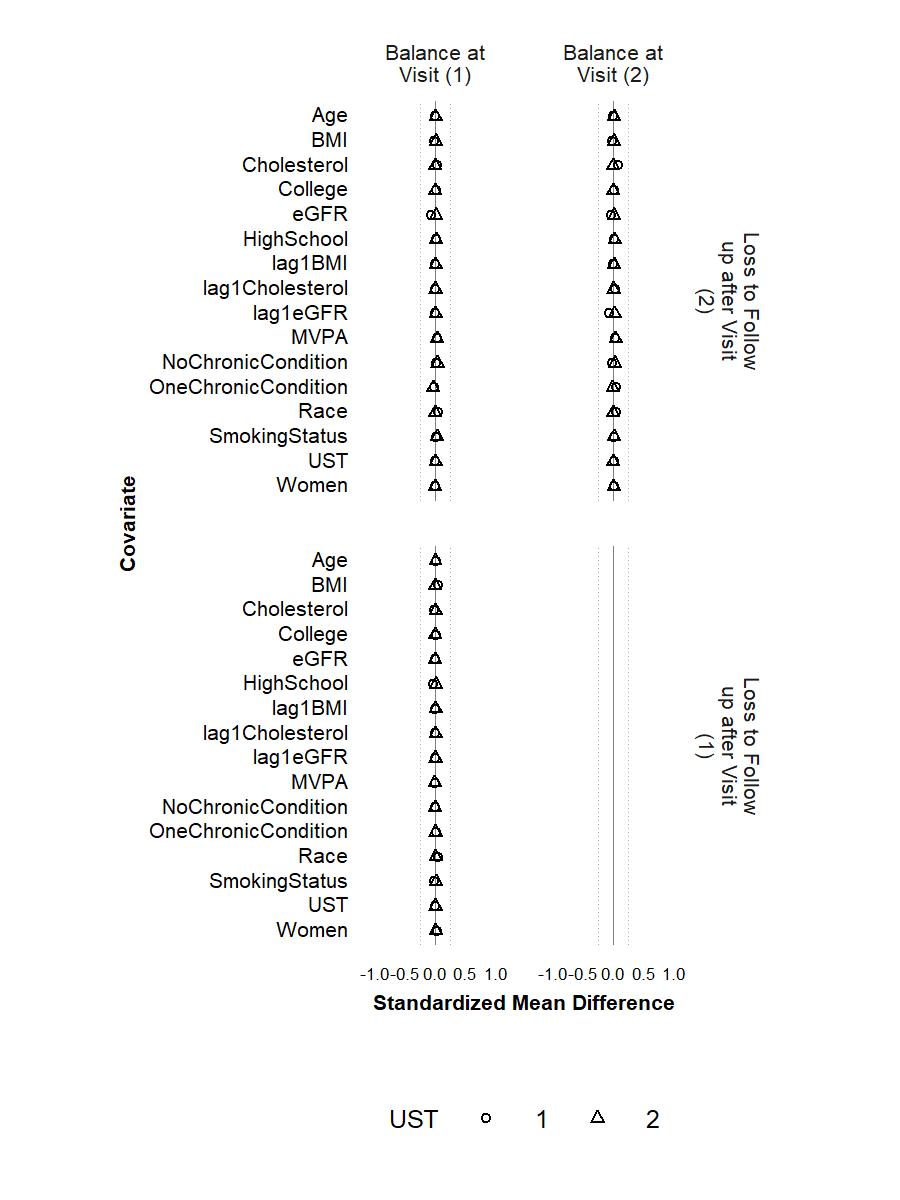

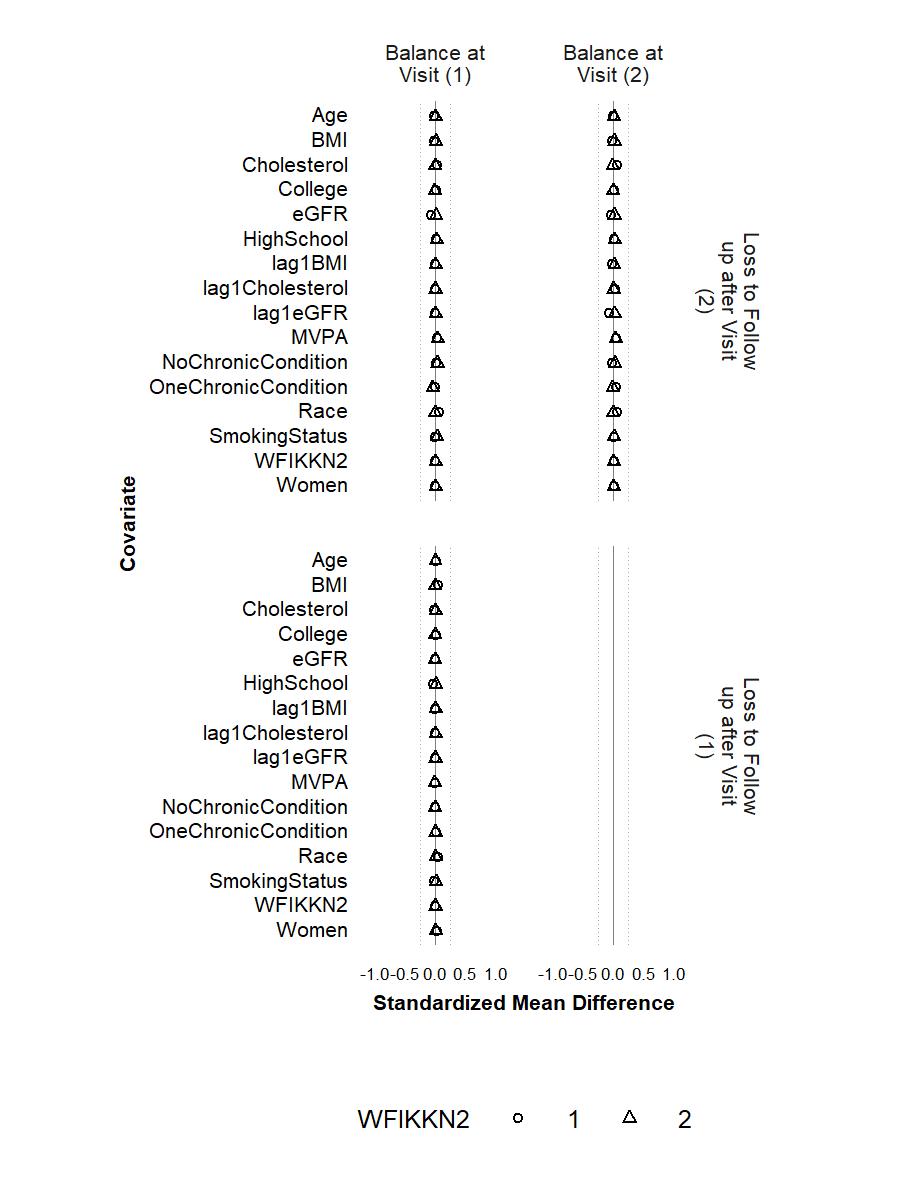

**Table S2.** Causal effects of long-term adherence to physical activity guideline on proteins associated with frailty. Long-term adherence to physical activity guideline was measured by achieving ≥150 minutes/week of MVPA at Visit 1 and Visit 3.

| **SeqID** | **Gene Symbol** | **UniProt ID** | **Target Full Name** | **ICE Method** | **IPW Method** |
| --- | --- | --- | --- | --- | --- |
|  |  |  |  | **Difference (95% CI)** | **Difference (95% CI)** |
| SeqId_7210_25 | APLP1 | P51693 | Amyloid-like protein 1 | **0.046 (0.001, 0.092)** | 0.043 (-0.008, 0.091) |
| SeqId_7970_315 | ART3 | Q13508 | Ecto-ADP-ribosyltransferase 3 | 0.024 (-0.015, 0.06) | 0.012 (-0.05, 0.075) |
| SeqId_7218_87 | ATP1B2 | P14415 | Sodium/potassium-transporting ATPase subunit beta-2 | **0.041 (0.0002, 0.087)** | **0.047 (0.004, 0.092)** |
| SeqId_8885_6 | CACNA2D3 | Q8IZS8 | Voltage-dependent calcium channel subunit alpha-2/delta-3 | **0.096 (0.054, 0.140)** | **0.105 (0.053, 0.152)** |
| SeqId_4129_72 | CFB | P00751 | Complement factor B | **-0.045 (-0.086, -0.001)** | -0.039 (-0.099, 0.022) |
| SeqId_2974_61 | CNTN1 | Q12860 | Contactin-1 | **0.046 (0.008, 0.092)** | 0.056 (-0.021, 0.126) |
| SeqId_4337_49 | CRP | P02741 | C-reactive protein | **-0.075 (-0.114, -0.033)** | **-0.06 (-0.114, -0.015)** |
| SeqId_4324_33 | CST2 | P09228 | Cystatin-SA | 0.019 (-0.025, 0.06) | -0.016 (-0.139, 0.056) |
| SeqId_17366_6 | DCUN1D1 | Q96GG9 | DCN1-like protein 1 | **-0.076 (-0.132, -0.033)** | -0.041 (-0.097, 0.025) |
| SeqId_3607_71 | DKK3 | Q9UBP4 | Dickkopf-related protein 3 | 0.027 (-0.022, 0.07) | 0.033 (-0.02, 0.088) |
| SeqId_9769_48 | DNER | Q8NFT8 | Delta and Notch-like epidermal growth factor-related receptor | **0.048 (0.006, 0.091)** | 0.051 (-0.0004, 0.096) |
| SeqId_5437_63 | FABP3 | P05413 | Fatty acid-binding protein, heart | **-0.056 (-0.093, -0.017)** | -0.033 (-0.097, 0.035) |
| SeqId_15386_7 | FABP4 | P15090 | Fatty acid-binding protein, adipocyte | **-0.056 (-0.100, -0.019)** | -0.035 (-0.094, 0.032) |
| SeqId_3438_10 | FSTL3 | O95633 | Follistatin-related protein 3 | -0.024 (-0.071, 0.021) | -0.029 (-0.092, 0.059) |
| SeqId_17456_53 | GOLM1 | Q8NBJ4 | Golgi membrane protein 1 | -0.01 (-0.051, 0.036) | -0.028 (-0.076, 0.022) |
| SeqId_18896_23 | HS6ST3 | Q8IZP7 | Heparan-sulfate 6-O-sulfotransferase 3 | 0.04 (-0.007, 0.091) | 0.042 (-0.047, 0.115) |
| SeqId_18901_26 | HSPA1B | P0DMV9 | Heat shock 70 kDa protein 1B | -0.033 (-0.076, 0.011) | -0.021 (-0.069, 0.037) |
| SeqId_15594_47 | HTRA1 | Q92743 | Serine protease HTRA1 | **-0.094 (-0.134, -0.049)** | **-0.079 (-0.124, -0.035)** |
| SeqId_2771_35 | IGFBP1 | P08833 | Insulin-like growth factor-binding protein 1 | **0.066 (0.021, 0.114)** | 0.044 (-0.012, 0.101) |
| SeqId_6478_2 | IGLON5 | A6NGN9 | IgLON family member 5 | 0.041 (-0.005, 0.082) | 0.031 (-0.037, 0.086) |
| SeqId_5353_89 | IL1RN | P18510 | Interleukin-1 receptor antagonist protein | 0.007 (-0.038, 0.057) | 0.019 (-0.038, 0.091) |
| SeqId_6408_2 | INHBC | P55103 | Inhibin beta C chain | **-0.065 (-0.111, -0.021)** | -0.044 (-0.095, 0.013) |
| SeqId_4883_56 | INS | P01308 | Insulin | -0.022 (-0.065, 0.036) | 0.022 (-0.053, 0.121) |
| SeqId_8484_24 | LEP | P41159 | Leptin | -0.024 (-0.056, 0.007) | -0.001 (-0.048, 0.049) |
| SeqId_2999_6 | LSAMP | Q13449 | Limbic system-associated membrane protein | 0.015 (-0.02, 0.054) | -0.001 (-0.053, 0.046) |
| SeqId_8080_24 | MSMP | Q1L6U9 | Prostate-associated microseminoprotein | -0.023 (-0.066, 0.022) | -0.03 (-0.078, 0.031) |
| SeqId_4498_62 | NCAM1 | P13591 | Neural cell adhesion molecule 1, 120 kDa isoform | **0.069 (0.032, 0.107)** | **0.067 (0.013, 0.113)** |
| SeqId_15573_110 | NCAN | O14594 | Neurocan core protein | **0.042 (0.005, 0.083)** | 0.039 (-0.009, 0.084) |
| SeqId_5107_7 | NOTCH1 | P46531 | Neurogenic locus notch homolog protein 1 | **0.066 (0.021, 0.116)** | **0.083 (0.032, 0.134)** |
| SeqId_13992_12 | NSF | P46459 | Vesicle-fusing ATPase | **-0.046 (-0.089, -0.008)** | **-0.052 (-0.095, -0.013)** |
| SeqId_19617_5 | PTGR1 | Q14914 | Prostaglandin reductase 1 | -0.021 (-0.062, 0.029) | -0.025 (-0.074, 0.03) |
| SeqId_9296_15 | PTPRD | P23468 | Receptor-type tyrosine-protein phosphatase delta | 0.038 (-0.0003, 0.077) | 0.02 (-0.03, 0.073) |
| SeqId_15515_2 | SAA1 | P0DJI8 | Serum amyloid A-1 protein | **-0.043 (-0.087, -0.001)** | -0.043 (-0.102, 0.011) |
| SeqId_7957_2 | SCG3 | Q8WXD2 | Secretogranin-3 | 0.030 (-0.014, 0.071) | 0.022 (-0.044, 0.077) |
| SeqId_2925_9 | SERPINE1 | P05121 | Plasminogen activator inhibitor 1 | **-0.065 (-0.114, -0.021)** | -0.039 (-0.086, 0.02) |
| SeqId_19563_3 | SEZ6L | Q9BYH1 | Seizure 6-like protein | 0.038 (-0.001, 0.081) | 0.024 (-0.036, 0.079) |
| SeqId_15539_15 | SLITRK1 | Q96PX8 | SLIT and NTRK-like protein 1 | 0.019 (-0.022, 0.058) | 0.005 (-0.057, 0.072) |
| SeqId_10565_19 | SLITRK3 | O94933 | SLIT and NTRK-like protein 3 | **0.052 (0.012, 0.089)** | 0.035 (-0.023, 0.104) |
| SeqId_8305_18 | SULF2 | Q8IWU5 | Extracellular sulfatase Sulf-2 | **-0.058 (-0.100, -0.018)** | -0.046 (-0.095, 0.003) |
| SeqId_18935_14 | TLR5 | O60602 | Toll-like receptor 5 | -0.036 (-0.078, 0.011) | -0.044 (-0.106, 0.017) |
| SeqId_8890_9 | TMEM132B | Q14DG7 | Transmembrane protein 132B | **0.053 (0.010, 0.096)** | 0.038 (-0.009, 0.088) |
| SeqId_13416_8 | TMEM132D | Q14C87 | Transmembrane protein 132D | **0.060 (0.014, 0.107)** | **0.085 (0.02, 0.144)** |
| SeqId_16307_22 | UNC5D | Q6UXZ4 | Netrin receptor UNC5D | 0.026 (-0.017, 0.068) | -0.002 (-0.07, 0.072) |
| SeqId_8364_74 | UST | Q9Y2C2 | Uronyl 2-sulfotransferase | 0.030 (-0.011, 0.078) | 0.055 (-0.036, 0.167) |
| SeqId_3235_50 | WFIKKN2 | Q8TEU8 | WAP, Kazal, immunoglobulin, Kunitz and NTR domain-containing protein 2 | 0.031 (-0.010, 0.073) | 0.013 (-0.035, 0.062) |

**Figure S5.** Balance of **individual chronic conditions, drinking status, family income, and histories of smoking status and number of chronic conditions** between participants who achieved ≥150 minutes/week of MVPA and those who did not at Visit 1 and Visit 3 after weighting with each of the proteins (except for CACNA2D3 and HITRA1) as the outcome. The circle represents the intervention strategy. The dashed lines denote ±0.25 SD.

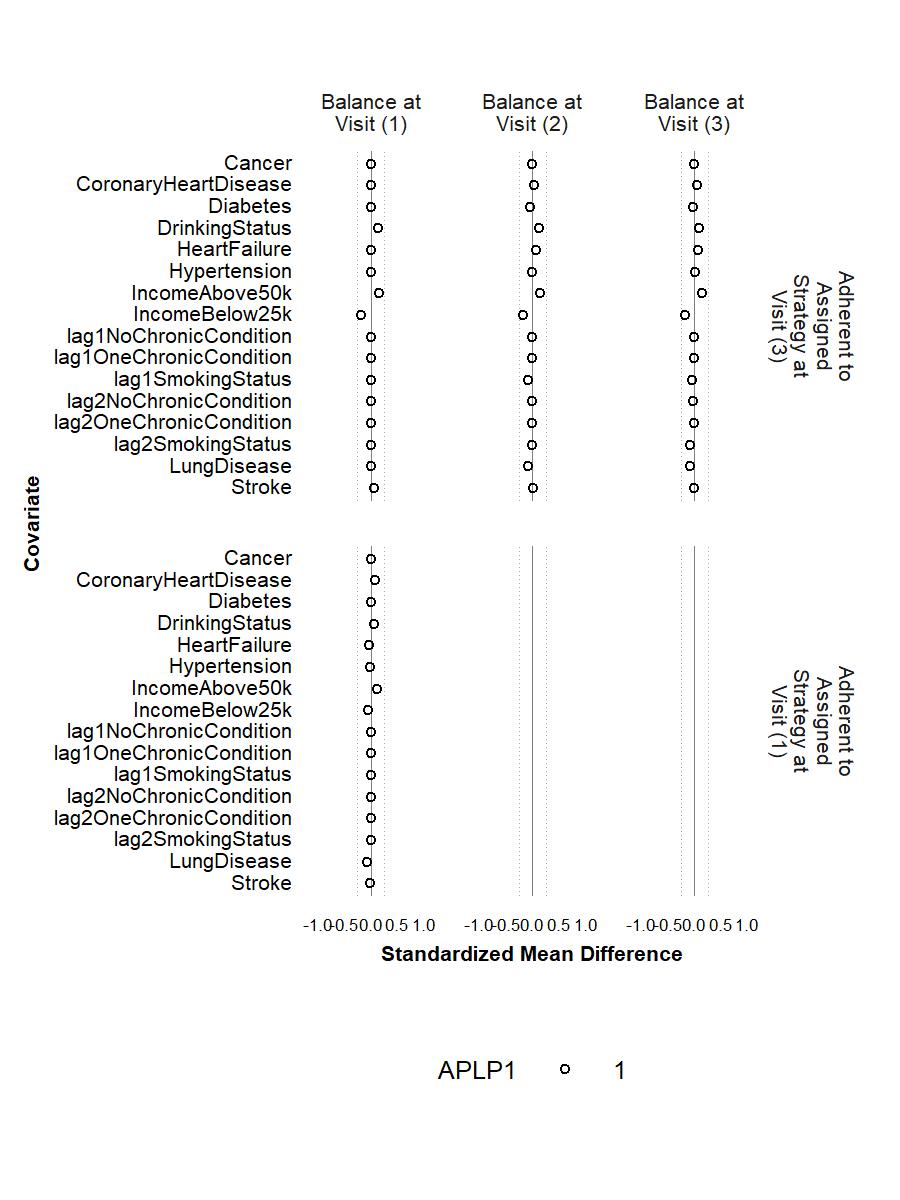

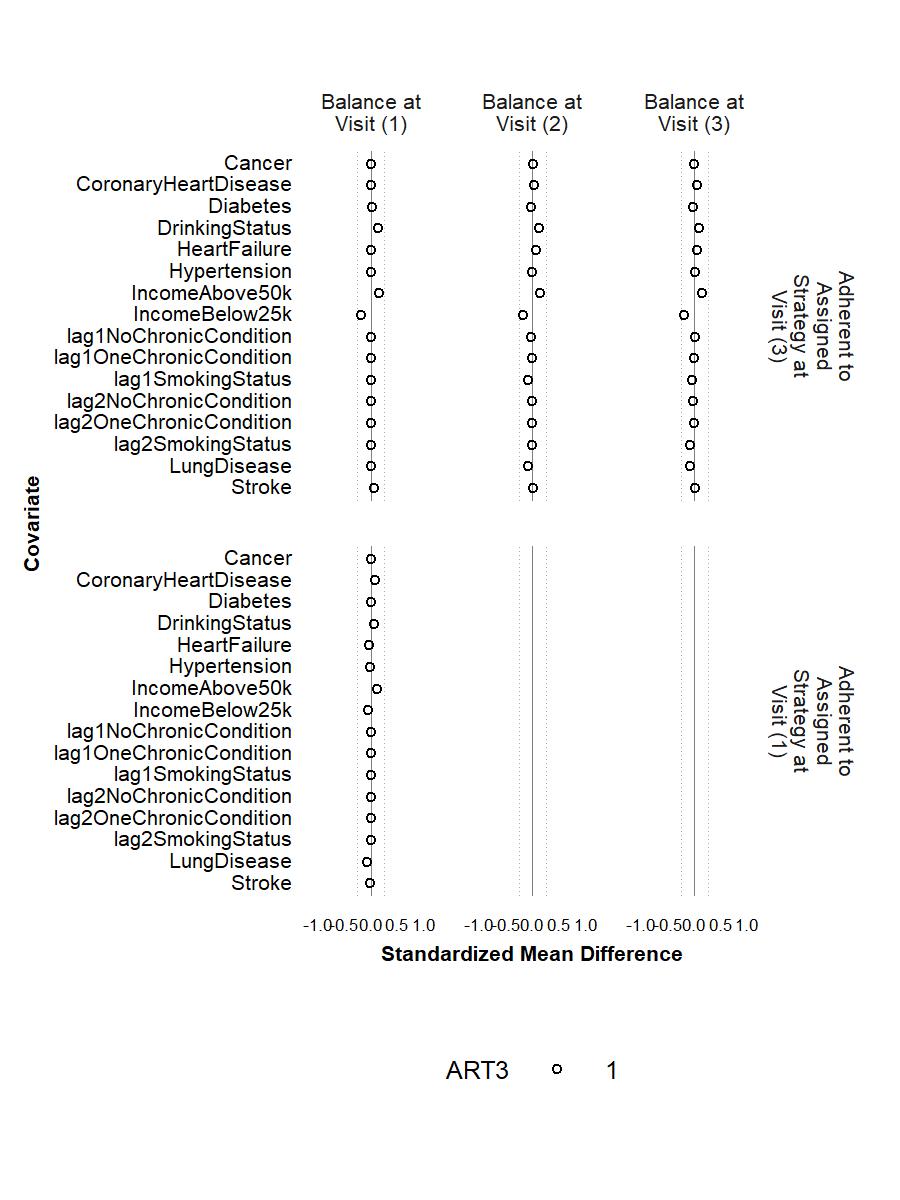

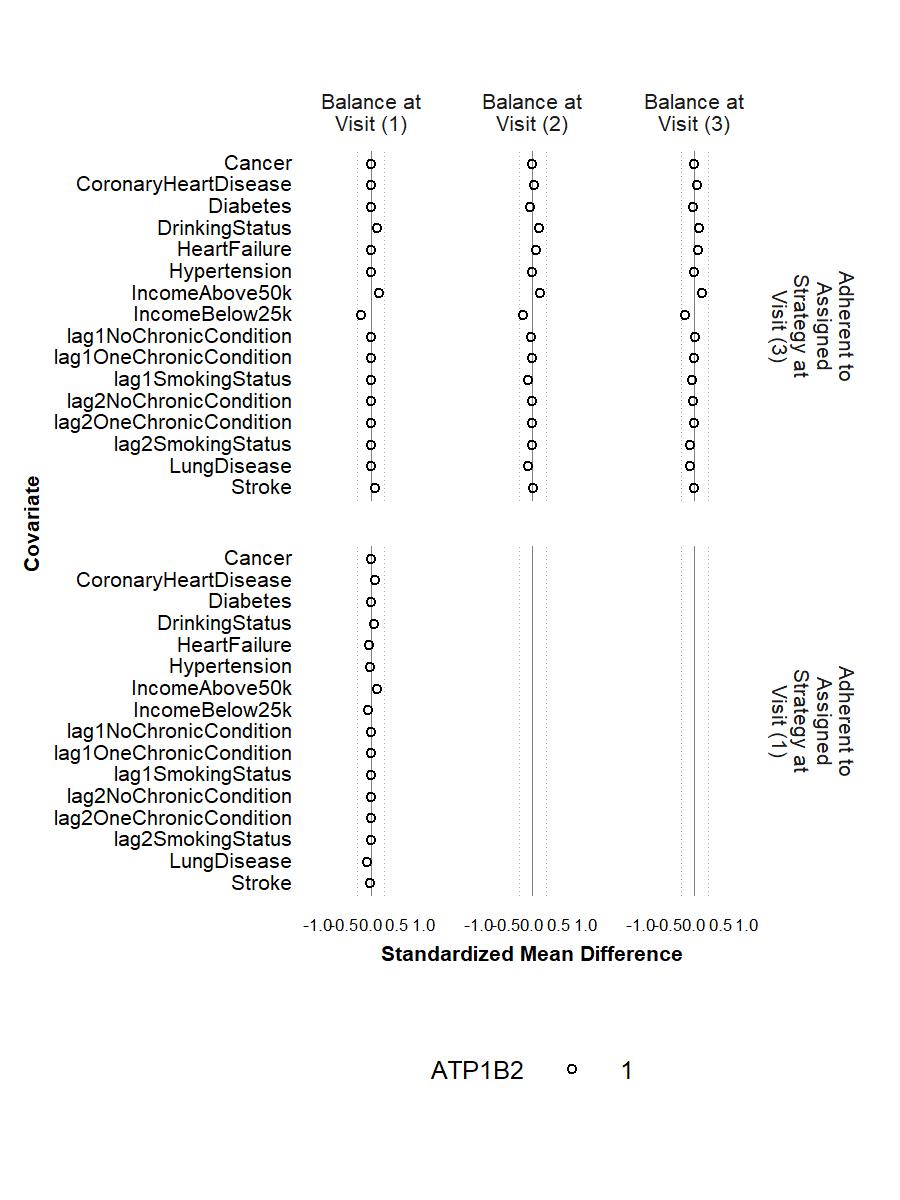

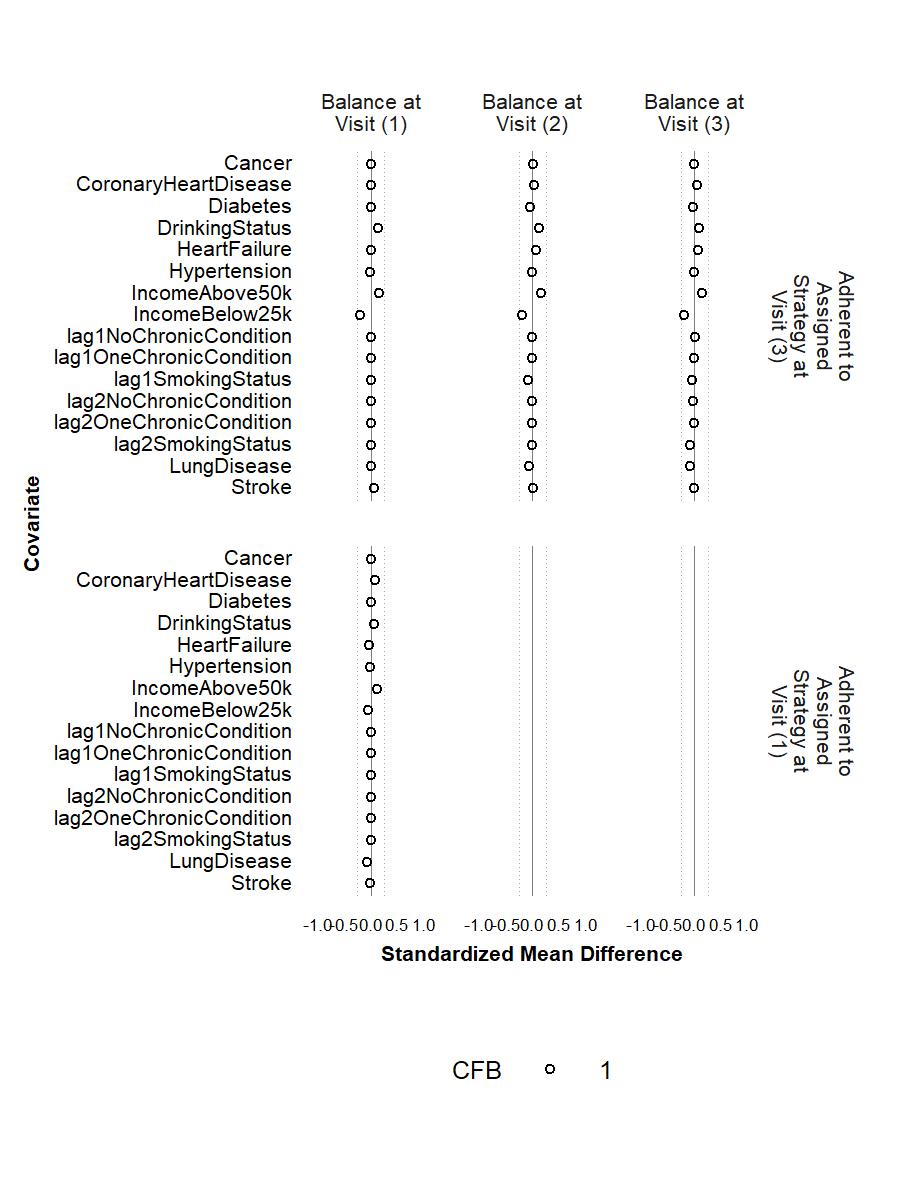

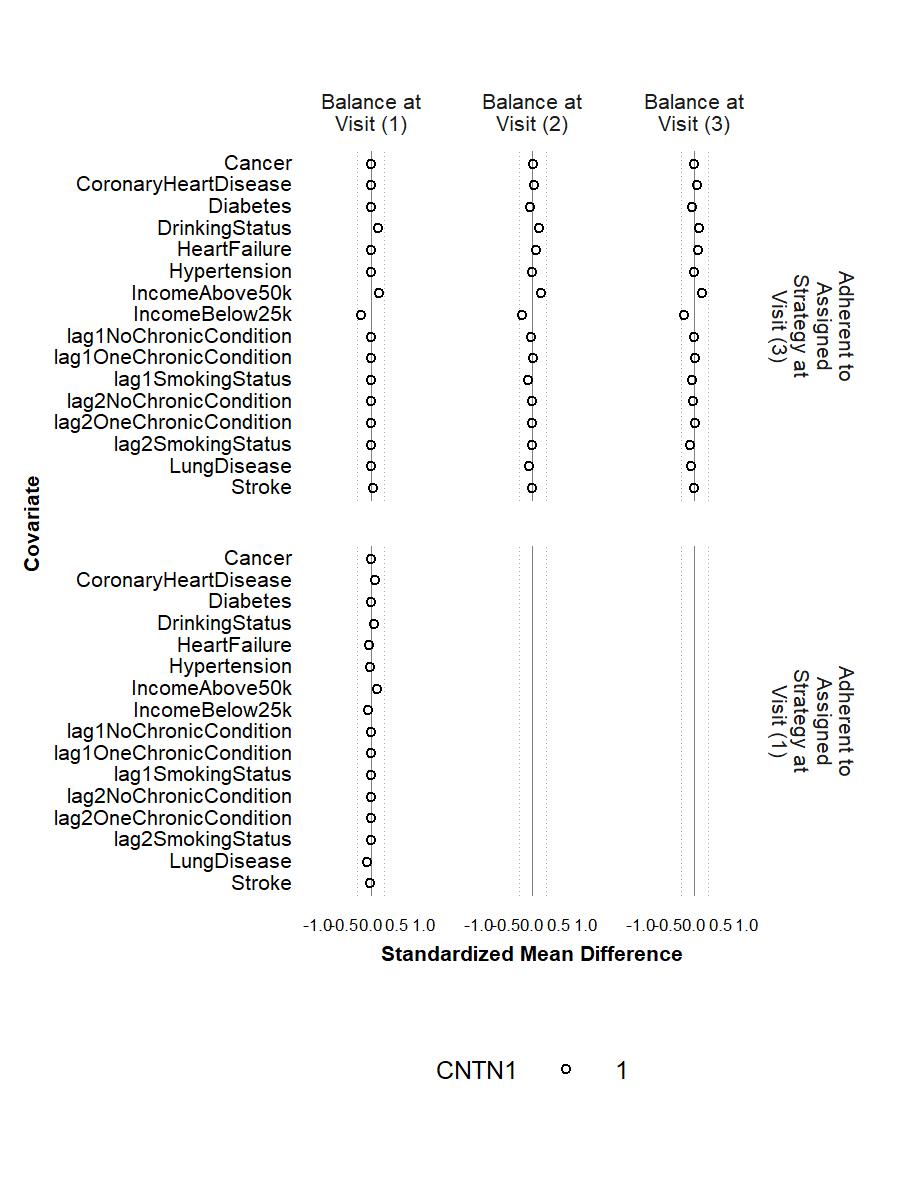

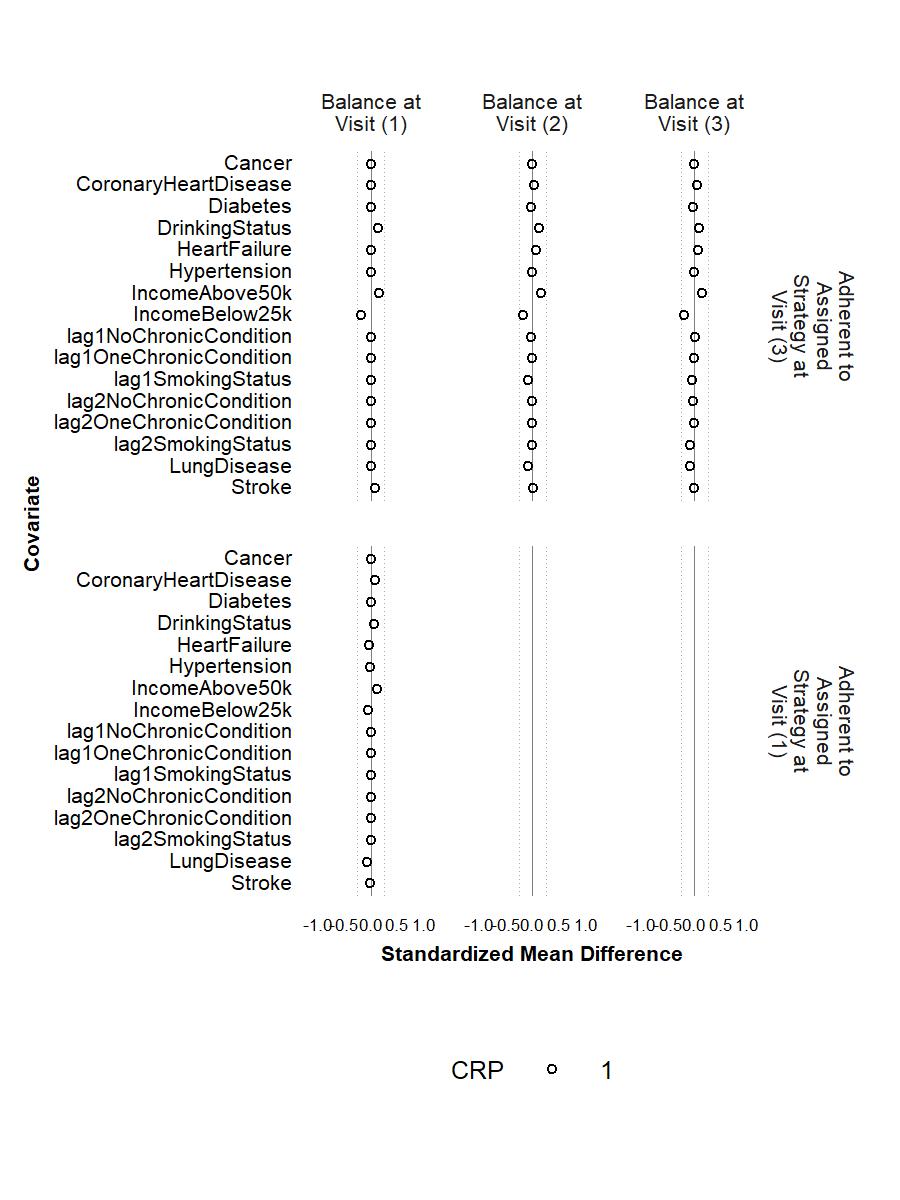

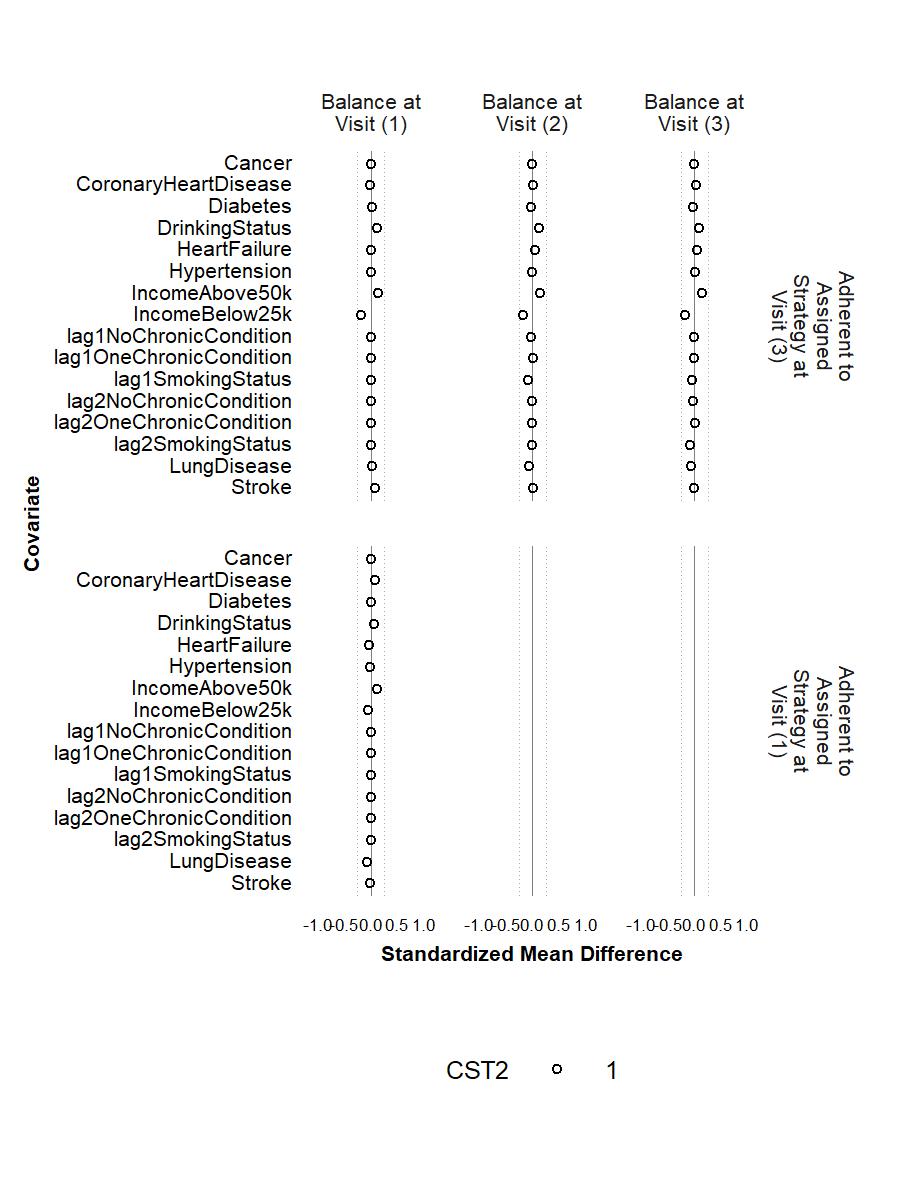

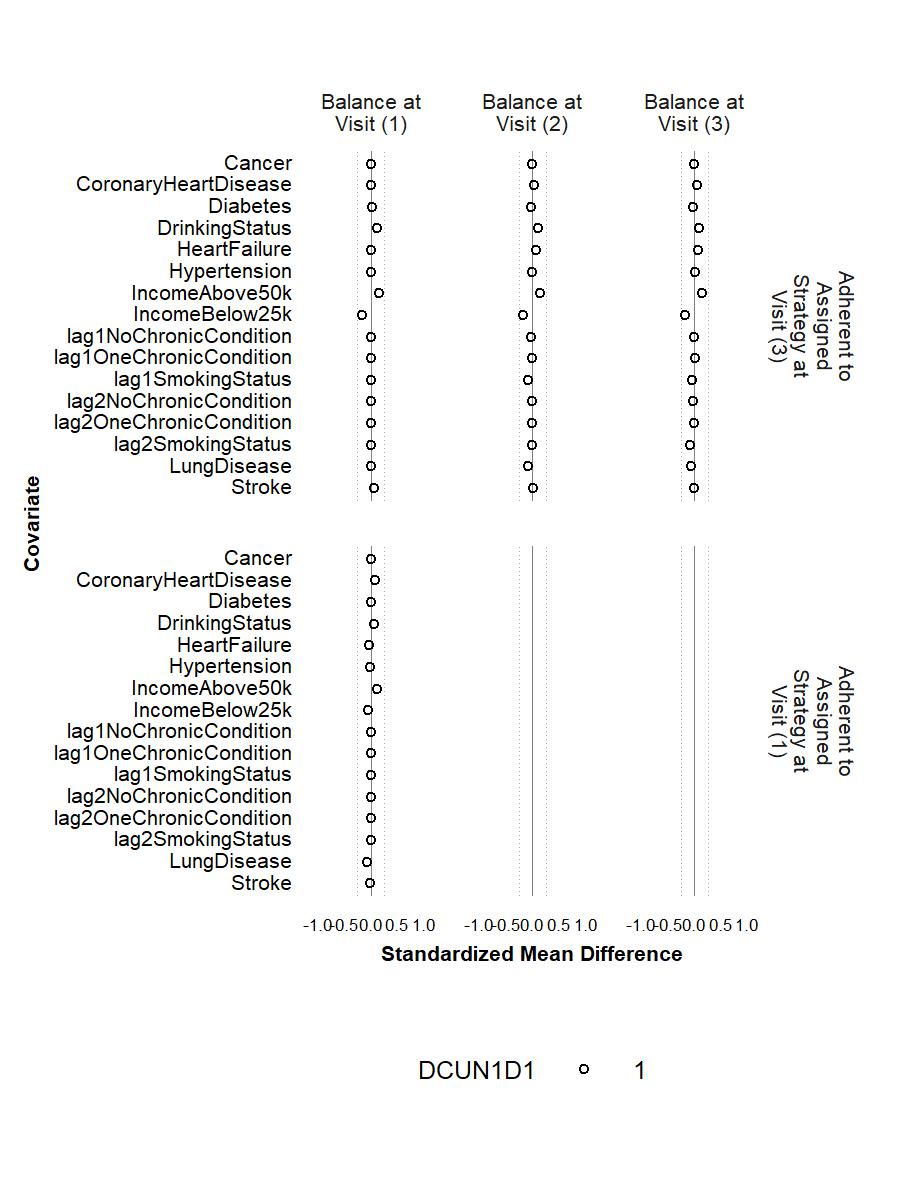

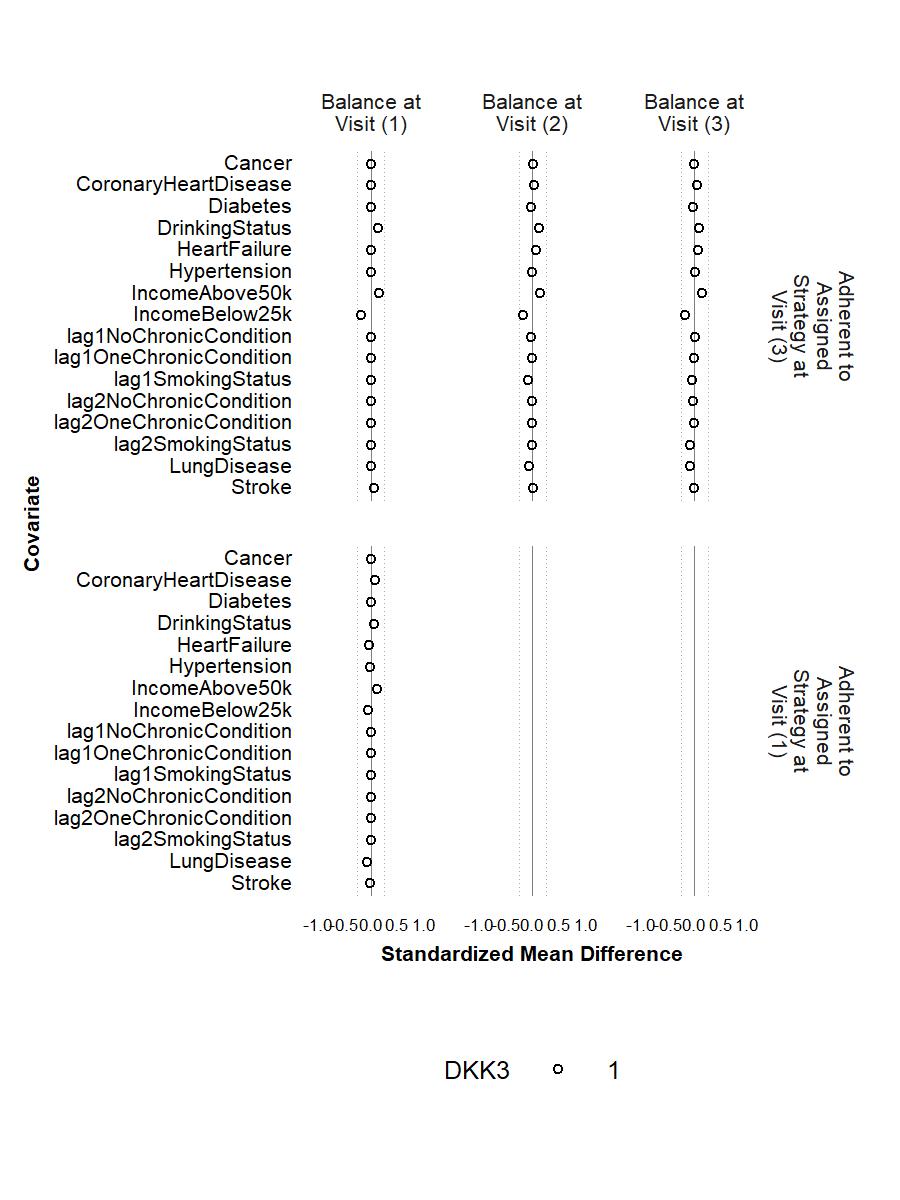

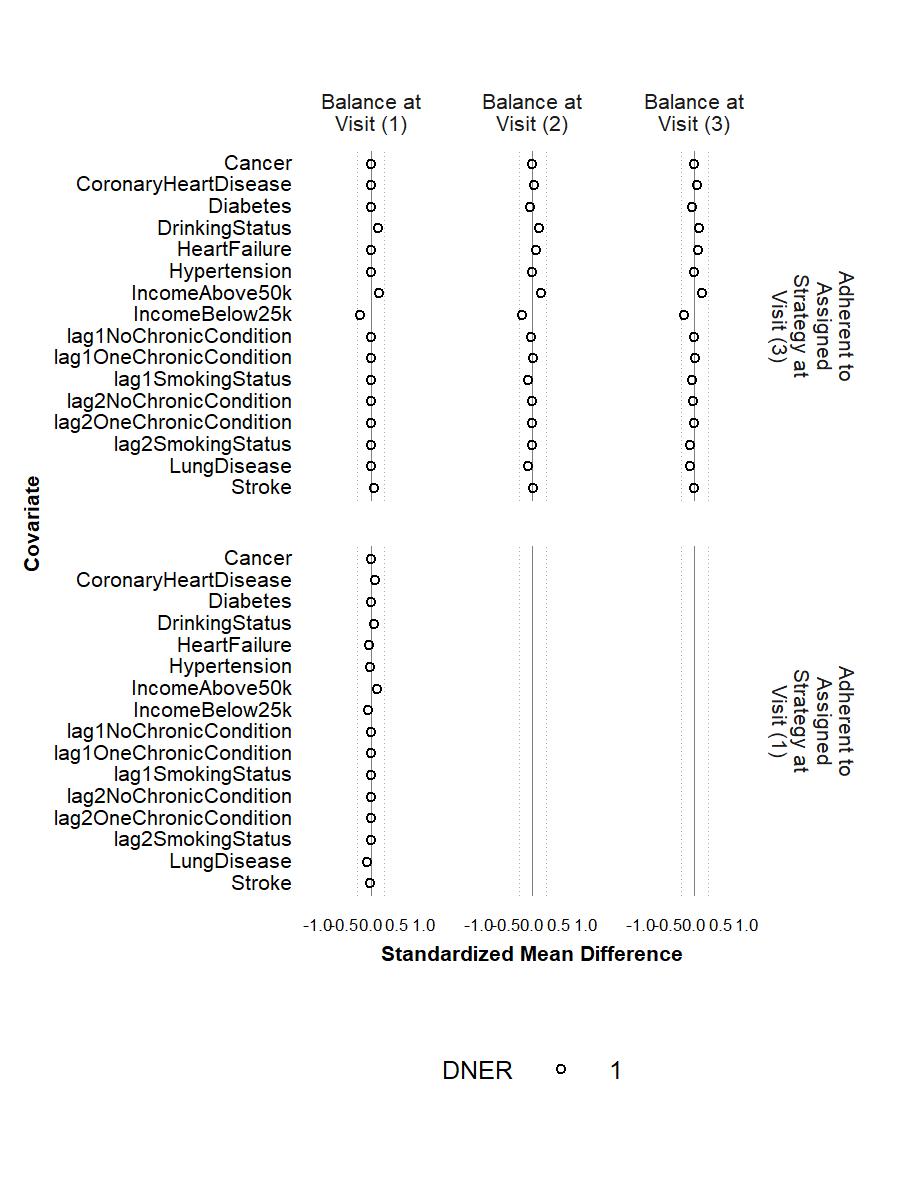

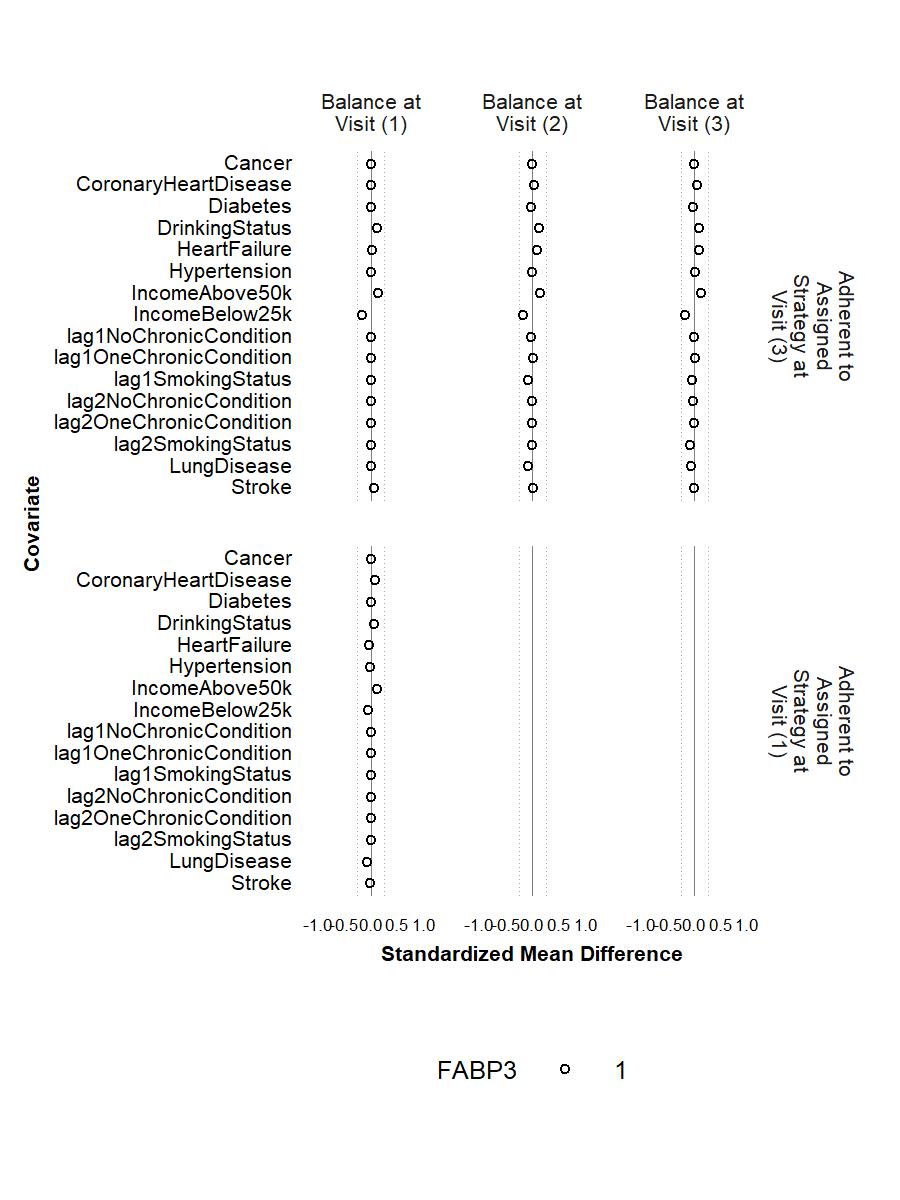

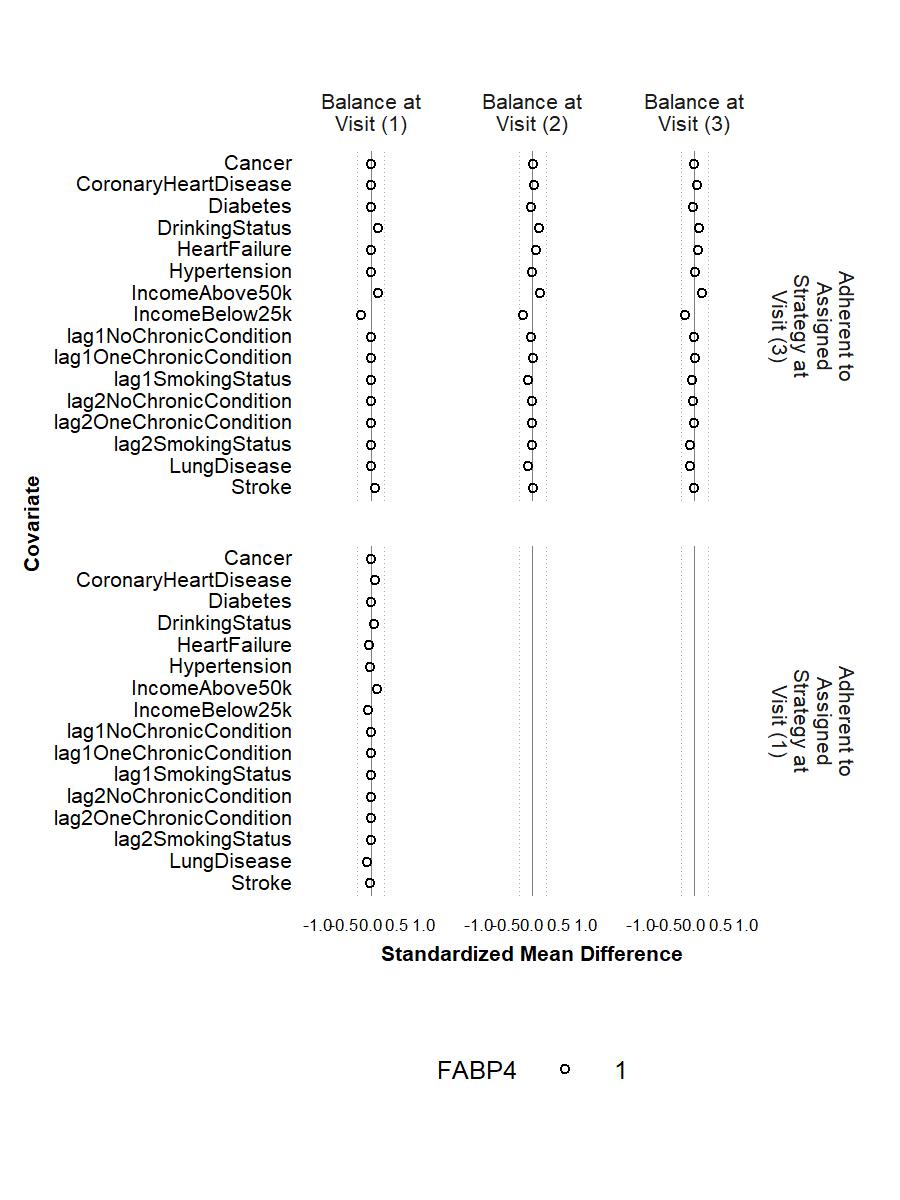

**Figure S6.** Balance of **individual chronic conditions, drinking status, family income, and histories of smoking status and number of chronic conditions** between participants who were lost and those who remained under study at Visits 1 and 2 after weighting with each of the proteins (except for CACNA2D3 and HITRA1) as the outcome. The circle represents the intervention strategy. The triangle represents the control strategy. The dashed lines denote ±0.25 SD.

**Figure S7.** Balance of **included covariates** between participants who achieved ≥150 minutes/week of MVPA and those who did not at Visit 1 and Visit 3 before weighting (A), after weighting with CACNA2D3 as the outcome (B) and after weighting with HITRA1 as the outcome (C) **among participant who did not have major chronic conditions at Visit 1**. The circle represents the intervention strategy. The dashed lines denote ±0.25 SD.

A.

B.

C.

**Figure S8.** Balance of **included covariates** between participants who were lost and those who remained under study at Visits 1 and 2 before weighting (A), after weighting with CACNA2D3 as the outcome (B) and after weighting with HITRA1 as the outcome (C) **among participant who did not have major chronic conditions at Visit 1.** The circle represents the intervention strategy. The triangle represents the control strategy. The dashed lines denote ±0.25 SD.

A.

B.

C.

**Figure S9.** Balance of **individual chronic conditions, drinking status, family income, and histories of smoking status and number of chronic conditions** between participants who achieved ≥150 minutes/week of MVPA and those who did not at Visit 1 and Visit 3 before weighting (A), after weighting with CACNA2D3 as the outcome (B) and after weighting with HITRA1 as the outcome (C) **among participant who did not have major chronic conditions at Visit 1**. The circle represents the intervention strategy. The dashed lines denote ±0.25 SD.

A.

B.

C.

**Figure S10.** Balance of **individual chronic conditions, drinking status, family income, and histories of smoking status and number of chronic conditions** between participants who were lost and those who remained under study at Visits 1 and 2 before weighting (A), after weighting with CACNA2D3 as the outcome (B) and after weighting with HITRA1 as the outcome (C) **among participant who did not have major chronic conditions at Visit 1**. The circle represents the intervention strategy. The triangle represents the control strategy. The dashed lines denote ±0.25 SD.

A.

B.

C.

**Figure S11.** The tipping points of an unmeasured confounder for the effect of MVPA at Visit 1 on 45 frailty-associated proteins.

**Figure S12.** Balance of **included covariates** between participants who achieved ≥150 minutes/week of MVPA and those who did not at Visit 1 and Visit 3 before weighting (A), after weighting with CACNA2D3 as the outcome (B) and after weighting with HITRA1 as the outcome (C) **after excluding participant who died before Visit 3**. The circle represents the intervention strategy. The dashed lines denote ±0.25 SD.

A.

B.

C.

å

**Figure S13.** Balance of **included covariates** between participants who were lost and those who remained under study at Visits 1 and 2 before weighting (A), after weighting with CACNA2D3 as the outcome (B) and after weighting with HITRA1 as the outcome (C) **after excluding participant who died before Visit 3.** The circle represents the intervention strategy. The triangle represents the control strategy. The dashed lines denote ±0.25 SD.

A.

B.

C.

**Figure S14.** Balance of **individual chronic conditions, drinking status, family income, and histories of smoking status and number of chronic conditions** between participants who achieved ≥150 minutes/week of MVPA and those who did not at Visit 1 and Visit 3 before weighting (A), after weighting with CACNA2D3 as the outcome (B) and after weighting with HITRA1 as the outcome (C) **after excluding participant who died before Visit 3**. The circle represents the intervention strategy. The dashed lines denote ±0.25 SD.

A.

B.

C.

**Figure S15.** Balance of **individual chronic conditions, drinking status, family income, and histories of smoking status and number of chronic conditions** between participants who were lost and those who remained under study at Visits 1 and 2 before weighting (A), after weighting with CACNA2D3 as the outcome (B) and after weighting with HITRA1 as the outcome (C) **after excluding participant who died before Visit 3**. The circle represents the intervention strategy. The triangle represents the control strategy. The dashed lines denote ±0.25 SD.

A.

B.

C.
